# Genomics link obesity and type 2 diabetes to Alzheimer’s disease to unveil novel biological insights

**DOI:** 10.64898/2026.02.10.26344393

**Authors:** César Cunha, Mario Garcia-Ureña, Raquel Martínez, María José Romero-Lado, Maria Victoria Fernandez, Ole A. Andreassen, Rebecca Sims, Magda Tsolaki, Kristel Sleegers, Mikko Hiltunen, Gaël Nicolas, Pascual Sánchez-Juan, Martin Ingelsson, Vilmantas Giedraitis, Roberta Ghidoni, Henne Holstege, Cornelia van Duijn, Sven van der Lee, Alfredo Ramirez, Céline Bellenguez, Jean-Charles Lambert, Ruth Frikke-Schmidt, EADB, Tuomas O. Kilpeläinen, Ruth J.F. Loos

**Affiliations:** Novo Nordisk Foundation Center for Basic Metabolic Research, University of Copenhagen, Denmark; Ace Alzheimer Center Barcelona, Universitat Internacional de Catalunya (UIC), Barcelona, Spain; Centre for Precision Psychiatry, Institute of Clinical Medicine, University of Oslo, Oslo, Norway; Division of Mental Health and Addiction, Oslo University Hospital, Oslo, Norway; KG Jebsen Centre for Neurodevelopmental disorders, University of Oslo, Oslo, Norway; Division of Psychological Medicine and Clinical Neuroscience, School of Medicine, Cardiff University, Wales, UK; Aristotle University of Thessaloniki, Greece; VIB Center for Molecular Neurology, VIB, Antwerp, Belgium; Department of Biomedical Sciences, University of Antwerp, Antwerp, Belgium; Institute of Biomedicine, University of Eastern Finland, Yliopistoranta 1E, 70211 Kuopio, Finland; Univ Rouen Normandie, Normandie Univ, Inserm U1245 and CHU Rouen, Department of Genetics and Reference Center for Developmental Disorders, Rouen, France; Alzheimer’s Centre Reina Sofia-CIEN Foundation, Madrid, Spain; CIBERNED, Network Center for Biomedical Research in Neurodegenerative Diseases, National Institute of Health Carlos III, Madrid, Spain; Department of Public Health and Caring Sciences, Molecular Geriatrics, Rudbeck Laboratory, Uppsala University, Uppsala, Sweden; Krembil Brain Institute, University Health Network, Toronto, Ontario, Canada; Tanz Centre for Research in Neurodegenerative Diseases, Departments of Medicine and Laboratory Medicine & Pathobiology, University of Toronto, Toronto, Ontario, Canada; Department of Public Health and Caring Sciences, Clinical Geriatrics, Uppsala University, Uppsala, Sweden; Molecular Markers Laboratory, IRCCS Istituto Centro San Giovanni di Dio Fatebenefratelli, Brescia, Italy; Genomics of Neurodegenerative Diseases and Aging, Human Genetics, Vrije Universiteit Amsterdam; Amsterdam UMC location VUmc, Amsterdam, The Netherlands; Alzheimer Center Amsterdam, Neurology, Vrije Universiteit Amsterdam, Amsterdam UMC location VUmc, Amsterdam, The Netherlands; Amsterdam Neuroscience, Neurodegeneration, Amsterdam, The Netherlands; VIB Center for Brain and Disease Research, ON5 Herestraat 49 - box 602, 3000 Leuven, Belgium Department of Neurosciences, Leuven Brain Institute, KU Leuven, Leuven, Belgium; Department of Epidemiology, ErasmusMC, Rotterdam, The Netherlands; Nuffield Department of Population Health Oxford University, Oxford, UK; Division of Neurogenetics and Molecular Psychiatry, Department of Psychiatry and Psychotherapy, Faculty of Medicine and University Hospital Cologne, University of Cologne, Cologne, Germany; Department for Cognitive Disorders and Old Age Psychiatry, University Hospital Bonn, Bonn, Germany; Department of Psychiatry & Glenn Biggs Institute for Alzheimer’s and Neurodegenerative Diseases, San Antonio, TX, USA; German Center for Neurodegenerative Diseases (DZNE), Bonn, Germany; CECAD; Université de Lille, Inserm, CHU Lille, Institut Pasteur de Lille, Lille, France; Department of Clinical Biochemistry, Copenhagen University Hospital - Rigshospitalet, Blegdamsvej 9, 2100 Copenhagen, Denmark; Department of Clinical Medicine, Faculty of Health and Medical Sciences, University of Copenhagen, Blegdamsvej 3B, 2100 Copenhagen, Denmark

**Keywords:** Alzheimer’s disease, Obesity, Type 2 diabetes, Shared genomics

## Abstract

Body mass index (BMI), type 2 diabetes (T2D) and related cardiometabolic features associate with Alzheimer’s disease (AD) risk, yet shared mechanisms remain poorly understood. By combining multi-trait, machine learning and single-cell transcriptomics with genotyping data for BMI and T2D, we investigate how these traits converge on shared genetic pathways to AD risk. Variant-level analyses reveal AD risk associates with genetically-driven hypotension and hypoglycaemia. We identify sixteen high-confidence effector genes in seven independent loci colocalizing between BMI/T2D and AD, mapping primarily to peripheral immune-metabolic tissues and cell-types. Eight high-confidence risk genes are currently targeted by ongoing therapeutic programmes, substantiating translatability relevance. Exploratory approaches using sex- and age-stratified genotyping data for BMI and T2D pinpoints potential sex-specific cardiometabolic liability linked to higher BMI-associated risk in women and T2D-driven risk in men. These findings elucidate shared cardiometabolic correlates with AD, prioritizes biological pathways, and reveal computationally-nominated drug candidates for preclinical testing.

## INTRODUCTION

Late-onset Alzheimer’s disease (AD) is a devastating disorder that affects patients and families alike. AD is the leading form of dementia^1^, an umbrella term encompassing multiple diseases that progressively deteriorate cognition severely enough to impair independence. Globally, over 55 million individuals live with dementia, affecting approximately one in nine individuals over age 65 and one in three over age 85^2^. With the global aging population projected to reach 2.4 billion individuals over 65 years by 2100^3^, from 800 million currently^4^, the global economic burden of AD is projected to escalate from $2.8 trillion in 2019^5^ to $16.9 trillion by 2050^5^.

Obesity and T2D are independent modifiable risk factors for dementia and AD^6–9^ Addressing modifiable risk factors could prevent 45% of dementia cases, with 12% directly attributable to cardiometabolic factors such as obesity, type 2 diabetes (T2D), high LDL cholesterol, and hypertension^10^ Midlife overweight and obesity elevate dementia risk by 33% and 63% respectively^11^. Likewise, diabetes elevates dementia risk by 59%^12^. Despite strong epidemiological associations, the molecular bases underlying obesity, T2D, and AD are poorly understood. It is unclear whether epidemiological findings stem from non-linear associations such as age-dependent and disease-stage reverse causation or sex-specific differences^13^. This uncertainty is compounded by the prevalent incorporation of AD-proxies, based on undiagnosed parental cases, in genetic association studies^14–16^.

The sex-specific prevalence in AD is striking, with women accounting for two-thirds of cases^17^ and exhibiting significantly higher age-adjusted odds of AD than men at older ages^18^ independent of their longer life expectancy. This contrasts with other neurodegenerative disorders, such as Parkinson’s disease, where men are at twice the risk^19^. Women also show higher susceptibility to APOE4-associated AD risk^20,21^, for which current therapeutic approaches are less effective^22^. The sex-specific prevalence, with obesity being more common in women^23^ and T2D in men^24^, has been proposed to influence the sex-specific risk of AD^20^.

Human genetic data enables systematic decoding of shared biology linking modifiable risk factors to the long preclinical and prodromal curves of AD. We mapped the shared genetic architecture linking obesity, T2D, and AD by integrating genetic correlation, colocalization, phenome-wide association, and protein-protein interaction analyses with deep learning and single-cell transcriptomic approaches to infer relevant biology of effector genes across tissues and cell-types.

## RESULTS

### Study design

Our primary multi-trait analyses used European-ancestry genotyping data from: GIANT consortium meta-analysis (BMI)^25^, T2D genomics Initiative meta-analysis (T2D risk^26^), and European Alzheimer & Dementia Biobank (EADB) consortium meta-analysis (AD risk)^27^ (**Table 1**). The primary AD meta-analysis (AD1) includes parental proxy cases (self-reported by offsprings, undiagnosed), risking signal dilution from potential misclassification (*e.g.* undiagnosed dementia subtypes or lack of neurodegenerative pathology^27–29^). To avoid confounding from the phenotyping of AD, we additionally analyzed AD2, which excludes AD-proxy cases, and AD3, which excludes AD-proxy cases, AD cases, age-matched and non-age-matched controls from the UK Biobank (UKB) to minimize age-related confounding and account for potential sample overlap from other GWAS including UKB participants (**Table 1**). The data were provided by the EADB consortium and has recently been made publicly available^30^.

**Table 1.** Dataset details. The original AD GWAS (AD1) includes clinically diagnosed AD cases (39,106), AD-proxy cases (46,828), age-matched controls (63,137) and non-age-matched controls (338,440). The AD-proxy cases and non-age-matched controls in AD1 derive from the UK Biobank (UKB). AD2 removes the 46,828 AD-proxy cases. AD3 does not include the UKB, resulting in the removal of 46,828 AD-proxy cases, 2,447 clinically diagnosed AD cases and 338,440 non-age-matched controls.

| Dataset | Cases | Controls | Total | Source |
| --- | --- | --- | --- | --- |
| AD1 | 39.106 cases<br>46.828 AD-proxy cases | 401.577 | 487.511 | EADB consortium meta-analysis<sup>1</sup> |
| AD2 | 39.106 cases | 401.577 | 440.683 | EADB consortium meta-analysis |
| AD3 | 36.659 cases | 63.137 | 99.796 |  |
| BMI | - | - | 681.275 | GIANT consortium meta-analysis<sup>2</sup> |
| BMI male | - | - | 374.756 | GIANT consortium<sup>3</sup> |
| BMI female | - | - | 434.794 |  |
| BMI male under 50 | - | - | 50.095 |  |
| BMI male over 50 | - | - | 93.021 |  |
| BMI female under 50 | - | - | 70.692 |  |
| BMI female over 50 | - | - | 106.497 |  |
| T2D | 180.834 | 1.159.055 | 1.339.889 | T2D genomics Initiative meta-analysis<sup>4</sup> |
| T2D male | 41.846 | 383.767 | 425.613 | T2D genomics Initiative<sup>5</sup> |
| T2D female | 30.053 | 434.336 | 464.389 |  |

Our secondary analysis investigated potential sex-specific genetic effects associated with BMI and T2D on AD risk, by evaluating sex-stratified genotyping data from the GIANT consortium (BMI)^31^ and T2D genomics initiative^32^. We additionally analyzed age-stratified BMI data (under/over age 50)^31^. Age 50 approximates the average age of menopause, a critical period affecting metabolic and hormonal risk factors^31^, and marks onset of asymptomatic AD preclinical pathology^33^. Our sex- and age-stratified pipeline is exploratory in nature given the lower sample sizes of stratified BMI and T2D genotyping data and the current unavailability of sex-specific AD genotyping data. The use of sex-combined AD data biases our findings towards the null. While measured indirectly through sex-stratified BMI and T2D genotyping data, if true sex-specific effects exist in AD genetics, using a sex-combined outcome would dilute, not inflate, the sex-specific signal we detect on the exposure side (BMI, T2D). Thus, our approach primarily reflects the sex-specific cardiometabolic liability and its differential shared genetic architecture with AD, not sex-specific AD risk *per se*, representing a conversative lower bound of the true sex-specific effects in AD.

All three AD datasets derive from EADB (AD2 and AD3 are subsets of AD1). All BMI data derives from the GIANT consortium (sex- and age-stratified data are subsets of the combined BMI meta-analysis). All T2D data derives from the DIAMANTE consortium (sex-stratified data are subsets of the combined T2D meta-analysis). We mitigate the risks associated with the overlapping data structure by stratifying our variant discovery into primary (sex-combined) and secondary (sex-stratified) analyses, minimizing the overlap between sex-combined and stratified datasets. The UKB is the only shared component between BMI, T2D and AD. The AD GWAS only used 2,447 cases from the UKB. Even if all UKB AD cases were present in the BMI/T2D GWAS, they would represent <0.5% of the total UKB sample. This issue is largely mitigated in AD2 (removal of AD-proxy cases from UKB) and absent in AD3 (complete removal of UKB participants).

Subsequent analyses included: (1) genetic correlations; (2) risk variant identification through multi-trait colocalization; (3) variant-level phenotyping through phenome-wide associations; (4) effector gene mapping through variant-to-gene linkage; (5) effector tissue and cell-type prioritization through multi-omic and deep learning perturbation analyses; (6) mechanistic detailing through enrichment analyses; and (7) genetic-backed drug target prioritization through gene-drug interaction analyses (**Fig. 1**).

**Fig. 1.**
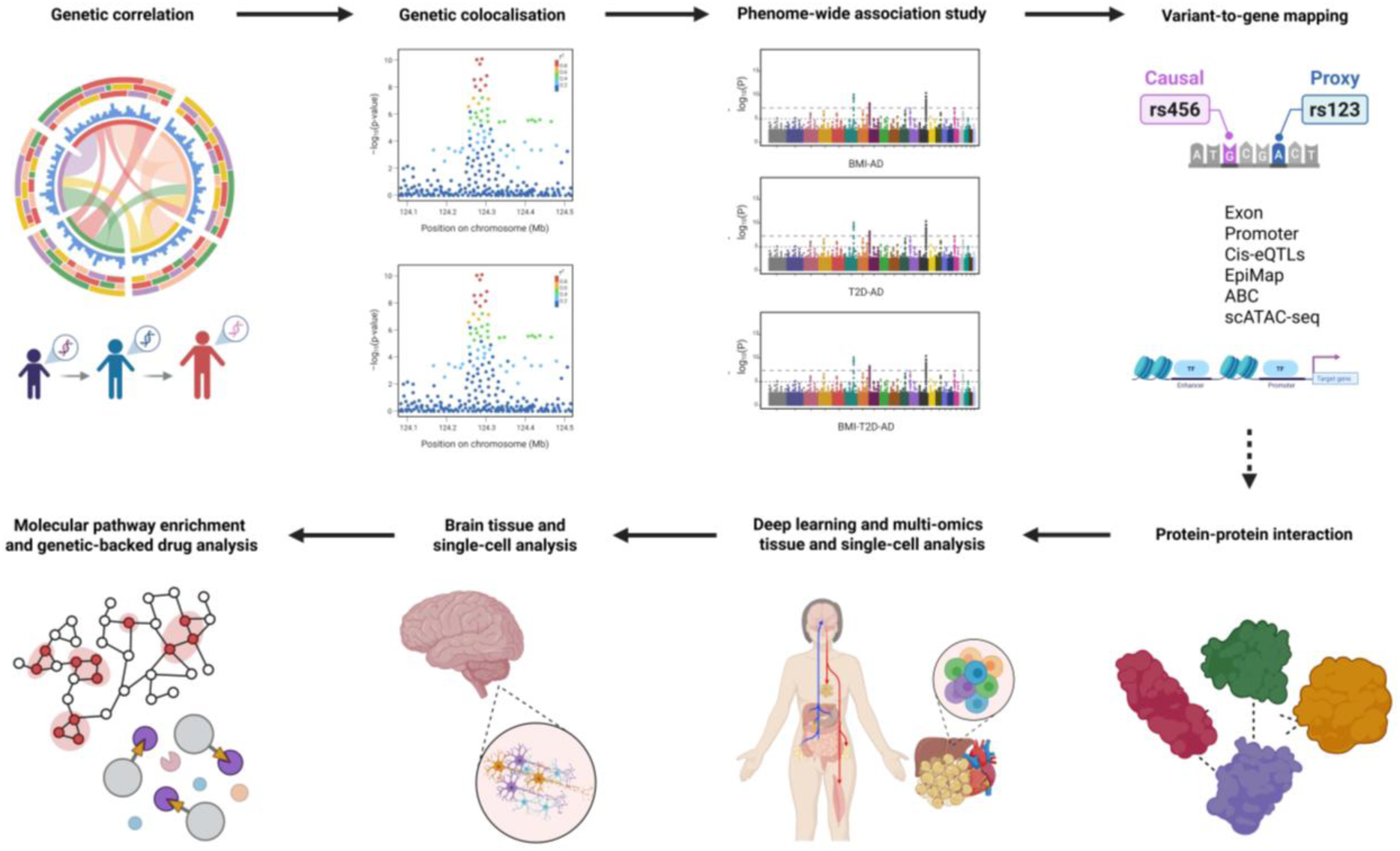
Schematic overview of the study. Created in BioRender.

### Genetic correlations suggest sex- and age-specific BMI liability linked to AD risk

We assessed genetic correlations (r_g_) between BMI, T2D, and AD using linkage disequilibrium score regression (**LDSC**; **Fig. 2**; **Table S1**; **Methods**).

**Fig. 2.**
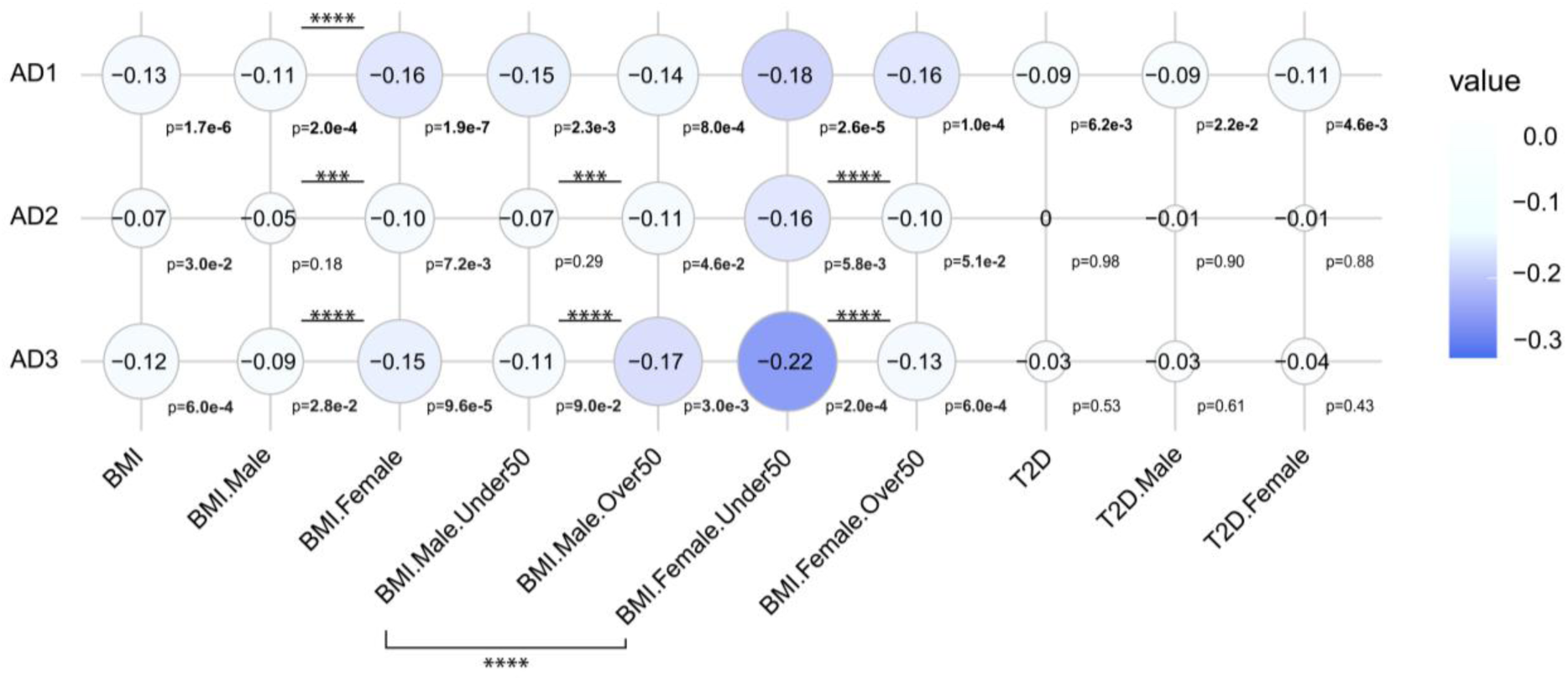
Cross-phenotype genetic correlation estimates. AD1: Includes AD proxy cases and non-age-matched controls. AD2: Excludes AD proxy cases, but retains non-age matched controls. AD3: Excludes AD proxy cases and non-age-matched controls. Sex-combined datasets: AD1, AD2, AD3, BMI, T2D. Sex and age-stratified datasets: BMI.Male, BMI.Female, BMI.Male.Under50, BMI.Male.Over50, BMI.Female.Under50, BMI.Female.Over50, T2D.Male, T2D.Female. The genetic correlation matrix was generated using the *corrplot* library in R (version 4.2.2). P-values are shown next to GC estimates. Statistical significance between genetic correlation estimates are stratified by p<0.05 (*); p<0.01 (**); p<0.001 (***); p<0.0001 (****) (**Table S1**).

In the primary sex-combined analysis, BMI showed negative genetic correlation with AD risk (r_g_= −0.13; p=1×10^−6^), linking lower BMI variants with higher AD risk. This correlation was significantly stronger in women than men across all AD datasets (p<4×10^−4^).

In the secondary sex-stratified analysis, divergent age patterns (≤50y versus >50y) emerged when excluding AD cases and controls from the UKB. In men, a stronger genetic correlation was observed for AD and BMI >50y versus ≤50y (p<6×10^−4^). In women, a stronger genetic correlation was observed for AD and BMI ≤50y versus >50y (p<6×10^−6^). The genetic correlation for BMI ≤50y is significantly stronger for women versus men (p<5×10^−6^), but non-significant >50y. These sex and age patterns were absent in AD-proxy- and UKB-inclusive data, indicating sex and age-dependent specificity between BMI and clinically diagnosed AD that disappeared when including AD-proxy cases.

No sex differences emerged in the genetic correlation between T2D and AD despite significant correlation in the overall population (r_g_= −0.09; p=6×10^−3^; **Table S1**). However, excluding AD-proxy cases and UKB abolished this correlation, suggesting that the shared T2D and AD architecture may be biased by AD-proxies or reflect broader dementia susceptibility rather than AD-specific biology. Sensitivity analyses confirm that the APOE region does not drive the estimates reported here, with genetic correlations remaining virtually unchanged after APOE exclusion (**Table S1**).

Overall, BMI showed stronger genetic correlation with AD than T2D (**Table S1**). Polygenic BMI risk in women is associated with stronger genetic susceptibility to AD. Marked age-sensitivity, particularly under 50 years for BMI in women, suggests non-linear, age- and sex-dependent shared genetic mechanisms with AD.

### Colocalization analysis identifies unique loci between BMI, T2D and AD

Genomic colocalization was performed following an initial filtering step where we identified genome-wide significant AD variants (p≤5×10^−8^) concurrently associated with BMI and/or T2D (p≤5×10^−4^), yielding 32 unique BMI-AD and 16 T2D-AD overlapped loci (**Table S2; Methods**). Colocalization analyses, complemented by locuszoom visual inspection (**Fig. 3**A), identified 14 shared causal variants in seven independent loci (r^2^<0.01; ±500 kb): four shared loci for BMI and AD (BMI-AD), two shared loci for T2D and AD (T2D-AD) and one shared locus for BMI, T2D and AD (BMI-T2D-AD) (**Fig. 3**B; **Methods**). The low proportion of overlapping loci that colocalize indicates that most cross-phenotype associations at these loci are not attributable to a single shared causal variant within the tested genomic windows.

**Fig. 3.**
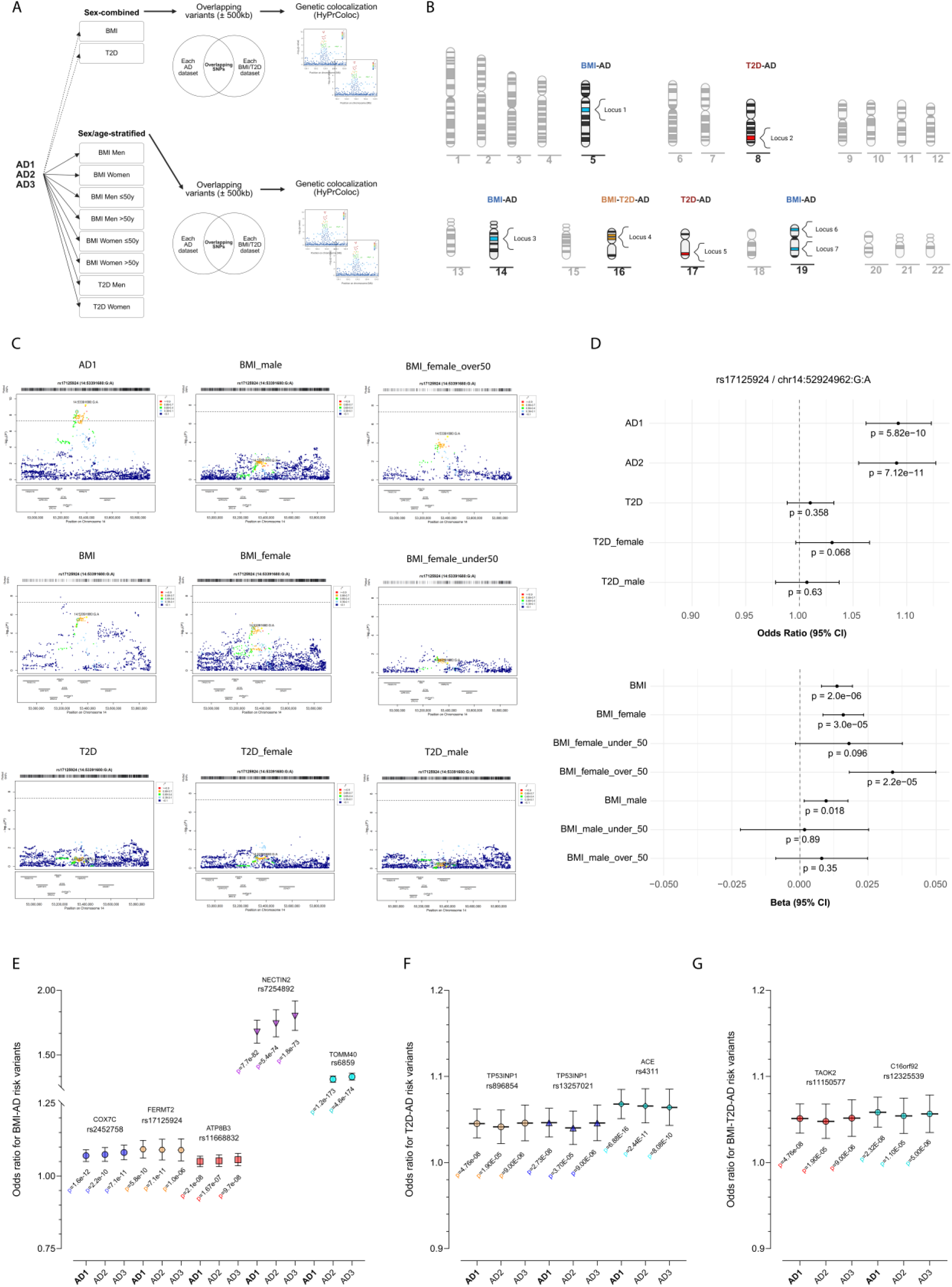
Genetic colocalization analysis. (**A**) Schematic overview of the multi-trait colocalized locus discovery pipeline, subdivided into the primary sex-combined and secondary sex-stratified analyses. (**B**) Karyotype representation of the colocalized loci for BMI-AD (chr5, chr14, chr19), T2D-AD (chr8, chr17) and BMI-T2D-AD (chr16). (**C**) Locuszoom plots for the BMI-AD rs17125924 locus (GRCh37). The overlapped variant is circled in green (**Table S3**) while the colocalized variant is shown in yellow (rs17125924). Variants in linkage disequilibrium (R^2^) are color-coded. (**D**) Forest plot for the BMI-AD rs17125924 locus showcasing odds ratio for dichotomous traits (AD and T2D) and effect size (beta) for the continuous trait (BMI). P-values are shown under each odds ratio/beta estimate. Odds ratio and p-values for (E) BMI-AD, (F) T2D-AD and (G) BMI-T2D-AD risk variants across AD datasets (AD1, AD2 and AD3).

We mapped each locus to the closest gene for orientation. COX7C and FERMT2 hits associate with increased BMI and AD risk. ATP8B3 and NECTIN2-TOMM40 (APOE) hits associate with higher AD risk but lower BMI. T2D-AD and BMI-T2D-AD hits are associated with higher AD risk but lower BMI and T2D risk (**Table 2**).

**Table 2.**
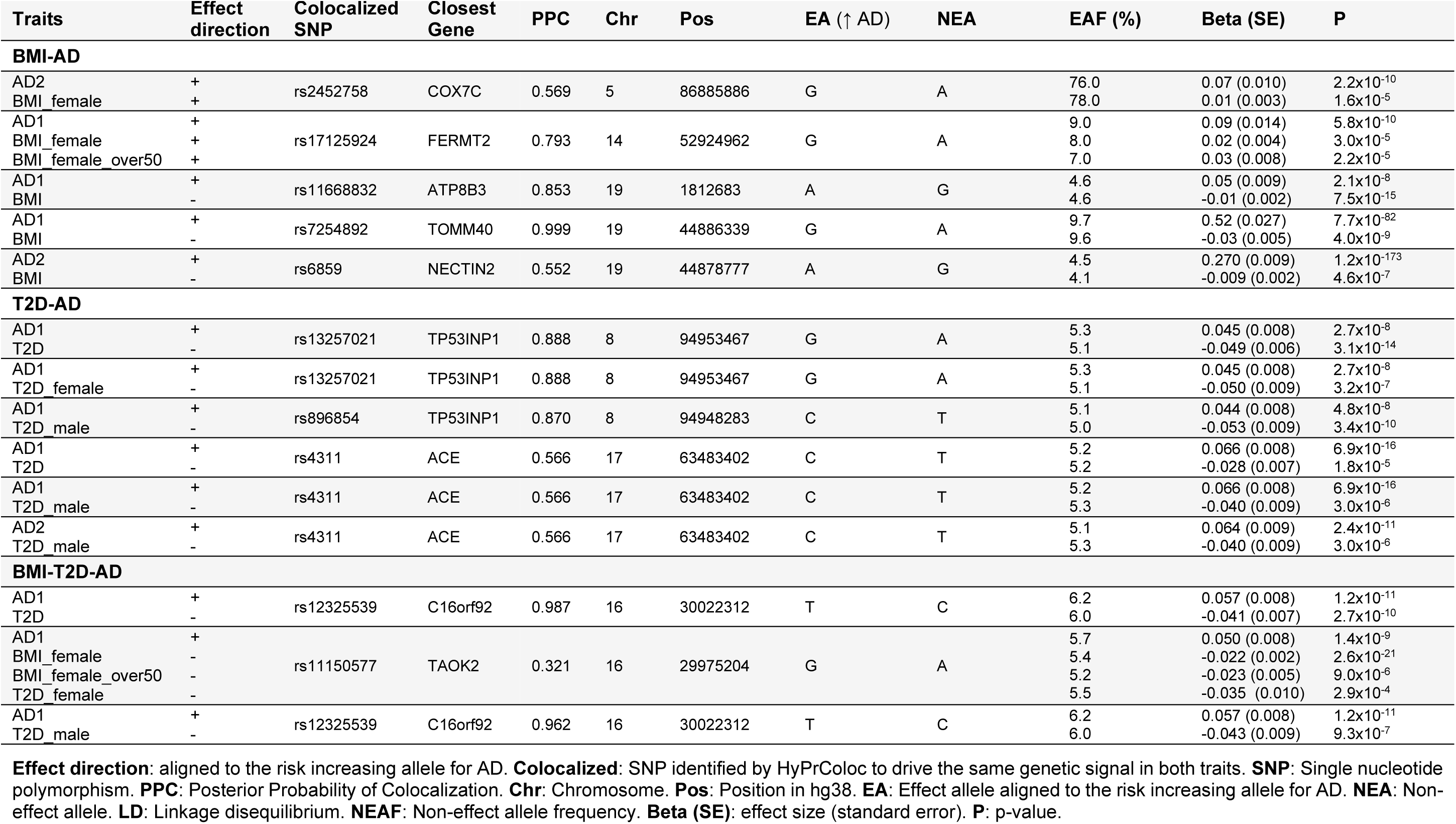
Colocalized loci linked to BMI-AD, T2D-AD and BMI-T2D-AD (full details in Table S3). Colocalization signals derived from the primary sex-combined analysis (BMI, T2D) and secondary (BMI_female, BMI_female_over50, T2D_female, T2D_male) pipelines are shown. Of note, the BMI-T2D-AD colocalization signals rs12325539 (sex-combined) and rs11150577 (sex-stratified) are in high LD (R^2^=0.77).

| Traits | Effect direction | Colocalized SNP | Closest Gene | PPC | Chr | Pos | EA (↑ AD) | NEA | EAF (%) | Beta (SE) | P |
| --- | --- | --- | --- | --- | --- | --- | --- | --- | --- | --- | --- |
| BMI-AD |  |  |  |  |  |  |  |  |  |  |  |
| AD2 | + | rs2452758 | COX7C | 0.569 | 5 | 86885886 | G | A | 76.0 | 0.07 (0.010) | 2.2x10<sup>-10</sup> |
| BMI_female | + |  |  |  |  |  |  |  | 78.0 | 0.01 (0.003) | 1.6x10<sup>-5</sup> |
| AD1 | + | rs17125924 | FERMT2 | 0.793 | 14 | 52924962 | G | A | 9.0 | 0.09 (0.014) | 5.8x10<sup>-10</sup> |
| BMI_female | + |  |  |  |  |  |  |  | 8.0 | 0.02 (0.004) | 3.0x10<sup>-5</sup> |
| BMI_female_over50 | + |  |  |  |  |  |  |  | 7.0 | 0.03 (0.008) | 2.2x10<sup>-5</sup> |
| AD1 | + | rs11668832 | ATP8B3 | 0.853 | 19 | 1812683 | A | G | 4.6 | 0.05 (0.009) | 2.1x10<sup>-8</sup> |
| BMI | - |  |  |  |  |  |  |  | 4.6 | -0.01 (0.002) | 7.5x10<sup>-15</sup> |
| AD1 | + | rs7254892 | TOMM40 | 0.999 | 19 | 44886339 | G | A | 9.7 | 0.52 (0.027) | 7.7x10<sup>-82</sup> |
| BMI | - |  |  |  |  |  |  |  | 9.6 | -0.03 (0.005) | 4.0x10<sup>-9</sup> |
| AD2 | + | rs6859 | NECTIN2 | 0.552 | 19 | 44878777 | A | G | 4.5 | 0.270 (0.009) | 1.2x10<sup>-173</sup> |
| BMI | - |  |  |  |  |  |  |  | 4.1 | -0.009 (0.002) | 4.6x10<sup>-7</sup> |
| T2D-AD |  |  |  |  |  |  |  |  |  |  |  |
| AD1 | + | rs13257021 | TP53INP1 | 0.888 | 8 | 94953467 | G | A | 5.3 | 0.045 (0.008) | 2.7x10<sup>-8</sup> |
| T2D | - |  |  |  |  |  |  |  | 5.1 | -0.049 (0.006) | 3.1x10<sup>-14</sup> |
| AD1 | + | rs13257021 | TP53INP1 | 0.888 | 8 | 94953467 | G | A | 5.3 | 0.045 (0.008) | 2.7x10<sup>-8</sup> |
| T2D_female | - |  |  |  |  |  |  |  | 5.1 | -0.050 (0.009) | 3.2x10<sup>-7</sup> |
| AD1 | + | rs896854 | TP53INP1 | 0.870 | 8 | 94948283 | C | T | 5.1 | 0.044 (0.008) | 4.8x10<sup>-8</sup> |
| T2D_male | - |  |  |  |  |  |  |  | 5.0 | -0.053 (0.009) | 3.4x10<sup>-10</sup> |
| AD1 | + | rs4311 | ACE | 0.566 | 17 | 63483402 | C | T | 5.2 | 0.066 (0.008) | 6.9x10<sup>-16</sup> |
| T2D | - |  |  |  |  |  |  |  | 5.2 | -0.028 (0.007) | 1.8x10<sup>-5</sup> |
| AD1 | + | rs4311 | ACE | 0.566 | 17 | 63483402 | C | T | 5.2 | 0.066 (0.008) | 6.9x10<sup>-16</sup> |
| T2D_male | - |  |  |  |  |  |  |  | 5.3 | -0.040 (0.009) | 3.0x10<sup>-6</sup> |
| AD2 | + | rs4311 | ACE | 0.566 | 17 | 63483402 | C | T | 5.1 | 0.064 (0.009) | 2.4x10<sup>-11</sup> |
| T2D_male | - |  |  |  |  |  |  |  | 5.3 | -0.040 (0.009) | 3.0x10<sup>-6</sup> |
| BMI-T2D-AD |  |  |  |  |  |  |  |  |  |  |  |
| AD1 | + | rs12325539 | C16orf92 | 0.987 | 16 | 30022312 | T | C | 6.2 | 0.057 (0.008) | 1.2x10<sup>-11</sup> |
| T2D | - |  |  |  |  |  |  |  | 6.0 | -0.041 (0.007) | 2.7x10<sup>-10</sup> |
| AD1 | + | rs11150577 | TAOK2 | 0.321 | 16 | 29975204 | G | A | 5.7 | 0.050 (0.008) | 1.4x10<sup>-9</sup> |
| BMI_female | - |  |  |  |  |  |  |  | 5.4 | -0.022 (0.002) | 2.6x10<sup>-21</sup> |
| BMI_female_over50 | - |  |  |  |  |  |  |  | 5.2 | -0.023 (0.005) | 9.0x10<sup>-6</sup> |
| T2D_female | - |  |  |  |  |  |  |  | 5.5 | -0.035 (0.010) | 2.9x10<sup>-4</sup> |
| AD1 | + | rs12325539 | C16orf92 | 0.962 | 16 | 30022312 | T | C | 6.2 | 0.057 (0.008) | 1.2x10<sup>-11</sup> |
| T2D_male | - |  |  |  |  |  |  |  | 6.0 | -0.043 (0.009) | 9.3x10<sup>-7</sup> |

**Effect direction:** aligned to the risk increasing allele for AD. **Colocalized:** SNP identified by HyPrColoc to drive the same genetic signal in both traits. **SNP:** Single nucleotide polymorphism. **PPC:** Posterior Probability of Colocalization. **Chr:** Chromosome. **Pos:** Position in hg38. **EA:** Effect allele aligned to the risk increasing allele for AD. **NEA:** Non-effect allele. **LD:** Linkage disequilibrium. **NEAF:** Non-effect allele frequency. **Beta (SE):** effect size (standard error). **P:** p-value.

To evaluate potential sex-informed cardiometabolic liability, we evaluated regional locus structures using locuszoom plots (**Fig. 3**C) and association metrics (p-value, beta) across traits (**Fig. 3**D; **Fig. S1**A-I**; Table S3**). Interestingly, the COX7C locus seems to be women and age-specific (**Fig. S1**A), exhibiting a locus peak and significant GWAS association from BMI in women (p=1.6×10^−5^) not found in BMI in men (*p*=0.13). Conversely, the ACE locus seems specific to men, showing no locus peak and weak genetic association from T2D GWAS in women (*p*=0.02), and significantly higher effect size in T2D in men versus women (p=1.7×10^−7^) (**Fig. S1**G; **Table S3**).

Effect sizes remained consistent across AD1 (includes AD-proxy cases), AD2 (excludes AD-proxy cases), and AD3 (primarily excludes AD-proxy cases and non-age-matched controls; **Fig. 3**E-G), indicating robustness of our colocalized loci to case misclassification and improper control age-matching. Overall, genetic colocalization revealed shared genetic loci between BMI/T2D and AD as well as suggestive women-predominant BMI-AD associations with age-sensitivity, while ACE T2D-AD effects were potentially specific to men.

### Phenome-wide association analyses pinpoint phenotype-specific biology

Phenome-wide association screening (n=1,314 phenotypes) of seven colocalized loci uncovered broad biological links (**Table S4**; **Methods**). BMI-AD variants consistently associated with lower blood pressure (**Fig. 4; Fig. S2**A-J), but exhibited mixed cardiometabolic associations independent of BMI effect direction: COX7C (women-specific) showed the standard obesity profile while FERMT2 resembled metabolically healthy obesity (elevated BMI, hematological inflammation markers, but lower blood and pulse pressure)^34,35^. Further, we examined whether colocalized variants associated with established AD endophenotypes (**Table S4**). We confirmed that variants within the broader APOE locus are linked to neuritic plaques and neurofibrillary tangles, CSF Aβ42 and pTau. We corroborate that all other colocalized variants associate with AD risk, but not AD biomarkers, suggesting alternative mechanisms of action linked to immune or metabolic pathways that likely precede amyloid or tau pathology.

**Fig. 4.**
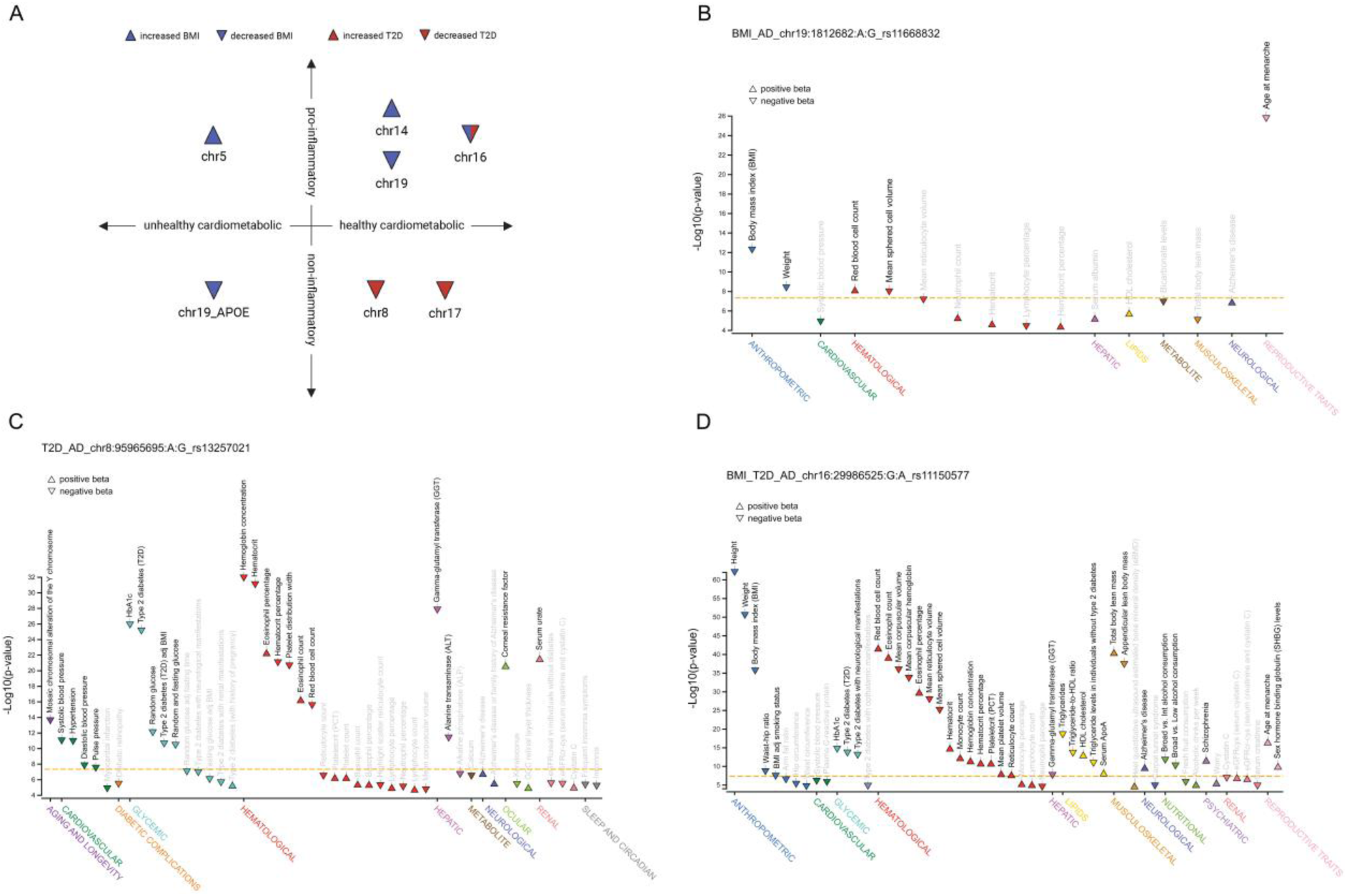
Phenome wide association study (PheWAS) analyses. (**A**) Summary of PheWAS associations of inflammatory and cardiometabolic profiles. PheWAS profile of the (**B**) rs11668832 BMI-AD locus (ATP8B3) in chr19, (**C**) rs13257021 T2D-AD locus (TP53INP1) in chr8 and (**D**) rs11150577 BMI-T2D-AD locus (TAOK2) in chr16. PheWAS p-value is set at a Bonferroni corrected threshold of 4.3×10^−5^ (orange dotted line). All PheWAS results are aligned to the risk-increasing allele for AD. Position in hg19.

Aligned to the risk increasing allele for AD, T2D-AD and BMI-T2D-AD loci associated with favorable metabolic profiles: lower glucose, HbA1c, and GGT (pro-inflammatory liver enzyme), with healthier lipid signatures and blood pressure (**Fig. 4**; **Fig. S2**A-J). Curiously, BMI loci potentially specific to women exhibited associations to SHBG and age at menarche, while the suggestive men-specific TP53INP1 locus (T2D-AD) associated with Y chromosome mosaic alterations, known to accelerate AD progression in men^36^, reinforcing our exploratory sex-specific variant-level findings.

Overall, BMI and T2D are linked to AD risk primarily through underlying cardiometabolic mechanisms, where genetically-driven hypotension and hypoglycemia associate with elevated AD risk.

### Variant-to-gene mapping identifies multi-omic functional links to effector genes

To map the colocalized loci between BMI, T2D risk, and AD risk to effector genes, we applied the combined SNP-to-gene (cS2G) approach that ranks genes based on a hierarchy of gene prioritization strategies (**Methods**).

We mapped colocalized loci to effector genes using hierarchical cS2G prioritization variants in the same credible set with the colocalized variant (**Table S5**). Supporting evidence came from nearest transcription start site annotation (closest gene)^37^, fine-mapped cis-sQTL data (54 tissues)^38^ and plasma pQTL data^39^ (**Table 3**; **Table S6**). High confidence genes were determined by a variant falling in the coding region – exon or promoter – of a gene, with other regulatory links also serving as strong evidence of the biological relevance of identified genes (**Table 3**).

**Table 3.**
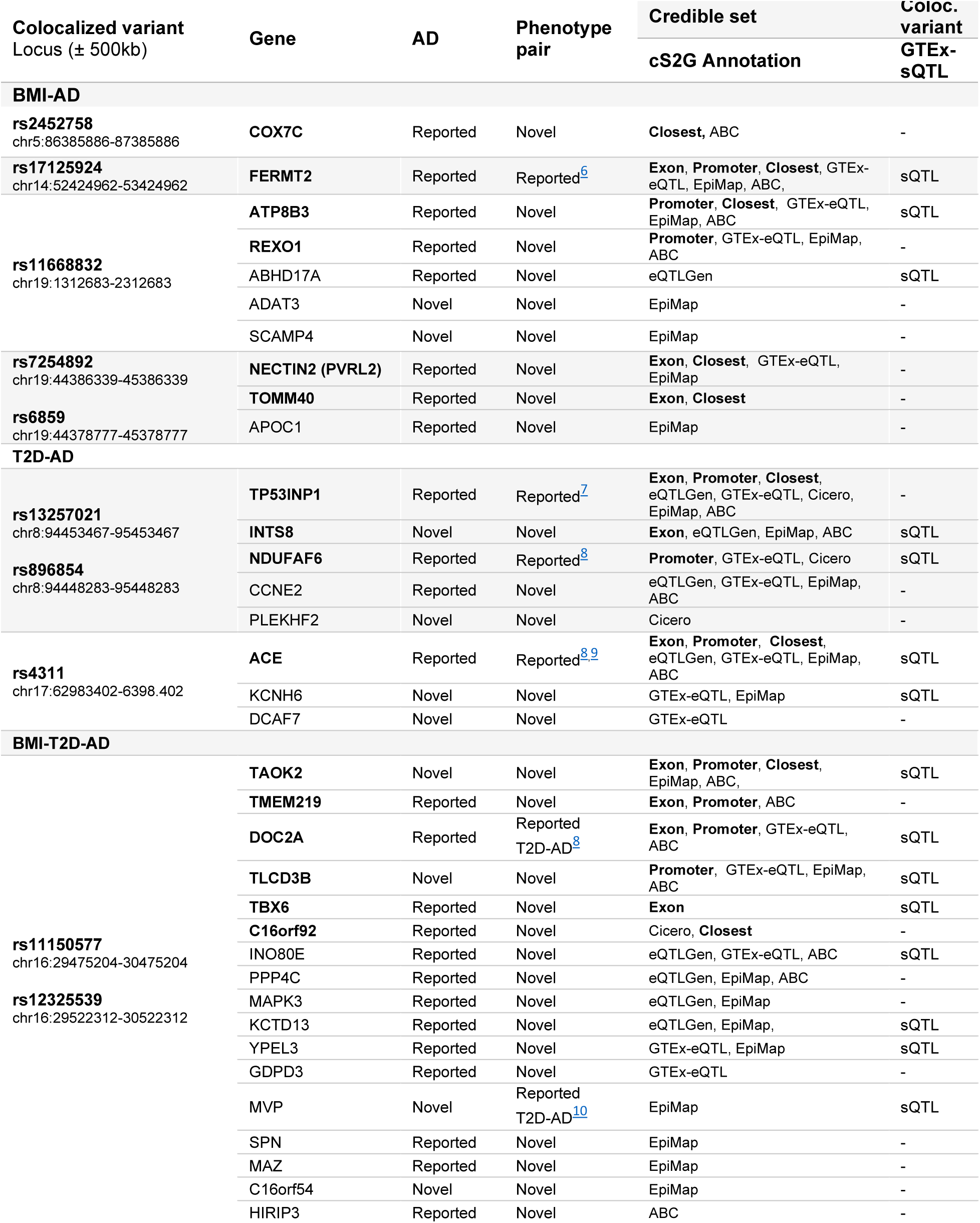
Genes with functional evidence across colocalized shared loci (all variants in the same credible set with the colocalized variant). The closest gene and high confidence genes (those with coding - exon or promoter - evidence are shown in bold. Genes in each locus are ranked by mapping confidence from cS2G (**Table S5**).

We identified 16 high confidence genes from a total of 35 unique effector genes across seven loci, of which 10 are novel genes for AD (high confidence: INTS8, TAOK2 and TLCD3B) and 29 are novel genes for phenotype pairs (high confidence: COX7C, ATP8B3, REXO1, NECTIN2, TOMM40, INTS8, TAOK2, TMEM219, TLCD3B and TBX6). We define novel hits as genes that have not been previously reported in the literature for the corresponding trait or combination of traits. To assess whether effector genes incorporate similar functional networks, we performed a protein-protein interaction (PPI) analysis (**Fig. 5; Table S7**). Protein-protein interaction (PPI) analysis (**Fig. 5**; **Table S7**) revealed that the genes in the BMI-T2D-AD locus formed a strong interaction network, suggesting shared mechanistic pathways. Most other loci lacked inter-gene interactions, indicating locus-specific biology. NECTIN2-TOMM40 (APOE) showed strong intra-locus but no cross-loci interactions, suggesting independence from APOE effects.

**Fig. 5.**
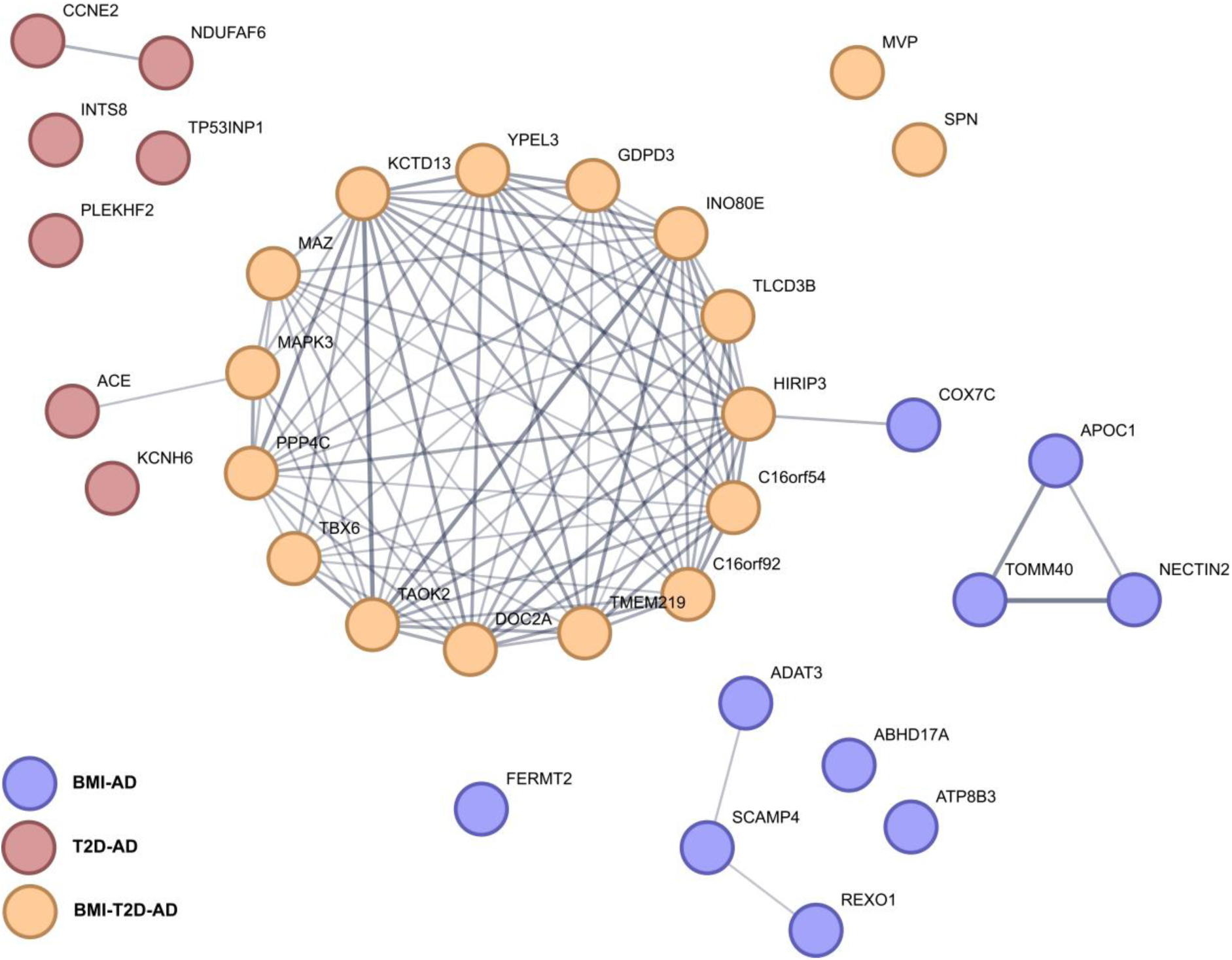
Protein-protein interaction (PPI) analysis. Genes from **Table 3** were included. Analysis performed in STRING (version 12.0), indicating both functional and physical protein associations with sufficient confidence (0.4) including data from text mining, experiments, databases, co-expression, neighborhood, gene fusion and co-occurrence. Line thickness indicates the strength of data support.

### Deep-learning and multi-omic analyses implicate peripheral biology in risk loci

We mapped tissue and cell-type associations using AlphaGenome^40^ for lead variant effects across functional genomic data-types (**Table S8**; **Fig. S3**; **Methods**), cS2G, splicing QTL (sQTL) and protein QTL (pQTL) functional links (**Table S5-S6**), and GTEx tissue enrichment^38^ via ShinyGO^41^ (**Fig. S4**; **Methods**). AlphaGenome corroborated cS2G and GTEx sQTLs, mapping risk loci primarily to peripheral immune and cardiometabolic tissues (**Fig. 6**A) with pervasive mesenchymal effects, particularly endothelial and immune cell-types (**Fig. 6**B). Notably, the suggestive women-specific COX7C locus induces strong expression changes in gonadal and reproductive tissues not predicted in other loci (**Fig. 6**C).

**Fig. 6.**
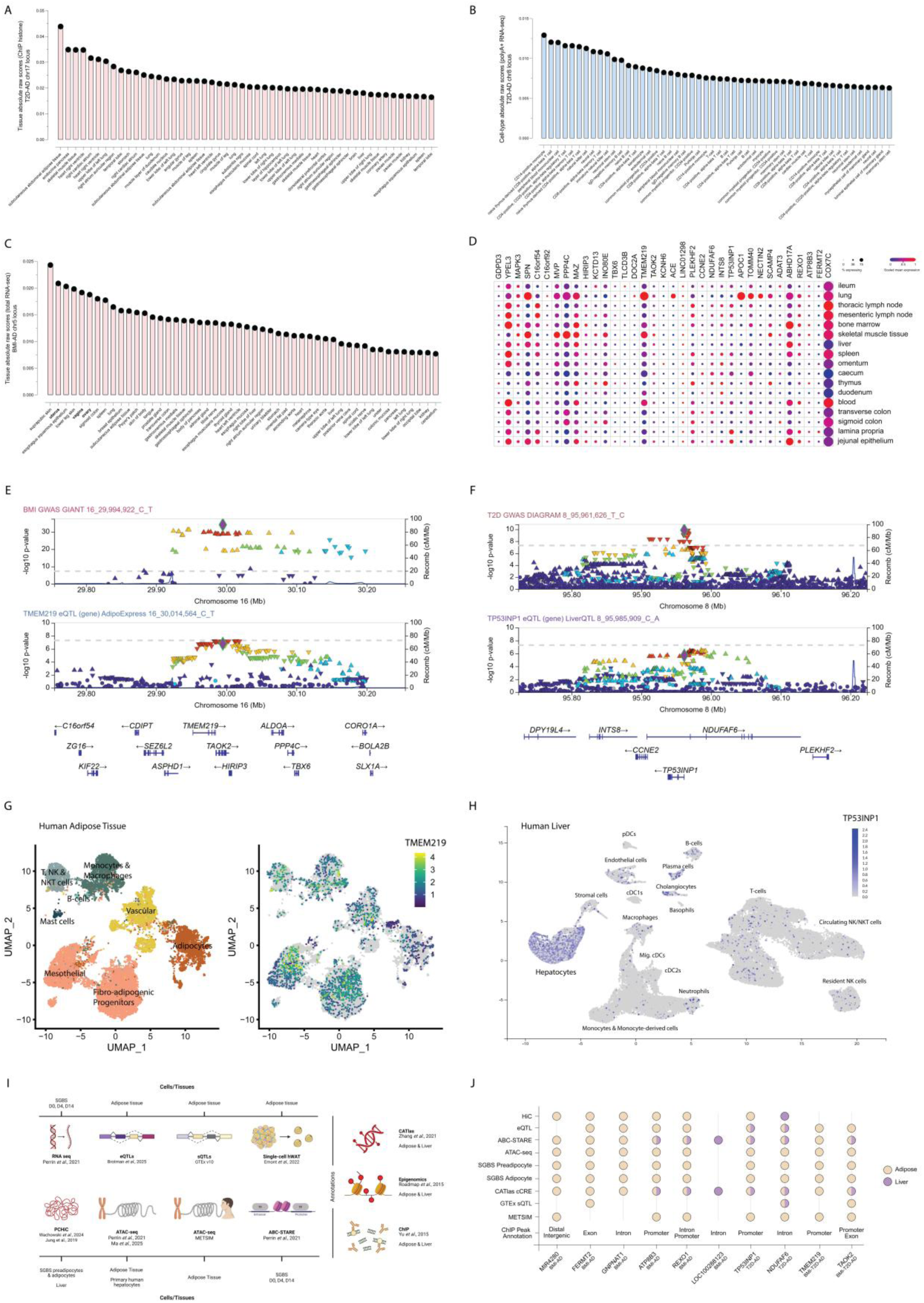
Immune and metabolic tissue association analyses. (**A**) AlphaGenome *in silico* perturbation prediction of the colocalized risk variant (rs4311) in the T2D-AD chr17 locus. The graph showcases the tissues where histone modifications (predicted from chromatin immunoprecipitation data) are most affected by perturbing the rs4311 variant. (**B**) AlphaGenome *in silico* perturbation prediction of the colocalized risk variant (rs13257021) in the T2D-AD chr8 locus. The graph showcases the cell-types where gene expression (predicted from poly(A) tail-containing RNA sequencing data) is most affected by perturbing the rs13257021 variant. (**C**) AlphaGenome *in silico* perturbation prediction of the colocalized risk variant (rs2452758) in the BMI-AD chr5 locus. The graph showcases the tissues where gene expression (predicted from total RNA sequencing data) is most affected by perturbing the rs2452758 variant. (**D**) Individual gene expression and % of expressing cells within the immune compartment of 17 peripheral tissues. (**E**) GWAS-eQTL colocalization locuszoom (LZ) region plot for BMI and TMEM219 eQTL in adipose tissue. (**F**) GWAS-eQTL colocalization LZ region plot for T2D and TP53INP1 eQTL in liver. (**G**) Human adipose tissue single-cell expression of TMEM219. (**H**) Human liver single-cell expression of TP53INP1. (**I**) Schematic overview of enhancer-to-gene (E2G) in adipose tissue and liver. (**J**) Summary associations of prioritized genes by E2G in adipose tissue and liver.

#### Peripheral immune niches

GTEx enrichment analyses implicated select loci in immune-related tissues (small intestine, colon, blood, spleen; **Fig. S4**). Single-cell transcriptomic analyses of the immune niches of 17 peripheral tissues^42^ revealed prevalent shared risk gene expression across immune cell-types (**Fig. 6**D; **Fig. S5**A-C). While not directly comparable, effector genes showed negligible central nervous system (CNS) microglia expression (**Fig. 7**B-C), the primary AD-enriched CNS cell-type^27^, supporting non-CNS immune effects. Despite no overall sex differences in immune-related expression, five genes showed significantly stronger expression in men (FDR 1%): *APOC1*, *NECTIN2*, *ADATA3*, *DOC2A*, and *ACE* (**Fig. S5**D-E; **Methods**). APOE-associated AD risk in women^21,43^ contrasts with the significant *APOC1*/*NECTIN2* enriched expression in men, suggesting that men might benefit from protective immune-associated effects from the APOE locus. In contrast, men show higher baseline ACE activity and greater ACE inhibitor response for hypertension^44^, suggesting that elevated ACE expression may contribute to specific immune signatures in men linking to hypertension-associated AD mechanisms.

**Fig. 7.**
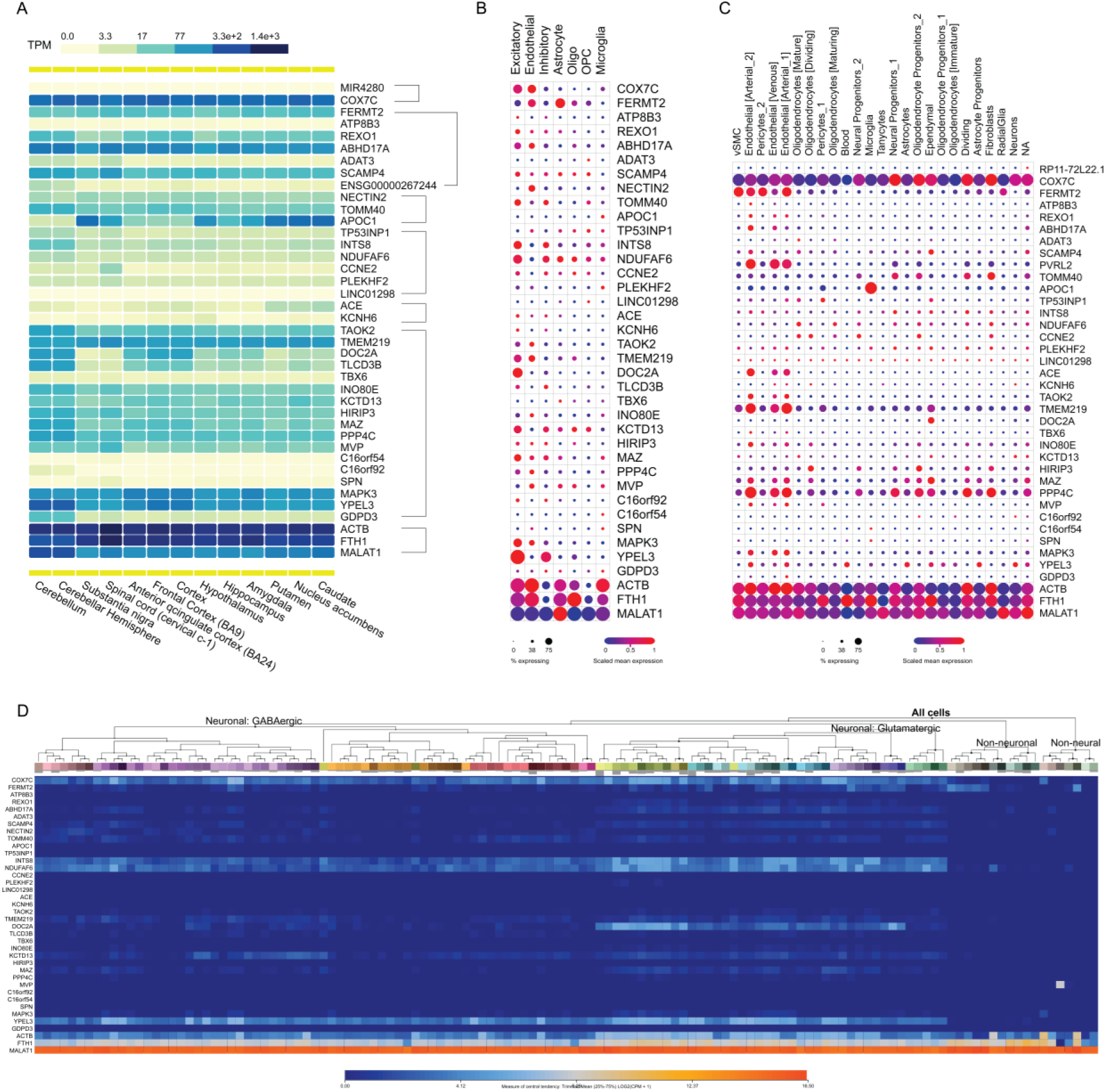
Regional and single-cell neural expression of high likelihood effector risk genes across colocalized loci. (**A**) Bulk tissue expression from multiple regions across the neurotypical brain (GTEx). (**B**) Single-cell gene expression in the neurotypical prefrontal cortex (PFC). (**C**) Single-cell gene expression in the neurotypical hypothalamus. (**D**) Single-cell gene expression in the middle temporal gyrus (MTG) of individuals with AD. ACTB, FTHI1 and MALAT1 were used as reference genes due to their high and ubiquitous expression levels across brain regions and cell-types^11^.

#### Peripheral cardiometabolic associations

GTEx enrichment extended to metabolic tissues (heart, vasculature, liver, adipose, muscle; **Fig. S4**). COLOCUS analysis evaluated colocalization between metabolic GWAS traits and eQTLs in ±100kb regions around lead variants (**Table 4**; **Fig. S6**; **Table S9**; **Methods**). Extensive colocalization emerged between metabolic GWAS traits and eQTLs in adipose tissue, liver, muscle, and pancreatic islets. We showcase BMI-TMEM219 colocalization in adipose tissue with broad cell-type expression (**Fig. 6**E, G) and T2D-TP53INP1 colocalization in liver with hepatocyte-specificity (**Fig. 6**F, H). The enhancer-to-gene (E2G) analysis^45^ corroborated cS2G associations and validated COLOCUS findings by reporting extensive links to regulatory regions in adipose tissue and hepatocytes using tissue-specific data (**Fig. 6**I; **Table S10**). E2G highlights the still uncharacterized LOC100288123 as a novel liver-specific RNA gene (**Fig. 6**J).

**Table 4.**
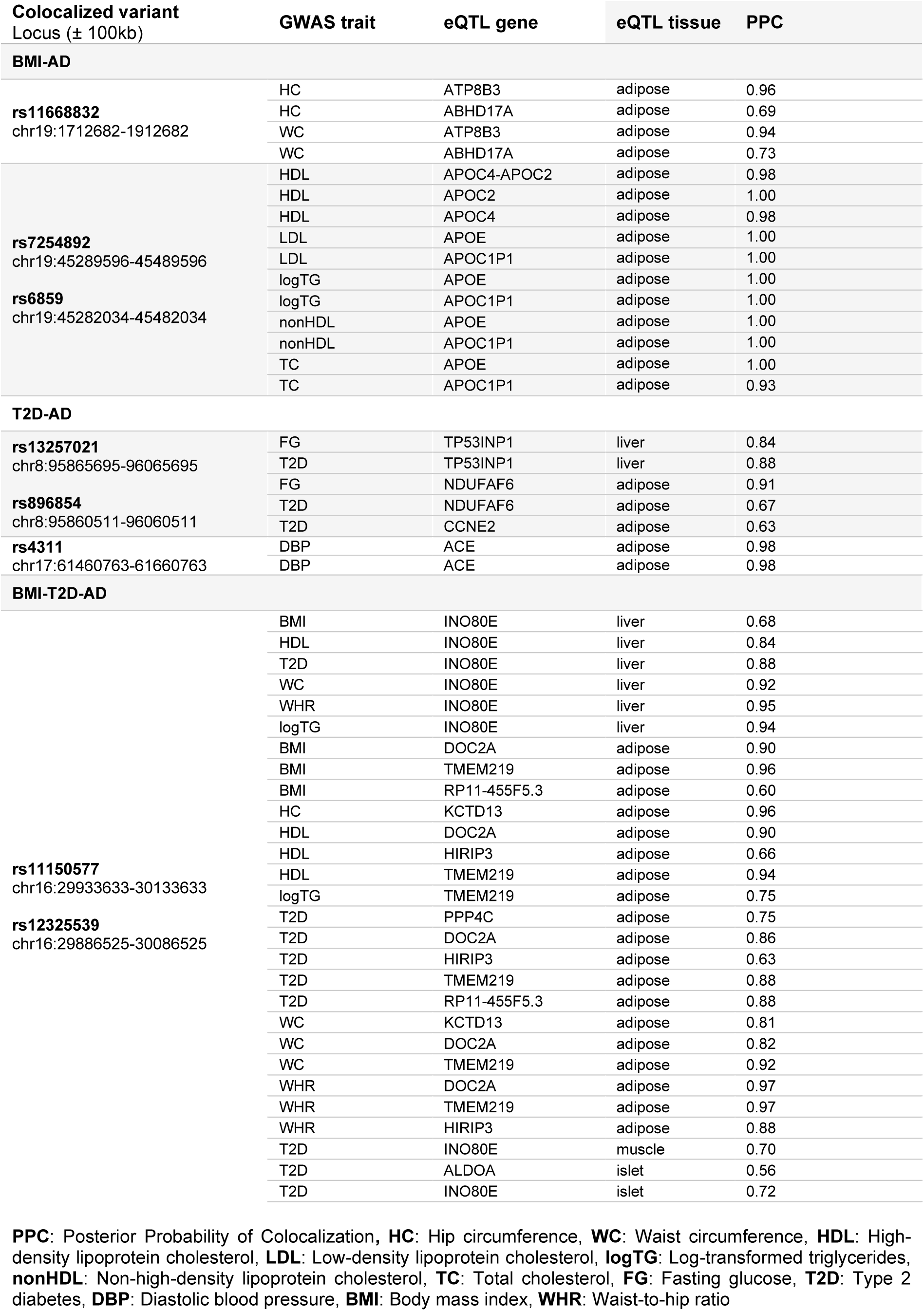
Summary table of colocalized GWAS-eQTL pairs (build 37) in metabolic tissues, including adipose, pancreatic islets, liver and muscle.

### Tissue expression and single-cell profiles reveal CNS regional differences and cell-type specificity

GTEx enrichment implicated brain regions in T2D loci with strong expression and splicing activity (**Fig. S4**, **S7**; **Table S5**). BMI-associated MIR4280 and T2D-associated MIR3150 (**Table S6**) mapped to regulatory activity in nucleus accumbens, cerebellum, cortex, and hippocampus. To examine CNS effects, we analyzed neurotypical GTEx data (13 brain regions), prefrontal cortex (PFC) and hypothalamus single-cell transcriptomics, and middle temporal gyrus (MTG) single-cell profiles from 5 AD donors (**Methods**).

Moderate bulk-level gene expression appeared primarily in cerebellum, cerebellar hemisphere and cortical regions (**Fig. 7**A). PFC excitatory neurons and hypothalamic endothelial cells were the primary expressing cell-types (**Fig. 7**B), contrasting with AD-genetic CNS microglia enrichment^27^. Regional gene expression differed substantially. ACE, an AD drug target^46^ and key driver of blood pressure regulation^47^, is primarily expressed in hypothalamic endothelial cells but not the PFC (**Fig. 7**B, C), linking the renin-angiotensin system to hypothalamic glucose homeostasis and AD risk^48^. Expected pancreatic TMEM219 expression contrasted with minimal expression in heart, vascular muscle, or retina (**Fig. S8**), implicating TMEM219 in CNS-adipose-immune axis regulation of metabolic-immune AD susceptibility. AD MTG neuronal cells expressed select risk genes (COX7C, INTS8, NDUFAF6, DOC2A, KCTD13, YPEL3; **Fig. 7**D), consistent with PFC neurotypical data (**Fig. 7**B).

Overall, CNS expression of BMI, T2D, and AD risk effector genes is limited in both neurotypical and AD brain. These results support a systemic immune-metabolic model of AD susceptibility.

### Pathway enrichment indicates novel systemic biology

Pathway enrichment (GO, KEGG, Reactome; **Fig. 8**; **Table S11**; **Methods**) and interactome analyses identified overarching immune and RNA pathways. Pervasive mechanisms included cell cycle, adhesion, autophagy, lipid metabolism, MAPK signaling, DNA/mitochondrial biology and cation channels. BMI-AD loci are uniquely involved in viral biology, T2D-AD loci in renin-angiotensin pathways, and BMI-T2D-AD loci in GTPase, glucagon signaling, and cardiac biology (**Fig. S9**A-C).

**Fig. 8.**
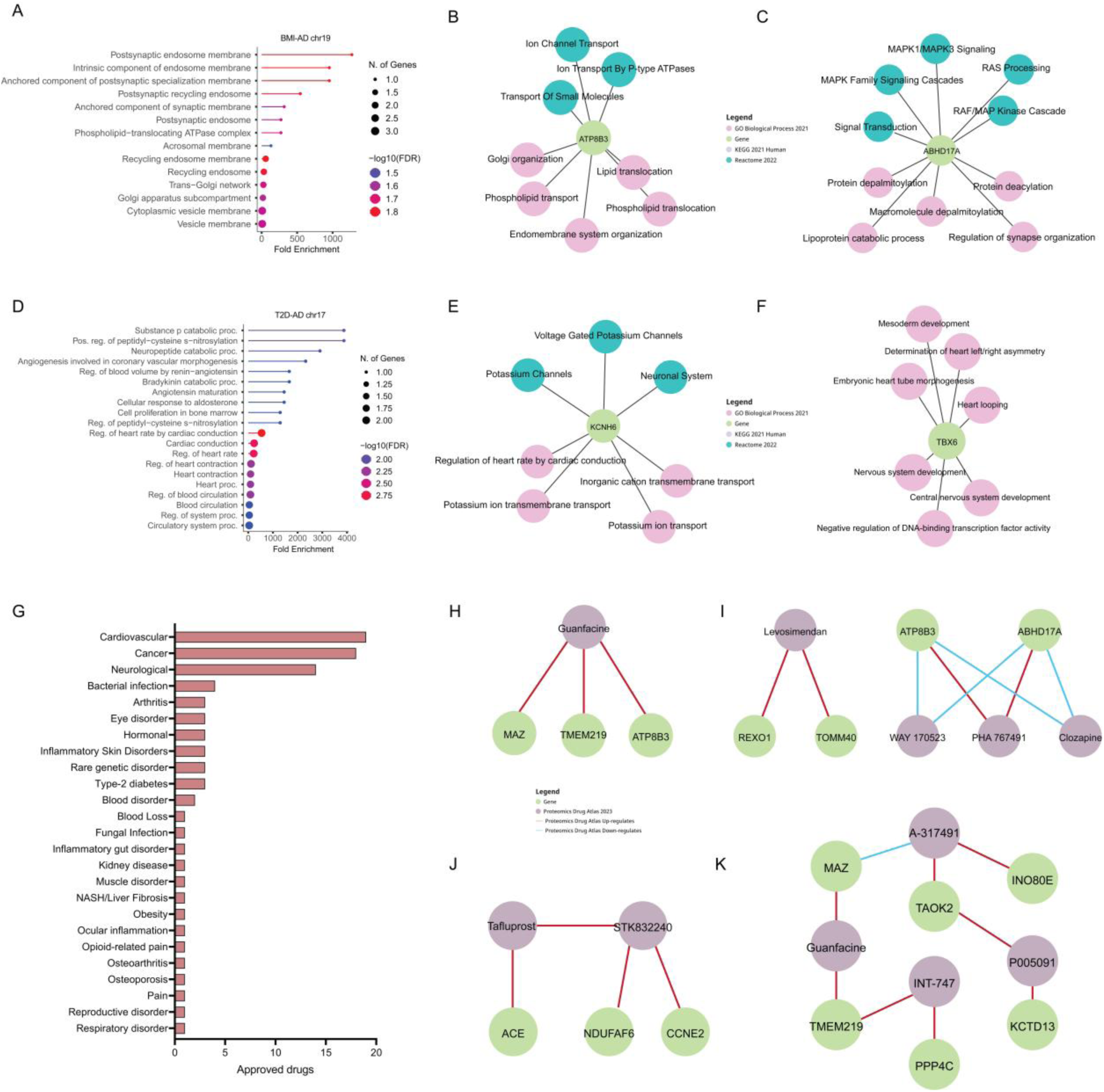
Enrichment and proteomic-based drug targeting analyses. (**A**) GO cellular components enrichment analysis for all genes with functional annotation in the BMI-AD chr19 locus. (**B**) Individual gene enrichment analysis for ATP8B3. (**C**) Individual gene enrichment analysis for ABHD17A. (**D**) GO cellular components enrichment analysis for genes with coding annotation in the T2D-AD chr17 locus. (**E**) Individual gene enrichment analysis for KCNH6. (**F**) Individual gene enrichment analysis for TBX6. (**G**) Summary of compound categories based on proteomic drug atlas hits for all effector genes. Proteomic drug atlas enrichment analysis for (**H**) all effector genes (minimum 3 links per drug target); (**I**) BMI-AD effector genes; (**J**) T2D-AD effector genes; BMI-T2D-AD effector genes. Enrichment analyses’ false discovery rate (FDR) was set at 0.05.

The ATP8B3 locus (BMI-AD) enriched for synaptic and endosomal biology (**Fig. 8**A), with ATP8B3 linked to lipid biology and ion channels (**Fig. 8**B). These findings are of particular interest given that TRPML1, a lysosomal calcium channel, has been shown to revert synaptic decline through lysosomal reacidification^49^. ABHD17A localizes to endosomes^50^ to regulate postsynaptic activity via PSD-95 depalmitoylation^51^ (calcium-NMDA-dependent^52^), modulate RAS-MAPK signaling^53^ and calcium-activated potassium channels^54^, unifying locus mechanisms. Both calcium channels^55^ and depalmitoylases^56,57^ are therapeutically interventionable targets.

The ACE locus (T2D-AD) enriched for cardiac and neuropeptide pathways (**Fig. 8**D). The same-locus effector gene *KCNH6* associates with potassium channels (**Fig. 8**E), corroborating cation channels as promising druggable targets. In line, TBX6 shows brain-heart developmental links (**Fig. 8**F), with cardiac pathways represented across multiple loci.

To link systemic and CNS biology, we analyzed secreted protein interactors (Human Protein Atlas; **Table S12**) to identify angiotensinogen (AGT, ACE interactor) and insulin-like growth factor binding protein 3 (IGFBP3, TMEM219 interactor) as potential mediators. AGT (liver/kidney-predominant) regulates renin-angiotensin-mediated blood pressure and emerges as a cardiometabolic therapeutic target^58^. IGFBP3, primarily produced in the liver, is upregulated in AD brain, interacts with humanin to prevent AD-related neuronal death^59^, and its knockdown reverses senescence^63,64^.

Overall, the broad evidence of molecular mechanisms spanning peripheral and central biological signatures support the systemic involvement of pathways mediating BMI and T2D risk in AD risk.

### Computationally nominated drug repurposing candidates are linked to preclinical development

We evaluated whether effector genes underlie ongoing therapeutic programs (**Table 5**). We identify 14 shared risk genes (high confidence: ACE, NECTIN2, TAOK2, TMEM219, ATP8B3, TP53INP1, FERMT2 and COX7C) currently targeted by ongoing therapeutic programmes, of which ACE is targeted by multiple approved drugs and phase 3 trials for cardiometabolic conditions. Additional risk genes are being targeted in preclinical development for cancer, liver fibrosis, autoimmune disease, hepatic steatosis, stroke, and mitochondrial disorders. These findings support the robustness of our multi-trait pipeline, highlighting extensive links between shared risk genes and current preclinical development.

**Table 5.** Therapeutic programmes targeting effector genes driving shared BMI, T2D and AD genetics.

| Gene | Status | Indication |
| --- | --- | --- |
| ACE (high confidence) | Multiple approved drugs<sup>12</sup><br>Phase 3<sup>13</sup> | Hypertension, cardiovascular disease, heart failure, dyslipidemia, kidney disease |
| MAPK3 | Preclinical<sup>14</sup> | Drug resistance, triple-negative breast cancer, solid tumors |
| NECTIN2 (high confidence) | Preclinical<sup>15</sup> | Ovarian cancer immunotherapy |
| TAOK2 (high confidence) | Preclinical<sup>16 17</sup> | Cancer |
| TMEM219 (high confidence) | Preclinical<sup>18 19</sup> | Liver fibrosis, gastrointestinal autoimmune disorders |
| ATP8B3 (high confidence) | Preclinical<sup>20</sup> | Colon adenocarcinoma immunotherapy |
| CCNE2 | Preclinical<sup>21 22</sup> | Breast cancer |
| ABHD17A | Preclinical<sup>23</sup> | NRAS-mutant cancers |
| GDPD3 | Preclinical<sup>24</sup> | Hepatic steatosis |
| TP53INP1 (high confidence) | Preclinical<sup>25</sup> | Castration-resistant prostate cancer |
| FERMT2 (high confidence) | Preclinical<sup>26</sup> | Prostate cancer |
| COX7C (high confidence) | Preclinical<sup>27</sup> | Stroke, mitochondrial disorders |
| MVP | Preclinical<sup>28</sup> | Pancreatic adenocarcinoma |
| PPP4C | Preclinical<sup>29</sup> | Ovarian cancer |

To identify novel targets of interest, proteomic-based drug targeting using EnrichR-KG (**Methods**) identified 81 approved drugs modulating effector gene expression (p<0.05), primarily for cardiovascular disorders (heart failure, hypertension), cancer and neurological conditions (schizophrenia, multiple sclerosis) (**Fig. 8**G; **Table S14**). We highlight [1] Guanfacine, a drug approved for ADHD and hypertension (**Fig. 8**H), [2] Clozapine, a drug approved for atypical antipsychotic and treatment-resistant schizophrenia (**Fig. 8I**), [3] Tafluprost, a prostaglandin analog approved for open angle glaucoma and ocular hypertension (**Fig. 8**J), [4] INT-747, a farnesoid X receptor agonist with preclinical indications for insulin sensitizing, hypertension, bile acid homeostasis, hepatic and intestinal inflammation, liver fibrosis, and cardiovascular disease^60–62^, and [5] A-317491, a P2X2/P2X3 antagonist with preclinical indications for neuropathic pain and chronic inflammation^63^ (**Fig. 8**K). Resmetirom, a thyroid hormone receptor-β agonist recently approved for NASH/liver fibrosis^64^, **Table S14**) modulates the novel gene SCAMP4, an endocytic trafficking protein. Remarkably, Levosimendan, a calcium sensitizer^65^ approved for heart failure (**Fig. 8**I), presents direct experimental evidence of inhibiting disulfide tau oligomerization, ameliorating tau pathology and rescuing cognitive decline in the AD-relevant Tau^P301L^-BiFC mouse model^66^.

Our findings demonstrate the potential of genetics-driven drug target prioritization for functional validation, and how drug mechanisms can accelerate the discovery of potentially novel disease biology.

## DISCUSSION

Obesity and type 2 diabetes are major modifiable risk factors for AD, and their shared genetic architecture has the potential to unveil actionable biology relevant to the long prodromal curve of AD. We identify loci associated with BMI/T2D and AD, with suggestive sex and age-specific associations. Besides brain involvement, peripheral immune-metabolic tissues and cell-types emerge as primary effectors of shared genetic risk mechanisms.

Our approach primarily reflects the genetic risk linked to variations in BMI and the pathobiology of T2D in AD development. Further, the exploratory sex-informed findings pertain to the sex-specificity of the cardiometabolic genetic architecture that overlaps with AD. All BMI-AD loci reported here are novel and exhibit suggestive sex- and age-specific effects in women. The broader APOE locus has been linked to T2D-AD^67^, but not directly to BMI-AD. We report a novel TP53INP1 lead variant (rs12546365) and male-specific ACE effects, expanding prior T2D-AD findings^68,67,69^. Further, we identify TAOK2-C16orf92 as the first jointly associated locus with BMI, T2D, and AD.

The integration of epidemiological and genetic findings within a broader life-course interpretation of cardiometabolic genetic risk in AD is remarkably nuanced. The relationship between cardiometabolic traits and AD risk is not static across the lifespan, but rather follows an age-dependent trajectory. Epidemiological evidence has long established that midlife obesity substantially elevates dementia risk^70–73^, while late-life obesity paradoxically associated with lower risk of incident dementia^70,74,75^. However, epidemiological observations of cardiometabolic risk in AD have not translated to clinical efficacy, as evidenced by the evoke and evoke+ trials^76^. Obesity at age 50, but not ages 60 or 70, are linked to higher dementia risk^70^. This midlife window coincides with the onset of asymptomatic amyloid accumulation, particularly in women^77^, which precedes clinical AD by up to two decades. Our exploratory use of sex-stratified BMI data (≤50 versus >50) was designed to capture genetic signals operating within this critical window, which we provide compelling evidence that the shared genetic architecture of BMI in women and AD is concentrated in the period preceding and surrounding menopausal transition. The late-life inversion of the BMI-AD risk profile, whereby lower BMI in older adults associates with higher dementia risk, is best understood as a composite of reverse causation from AD that distort observational estimates. This is supported by reverse mendelian randomization evidence showing that silent AD-related biological changes start reshaping body composition around age 50^78,79^.

We find that risk variants linked to lower BMI, and lower risk of T2D are generally linked to higher AD risk, in line with recent clinical evidence that compared people with underweight, normal weight and obesity to the risk of Aβ positivity^80^. Genetics capture lifelong risk, and potentially mechanism linked to earlier life or chronic additive exposure, and can dissect disease reverse causation, unlike epidemiological studies that often capture the risk profile of comorbidities or risk factors late in adulthood and early in preclinical AD. A critical consideration of our work is the potential for survival bias to confound results at the summary statistics data level. Individuals who develop AD may be depleted of genetic variants that predisposes to higher cardiometabolic risk. These individuals might be selectively removed from the population before the age of AD onset, thus enriching AD GWAS for protective cardiometabolic alleles^16^. This could obscure causal relationships and hampers the use of causal inference methods, as apparent protective associations could reflect selective survival rather than true biological antagonism. Our exclusion of AD-proxy cases and non-age-matched controls (AD3), as well as the sex- and age-stratified pipeline, were design to minimize this potential bias. The persistence and sensitivity of sex- and age-specific genetic correlations across AD datasets, but also at the individual variant level, support their biological authenticity rather than artefactual origin. Still, the lack of necessary information to fully adjust for these biases at the aggregated summary statistics level could be further mitigated using individual-level data.

The disconnect between epidemiological and genetic evidence might be partially explained by the use of BMI as the proxy-trait for obesity. We find that BMI-AD loci effects are independent of BMI direction but rather driven by underlying cardiometabolic associations. Genetically-driven low blood pressure and low glucose levels link to elevated AD risk, directly opposing the hypothesis motivating trials of GLP1R agonists for AD, aligning with observed lack of clinical efficacy in the evoke and evoke+ trials despite key biomarker engagement^76^. This potentially suggests that our computational approach managed to filter spurious shared loci whose biology might be confounded by multiple biological and/or environmental factors. Cross-sectional and longitudinal studies substantiate an increased prevalence of AD and dementia in older individuals with low blood pressure^81–83^, but with higher blood pressure in younger individuals^84^. Antihypertensive clinical trials are inconclusive^85^ and only recently genetic evidence was linking hypertension to AD in multivariate Mendelian randomization studies^16^ whereas multi-trait GWAS failed to identify strong links,^86^. Likewise, our findings that low glucose levels link to elevated AD risk align with AD patients showing strong glucose hypometabolism and insulin resistance in the brain^87,88^. As increased fasting plasma glucose levels directly associate with higher brain glucose concentrations^89^, our results suggest that lower peripheral glucose levels may potentiate long term cerebral energy deficits.

Cardiometabolic risk research in AD has primarily focused on T2D and AD, but our findings show that BMI-AD biology is more specific to AD while T2D-AD is more ubiquitous, in line with recent observations^8,90^. Further, we find suggestive evidence that women are at higher risk from cardiometabolic outcomes associated with variations in BMI and men from cardiometabolic outcomes associated with T2D, in line with recent longitudinal findings^91^. These observations align with higher prevalence of obesity in women^23^ and higher susceptibility to AD^17^. Our findings further indicate that women suffer more severe consequences associated with BMI variations driven by APOE, yet current approved drugs are less effective in women and APOE4 carriers^20^. Consistently, T2D and Parkinson’s disease (PD) share a stronger correlation compared to T2D and AD^92^, with PD being substantially more common in men^19^. For example, women show superior insulin sensitivity compared to men, but insulin resistance is not a relevant phenotype in our loci. On the other hand, clinical trials targeting the male-specific ACE locus (T2D-AD) show that men have significantly higher baseline plasma ACE activity than women and benefit more when taking ACE inhibitors for hypertension^44^. These considerations should prompt the field to examine sex-stratified data for clinical trials targeting genes or pathways evidencing sex-dysmorphic profiles.

Deciphering the genetic architecture and underlying biology of AD has been limited to the brain and its constituent cell-types. While T2D-related loci show brain associations, peripheral immune and metabolic tissues, not brain or microglia, primarily drive BMI and T2D-associated AD risk. This aligns with recent findings that APOE4 drives immune-related proteomic signatures enriched in circulating immune cells, directly linking AD’s primary genetic risk factor to peripheral immunity^93^.

Human genetic evidence boosts the likelihood of drugs’ clinical success by 2.6-fold^94^, with GWAS variants pointing to highly successful drug targets^95^. We identify multiple genetically-backed drug targets with vast repurposing and novel development potential. Even drug targets unrelated to the input cardiometabolic phenotypes are experiencing a substantial repurposing push for neurological disorders, exemplified by immunomodulatory cancer drugs like Lenalidomide currently in clinical trial for AD^96^. Importantly, while we observe a strong proportion of computationally-nominated targets linked to ongoing preclinical development, functional studies are needed to establish causal evidence.

Limitations of our work include the current unavailability of sex-specific AD data and small sample sizes of sex-stratified BMI and T2D data, that derive sex-specific inferences about AD risk indirectly and may underestimate the sex-specificity of the cardiometabolic genetic determinants overlapping with AD. Upcoming AD meta-analyses are set to publish sex-stratified AD genotyping data, though both studies face significant sample size limitations that would render their applicability in multi-trait pipelines quite limited^97,98^. Moreover, HyPrColoc stringency ensures reliability but may miss other true shared genetic signals. Also, assessing genome-wide significance alone may miss key insights for AD, exemplified by genetic variation linked to GLP1R driving obesity therapeutics but reaching BMI significance only in upcoming GWAS (5M+, unpublished). Future analyses incorporating more refined adiposity measures such as waist-to-hip ratio, waist-to-hip ratio adjusted for BMI or body fat percentage would be valuable extensions of our work, where methods that implicitly model shared genetic architecture could provide substantial complementary value. Importantly, the generalizability of our results might be limited outside European populations given the current sample size constraints in non-European large scale GWAS data for BMI, T2D and AD for multi-trait frameworks at comparable statistical power. Future work should prioritize the multi-ancestry discovery and replication of our findings. Further, our findings present strong biological inferences for experimental validation. Thus, future studies should focus on the experimental validation of our findings through perturbation analyses across relevant *in vitro* and animal models to dissect how peripheral AD risk mechanisms drive neuropathology.

Our findings enable informed therapeutic targeting of the shared genetic blueprint of BMI, T2D, and cardiometabolic-driven AD risk. Blood pressure interventions warrant more refined sex, age-range and disease stage eligibility criteria. Given prior focus on T2D-AD biology, BMI-related traits may yield outcomes more targeted to AD. The identification of peripheral tissues, not brain, as primary effectors of cardiometabolic risk in AD opens novel research directions for AD risk mitigation via genetically-backed candidates.

## METHODS

### GWAS summary statistics curation

All BMI and T2D summary statistics (sex-combined and sex-stratified) were originally sourced in the human reference genome GRCh37 (HG19) while the AD1, AD2 and AD3 summary statistics were made available in the human reference genome GRCh38 (HG38). Given that conversion rates are higher from HG19 to HG38^99^, we proceeded to liftover the BMI and T2D GWAS data from HG19 to HG38 using the “rtracklayer” R statistical software^100^ (version 4.2.2) package from the Bioconductor project^101^. BMI, T2D, AD2 and AD3 summary statistics required reference SNP cluster ID (rsID) annotation. Altogether, a total of ≈52.5 million chromosome positions were annotated using the “BSgenome” R statistical software (version 4.2.2) package from the Bioconductor project^102^. The APOE locus was retained in all analyses, reflecting a methodological consideration specific to our study design. The three AD meta-analyses used here (AD1, AD2, AD3) differ systematically in case ascertainment and control matching strategies. Regional APOE exclusion would result in unpredictable interactions with these pre-existing differences. Further, we aimed to evaluate sex-specific differences in cardiometabolic risk associated with the APOE region given the differential drug responses for women and APOE4 carriers^20^. To address potential APOE dominance concerns, we report: [1] full transparency of APOE findings (NECTIN2-TOMM40 locus), including novel BMI-AD associations with female-specific effects; and [2] protein-protein interaction analyses to assess non-APOE loci independence from APOE biology.

### Heritability and genetic correlation

We leveraged LD score regression^103^ (LDSC) to estimate narrow-sense heritability (additive genetic effects of common variants) and genetic correlation (GC) between AD, BMI and T2D datasets. We leveraged precalculated LD scores calculated in individuals of European ancestry from the 1000 Genomes Project (*eur_w_ld_chr*). Standard GC data curation and quality control procedures were followed^103,104^. In the secondary analysis, because sex-stratified summary statistics were used for BMI and T2D but not for AD, the analyses quantify the genetic overlap between sex-specific cardiometabolic genetic effects and population-average (weighted average of male and female) AD genetic effects. Further, we tested the differences between trait-pair genetic correlation estimates using:

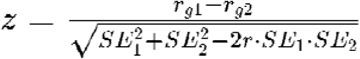

where rg is the genetic correlation for each trait-pair, SE are the standard errors, and r is the genetic correlation coefficient estimate between the different traits in both trait-pairs^101^. The genetic correlation coefficient r between non-AD traits accounts for their underlying genetic similarity. This parameter is key for controlling whether observed differences in genetic correlation estimates (rg) reflect true significant biological differences rather than shared genetic architecture of the traits themselves. For example, when comparing rg estimate differences between BMI.Male-AD1 and BMI.Female-AD1, r represents the genetic correlation between BMI.Male and BMI.Female, effectively quantifying their genetic similarity and correcting for it in statistical testing. We assumed a Bonferroni corrected p-value threshold of (0.05/27=1.85×10^−3^).

Importantly, the use of LDSC promotes literature generalization, but known LDSC limitations include the assumption of a single LD reference panel, potential sensitivity to extended LD regions (including APOE) and reduced performance when applied to meta-analytic GWAS derived from linear mixed models.

### Variant discovery

We conducted genetic overlap analyses using Python^105^ (version 3.9.13) between AD, BMI and T2D datasets in HG38. We defined the significance thresholds (*p*-value) of 5×10^−8^ for AD datasets and 5×10^−4^ for all BMI and T2D datasets to identify independent overlapping genetic variants following LD-clumping (*ld_clump()* function (r^2^=0.1; window=500kb) from the R package *MRCIEU/ieugwasr* based on the PLINK clumping method^106^ and 1000 Genomes EUR Phase 3 reference panel^107^). We performed phenotype overlap based on chromosome positions. This initial p-value thresholding of shared variants was performed to focus the colocalization analyses on high probability genomic regions to avoid statistical inflation. Further, the use of a less stringent p-value threshold for BMI and T2D datasets allowed us to keep methodological consistency across primary (sex-combined) and secondary (sex-stratified) analyses, where sample sizes are considerably smaller in sex-stratified and age-stratified BMI and T2D datasets. This methodological approach did not lead to false positive findings, as all colocalized loci, except for the ACE locus, exhibit genome-wide significant associations with sex-combined BMI/T2D despite the initial lower p-value thresholding (**Table 2**). The only colocalized variant failing to reach genome-wide significance for the risk factor – the ACE locus for T2D – is one of the most established shared risk loci in the literature with vast functional and therapeutic evidence in both AD and T2D. Further, our p-value thresholding approach was the most appropriate for the secondary sex-stratified pipeline, where comparable effect sizes between sex-combined and sex-stratified datasets for BMI and T2D confirm that their p-values fail to reach genome-wide significance simply driven by smaller sample sizes. The use of asymmetric significance thresholds reflects the distinct roles these traits play in the etiology of AD. The biological expectation that variants affecting intermediate, exposure or risk-factor phenotypes (BMI, T2D) through additive cascade mechanisms may exert smaller individual effects than those affecting disease endpoints (AD), and therefore may not reach genome-wide significance in individual cross-section GWAS but remain relevant in shared genetic architecture. The GLP1R locus exemplifies this principle: variants linked to GLP1R reach genome-wide significance for T2D^108^, but more modest associations in BMI^25^. Yet, therapeutic GLP1R agonists produce substantial body weight reduction^109^, demonstrating that variants below genome-wide significance for intermediate or risk-factor phenotypes can have substantial biological and therapeutic relevance. For AD, we applied the standard genome-wide significance threshold to minimize the propagation of systematic biases inherent to the use of AD-proxy phenotypes in all contemporary GWAS^16,110^.

We performed colocalization analysis on overlapping variants using HyPrColoc, a Bayesian method for identifying shared genetic associations between traits within genomic regions. Colocalization regions were defined as ±500kb windows around pre-filtered independent overlapped variants from the initial genetic overlap analysis. HyPrColoc analysis was conducted using GWAS summary statistics, including regression coefficients and standard errors for each variant and trait. The method tests whether multiple traits share causal variants in a genomic region and, when evidence for full colocalization is insufficient, employs a clustering algorithm to identify subsets of traits that colocalize at distinct causal variants. Default parameters were used for the analysis. Importantly, we did not pre-specify any posterior probability threshold in order to [1] keep methodological consistency across primary (sex-combined) and secondary (sex-stratified) analyses and [2] account for the possibility of 3 or more traits colocalizing where the joint probability of a spurious three-way colocalization is substantially lower than any single trait-pair, with the multi-trait colocalization calculation itself acting as a stringency filter. Moreover, given the smaller sample sizes in the sex-stratified pipeline, a predefined posterior probability threshold would fail to identify true signals that ended up confirmed as true colocalized genetic signals. Of note, none of the two-trait colocalized signals had a posterior probability of colocalization (PPC) under the standard of 0.5. The TAOK2 signal is the only one with a PPC lower than 0.5 due to it being a 4-trait colocalization hit, and is in high LD (R^2^=0.77) with C16orf92 (rs12325539) with a PPC of 0.987 in the same locus. To minimize the risk of false positives, we [1] performed sensitivity analyses to assess the robustness of identified colocalization clusters and [2] validated candidate colocalization signals through visual inspection of local Locuszoom plots to confirm signal, possible LD contamination and regional architecture.

All analyses were performed using the HyPrColoc R package^111^. The HLA region (chr6:28,510,120-33,480,577) was removed post-analysis. Locuszoom plots were generated using a package that creates regional Manhattan plots leveraging an european linkage disequilibrium reference panel (https://github.com/Geeketics/LocusZooms). We tested the differences between women-specific and men-specific beta-estimates (b_women_, b_men_) using:

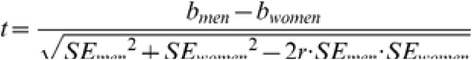

where SE are the standard errors and r is Spearman rank correlation coefficient across all SNPs across BMI and T2D strata. We assumed a Bonferroni corrected p-value threshold of (0.05/12=4.2×10^−3^).

### Phenome-wide association analysis

Phenome-wide association studies (PheWAS) were conducted using the Association to Function Knowledge Portal (A2F; a2fkp.org) to systematically test genetic variants for associations across a broad spectrum of phenotypes. We analyzed a total of 1,314 distinct phenotypes, encompassing diseases, disorders, and clinical traits derived from electronic health records and biobank data. We screened PheWAS associations for the seven colocalized loci aligned to the risk increasing allele for AD. To account for multiple testing, a statistical significance was determined using a Bonferroni-corrected p-value threshold of 4.3×10^−5^ (0.05/1,314 phenotypes). Results meeting the corrected significance threshold were considered evidence for phenome-wide significant associations. We examined whether colocalized genetic risk variants were associated with established AD endophenotypes by leveraging the GenomicsDB NIAGADS database^112^ and ADNI’s blood-based biomarker GWAS^113^.

### Variant-to-gene mapping

For establishing connections between genetic variants and their corresponding target genes, we utilized the cS2G methodology^114^. The analysis was conducted using the GRCh37/hg37 genome assembly. The cS2G approach integrates various SNP-to-gene association methods by incorporating functional annotations derived from seven component strategies, listed according to their assigned weights: Exon regions, Promoter sequences, fine-mapped cis-eQTL data from eQTLGen blood studies, fine-mapped cis-eQTL information from GTEx across 54 human tissue types, enhancer-gene associations from EpiMap spanning 833 cellular types, Activity-By-Contact (ABC) methodology covering 167 cellular types, and Cicero analysis for blood/basal conditions. The cS2G framework computes an integrated association score by applying weights to individual strategy scores, thereby identifying the most probable effector gene(s) for each single nucleotide polymorphism (SNP). We highlight the highest confidence genes for variants falling into coding exonic or promoter regions (**Table 3**). In the absence of coding evidence, the closest gene was considered the highest confidence link.

### AlphaGenome

We performed an in-silico perturbation screen using batch variant scoring from AlphaGenome^40^ to predict the functional consequences of colocalized BMI/T2D-AD variants across multiple functional domains. The effect of each colocalized variants was predicted within a 100kb genome window in humans. We used “UBERON” and “CL” ontologies to predict variant effects on up to 116 tissues and 106 cell-types, respectively. The colocalized variants within the APOE region were not considered in this analysis given their overwhelming predicted effects across all data-types. Importantly, neither data-type in AlphaGenome included central nervous system microglia. For each variant, we report the raw scores representing the predicted regulatory effect for each data-type. Raw scores are specific to individual data-types, thus not comparable between different them. We present variant effects by ordering raw scores on magnitude of effect from highest to lowest, thus identifying the tissues and cell-types where predicted variant impacts are strongest. The raw scores are shown as absolute values due to current limitations of deep neural network models in accurately predicting the direction of variant effects^115^. To contextualize the magnitude of variant effects, we report quantile scores which provide a percentile-based ranking of variant effects relative to the background genome (**Table S8**). The quantile score is computed by establishing an empirical background distribution using scores from common variants (minor allele frequency > 0.01 in any GnomAD v3 population) for each data-type. For example, a variant with a quantile score of 0.99 indicates that its predicted regulatory effects in a given tissue or cell-type ranks at the 99^th^ percentile among common variants. Variant effect predictions are evaluated across the following functional scorers:

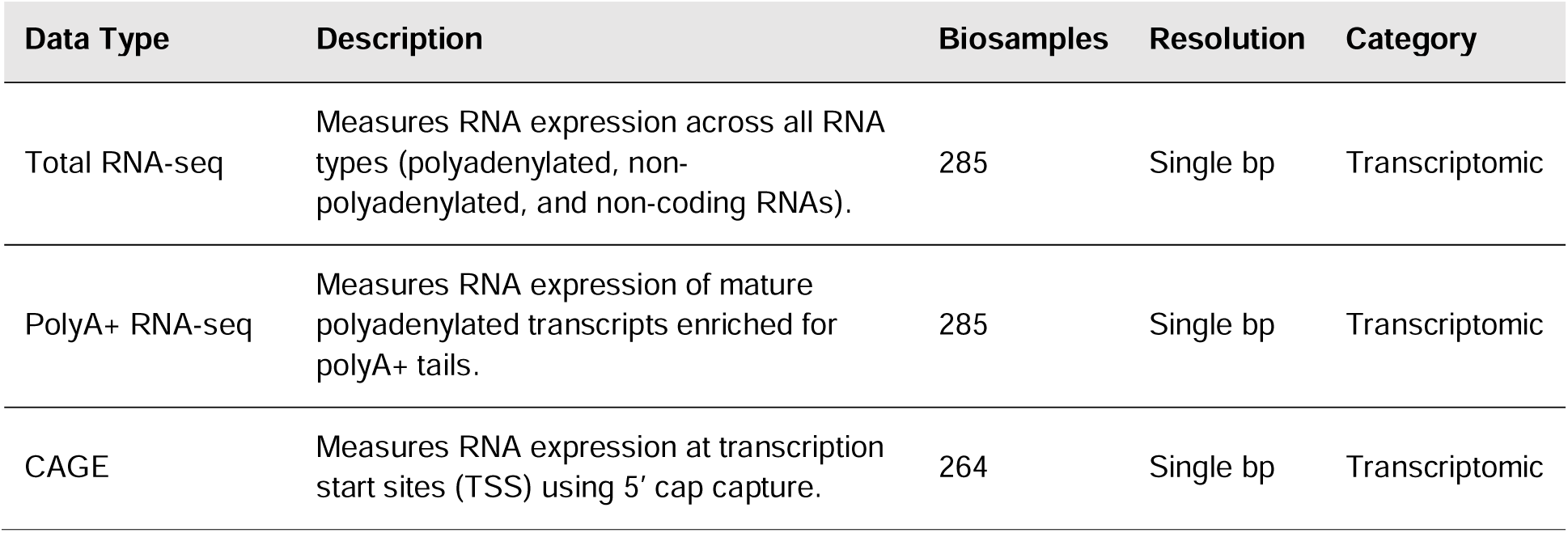
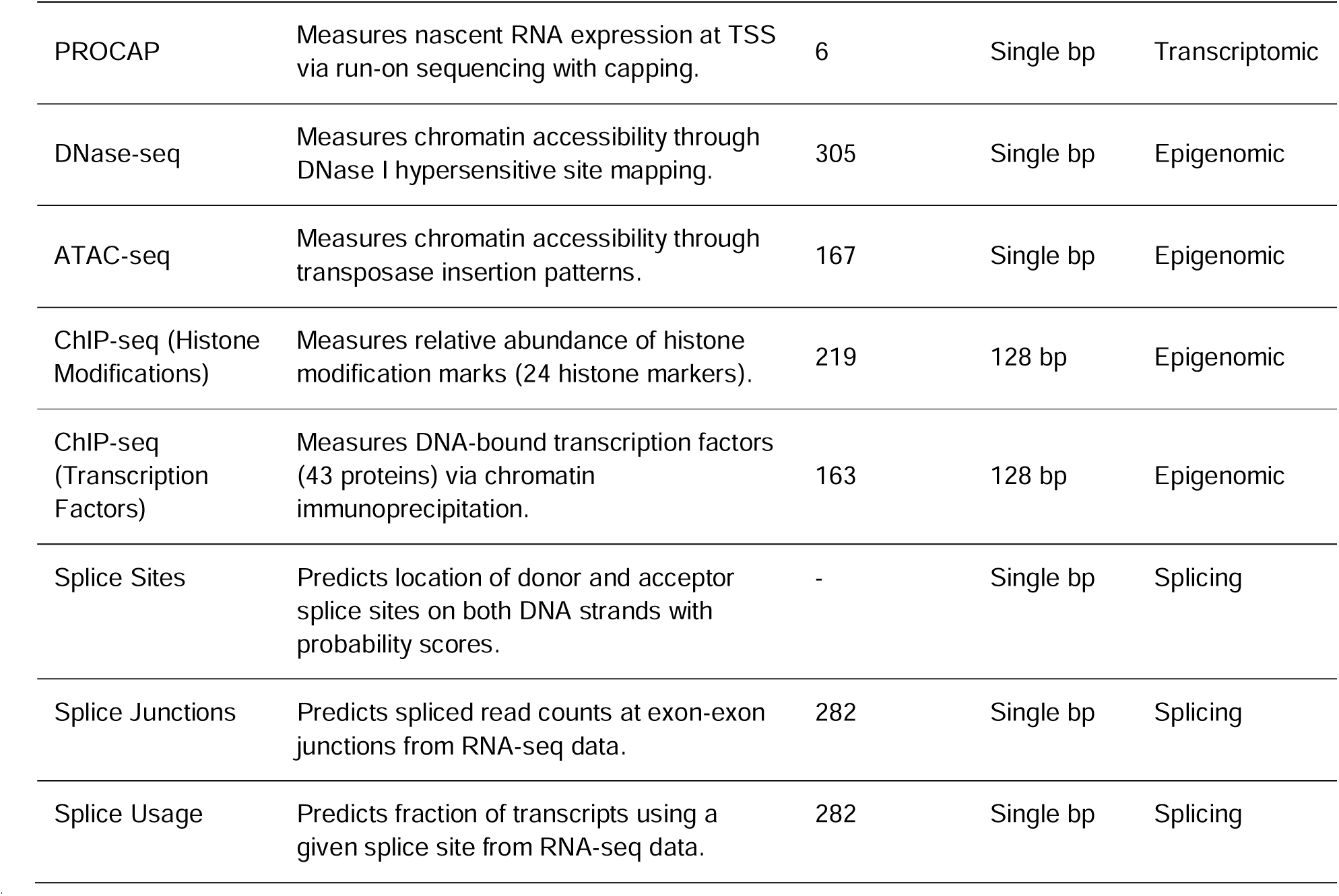

### GWAS-eQTL colocalization

We colocalized the lead cross-phenotype colocalized variants (from HyPrColoc) plus its credible sets (± 100kb) with eQTLs in metabolic tissues, including adipose, pancreatic islets, liver and muscle (build 37) using COLOCUS, a platform from HuGeAMP. COLOCUS implements the COLOC Bayesian algorithm, utilizing conditionally independent or fine-mapped signals identified through statistical fine-mapping approaches, with each signal represented by a lead variant and corresponding credible set. The GWAS traits analyzed in the eQTL colocalization analyses were selected based on the most relevant traits identified through the PheWAS analysis. The metabolic tissues were chosen based on tissue enrichment results from GTEx and variant effect predictions from AlphaGenome. Both required sufficient data availability for the analysis.

### Immune, adipose and liver analyses

We evaluated the individual gene expression and % of cells in the immune compartment of 17 peripheral tissues using the Single Cell Portal^116^, including ileum, lung, thoracic lymph node, mesenteric lymph node, bone marrow, skeletal muscle tissue, liver, spleen, omentum, caecum, thymus, duodenum, blood, transverse colon, sigmoid colon, lamina propria and jejunal epithelium^42^. Statistical sex-strata differences were evaluated using a Wilcoxon matched-pairs signed rank test (nonparametric) following the evaluation of data normality using the Anderson-Darling, D’Agostino-Pearson omnibus, Shapiro-Wilk and Kolmogorov-Smirnov normality tests. Outliers in **Fig. S5**D-E were identified using the Robust Regression and Outlier removal Using Nonlinear TukeyBiweight (ROUT) method (Q = 1%) implemented in GraphPad Prism (version 9.0.0). This method applies robust regression with a false discovery rate (FDR) of 1% to identify data points as outliers. We checked the single-cell expression in the human adipose tissue using the adipose tissues knowledge portal^117^ and the single-cell expression in the human liver^118^ using the liver cell atlas.

### Enhancer-to-gene

We adapted the E2G pipeline from Perrin *et al*.^45^ to map shared risk variants falling within regulatory genomic regions of human adipose tissue, liver, adipocytes and hepatocytes. The analysis was conducted in hg37. For the adipose tissue and adipocytes, we leveraged promoter capture HiC data from SGBS cells (both pre- and mature adipocytes)^119^, RNA-seq, ATAC-seq and ABC-STARE data from SGBS cells (days 0, 4 and 14)^45^, human adipose tissue ATAC-seq data from METSIM (10,197 Finnish men)^120^ and eQTL data from AdipoExpress (2344 human subcutaneous adipose tissue samples)^121^. We further supplemented E2G with peak annotations from ChIP using the ChIPseeker R package^122^, Cis-regulatory element annotations from single-cell epigenetic datasets using the Cis-element Atlas (CATlas)^123^, and Roadmap epigenomics data from samples E023 (mesenchymal stem cell (MSC) derived adipocyte cultured cells) and E063 (adipose nuclei)^124^. E023 derives from *in vitro* differentiated MSCs into adipocytes and models adipocyte development. E063 consists of nuclei isolated directly from adult adipose tissue and reflects the native epigenetic state of *in vivo* mature adipocytes. For the liver and hepatocytes, we leveraged promoter capture HiC data from the liver^125^, RNA-seq and ATAC-seq data from primary human hepatocytes^126^ and eQTL data from the most recent liver eQTL meta-analysis^127^. We further implemented ATAC peak annotations^122^ using the *annotatr* and *annotatePeak* R packages and Roadmap epigenomics data from liver samples. We complemented E2G with e/sQTL data from GTEx^128^ (version 10).

### Brain analysis

We performed single-cell analyses using the Single Cell Portal^105^ in the prefrontal cortex^118^ (PFC, key AD region^129^) and hypothalamus^119^ (key region in obesity, T2D and metabolism^48,130^). We used the Allen Brain Atlas^120^ to perform exploratory analyses of the single-cell diversity of the middle temporal gyrus of individuals with AD (MTG 10x SEA-AD; vulnerable AD brain region^131^) (166,868 total nuclei from 5 post-mortem human AD brain specimens, median age of 80)^121^.

### Enrichment analysis

We performed enrichment analyses in GO Biological Processes, GO Cellular Components, GO Molecular Functions, KEGG and GTEx tissues^38^ using ShinyGO^41^ (v0.85) and an FRD cutoff of 0.05. The GTEx tissue gene expression profiles dataset contains gene expression data from 17,382 RNA-seq samples of 54 different human tissues (plus 2 cell lines) from 838 postmortem donors. We performed enrichment analyses based on the “Co.Expression GTEx.Tissue Expression.Down,”, “Co.Expression GTEx.Tissue Expression.Up.” and “Co-expression.GTEx.Tissues.V8.2023” categories. This analysis evaluates whether shared risk genes are enriched within genes showing elevated or decreased expression in specific tissues. Additionally, we performed an over-representation analysis based on GO terms, KEGG, Reactome and Proteomic Drug Atlas using EnrichR-KG^132^. Of note, the proteomic drug targeting was originally performed on human immortalized cell lines^133^.

## Supporting information

EADB Supplementary Information

## Data Availability

All data produced in the present work are contained in the manuscript

https://www.ebi.ac.uk/gwas/studies/GCST90027158

https://www.ebi.ac.uk/gwas/publications/30239722

https://www.ebi.ac.uk/gwas/publications/35551307

## EADB ACKNOWLEDGMENTS

The EADB consortium groups together 20,464 Alzheimer’s disease (AD) cases and 22,244 controls after quality controls from 16 European countries (Austria, Belgium, Bulgaria, Czech Republic, Denmark, Finland, France, Germany, Greece, Italy, Portugal, Spain, Sweden, Switzerland, The Netherlands and the UK). These samples were genotyped in three independent centers (France, Germany and the Netherlands) leading to define three nodes: EADB-France, EADB-Germany and EADB-Netherlands. In addition, EADB also included Australian partners. We thank the numerous participants, researchers, and staff from many studies who collected and contributed to the data. We thank the high performance computing service of the University of Lille. We thank all the CEA-CNRGH staff who contributed to sample preparation and genotyping for their excellent technical assistance. This work was supported by a grant (European Alzheimer DNA BioBank, EADB) from the EU Joint Programme – Neurodegenerative Disease Research (JPND). Inserm UMR1167 is also funded by Inserm, Institut Pasteur de Lille, the Lille Métropole Communauté Urbaine, the French government’s LABEX DISTALZ program (development of innovative strategies for a transdisciplinary approach to Alzheimer’s disease). The Bulgarian cohort was partially supported by the European Union-NextGenerationEU, through the National Recovery and Resilience Plan of the Republic of Bulgaria, project BG-RRP-2.004-0004-C01, Strategic research and innovation program for development of Medical university - Sofia.

## SUPPLEMENTARY AUTHOR INFORMATION

Céline Bellenguez**^1^**, Benjamin Grenier-Boley**^1^**, Sami Heikkinen**^2^**, Pablo García-González**^3-5^**, Atahualapa Castillo-Morales**^6^**, Fahri Küçükali**^7,8^**, Najaf Amin**^9^**, Itziar de Rojas**^3,4,10^**, Victoria Alvarez**^11,12^**, Beatrice Arosio**^13,14^**, Jesper Qvist Thomassen**^15^**, Sverre Bergh**^16,17^**, Merce Boada**^3,4^**, Barbara Borroni**^18,19^**, Paola Bossù**^20^**, María J. Bullido**^4,21,22^**, Ángel Carracedo**^23,24^**, Antonio Daniele**^25,26^**, Alexandre de Mendonça**^27^**, Jürgen Deckert**^28^**, Martin Dichgans**^29-31^**, Srdjan Djurovic**^32,33^**, Oriol Dols-Icardo**^4,34^**, Carole Dufouil**^35^**, Emrah Düzel**^36,37^**, Valentina Escott-Price**^38^**, Tormod Fladby**^39,40^**, Laura Fratiglioni**^41,42^**, Daniela Galimberti**^43,44^**, Jose Maria García-Alberca**^4,45^**, Guillermo Garcia-Ribas**^46^**, Vilmantas Giedraitis**^47^**, Caroline Graff**^41,48^**, Timo Grimmer**^49^**, Edna Grünblatt**^50-52^**, OIivier Hanon**^53^**, Lucrezia Hausner**^54^**, Stefanie Heilmann-Heimbach**^55^**, Jakub Hort**^56,57^**, Frank Jessen**^58-60^**, Ville Leinonen**^61,62^**, Anssi Lipponen**^2^**, Jiao Luo**^15, 63^**, Henna Martiskainen**^2^**, Carlo Masullo**^64^**, Patrizia Mecocci**^42,65^**, Shima Mehrabian**^66^**, Pablo Mir**^4,67,68^**, Susanne Moebus**^69^**, Laura Molina Porcel**^70,71^**, Fermin Moreno**^4,72,73^**, Benedetta Nacmias**^74,75^**, Lucilla Parnetti**^76^**, Pau Pastor**^77,78^**, Jordi Pérez-Tur**^4,79,80^**, Oliver Peters**^81,82^**, Yolande A.L. Pijnenburg**^83,84^**, Gerard Piñol-Ripoll**^85,86^**, Julius Popp**^87-90^**, Innocenzo Rainero**^91^**, Luis M Real**^92,93^**, Steffi Riedel-Heller**^94^**, Eloy Rodriguez-Rodriguez**^4,95^**, Arvid Rongve**^96^**, Giacomina Rossi**^97^**, Jose Luis Royo**^98^**, Dan Rujescu**^99,100^**, Ingvild Saltvedt**^101^**, María Eugenia Sáez**^102^**, Raquel Sánchez-Valle**^103^**, Florentino Sanchez-Garcia**^104^**, Nicolai Sandau**^15^**, Nikolaos Scarmeas**^105,106^**, Katja Scheffler**^107,108^**, Norbert Scherbaum**^109^**, Anja Schneider**^59,110^**, Geir Selbæk**^16,111,112^**, Davide Seripa**^113^**, Vincenzo Solfrizzi**^114,115^**, Marco Spallazzi**^116^**, Alessio Squassina**^117^**, Eystein Stordal**^118^**, Niccoló Tesi**^84,119,120^**, Betty Tijms**^83,84^**, Latchezar Traykov**^66^**, Lucio Tremolizzo**^121,122^**, Kumar P. Tripathi**^123,124^**, Julie Williams**^6,125^**, Jens Wiltfang**^126-128^**, Dag Aarsland**^129-131^**, Philippe Amouyel**^1^**, Stéphanie Debette**^35^**, Gael Nicolas**^132^**, Sven van der Lee**^83,84,133^**, Henne Holstege**^83,84,119,134,135^**, Maria Victoria Fernandez**^3^**, Magda Toslaki**^136^**, Patrick Gavin Kehoe**^137^**, Kristel Sleegers**^7,8^**, Martin Ingelsson**^47,138,139^**, Roberta Ghidoni**^19^**, Ole A Andreassen**^33,140,141^**, Peter A. Holmans**^125^**, Pascual Sánchez-Juan**^4,142^**, Rebecca Sims**^125^**, Alfredo Ramirez**^59,60,110,123,143^**, Agustin Ruiz**^3,4,144^**, Mikko Hiltunen**^2^**, Cornelia van Duijn**^9,145^**, Ruth Frikke-Schmidt**^15,146^**, Jean-Charles Lambert**^1^**

^1.^ Université de Lille, Inserm, CHU Lille, Institut Pasteur de Lille, Lille, France.

^2.^ Institute of Biomedicine, University of Eastern Finland, Yliopistoranta 1E, 70211 Kuopio, Finland

^3.^ Ace Alzheimer Center Barcelona, Universitat Internacional de Catalunya (UIC), Barcelona, Spain

^4.^ Networking Research Center on Neurodegenerative Diseases (CIBERNED), Instituto de Salud Carlos III, Madrid, Spain

^5.^ Doctorate in Biotechnology, Faculty of Pharmacy and Food Sciences, University of Barcelona, Barcelona, Spain.

^6.^ UK Dementia Research Institute, School of Medicine, Cardiff University, Wales, UK

^7.^ VIB Center for Molecular Neurology, VIB, Antwerp, Belgium

^8.^ Department of Biomedical Sciences, University of Antwerp, Antwerp, Belgium

^9.^ Nuffield Department of Population Health, University of Oxford, Oxford, UK

^10.^ Luxembourg Centre for Systems Biomedicine (LCSB), University of Luxembourg, Esch-sur-Alzette, Luxembourg

^11.^ Laboratorio de Genética. Hospital Universitario Central de Asturias, Oviedo, Spain

^12.^ Instituto de Investigación Sanitaria del Principado de Asturias (ISPA)

^13.^ Department of Clinical Sciences and Community Health, University of Milan, Milan, Italy

^14.^ Geriatric Unit, Fondazione IRCCS Ca’ Granda Ospedale Maggiore Policlinico, Milan, Italy

^15.^ Department of Clinical Biochemistry, Copenhagen University Hospital - Rigshospitalet, Blegdamsvej 9, 2100 Copenhagen, Denmark

^16.^ The Research Centre for Age-related Functional Decline and Disease, Innlandet Hospital Trust, Ottestad, Norway

^17.^ Norwegian National Centre for Aging and Health, Vestfold Health Trust, Tønsberg, Norway

^18.^ Department of Clinical and Experimental Sciences, University of Brescia, Italy

^19.^ Molecular Markers Laboratory, IRCCS Istituto Centro San Giovanni di Dio Fatebenefratelli, Brescia, Italy

^20.^ Laboratory of Experimental Neuropsychobiology, Clinical Neuroscience and Neurorehabilitation Department, IRCCS Santa Lucia Foundation, Rome, Italy

^21.^ Centro de Biología Molecular Severo Ochoa (UAM-CSIC)

^22.^ Instituto de Investigacion Sanitaria ‘Hospital la Paz’ (IdIPaz), Madrid, Spain

^23.^ Grupo de Medicina Xenómica, CIBERER, CIMUS. Universidade de Santiago de Compostela, Santiago de Compostela, Spain.

^24.^ Fundación Pública Galega de Medicina Xenómica-IDIS, Santiago de Compostela, Spain.

^25.^ Department of Neuroscience, Università Cattolica del Sacro Cuore, Rome, Italy

^26.^ Neurology Unit, IRCCS Fondazione Policlinico Universitario A. Gemelli, Rome, Italy

^27.^ Faculty of Medicine, University of Lisbon, Portugal

^28.^ Department of Psychiatry, Psychosomatics and Psychotherapy, Center of Mental Health, University Hospital of Würzburg, Germany

^29.^ Institute for Stroke and Dementia Research (ISD), University Hospital, LMU Munich, Munich, Germany.

^30.^ German Center for Neurodegenerative Diseases (DZNE), Munich, Germany

^31.^ Munich Cluster for Systems Neurology (SyNergy), Munich, Germany

^32.^ Department of Medical Genetics, Oslo University Hospital, Oslo, Norway

^33.^ Centre for Precision Psychiatry, Institute of Clinical Medicine, University of Oslo, Oslo, Norway

^34.^ Department of Neurology, Institut de Recerca Sant Pau, Hospital de la Santa Creu i Sant Pau, Universitat Autònoma de Barcelona, Barcelona, Spain.

^35.^ University of Bordeaux, Inserm Bordeaux Population Health Research Center U1219, Bordeaux, France.

^36.^ German Center for Neurodegenerative Diseases (DZNE), Magdeburg, Germany

^37.^ Institute of Cognitive Neurology and Dementia Research (IKND), Otto-von-Guericke University, Magdeburg, Germany

^38.^ UKDRI@ Cardiff, School of Medicine, Cardiff University, Cardiff, UK

^39.^ Institute of Clinical Medicine, Campus Ahus, University of Oslo, Oslo, Norway

^40.^ Department of Neurology, Akershus University Hospital, Lørenskog, Norway

^41.^ Karolinska University Hospital, Theme inflammation and Aging, Sweden

^42.^ Karolinska Institutet, Center for Alzheimer Research, Department NVS, Division of ARC, Stockholm, Sweden

^43.^ Neurodegenerative Diseases Unit, Fondazione IRCCS Ca’ Granda, Ospedale Policlinico, Milan, Italy

^44.^ Dept. of Biomedical, Surgical and Dental Sciences, University of Milan, Milan, Italy

^45.^ Alzheimer Research Center & Memory Clinic, Andalusian Institute for Neuroscience, Málaga, Spain.

^46.^ Hospital Universitario Ramon y Cajal, IRYCIS, Madrid

^47.^ Department of Public Health and Caring Sciences, Clinical Geriatrics, Uppsala University, Uppsala, Sweden

^48.^ Karolinska Institutet, Center for Alzheimer Research, Department NVS, Division of Neurogeriatrics, Bioclinicum J10:20, 171 64 Stockholm Sweden

^49.^ Center for Cognitive Disorders, Department of Psychiatry and Psychotherapy, Technical University of Munich, School of Medicine, Munich, Germany

^50.^ Department of Child and Adolescent Psychiatry and Psychotherapy, University Hospital of Psychiatry Zurich, University of Zurich, Zurich, Switzerland

^51.^ Neuroscience Center Zurich, University of Zurich and ETH Zurich, Switzerland

^52.^ Zurich Center for Integrative Human Physiology, University of Zurich, Switzerland

^53.^ Université Paris Cité, EA 4468, APHP, Hospital Broca, Memory Resource and Research Centre of de Paris-Broca-Ile de France, Paris, France

^54.^ Department of Geriatric Psychiatry, Central Institute for Mental Health Mannheim, Faculty Mannheim, University of Heidelberg, Germany

^55.^ Institute of Human Genetics, University of Bonn, School of Medicine & University Hospital Bonn, Bonn, Germany

^56.^ Memory Clinic, Department of Neurology, Charles University, Second Faculty of Medicine and Motol University Hospital, Czech Republic

^57.^ International Clinical Research Center, St. Anne’s University Hospital Brno, Brno, Czech Republic

^58.^ Department of Psychiatry and Psychotherapy, Faculty of Medicine and University Hospital Cologne, University of Cologne, Cologne, Germany

^59.^ German Center for Neurodegenerative Diseases (DZNE), Bonn, Germany

^60.^ Cluster of Excellence Cellular Stress Responses in Aging-associated Diseases (CECAD), University of Cologne, Cologne, Germany

^61.^ Department of Neurosurgery, Kuopio University Hospital, Puijonlaaksontie 2 FI-70210 Kuopio, Finland

^62.^ Institute of Clinical Medicine-Neurosurgery, University of Eastern Finland, Kuopio, Finland

^63.^ Novo Nordisk Foundation Center for Basic Metabolic Research, Faculty of Health and Medical Science, University of Copenhagen, Blegdamsvej 3, 2200 Copenhagen

^64.^ Institute of Neurology, Catholic University of the Sacred Heart, Rome, Italy

^65.^ Division of Gerontology and Geriatrics, Department of Medicine and Surgery, University of Perugia, Italy

^66.^ Clinic of Neurology, UH “Alexandrovska”, Medical University - Sofia, Sofia, Bulgaria

^1.^ 67. Unidad de Trastornos del Movimiento, Servicio de Neurología y Neurofisiología. Instituto de Biomedicina de Sevilla (IBiS), Hospital Universitario Virgen del Rocío/CSIC/Universidad de Sevilla, Seville, Spain

^68.^ Departamento de Medicina, Facultad de Medicina, Universidad de Sevilla, Seville, Spain

^69.^ Institute for Urban Public Health, University Hospital of University Duisburg-Essen, Essen, Germany.

^70.^ Neurological Tissue Bank - Biobanc-Hospital Clinic-IDIBAPS, Barcelona, Spain

^71.^ Alzheimer’s disease and other cognitive disorders Unit. Neurology Department, Hospital Clinic, Barcelona, Spain

^72.^ Department of Neurology. Hospital Universitario Donostia. San Sebastian, Spain

^73.^ Neurosciences Area. Instituto Biodonostia. San Sebastian, Spain

^74.^ Department of Neuroscience, Psychology, Drug Research and Child Health University of Florence, Florence, Italy

^75.^ IRCCS Fondazione Don Carlo Gnocchi, Florence, Italy

^76.^ Centre for Memory Disturbances, Lab of Clinical Neurochemistry, Section of Neurology, University of Perugia, Perugia, Italy

^77.^ Fundació Docència i Recerca MútuaTerrassa, Terrassa, Barcelona, Spain

^78.^ Memory Disorders Unit, Department of Neurology, Hospital Universitari Mutua de Terrassa, Terrassa, Barcelona, Spain

^79.^ Unitat de Genètica Molecular, Institut de Biomedicina de València-CSIC, Valencia, Spain

^80.^ Unidad Mixta de Neurologia Genètica, Instituto de Investigación Sanitaria La Fe, Valencia, Spain.

^81.^ German Center for Neurodegenerative Diseases (DZNE), Berlin, Germany

^82.^ Charité – Universitätsmedizin Berlin, corporate member of Freie Universität Berlin, Humboldt-Universität zu Berlin, and Berlin Institute of Health, Institute of Psychiatry and Psychotherapy, Hindenburgdamm 30, 12203 Berlin, Germany

^83.^ Alzheimer Center Amsterdam, Neurology, Vrije Universiteit Amsterdam, Amsterdam UMC location VUmc, Amsterdam, The Netherlands

^84.^ Amsterdam Neuroscience, Neurodegeneration, Amsterdam, The Netherlands

^85.^ Unitat Trastorns Cognitius, Hospital Universitari Santa Maria de Lleida, Lleida, Spain

^86.^ Institut de Recerca Biomedica de Lleida (IRBLLeida), Lleida, Spain

^87.^ Old Age Psychiatry, Department of Psychiatry, Lausanne University Hospital, Lausanne, Switzerland

^88.^ Department of Geriatric Psychiatry, University Hospital of Psychiatry Zürich, Zürich, Switzerland

^89.^ Institute for Regenerative Medicine, University of Zürich, Switzerland

^90.^ Center for memory disorders and Alzheimer’s disease, Klinik für Neurologie Hirslanden, Zürich, CH

^91.^ Department of Neuroscience “Rita Levi Montalcini”, University of Torino, Torino, Italy

^92.^ Centro de Investigación Biomédica en Red de Enfermedades Infecciosas (CIBERINFEC), Instituto de Salud Carlos III, Madrid, Spain

^93.^ Instituto de Biomedicina de Sevilla, IBiS/Hospital Universitario Virgen de Valme /CSIC/ Departamento de Bioquímica Médica, Biología Molecular e Inmunología, Universidad de Sevilla, Sevilla, Spain

^94.^ Institute of Social Medicine, Occupational Health and Public Health, University of Leipzig, 04103 Leipzig, Germany.

^95.^ Neurology Service, Marqués de Valdecilla University Hospital (University of Cantabria and IDIVAL), Santander, Spain.

^96.^ Haugesund Hospital, Department of Research and Innovation, Haugesund, Norway

^97.^ Unit of Neurology V - Neuropathology, Fondazione IRCCS Istituto Neurologico Carlo Besta, Milan, Italy

^98.^ Depatamento de Especialidades Quirúrgicas, Bioquímica e Inmunología. Facultad de Medicina. Universidad de Málaga. Málaga, Spain

^99.^ Department of Psychiatry and Psychotherapy, Medical University Vienna, Vienna, Austria

^100.^ Martin-Luther-University Halle-Wittenberg, University Clinic and Outpatient Clinic for Psychiatry, Psychotherapy and Psychosomatics, Halle (Saale), Germany

^101.^ Department of Neuromedicine and Movement Science, Faculty of Medicine and Health Sciences, Norwegian University of Science and Technology (NTNU), Department of Geriatrics, St. Olav’s Hospital, Trondheim University Hospital, Trondheim, Norway

^102.^ CAEBI, Centro Andaluz de Estudios Bioinformáticos, Sevilla, Spain.

^103.^ Alzheimer’s disease and other cognitive disorders unit. Service of Neurology. Hospital Clínic of Barcelona. Institut d’Investigacions Biomèdiques August Pi i Sunyer, University of Barcelona, Barcelona, Spain

^104.^ Department of Immunology, Hospital Universitario Doctor Negrín, Las Palmas de Gran Canaria, Las Palmas, Spain.

^105.^ Taub Institute for Research in Alzheimer’s Disease and the Aging Brain, The Gertrude H. Sergievsky Center, Depatment of Neurology, Columbia University, New York, NY

^106.^ 1st Department of Neurology, Aiginition Hospital, National and Kapodistrian University of Athens, Medical School, Greece

^107.^ Department of Neurology and Clinical Neurophysiology, Trondheim University Hospital, Trondheim, Norway

^108.^ Department of Neuromedicine and Movement Science, Norwegian University of Science and Technology (NTNU), Trondheim, Norway

^109.^ Department of Psychiatry and Psychotherapy, LVR-Klinikum Essen, University of Duisburg-Essen, Germany Medical Faculty, Germany

^110.^ Department for Cognitive Disorders and Old Age Psychiatry, University Hospital Bonn, Bonn, Germany.

^111.^ Norwegian National Advisory Unit on Aging and Health, Vestfold Health Trust, Tønsberg, Norway

^112.^ Institute of Health and Society, Faculty of Medicine, University of Oslo, Oslo, Norway

^113.^ Department of Hematology and Stem Cell Transplant, Vito Fazzi Hospital, Lecce, Italy

^114.^ Interdisciplinary Department of Medicine, Geriatric Medicine and Memory Unit, University of Bari “A. Moro, Bari, Italy

^115.^ Academic Division “C. Frugoni” & Hospital Division of Internal and Geriatric Medicine, Policlinico Hospital, Bari, Italy

^116.^ Department of Medicine and Surgery, Unit of Neurology, University-Hospital of Parma, Parma, Italy

^117.^ Department of Biomedical Sciences, Section of Neuroscience and Clinical Pharmacology, University of Cagliari, Cagliari, Italy

^118.^ Namsos Hospital, Namsos, Norway

^119.^ Genomics of Neurodegenerative Diseases and Aging, Human Genetics, Vrije Universiteit Amsterdam, Amsterdam UMC location VUmc, Amsterdam, The Netherlands

^120.^ Delft Bioinformatics Lab, Delft University of Technology, Delft, The Netherlands

^121.^ Neurology Unit, IRCCS “San Gerardo dei Tintori”, Monza, Italy

^122.^ School of Medicine and Surgery, University of Milano-Bicocca, Monza, Italy

^123.^ Division of Neurogenetics and Molecular Psychiatry, Department of Psychiatry and Psychotherapy, Faculty of Medicine and University Hospital Cologne, University of Cologne, Cologne, Germany

^124.^ Estudios en Neurociencias y Sistemas Complejos (ENyS) CONICET-HEC-UNAJ

^125.^ Division of Psychological Medicine and Clinical Neuroscience, School of Medicine, Cardiff University, Wales, UK

^126.^ Department of Psychiatry and Psychotherapy, University Medical Center Goettingen, Goettingen, Germany

^127.^ German Center for Neurodegenerative Diseases (DZNE), Goettingen, Germany

^128.^ Medical Science Department, iBiMED, Aveiro, Portugal

^129.^ Department of Old Age Psychiatry, Division of Academic Psychiatry, Institute of Psychiatry, Psychology and Neuroscience, King’s College London, United Kingdom

^130.^ Wolfson Centre for Age Related Diseases, Division of Neuroscience of the Institute of Psychiatry, Psychology and Neuroscience, King’s College London, United Kingdom

^131.^ Stavanger University Hospital, Stavanger, Norway

^132.^ Univ Rouen Normandie, Normandie Univ, Inserm U1245 and CHU Rouen, Department of Genetics and Reference Center for Developmental Disorders, Rouen, France.

^133.^ Genomics of Neurodegenerative Diseases and Aging, Human Genetics, Vrije Universiteit Amsterdam, Amsterdam UMC location VUmc, Amsterdam, The Netherlands.

^134.^ VIB Center for Brain and Disease Research, ON5 Herestraat 49 - box 602, 3000 Leuven, Belgium

^135.^ Department of Neurosciences, Leuven Brain Institute, KU Leuven, Leuven, Belgium.

^136.^ Aristotle University of Thessaloniki, Greece

^137.^ Bristol Medical School, University of Bristol, Bristol, UK

^138.^ Krembil Brain Institute, University Health Network, Toronto, Ontario, Canada

^139.^ Tanz Centre for Research in Neurodegenerative Diseases, Departments of Medicine and Laboratory Medicine & Pathobiology, University of Toronto, Toronto, Ontario, Canada

^140.^ Division of Mental Health and Addiction, Oslo University Hospital, Oslo, Norway

^141.^ KG Jebsen Centre for Neurodevelopmental disorders, University of Oslo, Oslo, Norway

^142.^ Alzheimer’s Centre Reina Sofia-CIEN Foundation, Madrid, Spain.

^143.^ Department of Psychiatry & Glenn Biggs Institute for Alzheimer’s and Neurodegenerative Diseases, San Antonio, TX, USA

^144.^ Biggs Institute for Alzheimer’s and Neurodegenerative Diseases, University of Texas Health Science Center, San Antonio, Texas, USA.

^145.^ Department of Epidemiology, ErasmusMC, Rotterdam, The Netherlands

^146.^ Department of Clinical Medicine, Faculty of Health and Medical Sciences, University of Copenhagen, Blegdamsvej 3B, 2100 Copenhagen, Denmark

## TABLE AND FIGURE REFERENCES

1. Bellenguez, C. *et al.* New insights into the genetic etiology of Alzheimer’s disease and related dementias. *Nat Genet* **54**, 412-436 (2022).

2. Pulit, S.L. *et al.* Meta-analysis of genome-wide association studies for body fat distribution in 694 649 individuals of European ancestry. *Hum Mol Genet* **28**, 166-174 (2019).

3. Winkler, T.W. *et al.* The Influence of Age and Sex on Genetic Associations with Adult Body Size and Shape: A Large-Scale Genome-Wide Interaction Study. *PLoS Genet* **11**, e1005378 (2015).

4. Mahajan, A. *et al.* Multi-ancestry genetic study of type 2 diabetes highlights the power of diverse populations for discovery and translation. *Nat Genet* **54**, 560-572 (2022).

5. Mahajan, A. *et al.* Fine-mapping type 2 diabetes loci to single-variant resolution using high-density imputation and islet-specific epigenome maps. *Nat Genet* **50**, 1505-1513 (2018).

6. Broce, I.J. *et al.* Dissecting the genetic relationship between cardiovascular risk factors and Alzheimer’s disease. *Acta Neuropathol* **137**, 209-226 (2019).

7. Wang, X.F. *et al.* Linking Alzheimer’s disease and type 2 diabetes: Novel shared susceptibility genes detected by cFDR approach. *Journal of the Neurological Sciences* **380**, 262-272 (2017).

8. Hardy, J., de Strooper, B. & Escott-Price, V. Diabetes and Alzheimer’s disease: shared genetic susceptibility? *Lancet Neurol* **21**, 962-964 (2022).

9. Yuan, X. *et al.* The common genes involved in the pathogenesis of Alzheimer’s disease and type 2 diabetes and their implication for drug repositioning. *Neuropharmacology* **223**, 109327 (2023).

10. Chung, Y., Lee, H. & the Alzheimer’s Disease Neuroimaging, I. Correlation between Alzheimer’s disease and type 2 diabetes using non-negative matrix factorization. *Sci Rep* **11**, 15265 (2021).

11. McKenzie, A.T. *et al.* Brain Cell Type Specific Gene Expression and Co-expression Network Architectures. *Sci Rep* **8**, 8868 (2018).

12. Zheng, W. *et al.* Small molecule angiotensin converting enzyme inhibitors: A medicinal chemistry perspective. *Front Pharmacol* **13**, 968104 (2022).

13. Heerspink, H.J.L. *et al.* Sparsentan in patients with IgA nephropathy: a prespecified interim analysis from a randomised, double-blind, active-controlled clinical trial. *Lancet* **401**, 1584-1594 (2023).

14. Xiao, H. *et al.* A first-in-class selective inhibitor of ERK1/2 and ERK5 overcomes drug resistance with a single-molecule strategy. *Signal Transduct Target Ther* **10**, 70 (2025).

15. Sim, Y.H., Um, Y.J., Park, J.Y., Seo, M.D. & Park, S.G. A Novel Antibody-Drug Conjugate Targeting Nectin-2 Suppresses Ovarian Cancer Progression in Mouse Xenograft Models. *Int J Mol Sci* **23**(2022).

16. Piala, A.T. *et al.* Discovery of novel TAOK2 inhibitor scaffolds from high-throughput screening. *Bioorg Med Chem Lett* **26**, 3923-7 (2016).

17. Koo, C.Y. *et al.* Targeting TAO Kinases Using a New Inhibitor Compound Delays Mitosis and Induces Mitotic Cell Death in Centrosome Amplified Breast Cancer Cells. *Mol Cancer Ther* **16**, 2410-2421 (2017).

18. Amabile, G. *et al.* Targeting of the IGFBP3/TMEM219 pathway restores intestinal stem cells capability of healing mucosa in gastrointestinal autoimmune disorders. *Pharmacol Res* **219**, 107901 (2025).

19. Mokhtariye, A., Varshosaz, J., Mohammadalipour, A., Hashemnia, M. & Mofid, M.R. Xanthosomes loaded with recombinant TMEM219 for targeting IGFBP-3: A novel approach for targeted drug delivery to ameliorate liver fibrosis. *Life Sci* **377**, 123748 (2025).

20. Zhang, W. *et al.* The aminophospholipid transporter, ATP8B3, as a potential biomarker and target for enhancing the therapeutic effect of PD-L1 blockade in colon adenocarcinoma. *Genomics* **116**, 110907 (2024).

21. Caldon, C.E. *et al.* Cyclin E2 overexpression is associated with endocrine resistance but not insensitivity to CDK2 inhibition in human breast cancer cells. *Mol Cancer Ther* **11**, 1488-99 (2012).

22. Abdel-Magid, A.F. Potential of Cyclin-Dependent Kinase Inhibitors as Cancer Therapy. *ACS Med Chem Lett* **12**, 182-184 (2021).

23. Decker, M. *et al.* In vivo and In vitro Chemical Synthetic Lethality of ABHD17 and MEK Inhibition in NRAS-Mutant Acute Myeloid Leukemia. *Blood* **144**, 5808-5809 (2024).

24. Key, C.C. *et al.* Human GDPD3 overexpression promotes liver steatosis by increasing lysophosphatidic acid production and fatty acid uptake. *J Lipid Res* **61**, 1075-1086 (2020).

25. Giusiano, S. *et al.* TP53INP1 as new therapeutic target in castration-resistant prostate cancer. *Prostate* **72**, 1286-94 (2012).

26. Bialkowska, K. *et al.* Role of Kindlin 2 in prostate cancer. *Sci Rep* **14**, 19809 (2024).

27. Jia, J. *et al.* Effect of Dl-3-n-butylphthalide on mitochondrial Cox7c in models of cerebral ischemia/reperfusion injury. *Front Pharmacol* **14**, 1084564 (2023).

28. Wu, X. *et al.* Unraveling the role of Major Vault Protein as a novel immune-related biomarker that promotes the proliferation and migration in pancreatic adenocarcinoma. *Front Immunol* **15**, 1399222 (2024).

29. Raja, R. *et al.* PP4 inhibition sensitizes ovarian cancer to NK cell-mediated cytotoxicity via STAT1 activation and inflammatory signaling. *J Immunother Cancer* **10**(2022).

