## Supplementary Figures for "Genomics link obesity and type 2 diabetes to Alzheimer’s disease to unveil novel biological insights"

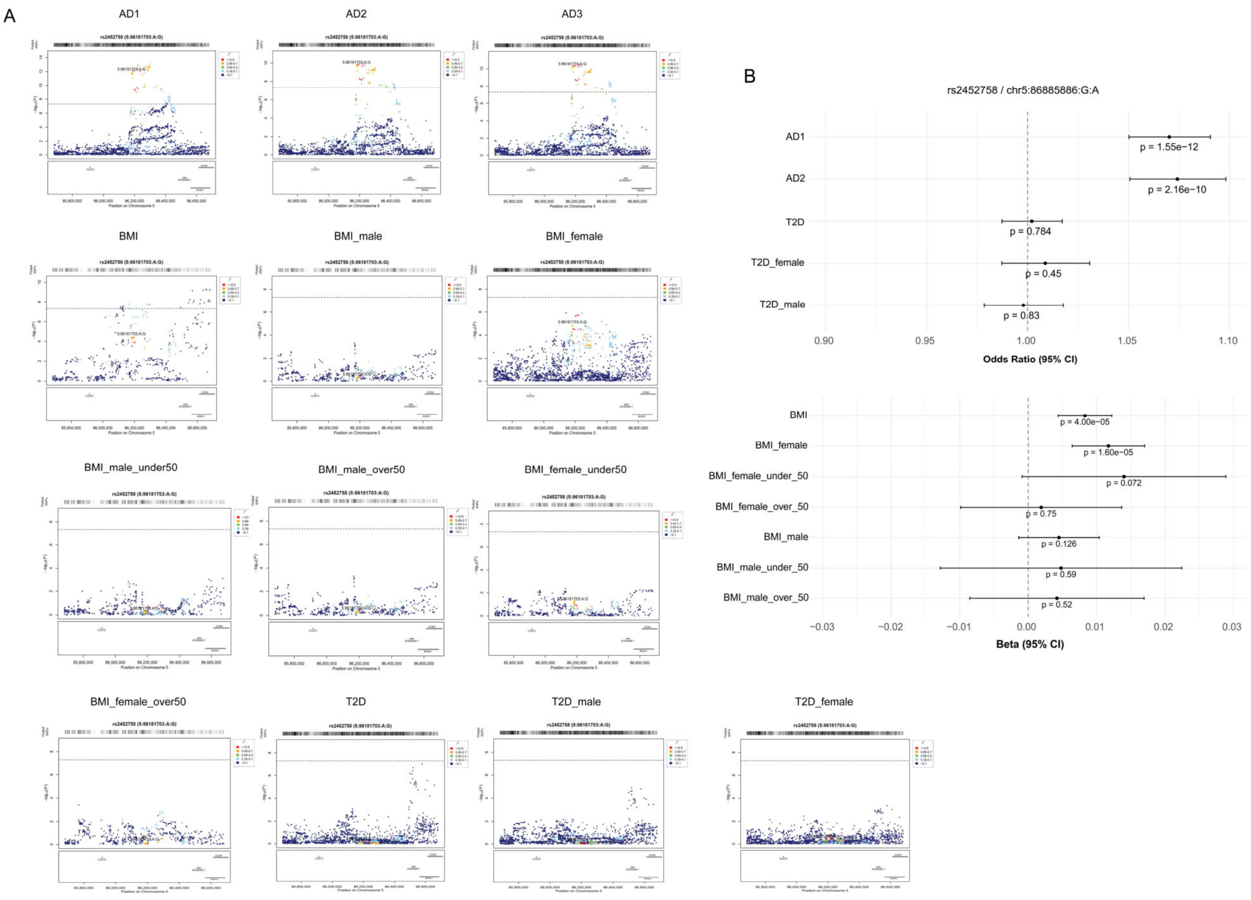


**Fig. S1**A | **Genetic colocalization analysis**. (**A**) Locuszoom plots for the BMI-AD rs2452758 locus (GRCh37). The overlapped variant is circled in green (**Table S3**) while the colocalized variant is shown in yellow (rs2452758). Variants in linkage disequilibrium ($R^2$) are color-coded. (**B**) Forest plot for the BMI-AD rs2452758 locus showcasing odds ratio for dichotomous traits (AD and T2D) and effect size (beta) for the continuous trait (BMI). P-values are shown under each odds ratio/beta estimate. Odds ratio and p-values for BMI-AD, T2D-AD and BMI-T2D-AD risk variants across AD datasets (AD1, AD2 and AD3).

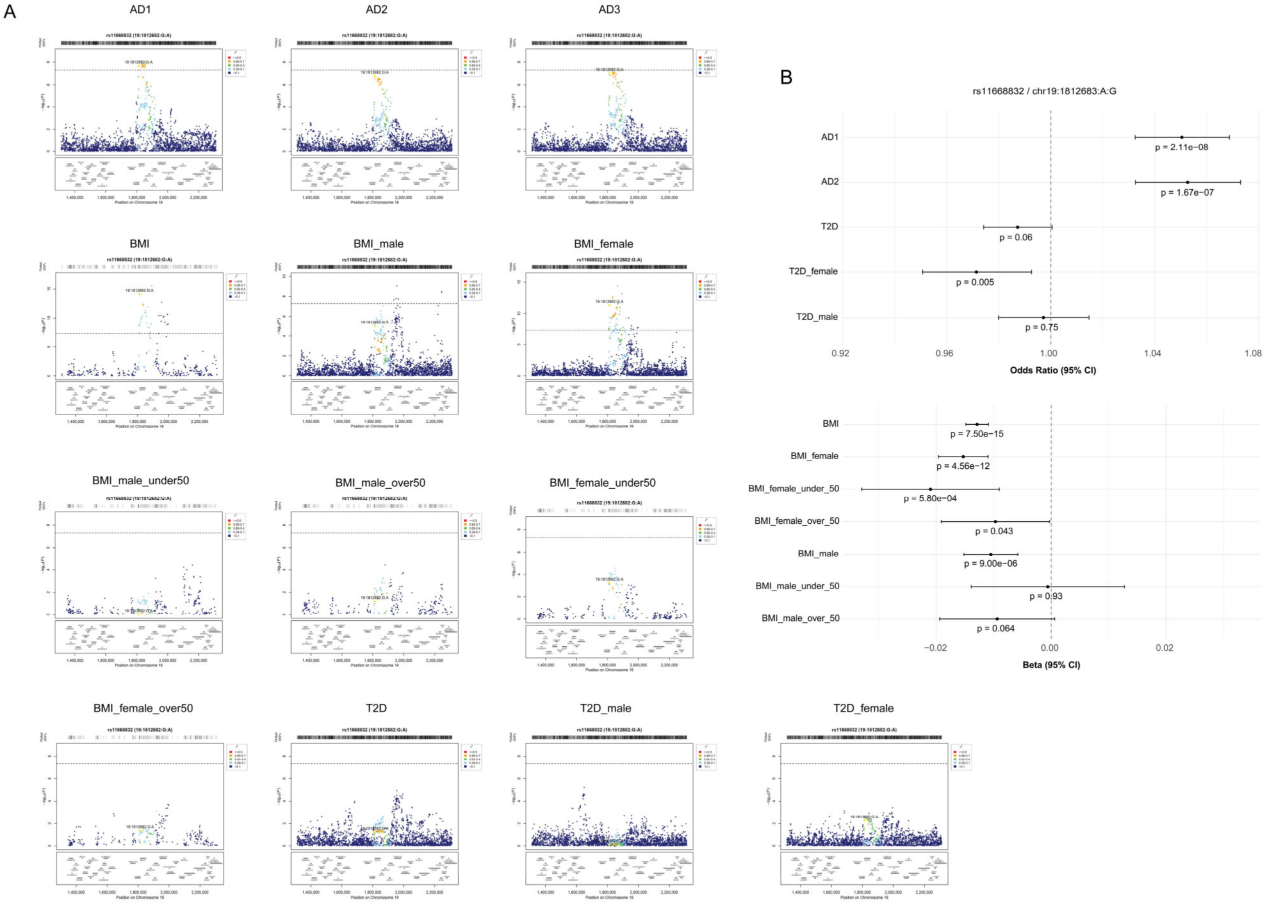


**Fig. S1**B | **Genetic colocalization analysis**. (**A**) Locuszoom plots for the BMI-AD rs11668832 locus (GRCh37). The overlapped variant is circled in green (**Table S3**) while the colocalized variant is shown in yellow (rs11668832). Variants in linkage disequilibrium ($R^2$) are color-coded. (**B**) Forest plot for the BMI-AD rs11668832 locus showcasing odds ratio for dichotomous traits (AD and T2D) and effect size (beta) for the continuous trait (BMI). P-values are shown under each odds ratio/beta estimate. Odds ratio and p-values for BMI-AD, T2D-AD and BMI-T2D-AD risk variants across AD datasets (AD1, AD2 and AD3).

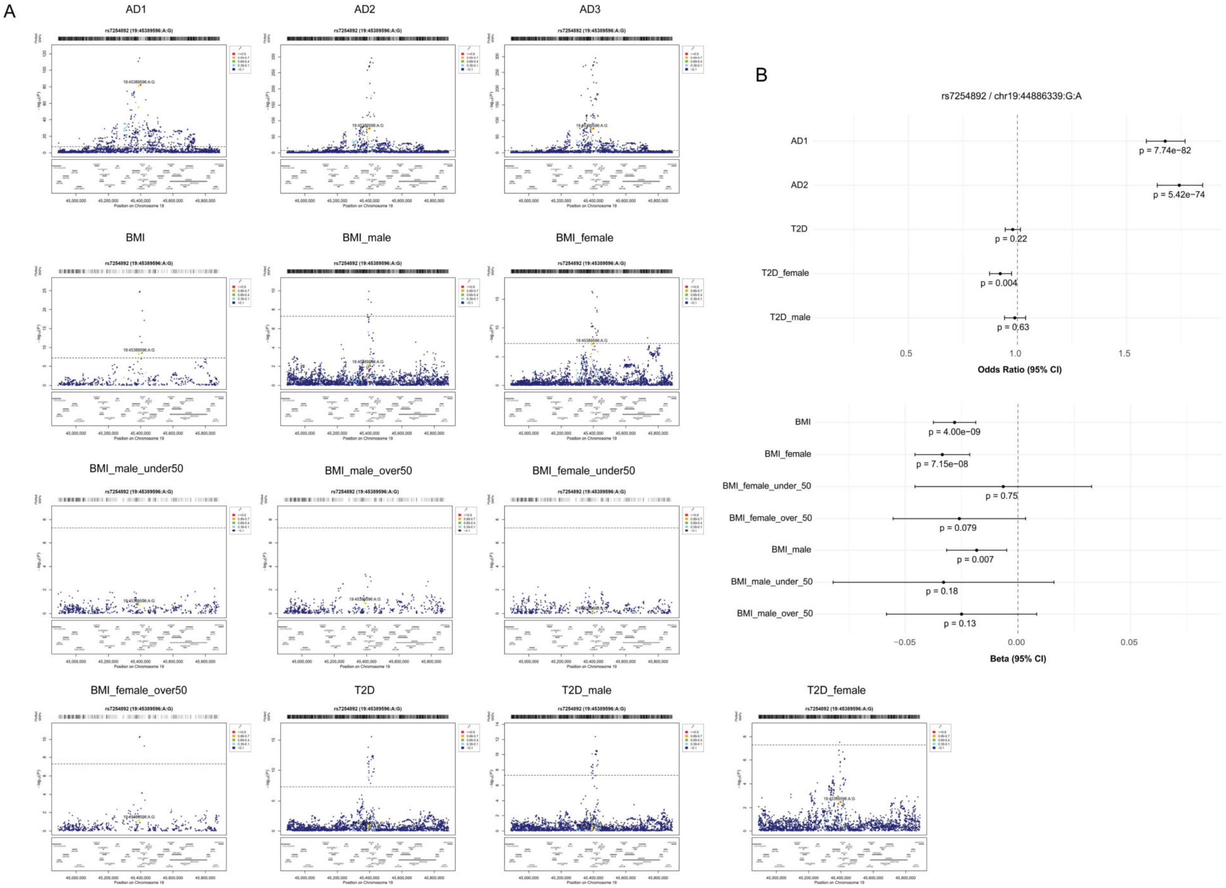


**Fig. S1**C | **Genetic colocalization analysis**. (**A**) Locuszoom plots for the BMI-AD rs7254892 locus (GRCh37). The overlapped variant is circled in green (**Table S3**) while the colocalized variant is shown in yellow (rs7254892). Variants in linkage disequilibrium ($R^2$) are color-coded. (**B**) Forest plot for the BMI-AD rs7254892 locus showcasing odds ratio for dichotomous traits (AD and T2D) and effect size (beta) for the continuous trait (BMI). P-values are shown under each odds ratio/beta estimate. Odds ratio and p-values for BMI-AD, T2D-AD and BMI-T2D-AD risk variants across AD datasets (AD1, AD2 and AD3).

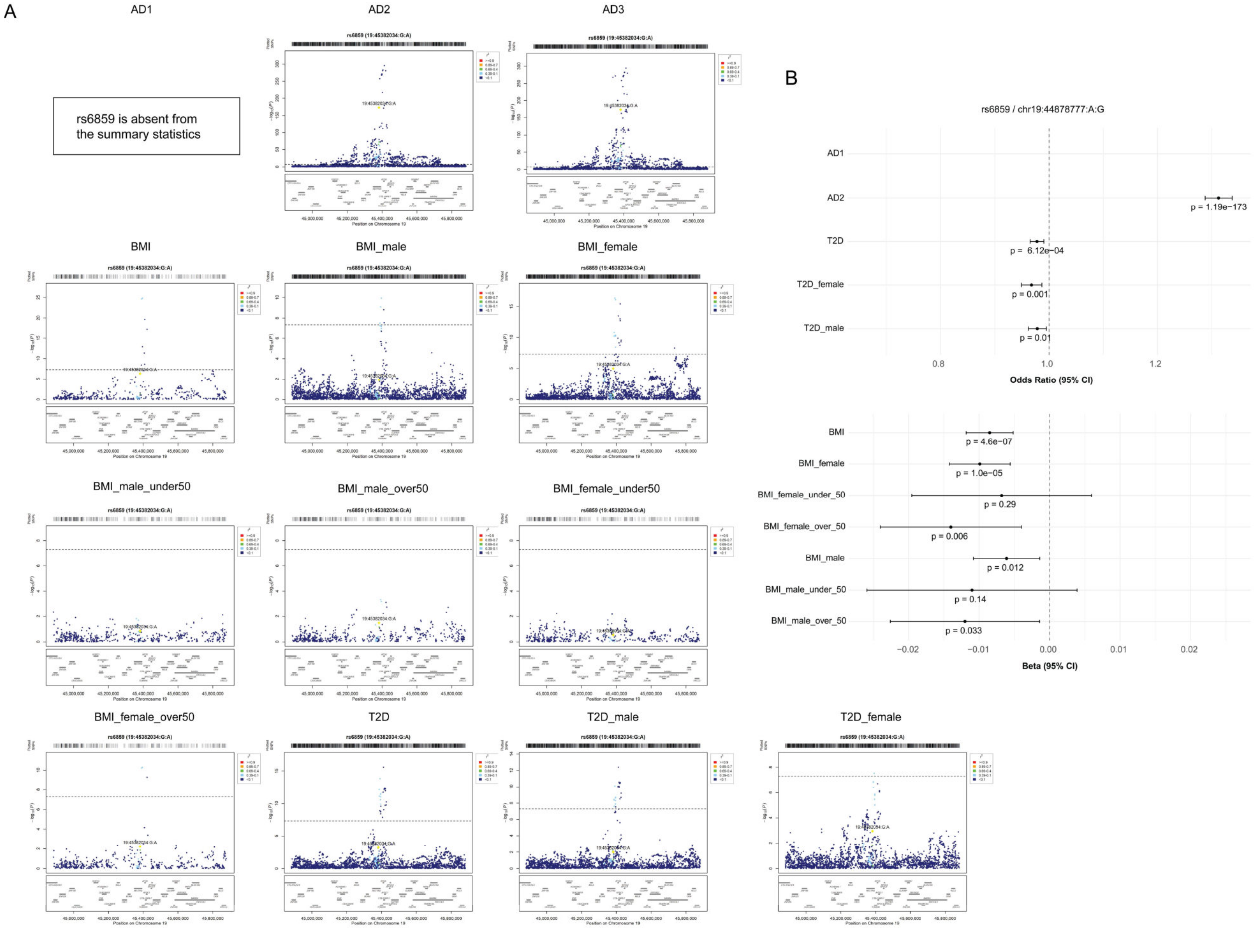


**Fig. S1**D | **Genetic colocalization analysis**. (**A**) Locuszoom plots for the BMI-AD rs6859 locus (GRCh37). The overlapped variant is circled in green (**Table S3**) while the colocalized variant is shown in yellow (rs6859). Variants in linkage disequilibrium ($R^2$) are color-coded. (**B**) Forest plot for the BMI-AD rs6859 locus showcasing odds ratio for dichotomous traits (AD and T2D) and effect size (beta) for the continuous trait (BMI). P-values are shown under each odds ratio/beta estimate. Odds ratio and p-values for BMI-AD, T2D-AD and BMI-T2D-AD risk variants across AD datasets (AD1, AD2 and AD3).

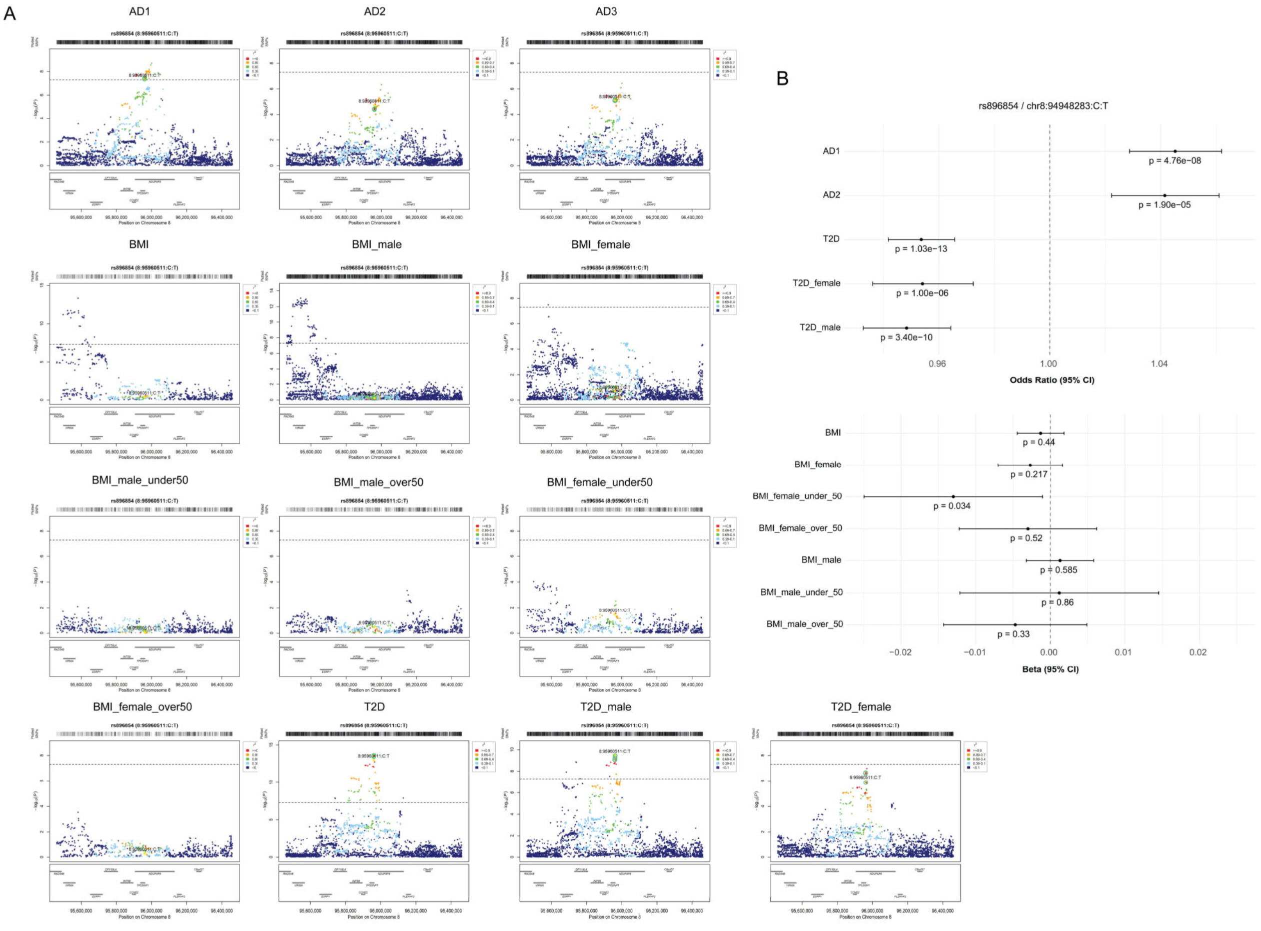


**Fig. S1**E | **Genetic colocalization analysis**. (**A**) Locuszoom plots for the T2D-AD rs896854 locus (GRCh37). The overlapped variant is circled in green (**Table S3**) while the colocalized variant is shown in yellow (rs896854). Variants in linkage disequilibrium ($R^2$) are color-coded. (**B**) Forest plot for the T2D-AD rs896854 locus showcasing odds ratio for dichotomous traits (AD and T2D) and effect size (beta) for the continuous trait (BMI). P-values are shown under each odds ratio/beta estimate. Odds ratio and p-values for BMI-AD, T2D-AD and BMI-T2D-AD risk variants across AD datasets (AD1, AD2 and AD3).

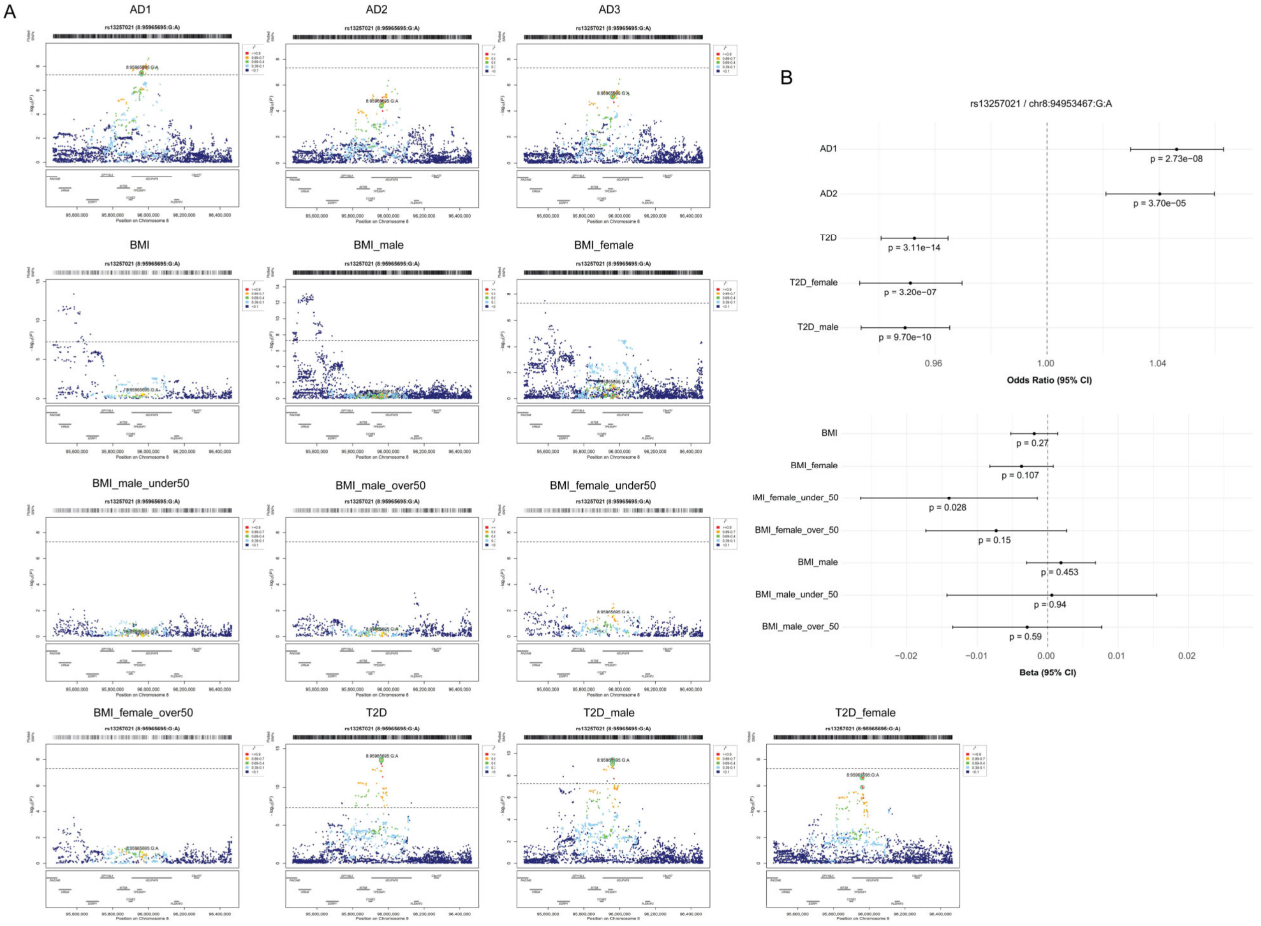


**Fig. S1**F | **Genetic colocalization analysis**. (**A**) Locuszoom plots for the T2D-AD rs13257021 locus (GRCh37). The overlapped variant is circled in green (**Table S3**) while the colocalized variant is shown in yellow (rs13257021). Variants in linkage disequilibrium ($R^2$) are color-coded. (**B**) Forest plot for the T2D-AD rs13257021 locus showcasing odds ratio for dichotomous traits (AD and T2D) and effect size (beta) for the continuous trait (BMI). P-values are shown under each odds ratio/beta estimate. Odds ratio and p-values for BMI-AD, T2D-AD and BMI-T2D-AD risk variants across AD datasets (AD1, AD2 and AD3).

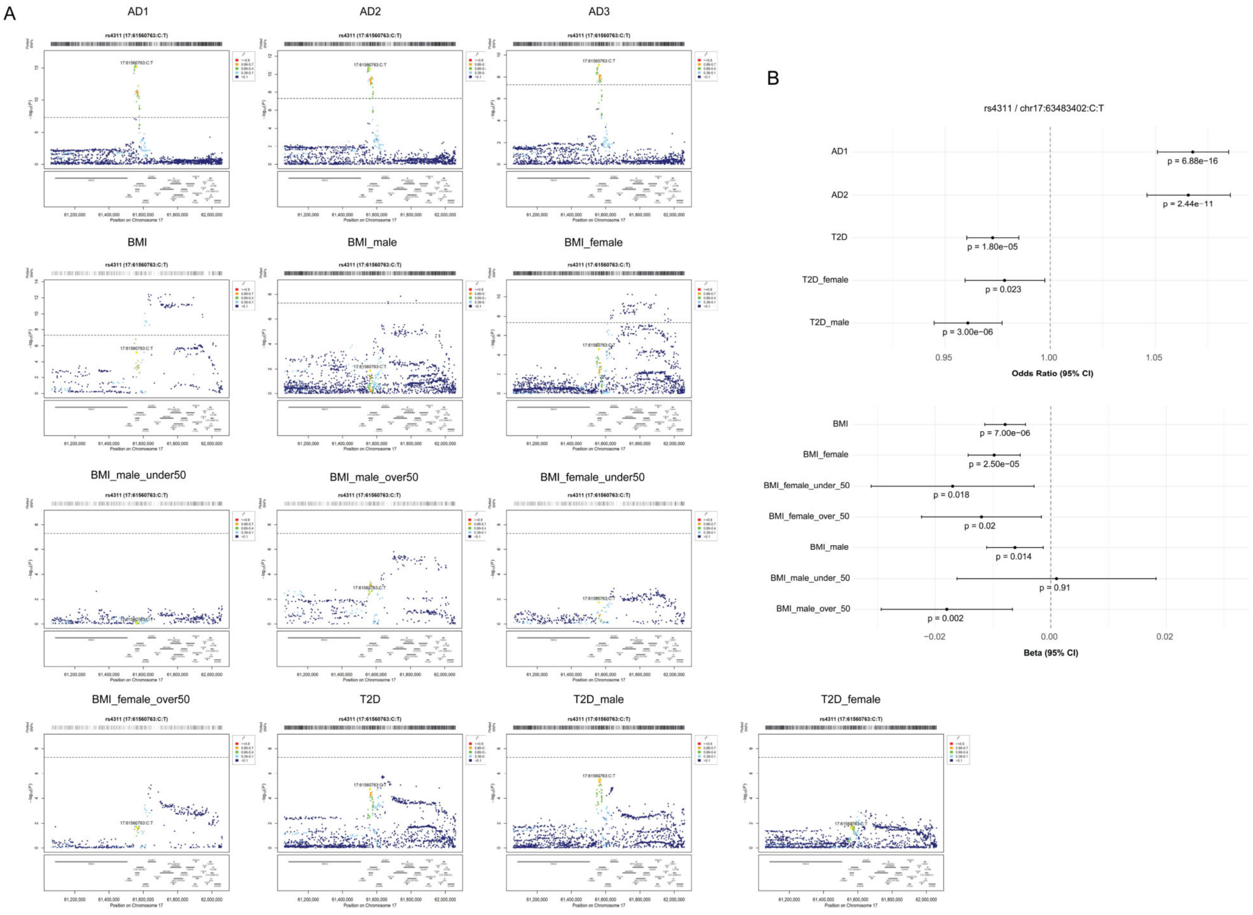


**Fig. S1**G | **Genetic colocalization analysis**. (**A**) Locuszoom plots for the T2D-AD rs4311 locus (GRCh37). The overlapped variant is circled in green (**Table S3**) while the colocalized variant is shown in yellow (rs4311). Variants in linkage disequilibrium ($R^2$) are color-coded. (**B**) Forest plot for the T2D - AD rs4311 locus showcasing odds ratio for dichotomous traits (AD and T2D) and effect size (beta) for the continuous trait (BMI). P-values are shown under each odds ratio/beta estimate. Odds ratio and p-values for BMI-AD, T2D-AD and BMI-T2D-AD risk variants across AD datasets (AD1, AD2 and AD3).

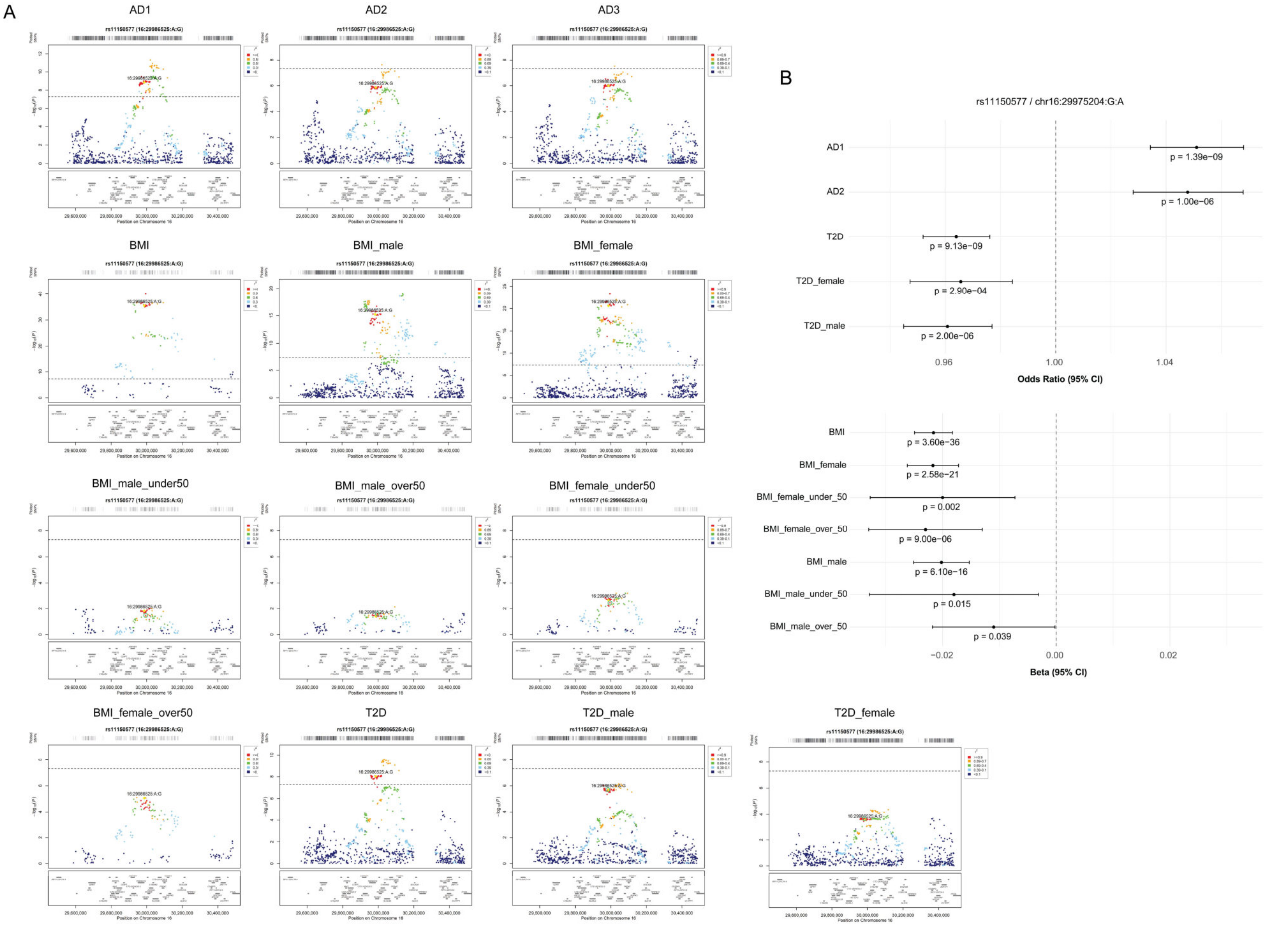


**Fig. S1**H | **Genetic colocalization analysis**. (**A**) Locuszoom plots for the BMI-T2D-AD rs11150577 locus (GRCh37). The overlapped variant is circled in green (**Table S3**) while the colocalized variant is shown in yellow (rs11150577). Variants in linkage disequilibrium ($R^2$) are color-coded. (**B**) Forest plot for the BMI-T2D-AD rs11150577 locus showcasing odds ratio for dichotomous traits (AD and T2D) and effect size (beta) for the continuous trait (BMI). P-values are shown under each odds ratio/beta estimate. Odds ratio and p-values for BMI-AD, T2D-AD and BMI-T2D-AD risk variants across AD datasets (AD1, AD2 and AD3).

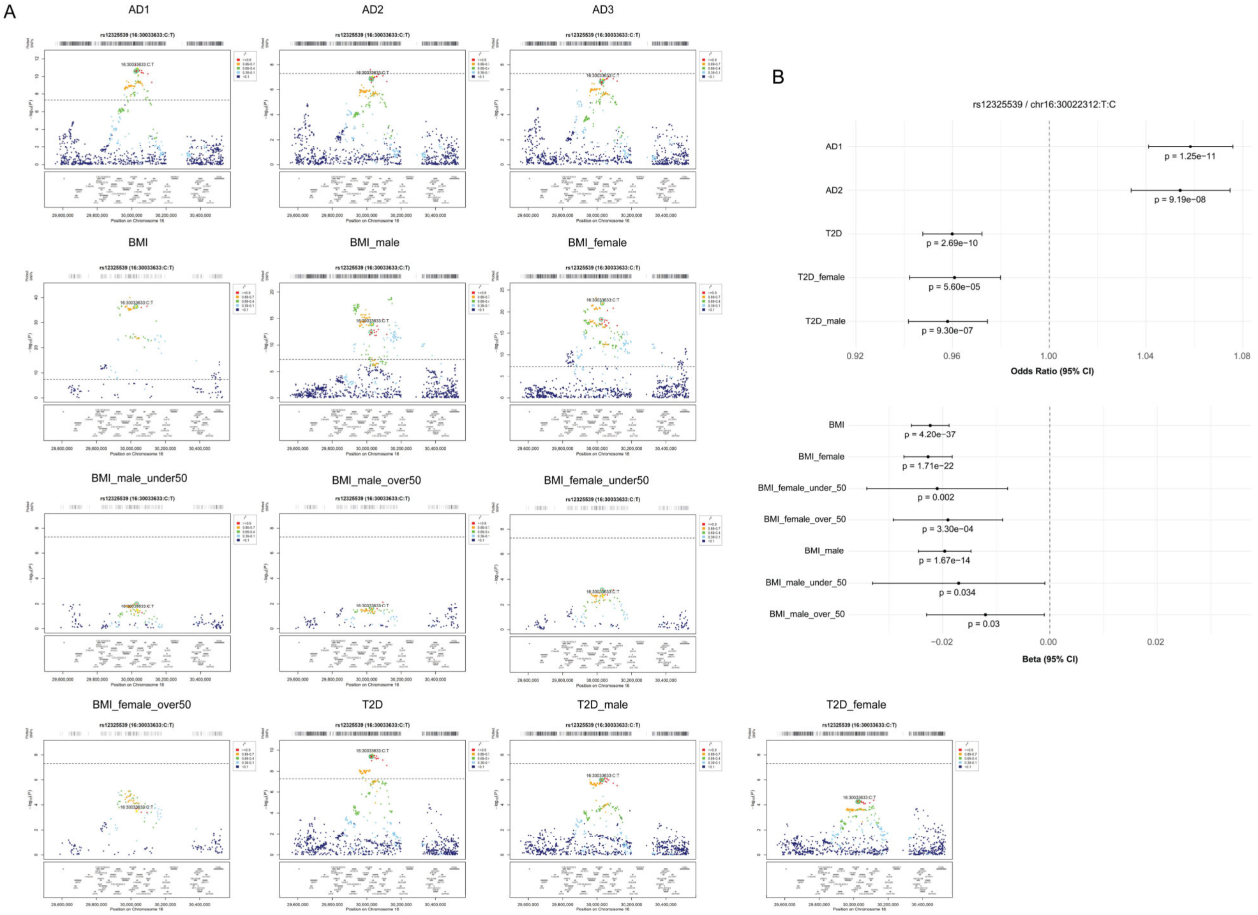


**Fig. S1l | Genetic colocalization analysis**. (**A**) Locuszoom plots for the BMI-T2D-AD rs12325539 locus (GRCh37). The overlapped variant is circled in green (**Table S3**) while the colocalized variant is shown in yellow (rs12325539). Variants in linkage disequilibrium ($R^2$) are color-coded. (**B**) Forest plot for the BMI-T2D-AD rs12325539 locus showcasing odds ratio for dichotomous traits (AD and T2D) and effect size (beta) for the continuous trait (BMI). P-values are shown under each odds ratio/beta estimate. Odds ratio and p-values for BMI-AD, T2D-AD and BMI-T2D-AD risk variants across AD datasets (AD1, AD2 and AD3).

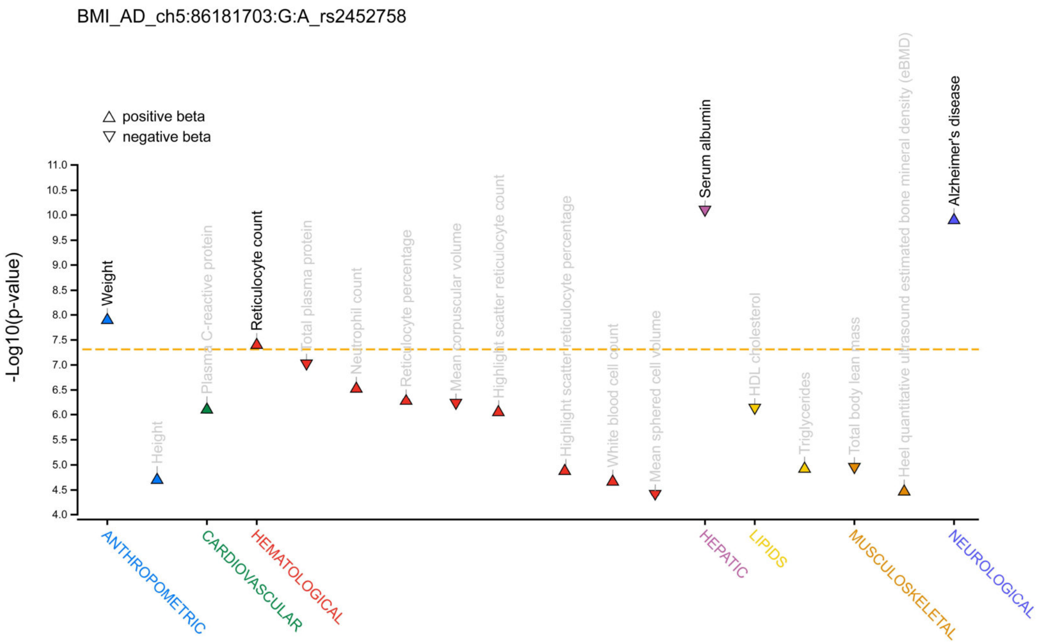


**Fig. S2**A | **Phenome wide association study (PheWAS)**. Results are aligned to the risk-increasing allele for AD. Position in hg19.

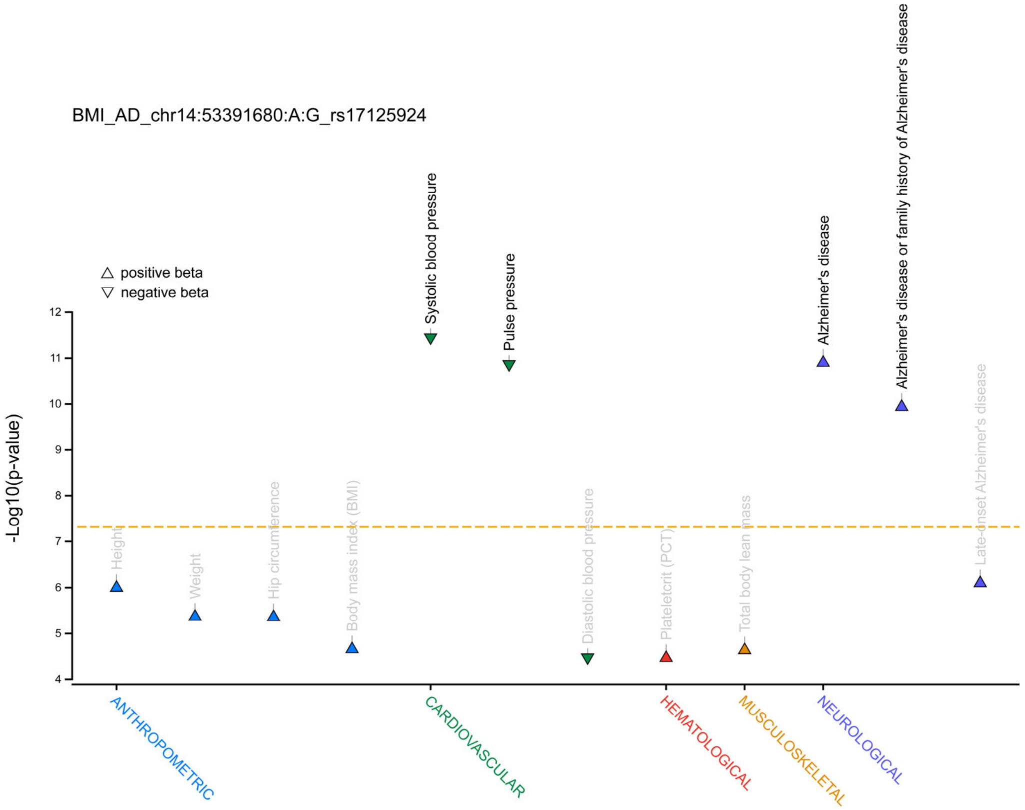


**Fig. S2**B | **Phenome wide association study (PheWAS)**. Results are aligned to the risk-increasing allele for AD. Position in hg19.

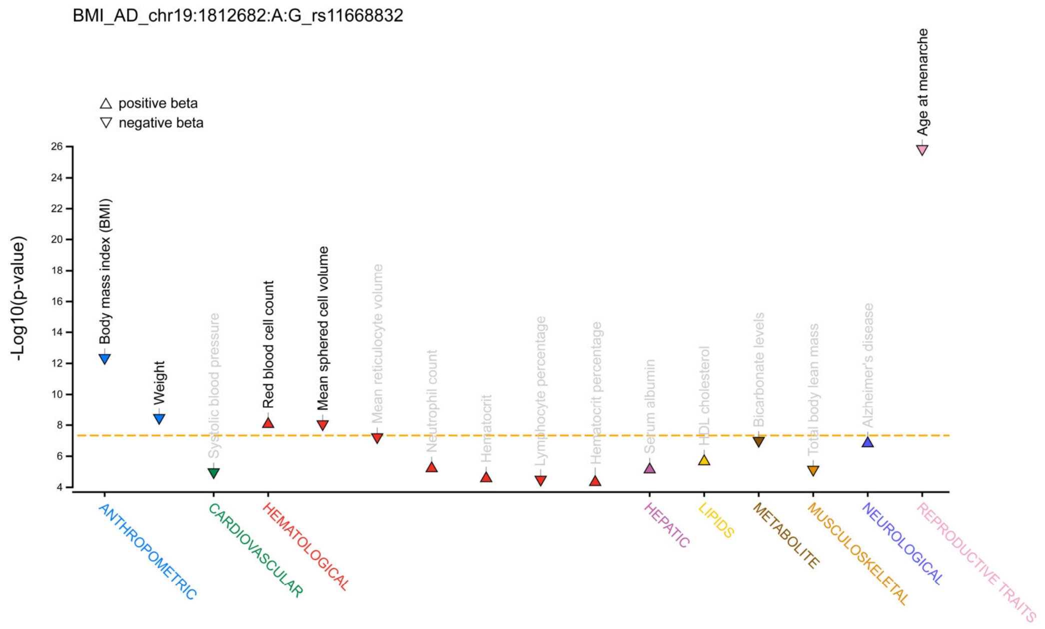


**Fig. S2**C | **Phenome wide association study (PheWAS)**. Results are aligned to the risk-increasing allele for AD. Position in hg19.

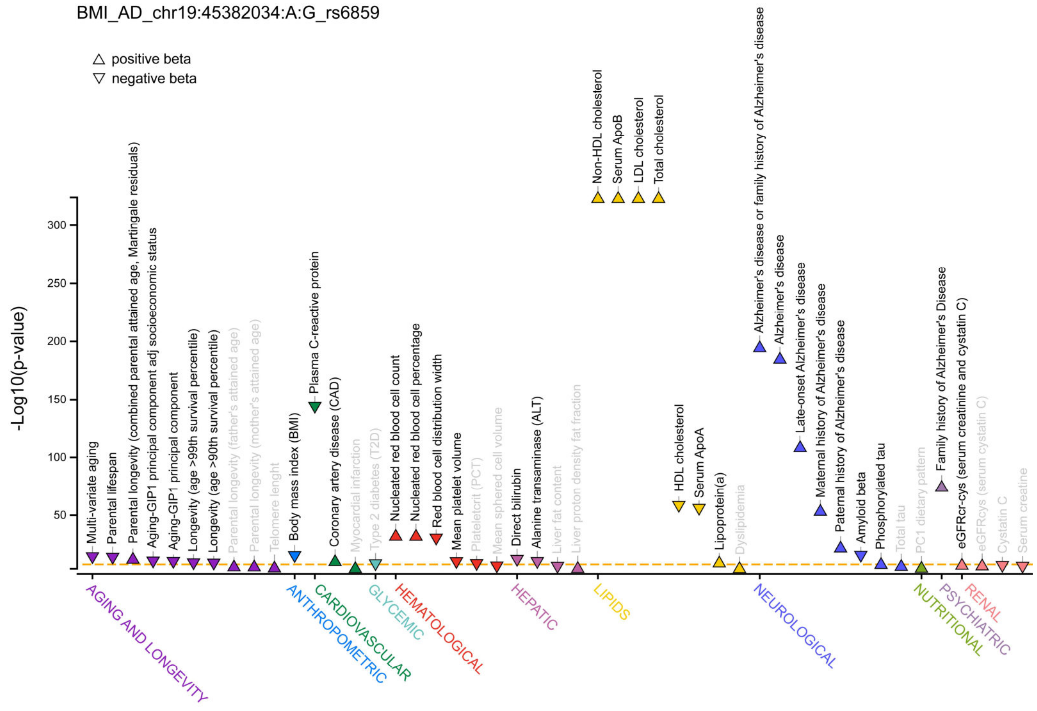


**Fig. S2**D | **Phenome wide association study (PheWAS)**. Results are aligned to the risk-increasing allele for AD. Position in hg19.

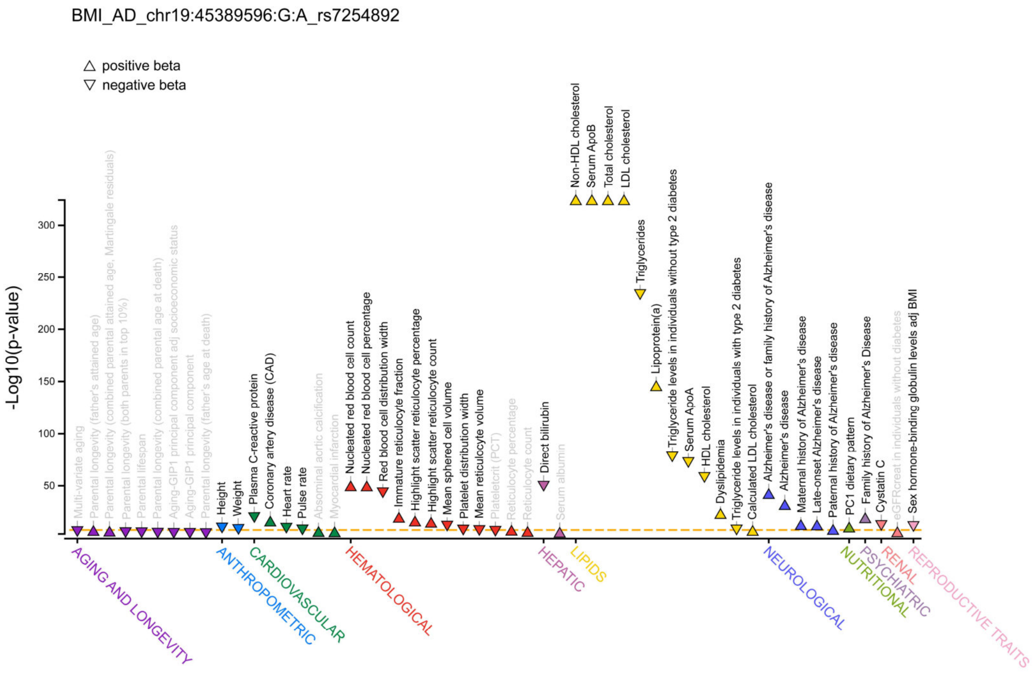


**Fig. S2**E | **Phenome wide association study (PheWAS)**. Results are aligned to the risk-increasing allele for AD. Position in hg19.

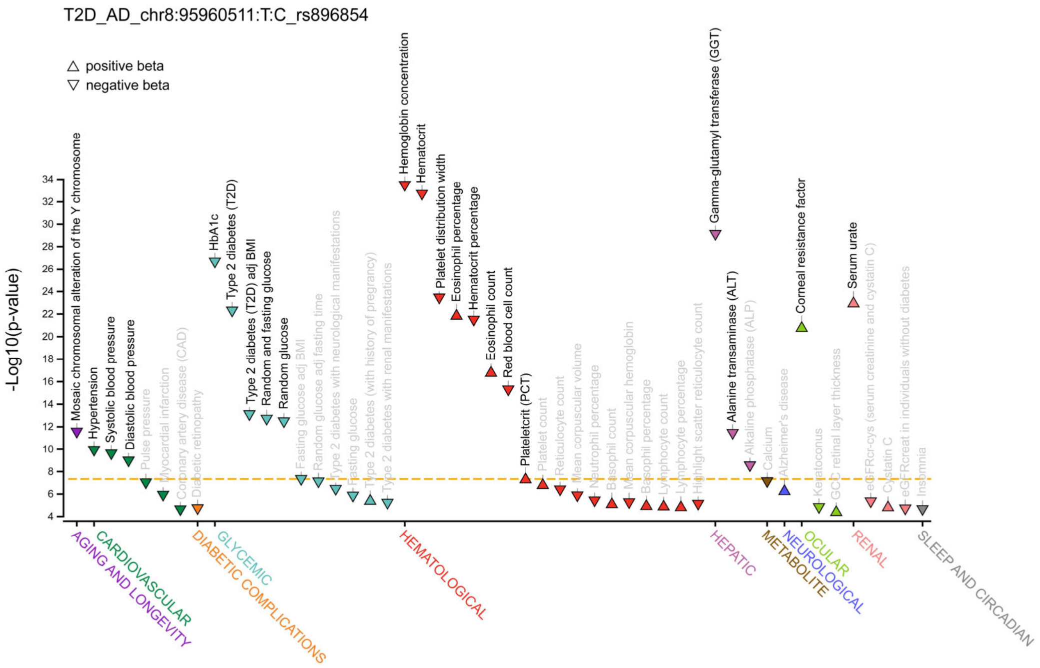


**Fig. S2**F | **Phenome wide association study (PheWAS)**. Results are aligned to the risk-increasing allele for AD. Position in hg19.

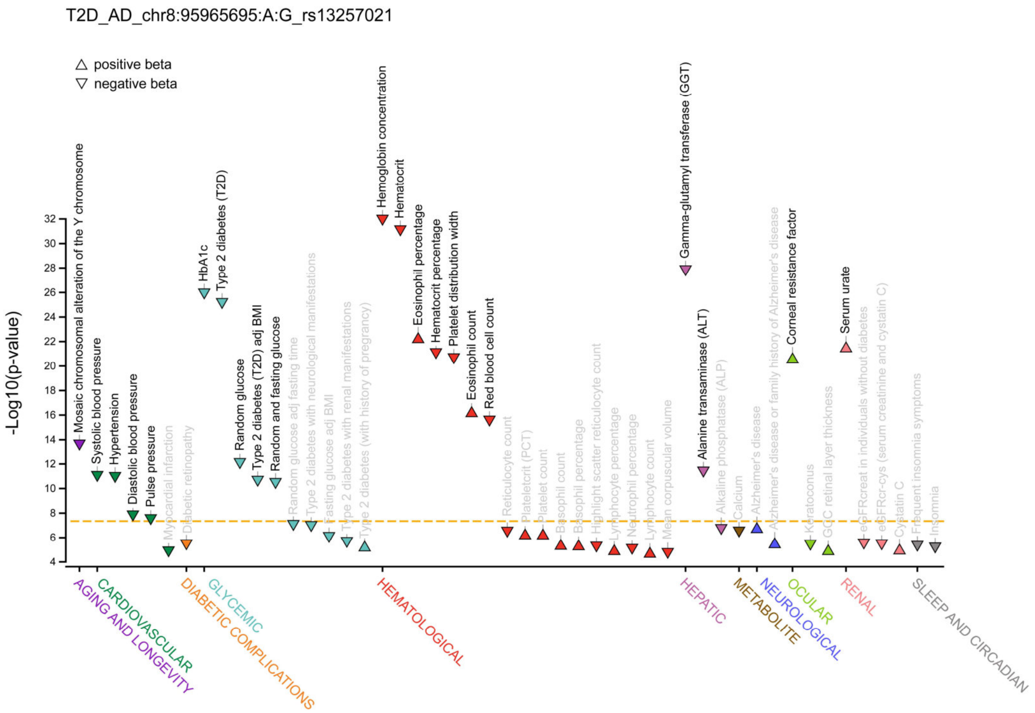


**Fig. S2**G | **Phenome wide association study (PheWAS)**. Results are aligned to the risk-increasing allele for AD. Position in hg19.

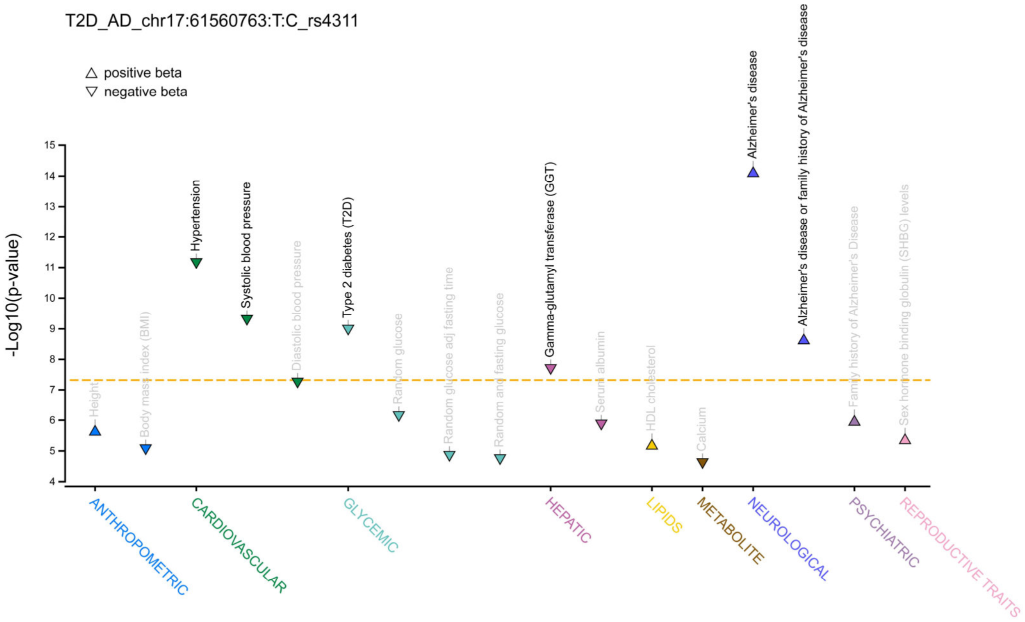


**Fig. S2**H | **Phenome wide association study (PheWAS)**. Results are aligned to the risk-increasing allele for AD. Position in hg19.

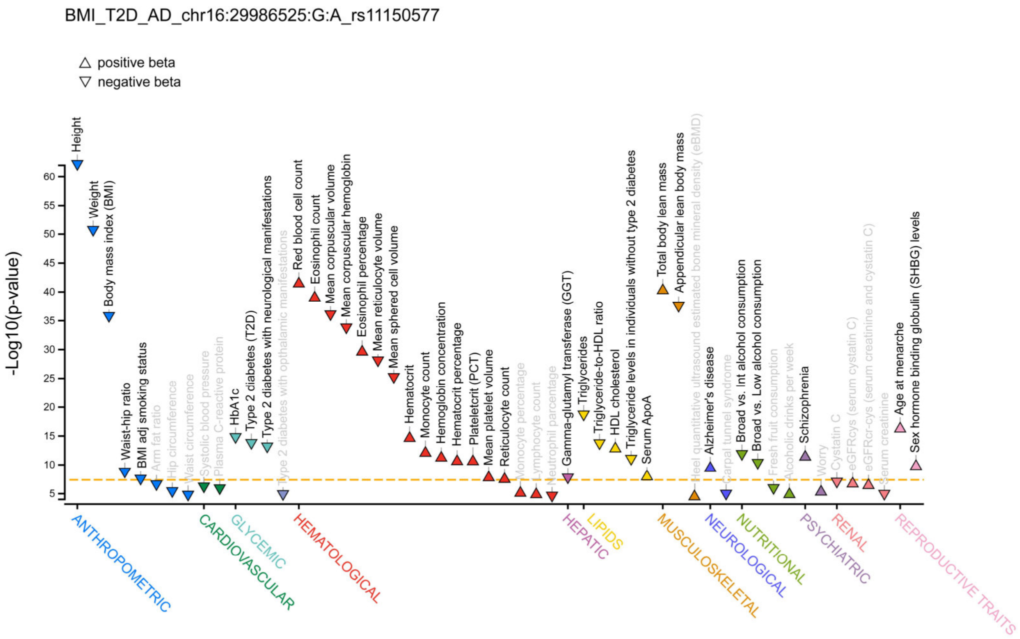


**Fig. S2**I | **Phenome wide association study (PheWAS)**. Results are aligned to the risk-increasing allele for AD. Position in hg19.

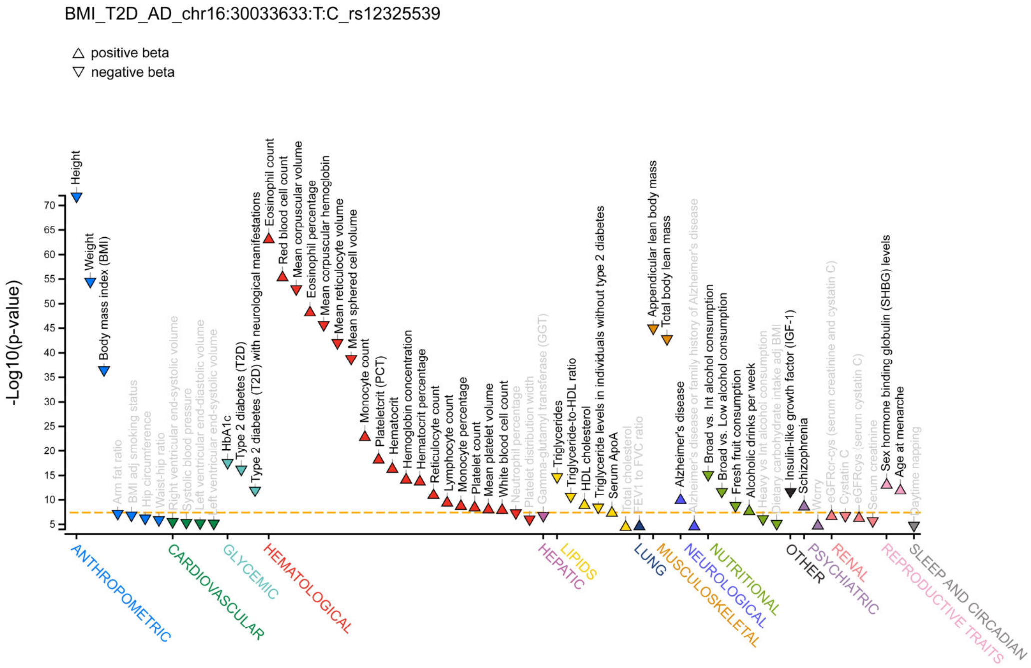


**Fig. S2**J | **Phenome wide association study (PheWAS)**. Results are aligned to the risk-increasing allele for AD. Position in hg19.

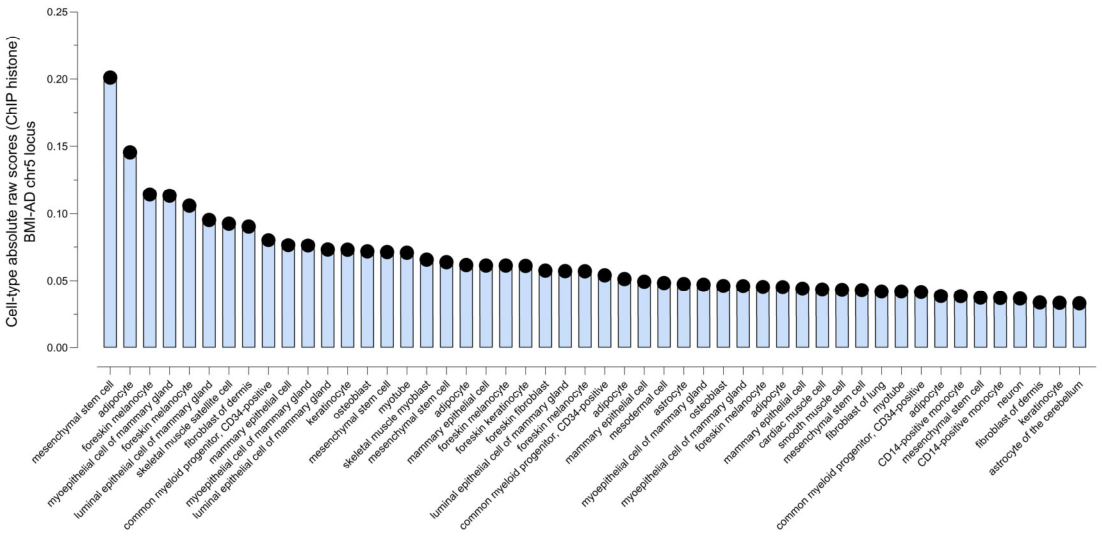


**Fig. S3**A | **AlphaGenome**. Top cell-type predictions for the effects of rs2452758 in histone modifications.

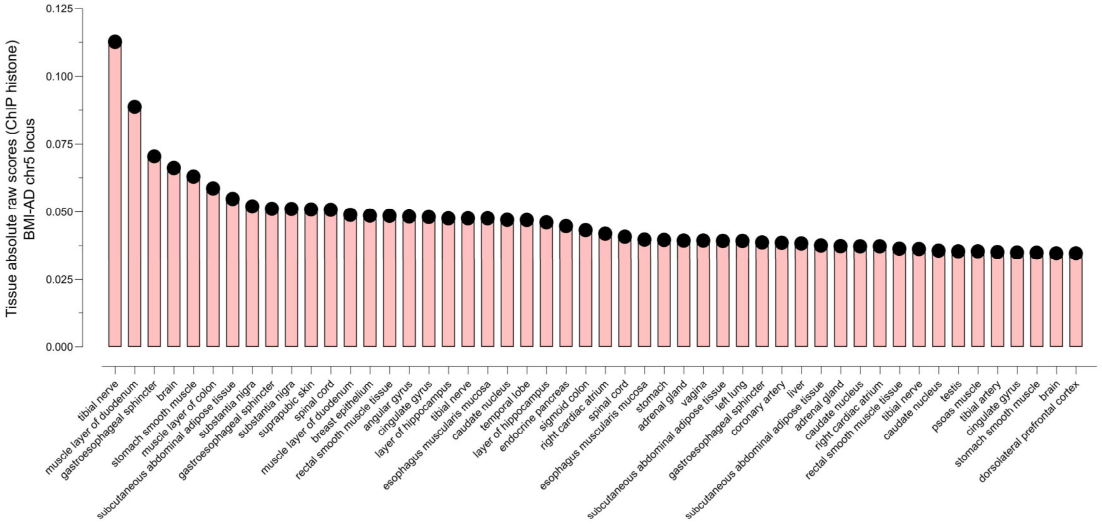


**Fig. S3**B | **AlphaGenome**. Top tissue predictions for the effects of rs2452758 in histone modifications.

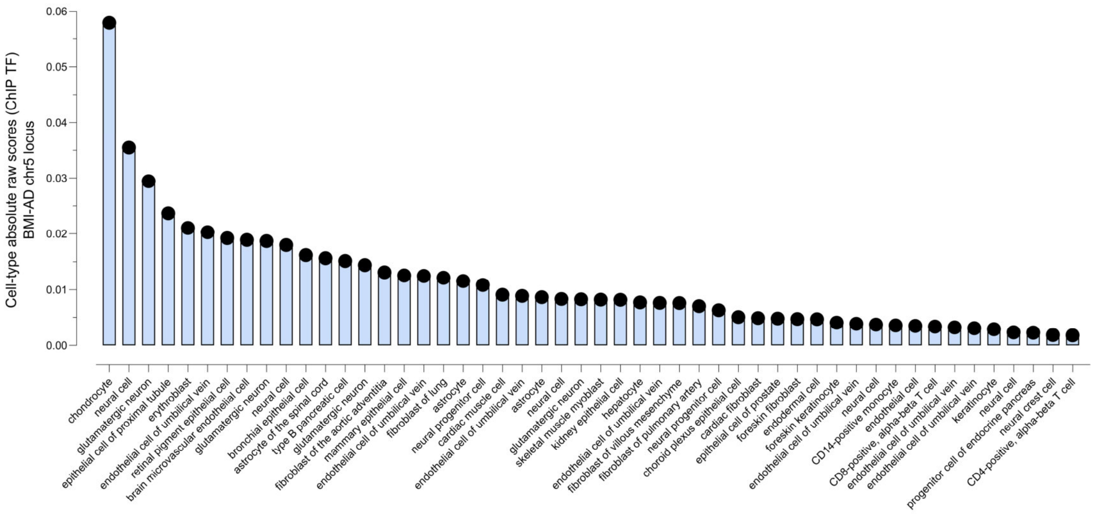


**Fig. S3**C | **AlphaGenome**. Top cell-type predictions for the effects of rs2452758 in transcription factor binding.

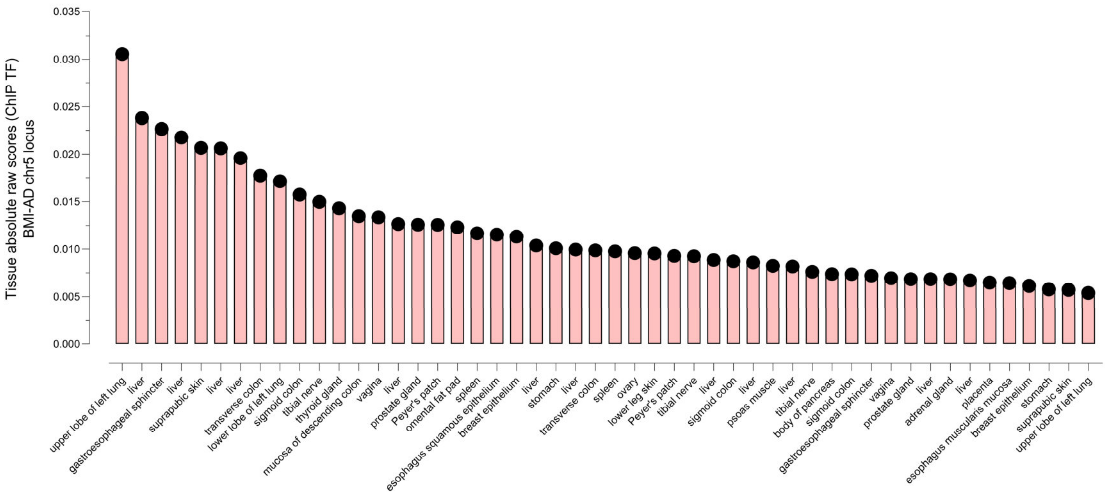


**Fig. S3**D | **AlphaGenome**. Top tissue predictions for the effects of rs2452758 in transcription factor binding.

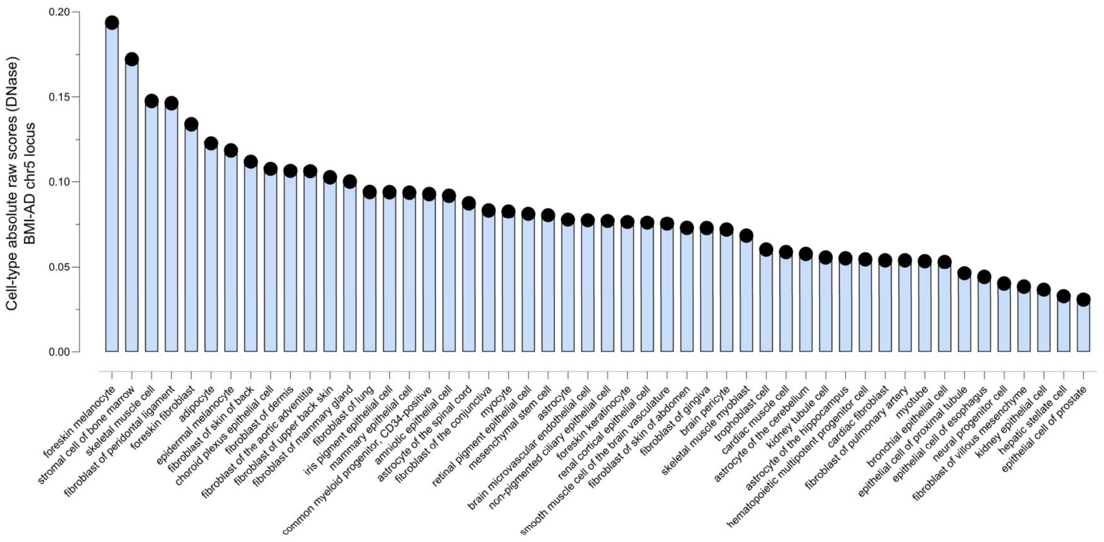


**Fig. S3**E | **AlphaGenome**. Top cell-type predictions for the effects of rs2452758 in chromatin accessibility assessed by DNase.

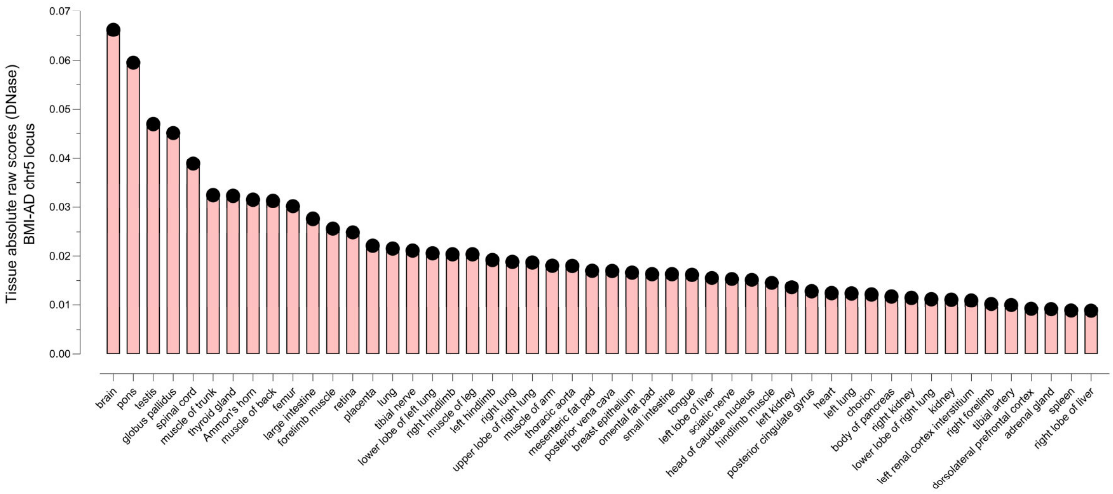


**Fig. S3**F | **AlphaGenome**. Top tissue predictions for the effects of rs2452758 in chromatin accessibility assessed by DNase.

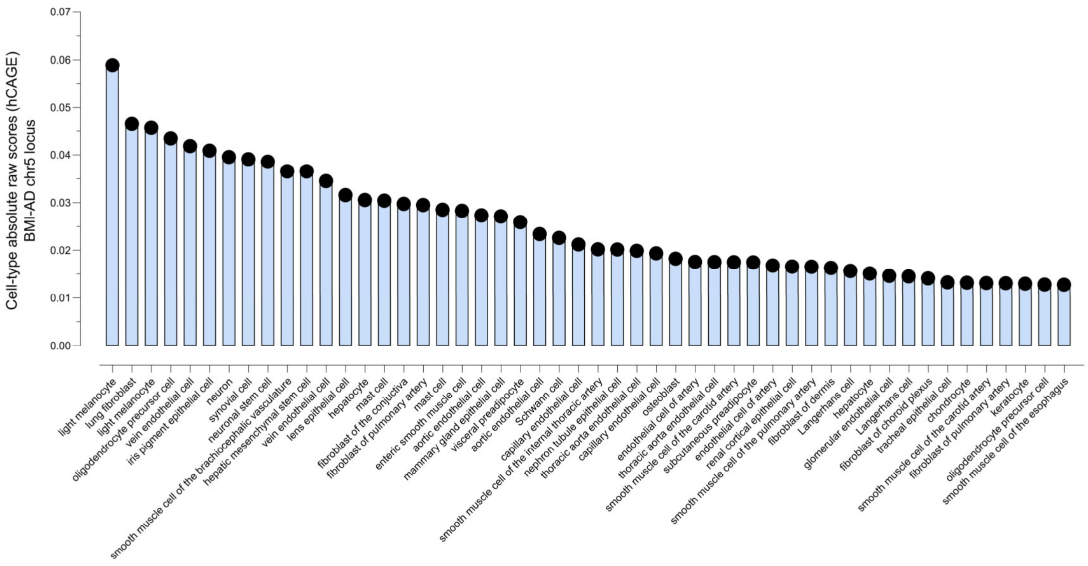


**Fig. S3**G | **AlphaGenome**. Top cell-type predictions for the effects of rs2452758 in gene expression assessed by hCAGE.

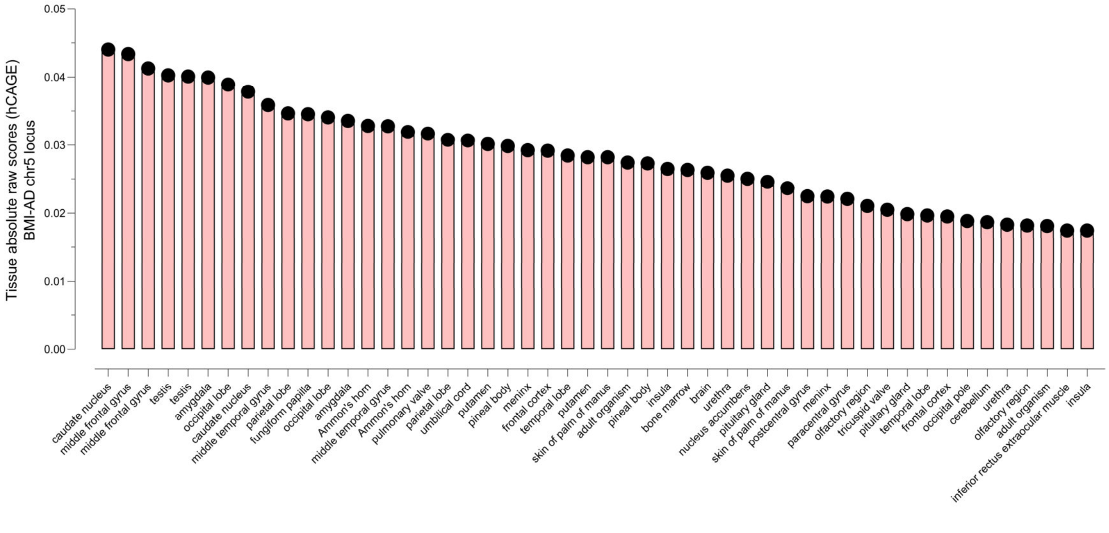


**Fig. S3**H | **AlphaGenome**. Top tissue predictions for the effects of rs2452758 in gene expression assessed by hCAGE.

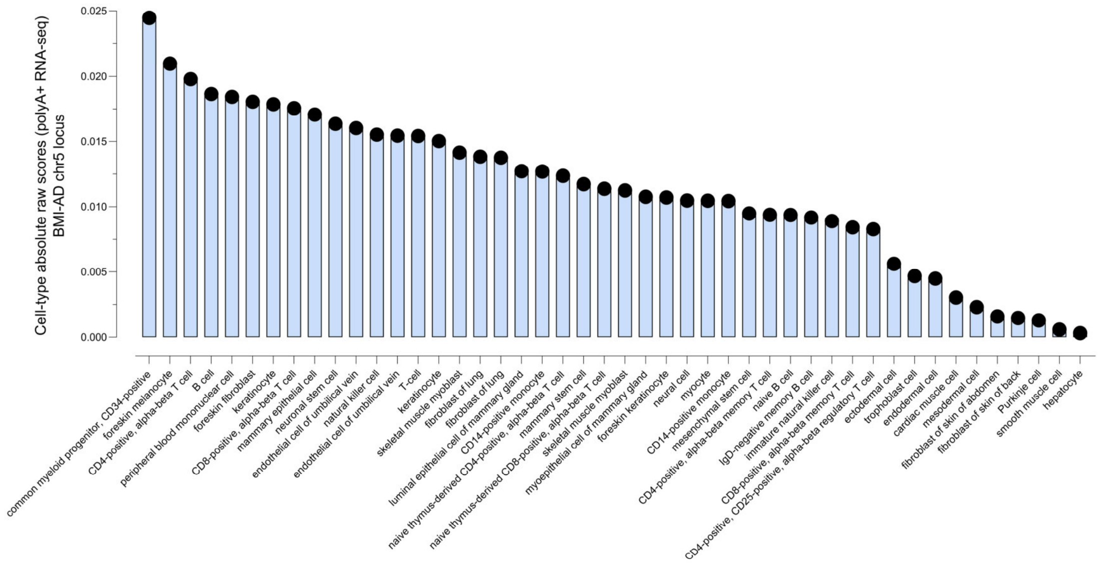


**Fig. S3**I | **AlphaGenome**. Top tissue predictions for the effects of rs2452758 in gene expression assessed by polyA+ RNA-seq.

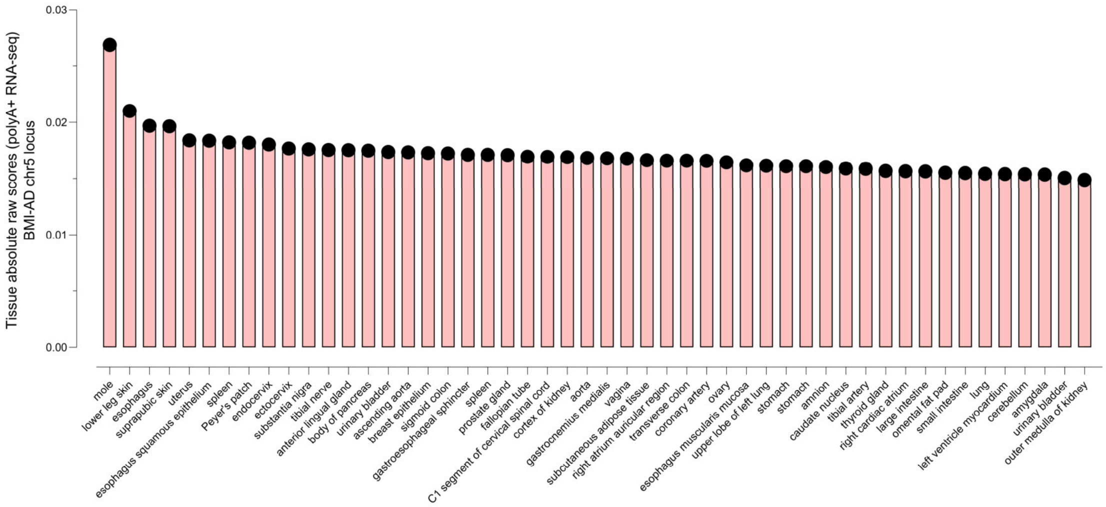


**Fig. S3**J | **AlphaGenome**. Top cell-type predictions for the effects of rs2452758 in gene expression assessed by polyA+ RNA-seq.

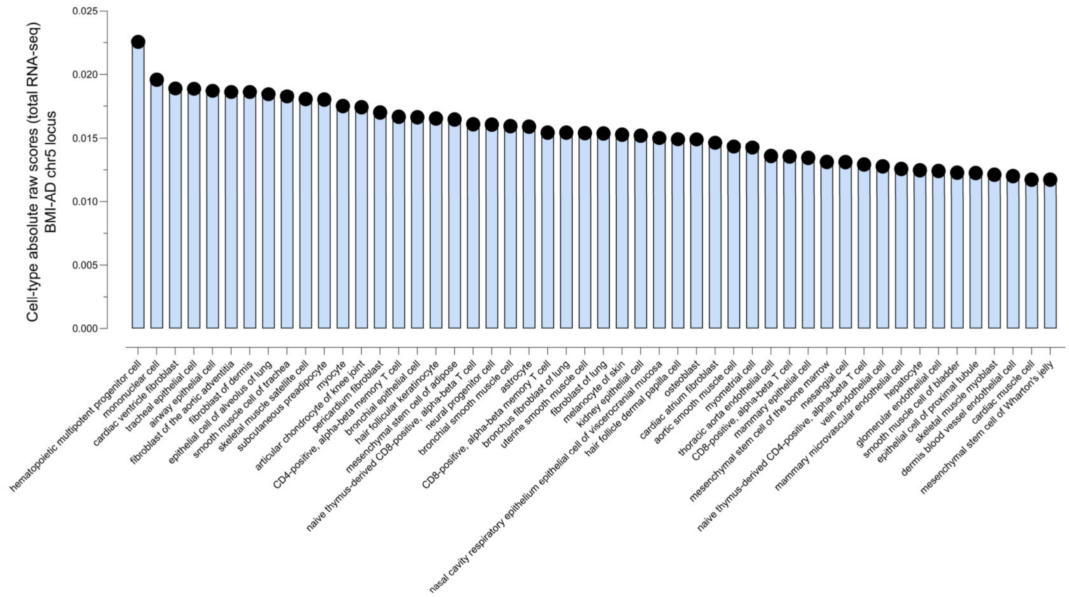


**Fig. S3**K | **AlphaGenome**. Top tissue predictions for the effects of rs2452758 in gene expression assessed by total RNA-seq.

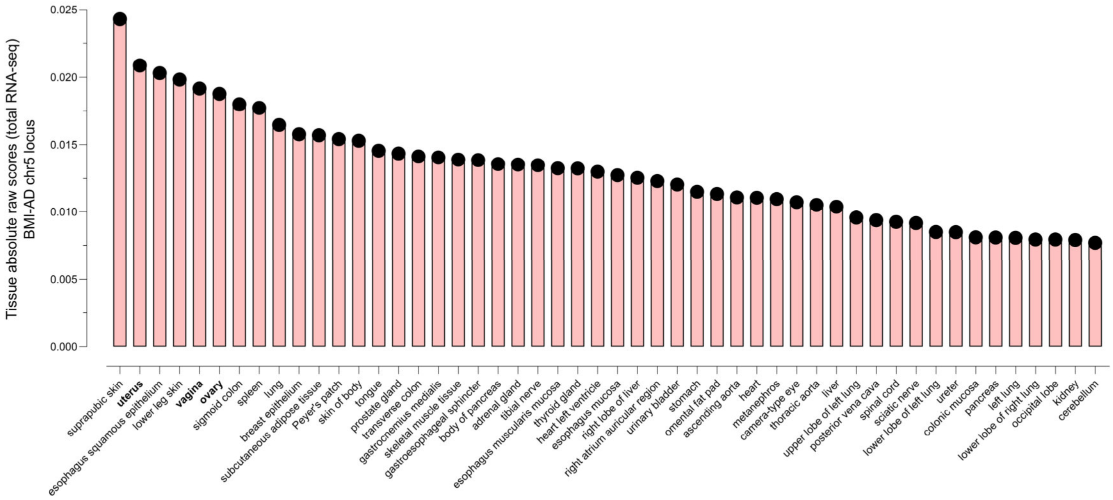


**Fig. S3**L | **AlphaGenome**. Top tissue predictions for the effects of rs2452758 in gene expression assessed by total RNA-seq.

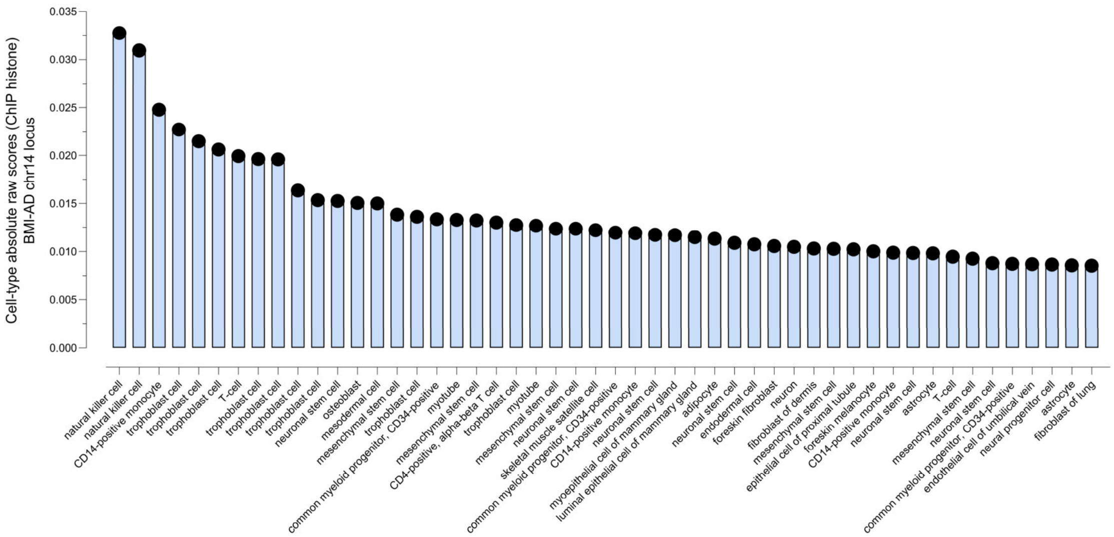


**Fig. S3**M | **AlphaGenome**. Top cell-type predictions for the effects of rs17125924 in histone modifications.

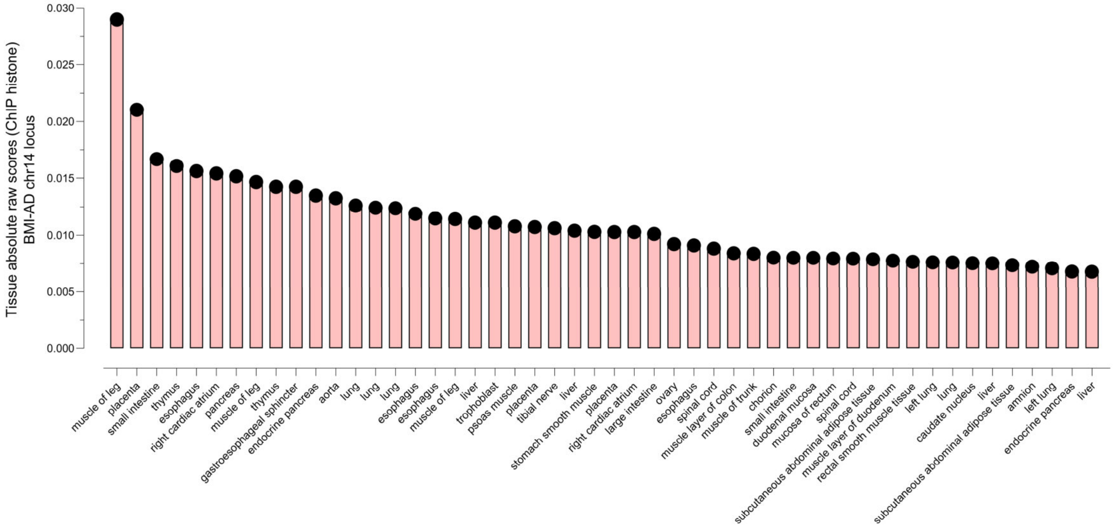


**Fig. S3**N | **AlphaGenome**. Top tissue predictions for the effects of rs17125924 in histone modifications.

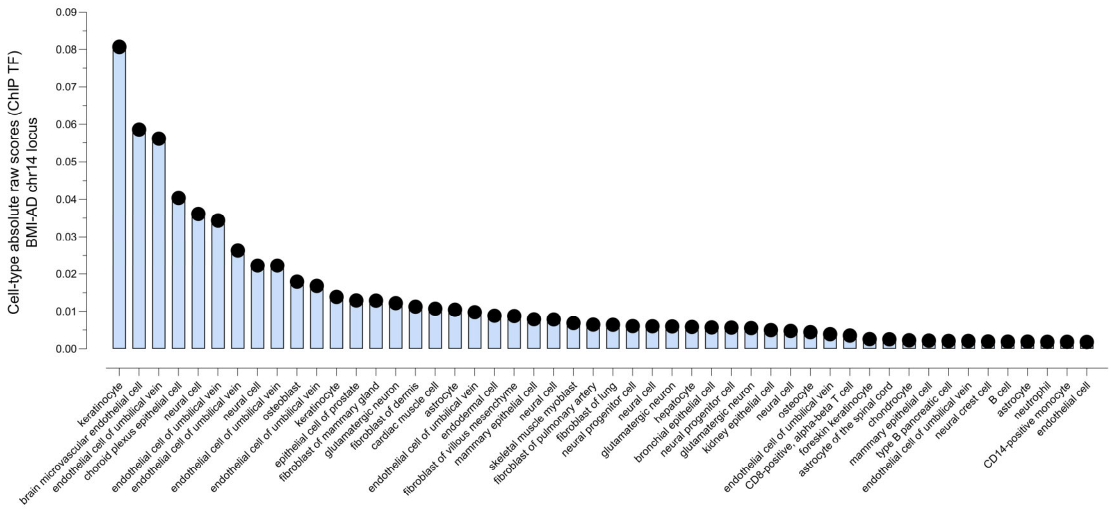


**Fig. S3**O | **AlphaGenome**. Top cell-type predictions for the effects of rs17125924 in transcription factor binding.

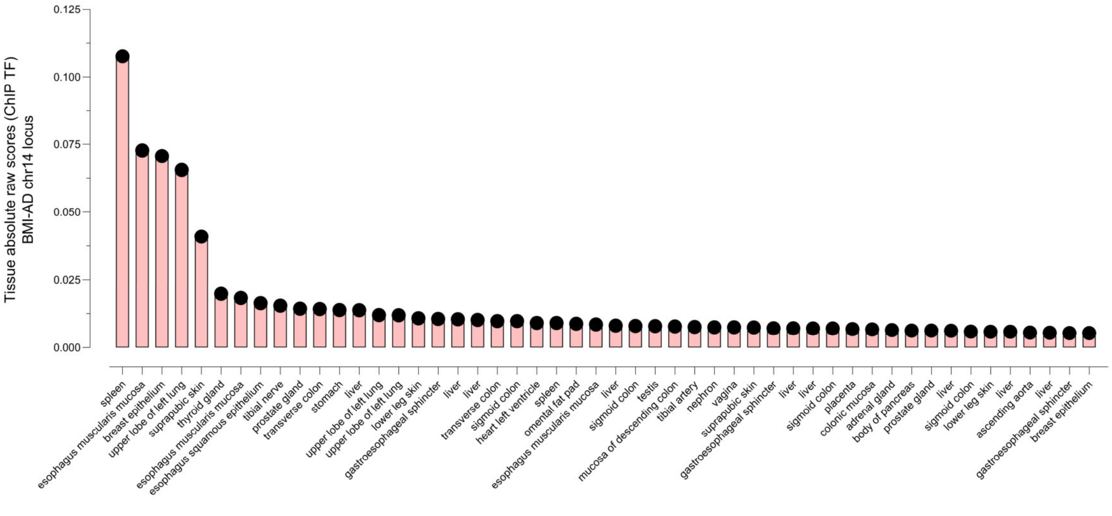


**Fig. S3**P | **AlphaGenome**. Top tissue predictions for the effects of rs17125924 in transcription factor binding.

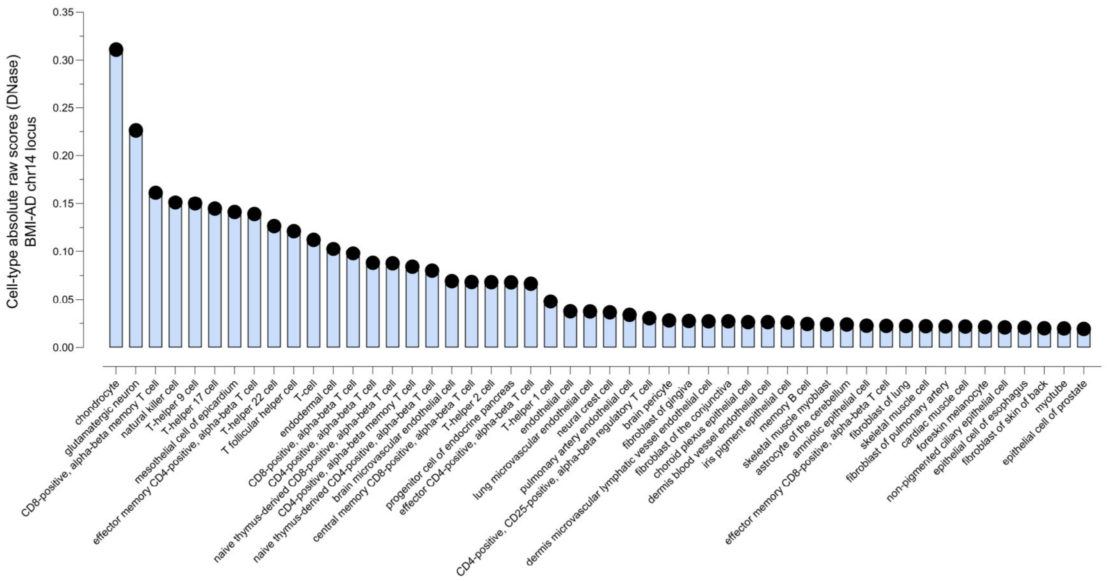


**Fig. S3**Q | **AlphaGenome**. Top cell-type predictions for the effects of rs17125924 in chromatin accessibility assessed by DNase.

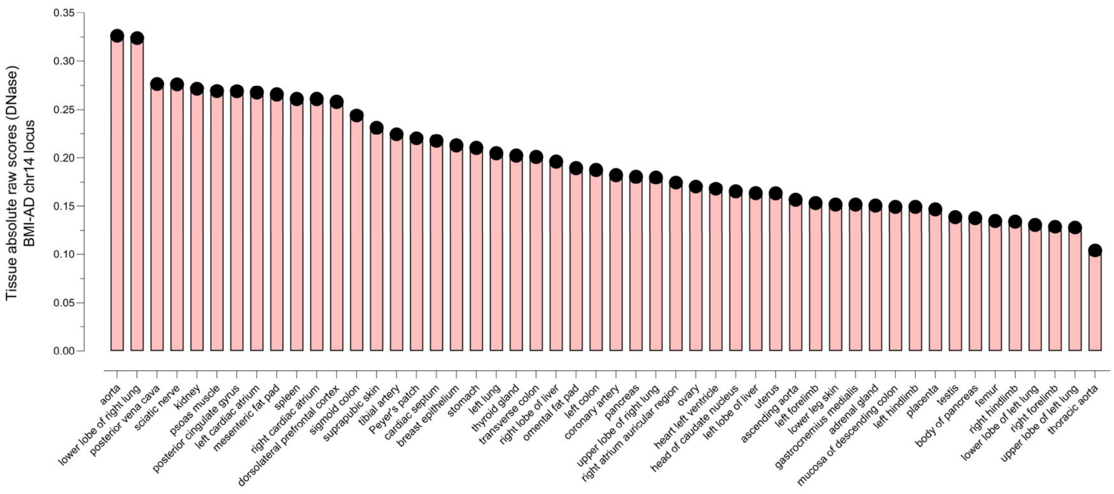


**Fig. S3**R | **AlphaGenome**. Top tissue predictions for the effects of rs17125924 in chromatin accessibility assessed by DNase.

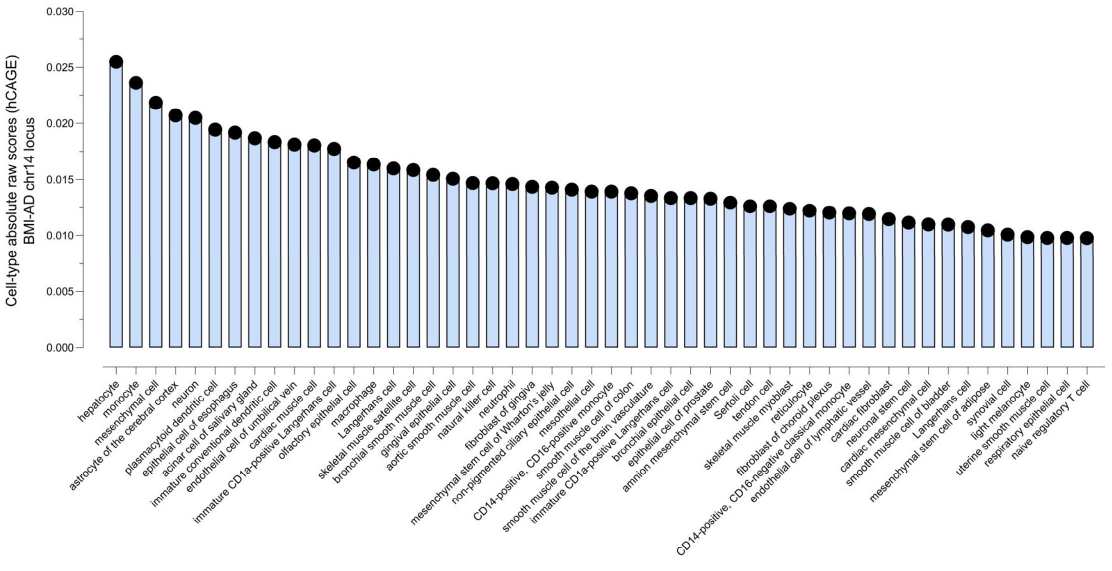


**Fig. S3**S | **AlphaGenome**. Top cell-type predictions for the effects of rs17125924 in gene expression assessed by hCAGE.

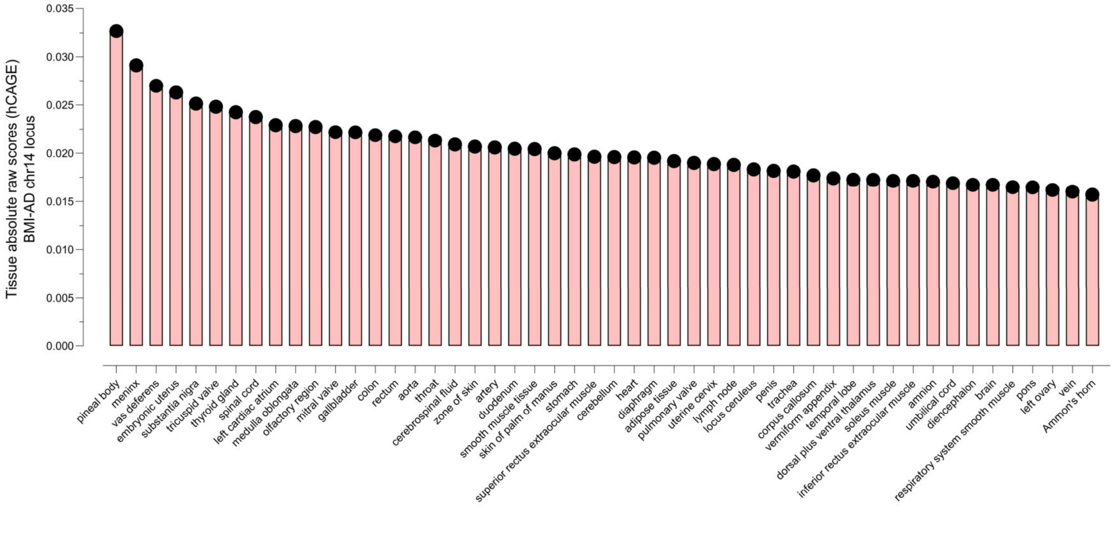


**Fig. S3**T | **AlphaGenome**. Top tissue predictions for the effects of rs17125924 in gene expression assessed by hCAGE.

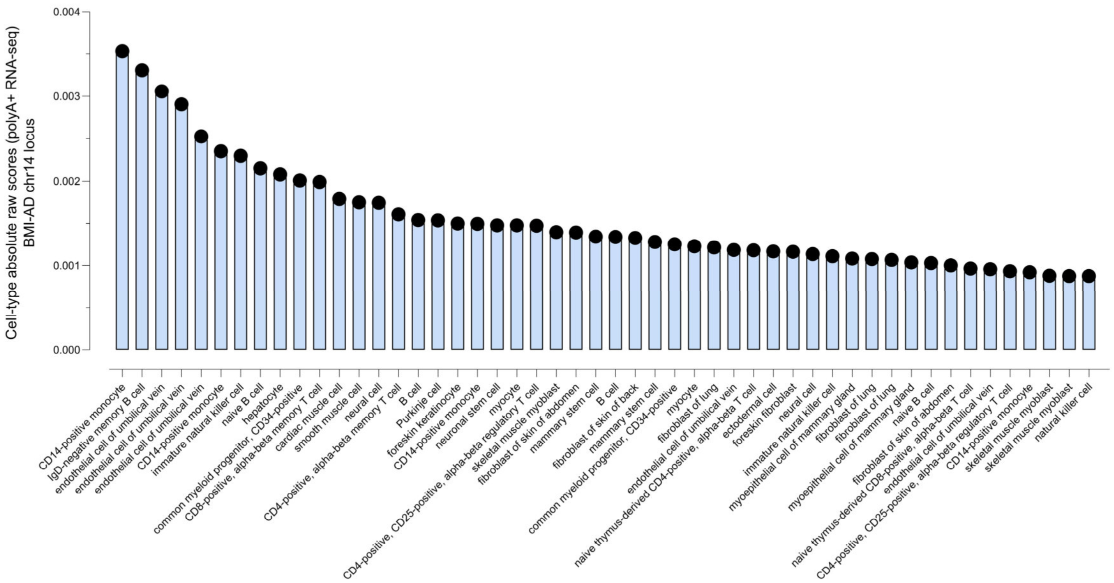


**Fig. S3**U | **AlphaGenome**. Top cell-type predictions for the effects of rs17125924 in gene expression assessed by polyA+ RNA-seq.

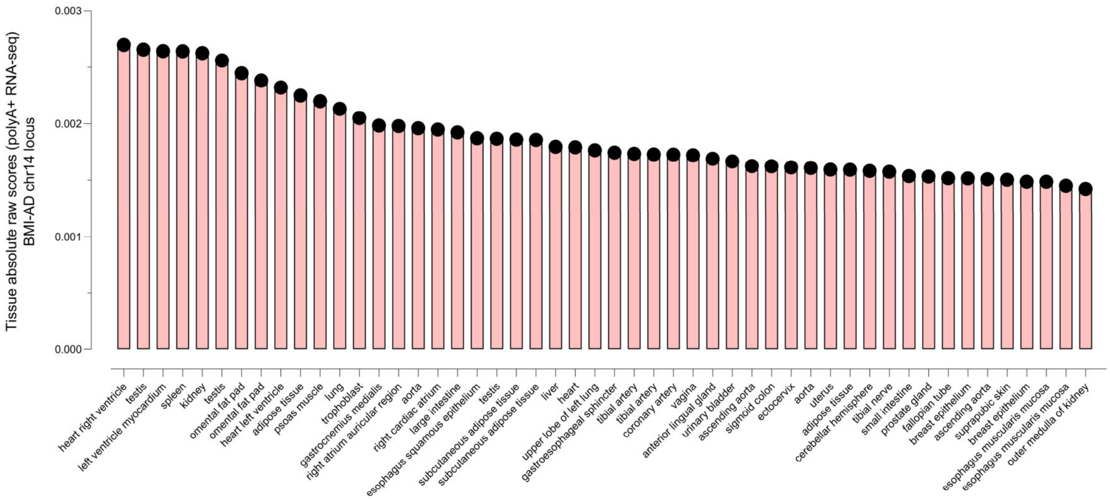


**Fig. S3**V | **AlphaGenome**. Top tissue predictions for the effects of rs17125924 in gene expression assessed by polyA+ RNA-seq.

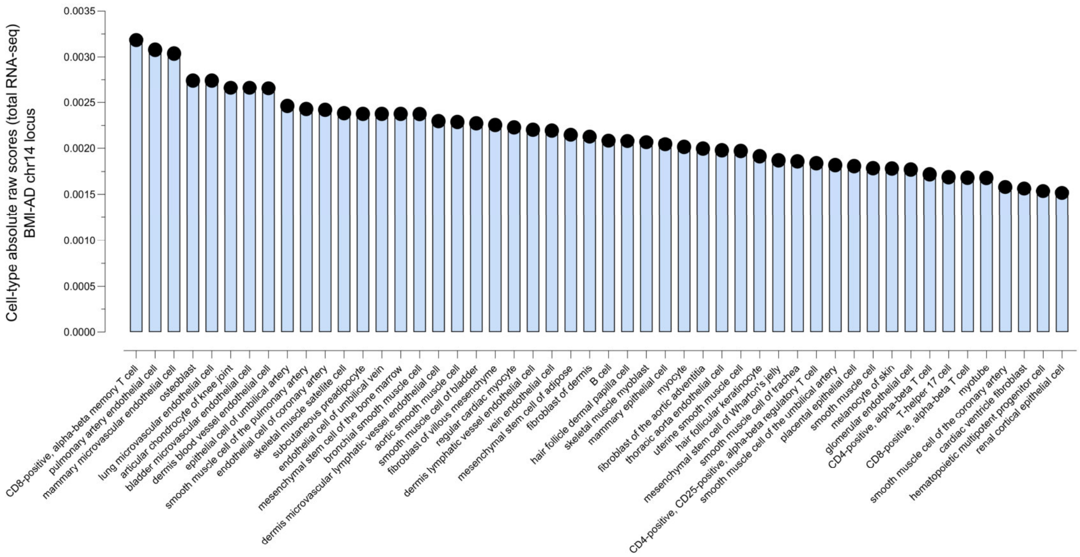


**Fig. S3**W | **AlphaGenome**. Top cell-type predictions for the effects of rs17125924 in gene expression assessed by total RNA-seq.

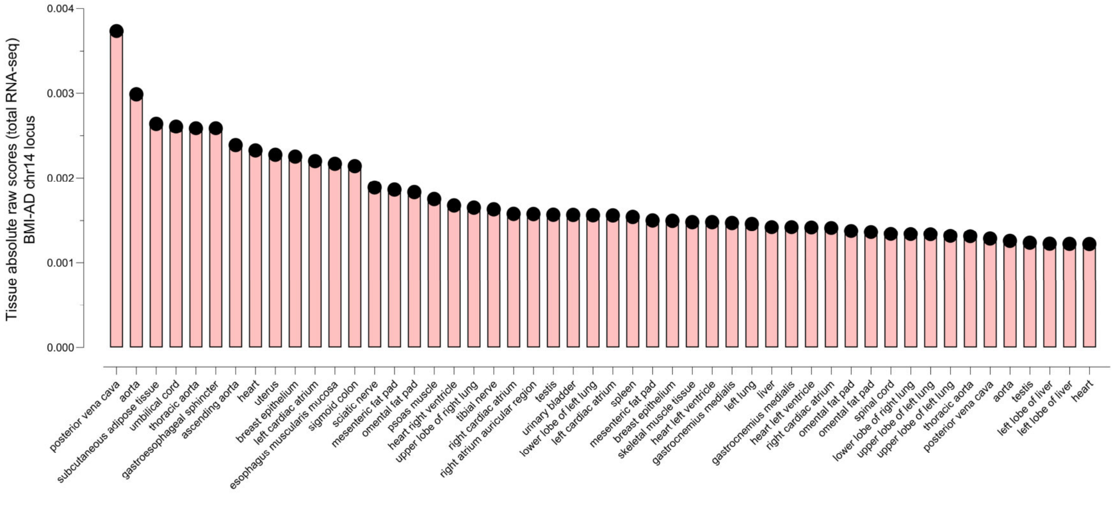


**Fig. S3**X | **AlphaGenome**. Top tissue predictions for the effects of rs17125924 in gene expression assessed by total RNA-seq.

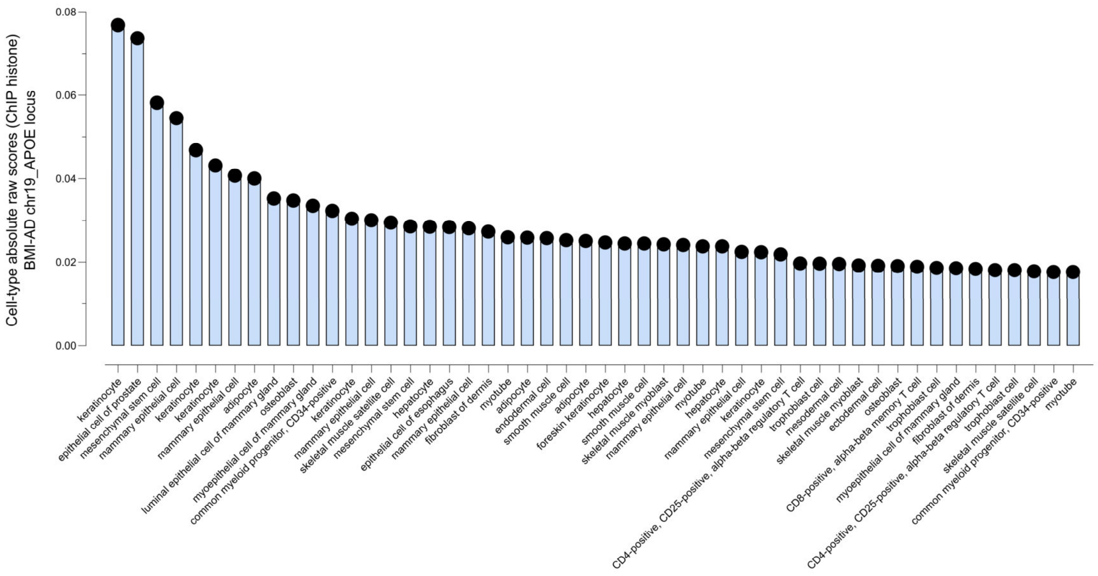


**Fig. S3**Y | **AlphaGenome**. Top cell-type predictions for the effects of rs7254892 in histone modifications.

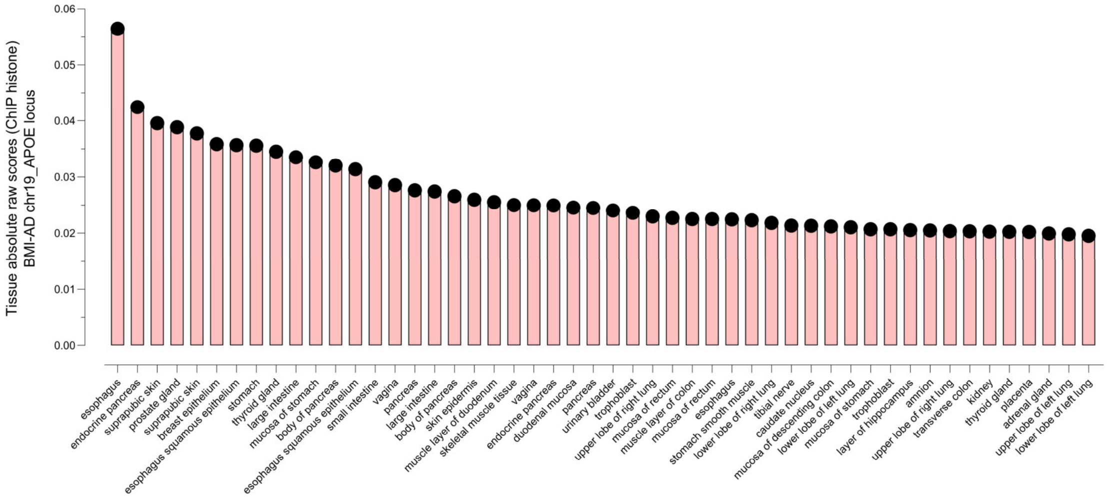


**Fig. S3**Z | **AlphaGenome**. Top tissue predictions for the effects of rs7254892 in histone modifications.

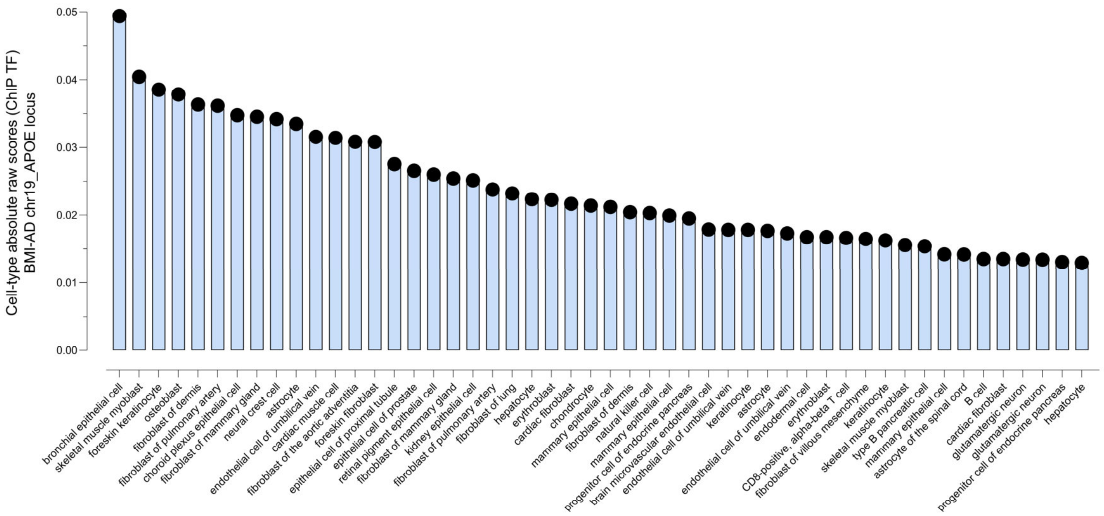


**Fig. S3**A1 | **AlphaGenome**. Top cell-type predictions for the effects of rs7254892 in transcription factor binding.

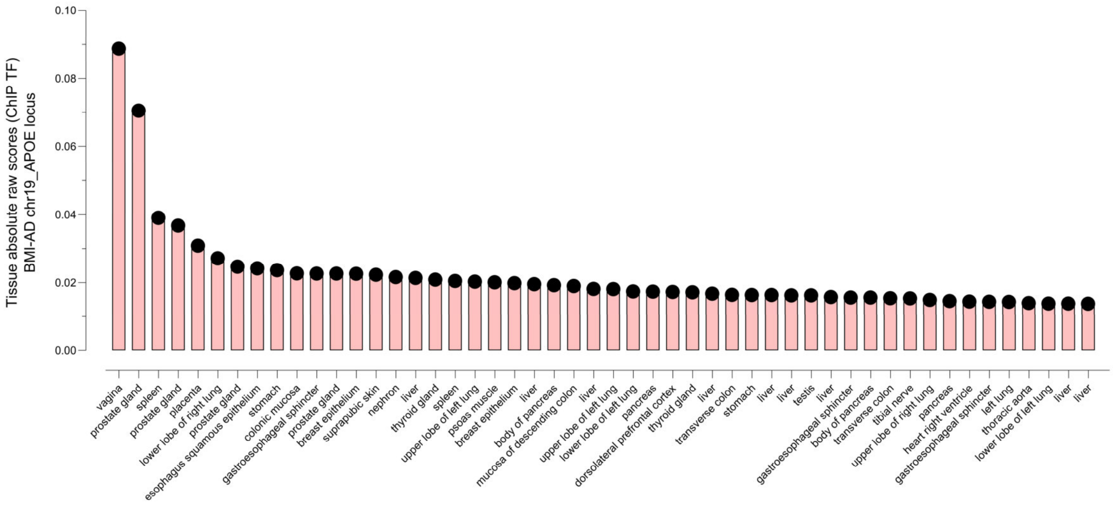


**Fig. S3**B1 | **AlphaGenome**. Top tissue predictions for the effects of rs7254892 in transcription factor binding.

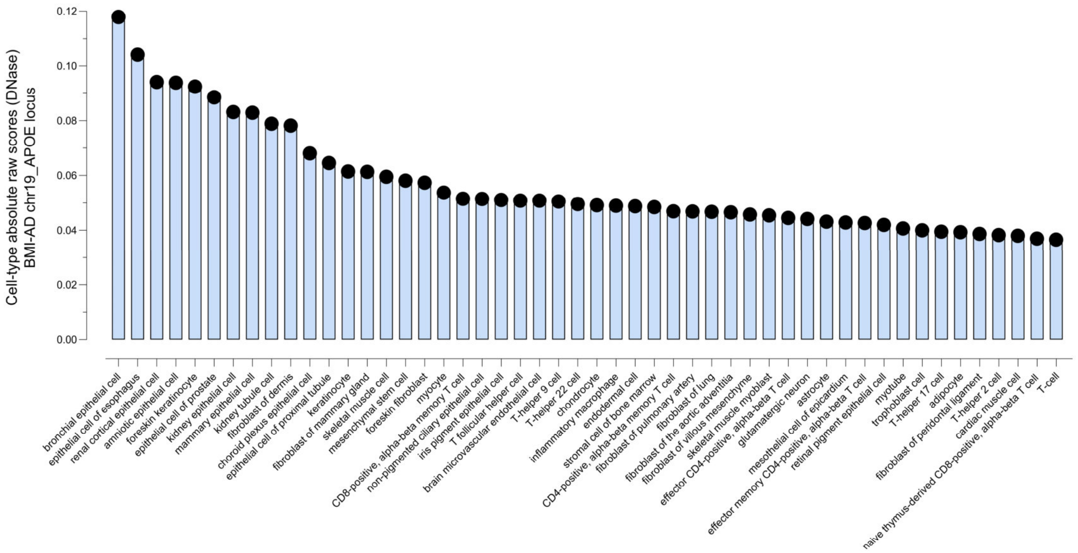


**Fig. S3**C1 | **AlphaGenome**. Top cell-type predictions for the effects of rs7254892 in chromatin accessibility assessed by DNase.

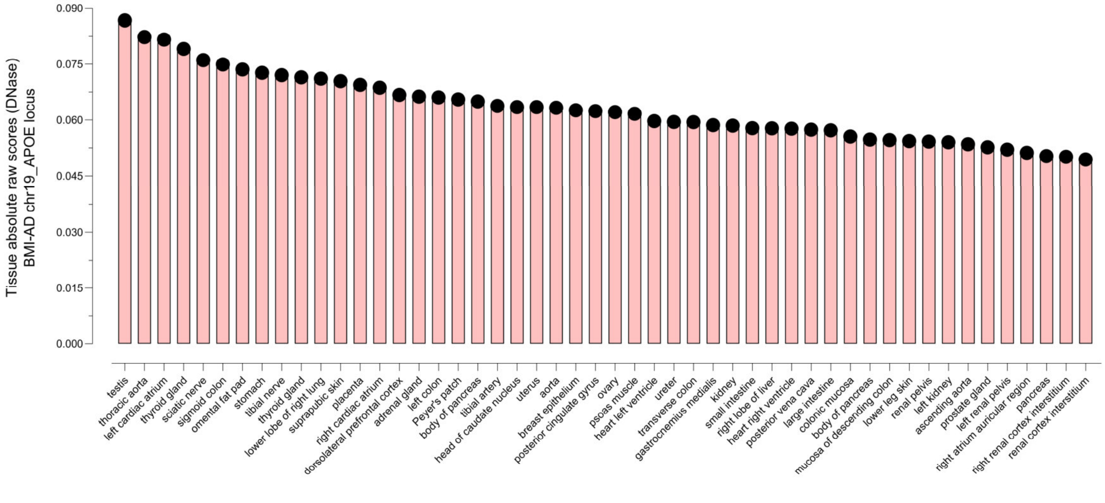


**Fig. S3**D1 | **AlphaGenome**. Top tissue predictions for the effects of rs7254892 in chromatin accessibility assessed by DNase.

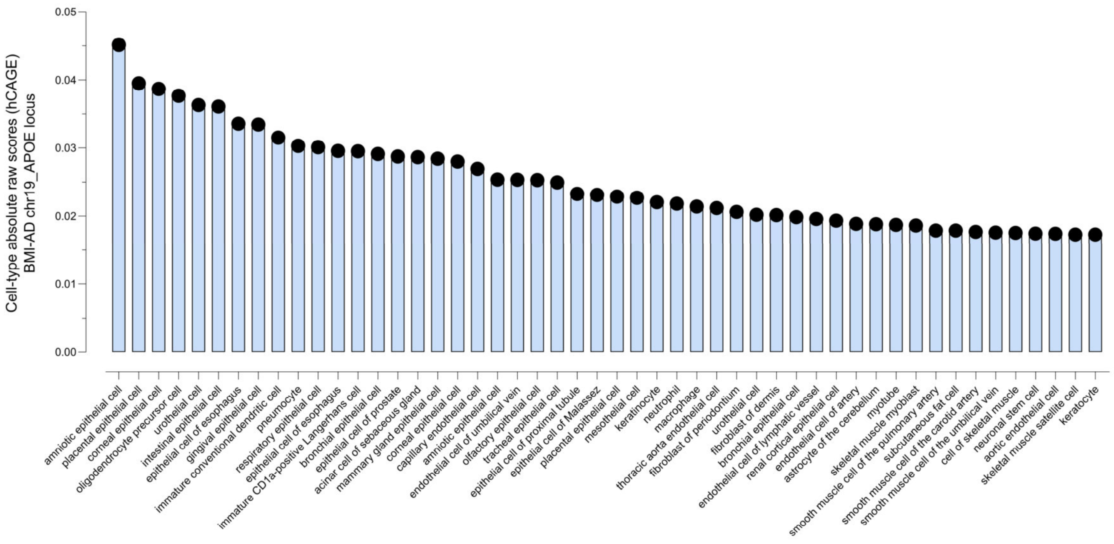


**Fig. S3**E1 | **AlphaGenome**. Top cell-type predictions for the effects of rs7254892 in gene expression assessed by hCAGE.

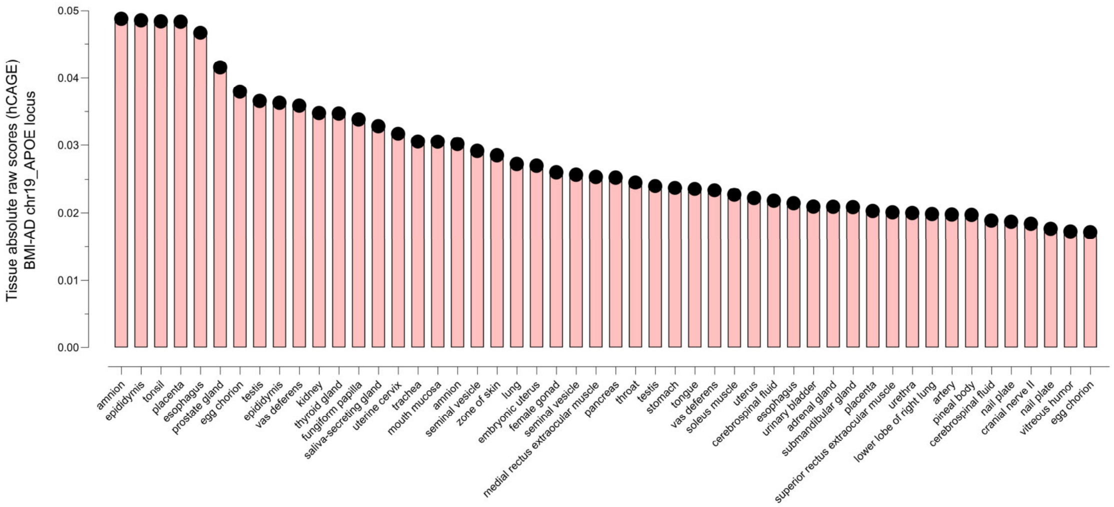


**Fig. S3**F1 | **AlphaGenome**. Top tissue predictions for the effects of rs7254892 in gene expression assessed by hCAGE.

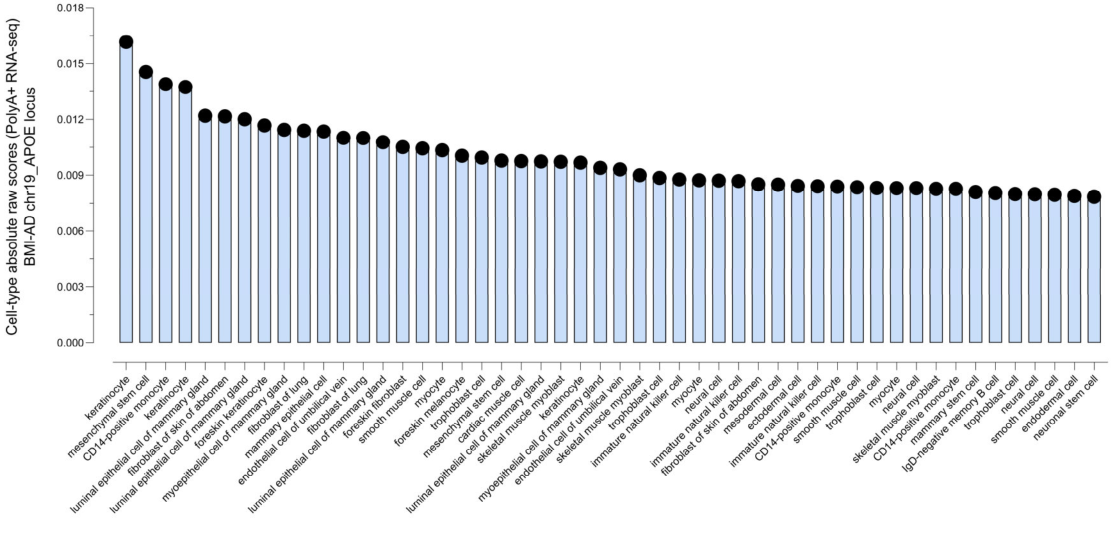


**Fig. S3**G1 | **AlphaGenome**. Top cell-type predictions for the effects of rs7254892 in gene expression assessed by polyA+ RNA-seq.

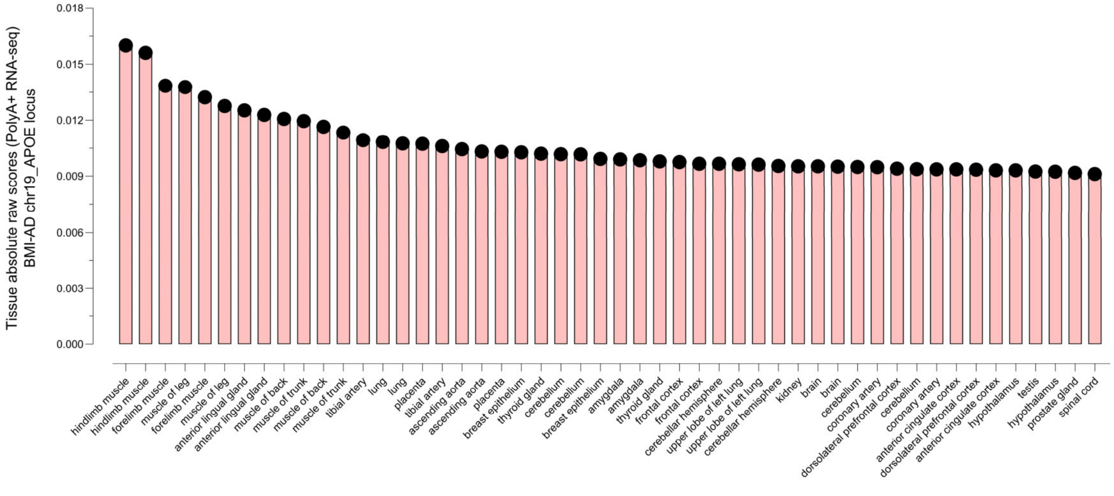


**Fig. S3**H1 | **AlphaGenome**. Top tissue predictions for the effects of rs7254892 in gene expression assessed by polyA+ RNA-seq.

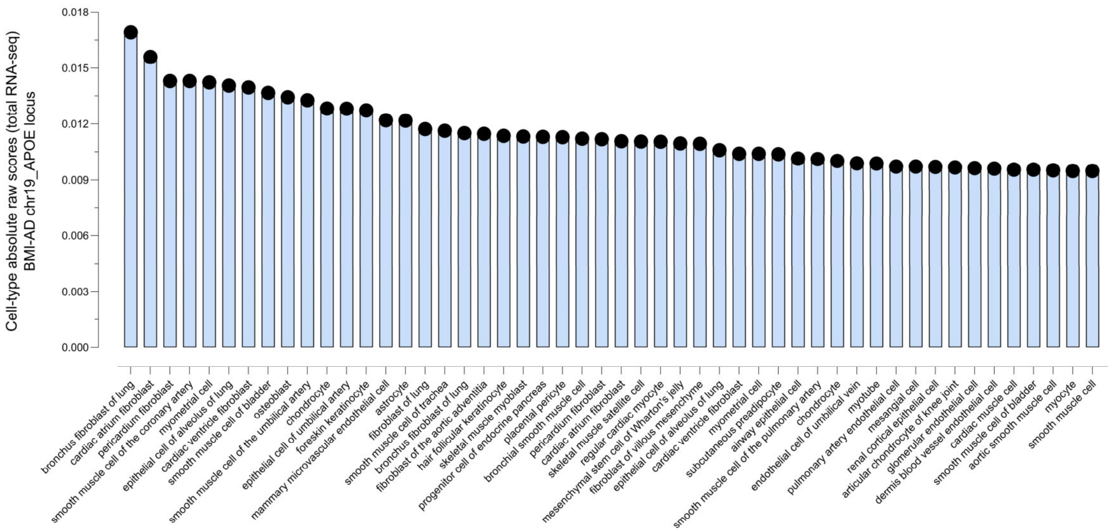


**Fig. S3**I1 | **AlphaGenome**. Top cell-type predictions for the effects of rs7254892 in gene expression assessed by total RNA-seq.

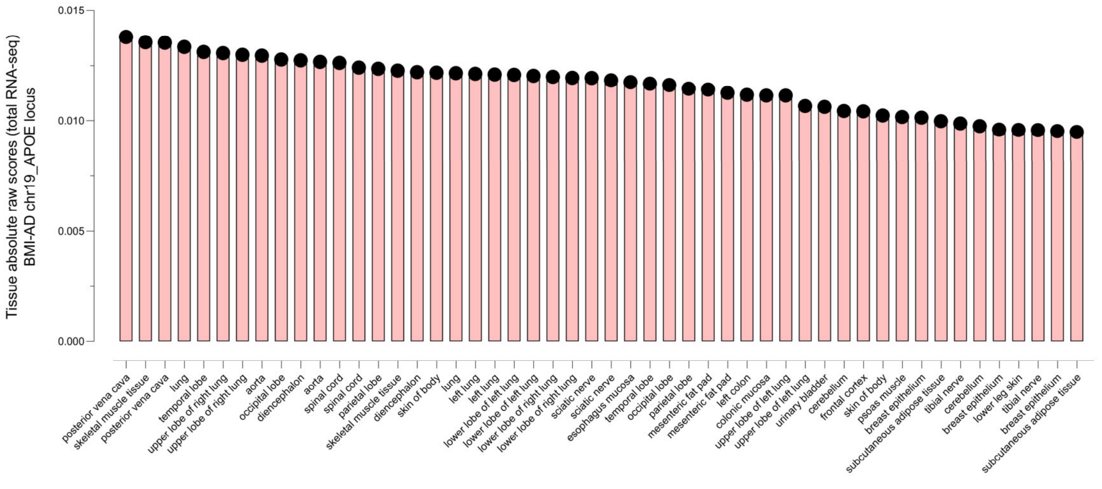


**Fig. S3**J1 | **AlphaGenome**. Top tissue predictions for the effects of rs7254892 in gene expression assessed by total RNA-seq.

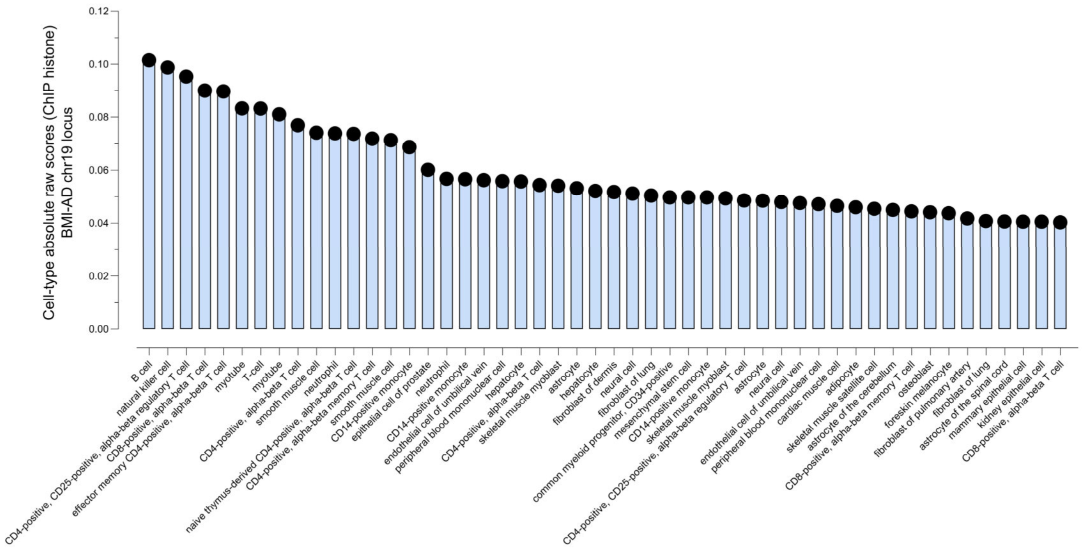


**Fig. S3**K1 | **AlphaGenome**. Top cell-type predictions for the effects of rs11668832 in histone modifications.

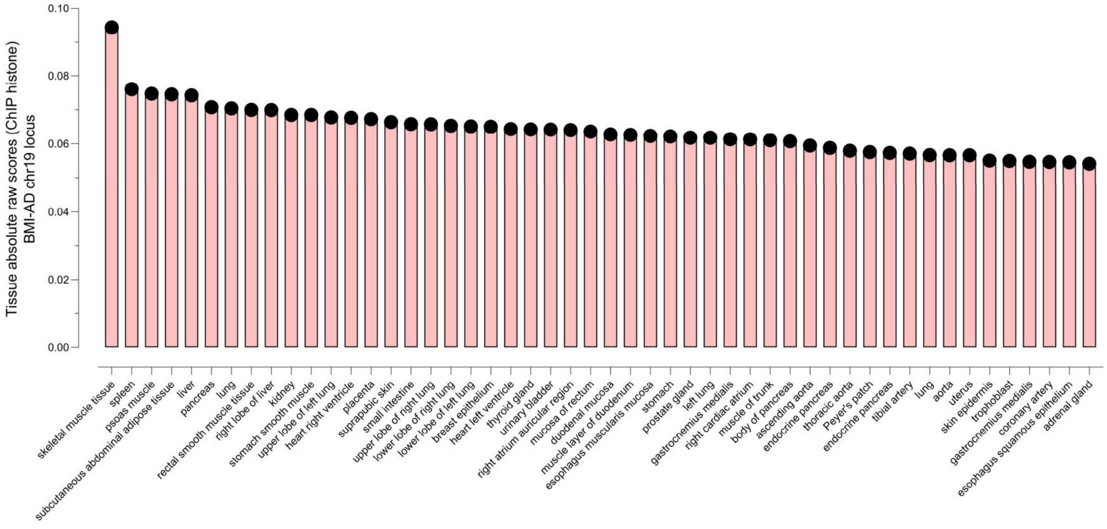


**Fig. S3**L1 | **AlphaGenome**. Top tissue predictions for the effects of rs11668832 in histone modifications.

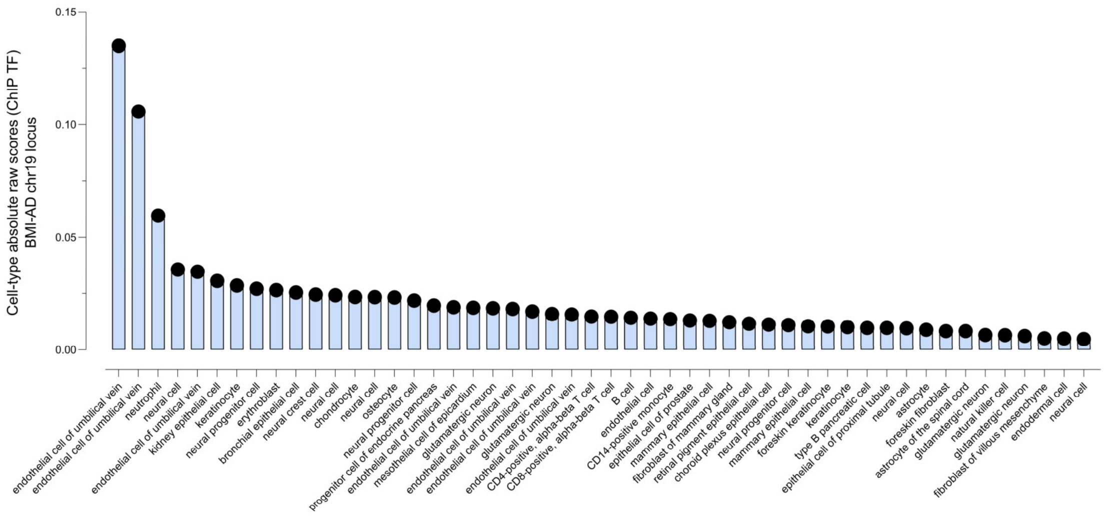


**Fig. S3**M1 | **AlphaGenome**. Top cell-type predictions for the effects of rs11668832 in transcription factor binding.

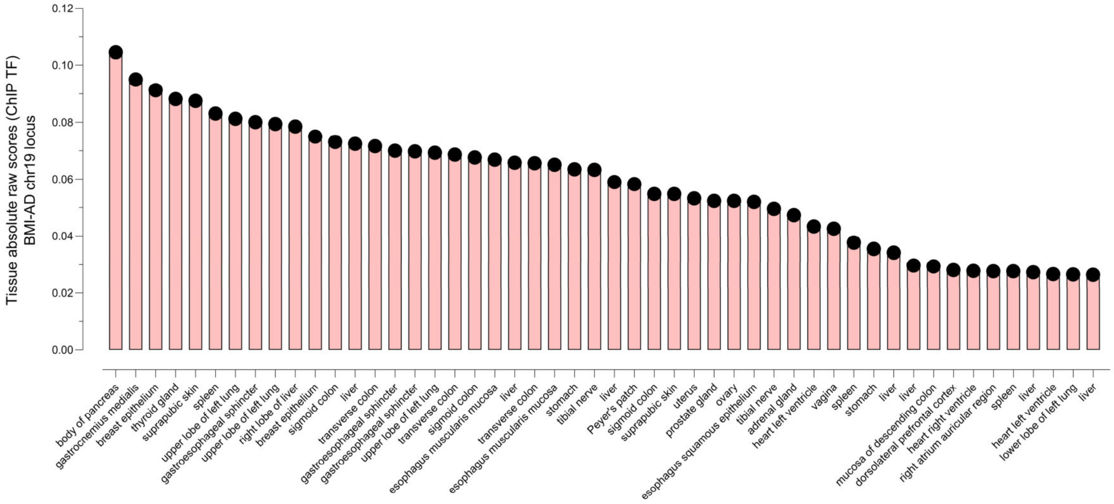


**Fig. S3**N1 | **AlphaGenome**. Top tissue predictions for the effects of rs11668832 in transcription factor binding.

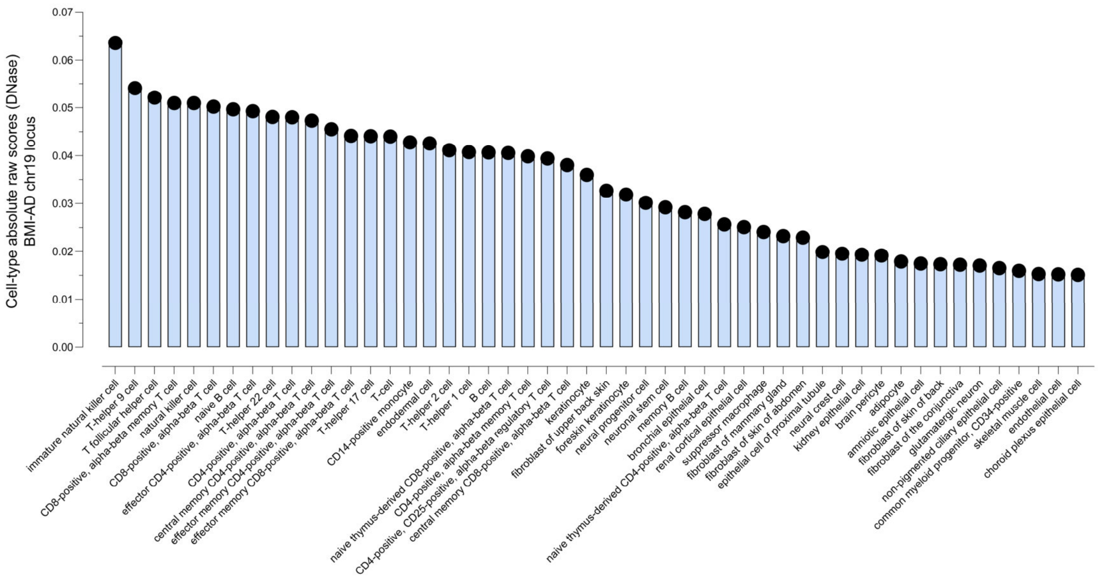


**Fig. S3**O1 | **AlphaGenome**. Top cell-type predictions for the effects of rs11668832 in chromatin accessibility assessed by DNase.

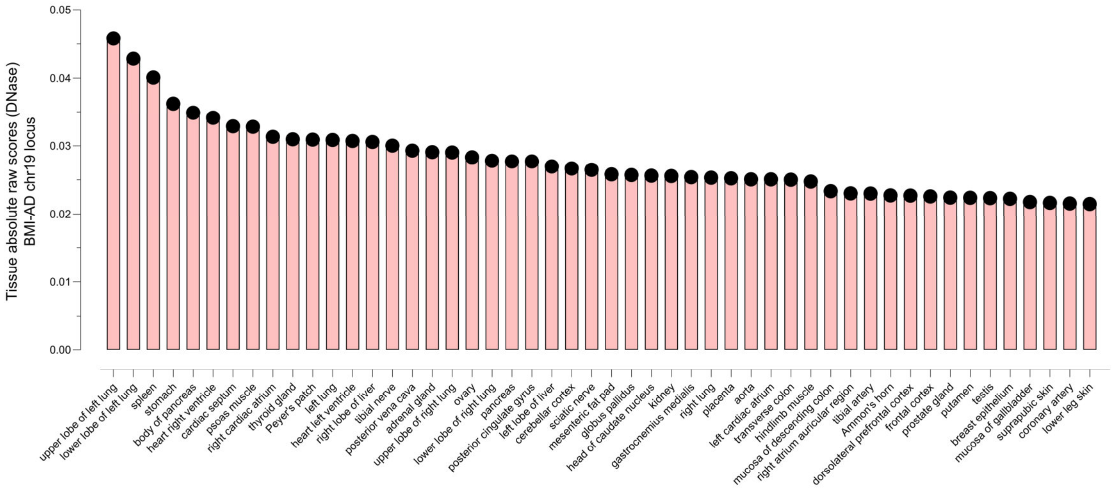


**Fig. S3**P1 | **AlphaGenome**. Top tissue predictions for the effects of rs11668832 in chromatin accessibility assessed by DNase.

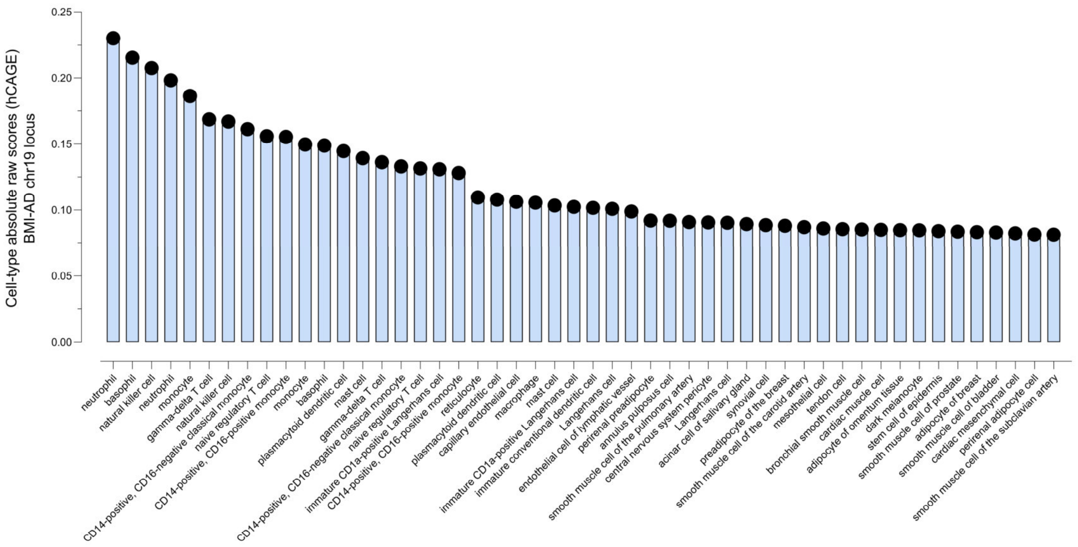


**Fig. S3**Q1 | **AlphaGenome**. Top cell-type predictions for the effects of rs11668832 in gene expression assessed by hCAGE.

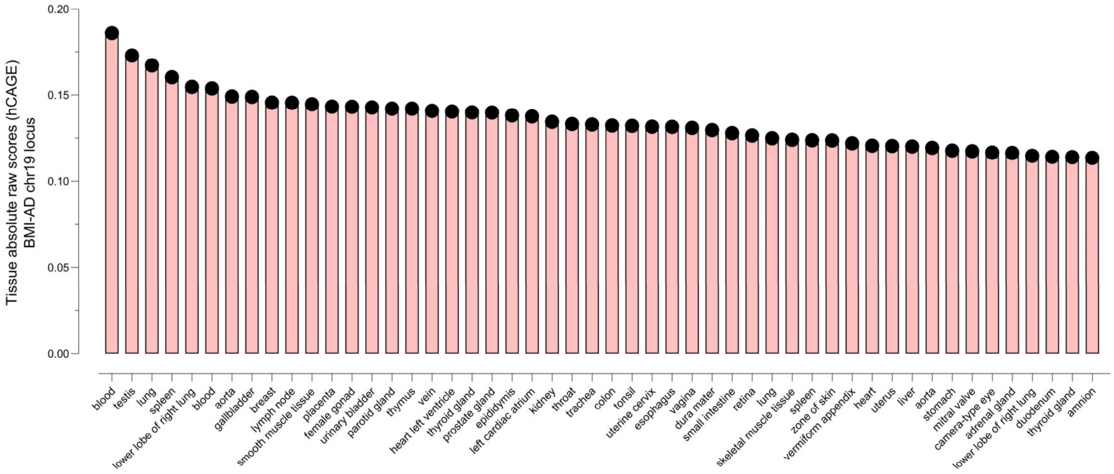


**Fig. S3**R1 | **AlphaGenome**. Top tissue predictions for the effects of rs11668832 in gene expression assessed by hCAGE

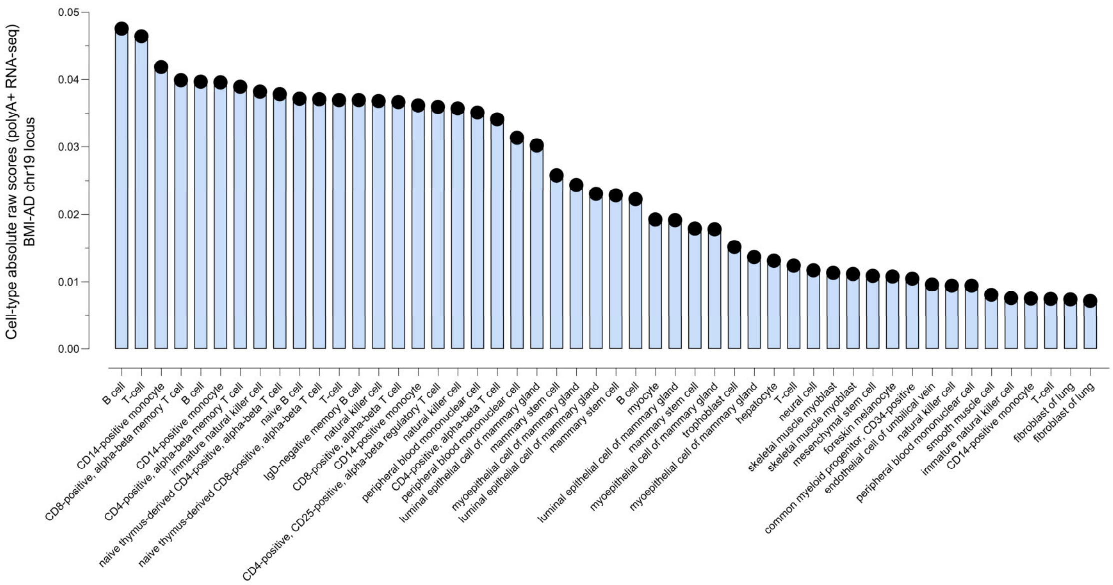


**Fig. S3**S1 | **AlphaGenome**. Top cell-type predictions for the effects of rs11668832 in gene expression assessed by polyA+ RNA-seq.

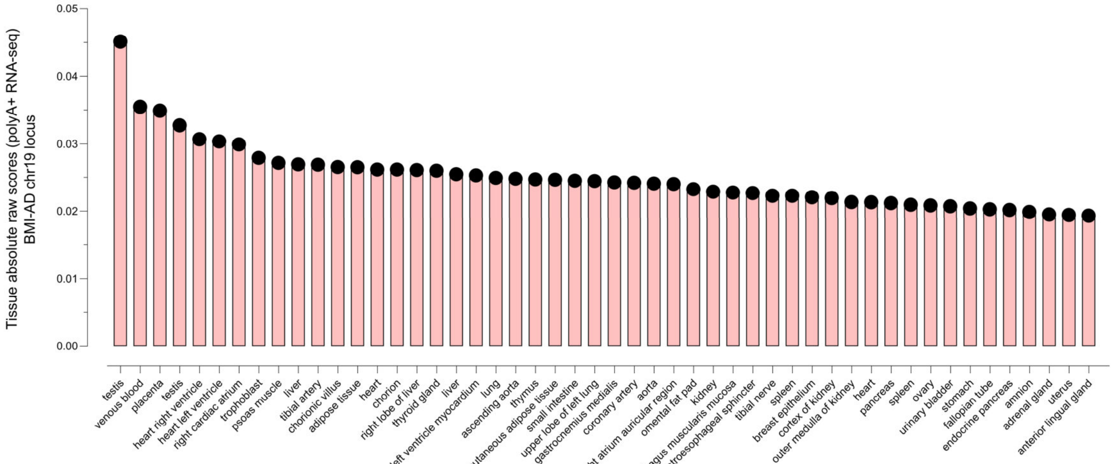


**Fig. S3**T1 | **AlphaGenome**. Top tissue predictions for the effects of rs11668832 in gene expression assessed by polyA+ RNA-seq.

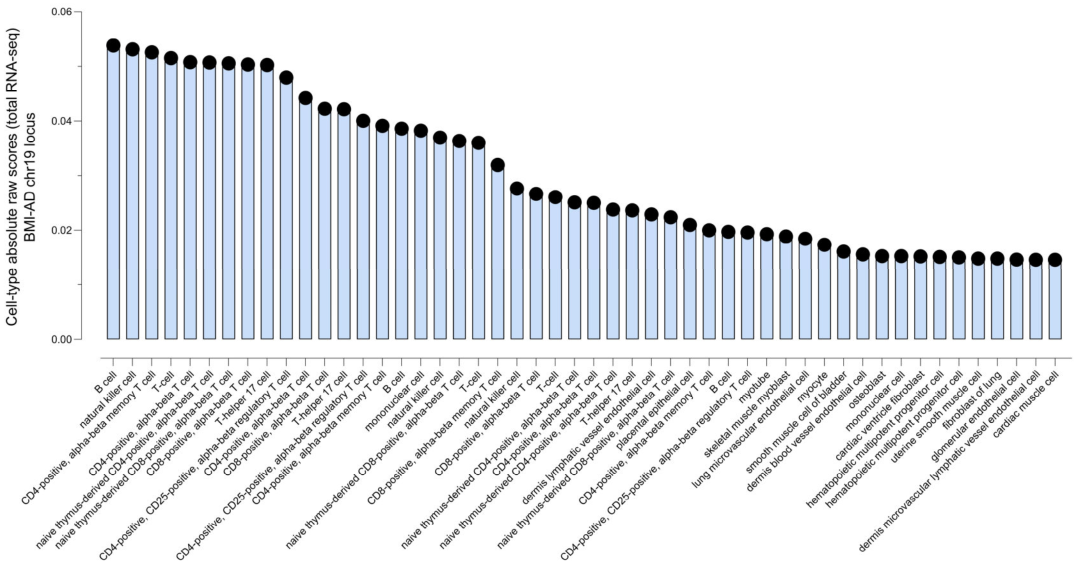


**Fig. S3**U1 | **AlphaGenome**. Top cell-type predictions for the effects of rs11668832 in gene expression assessed by total RNA-seq.

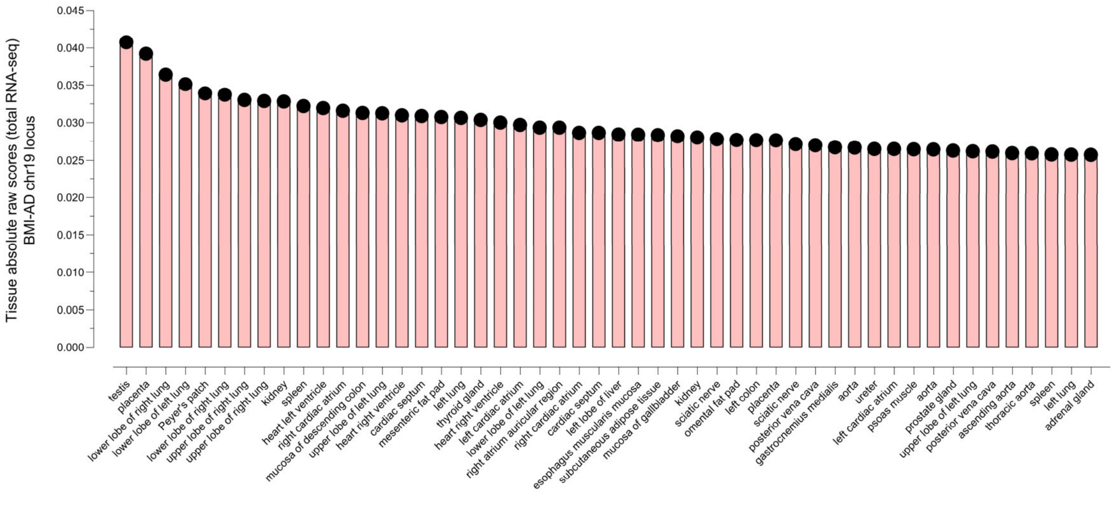


**Fig. S3**U1 | **AlphaGenome**. Top tissue predictions for the effects of rs11668832 in gene expression assessed by total RNA-seq.

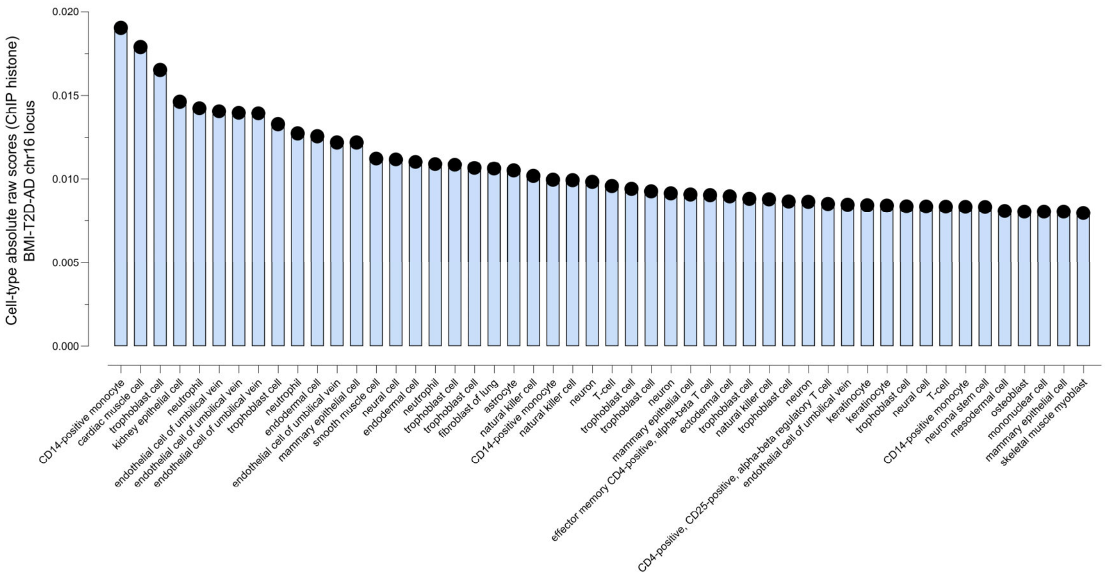


**Fig. S3**V1 | **AlphaGenome**. Top cell-type predictions for the effects of rs11150577 in histone modifications.

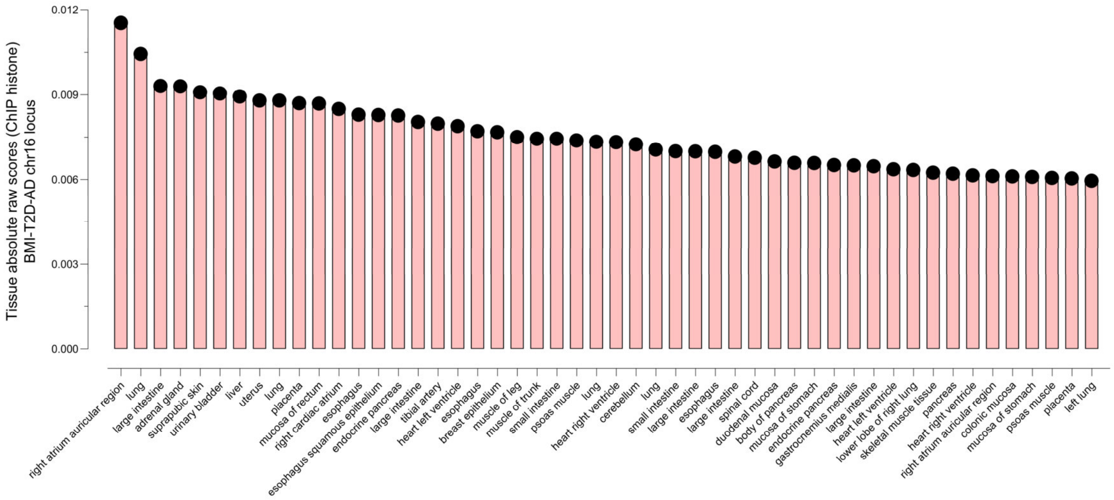


**Fig. S3**W1 | **AlphaGenome**. Top tissue predictions for the effects of rs11150577 in histone modifications.

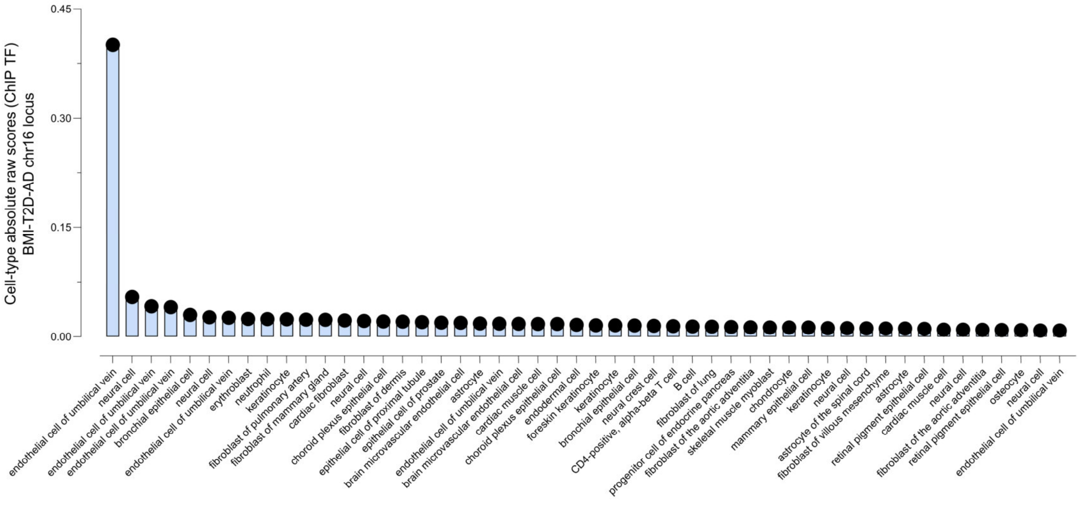


**Fig. S3**X1 | **AlphaGenome**. Top cell-type predictions for the effects of rs11150577 in transcription factor binding.

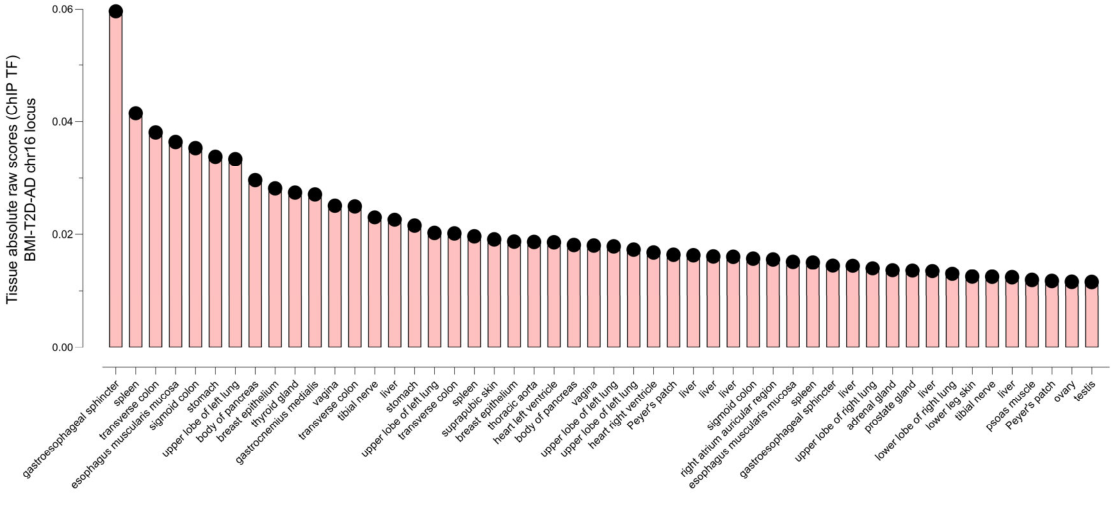


**Fig. S3**Y1 | **AlphaGenome**. Top cell-type predictions for the effects of rs11150577 in transcription factor binding.

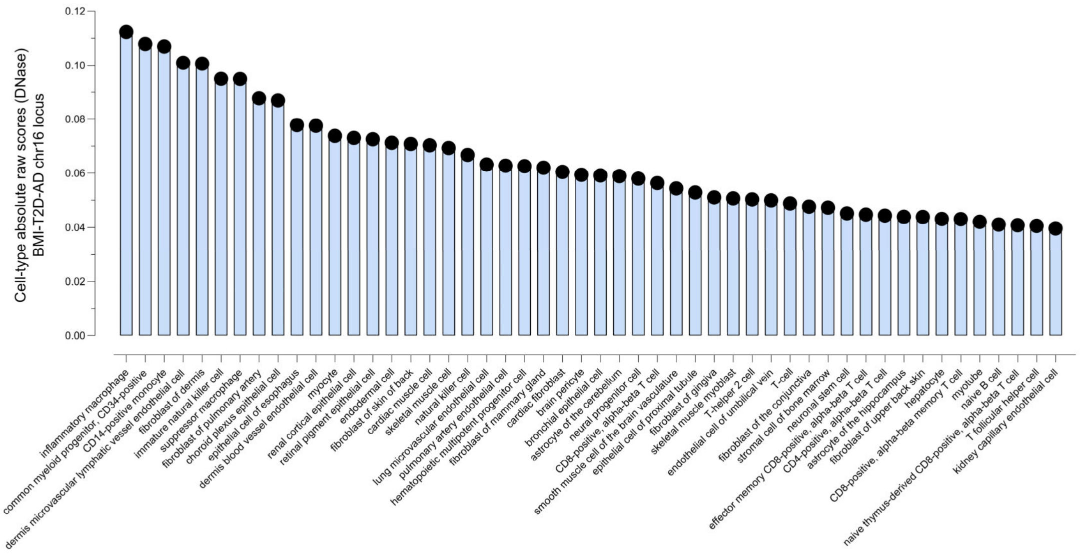


**Fig. S3**Z1 | **AlphaGenome**. Top cell-type predictions for the effects of rs11150577 in chromatin accessibility assessed by DNase.

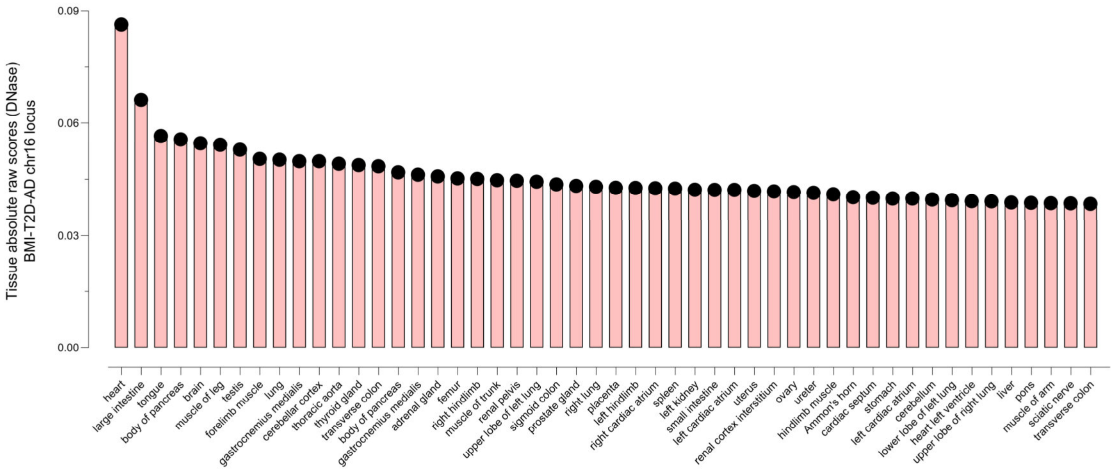


**Fig. S3**A2 | **AlphaGenome**. Top tissue predictions for the effects of rs11150577 in chromatin accessibility assessed by DNase.

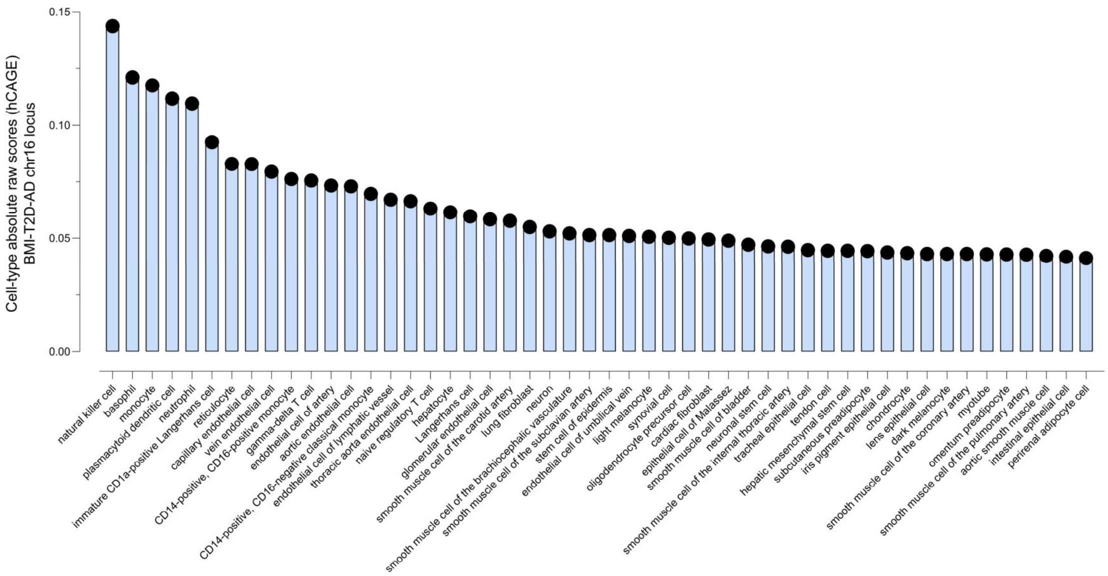


**Fig. S3**B2 | **AlphaGenome**. Top cell-type predictions for the effects of rs11150577 in gene expression assessed by hCAGE.

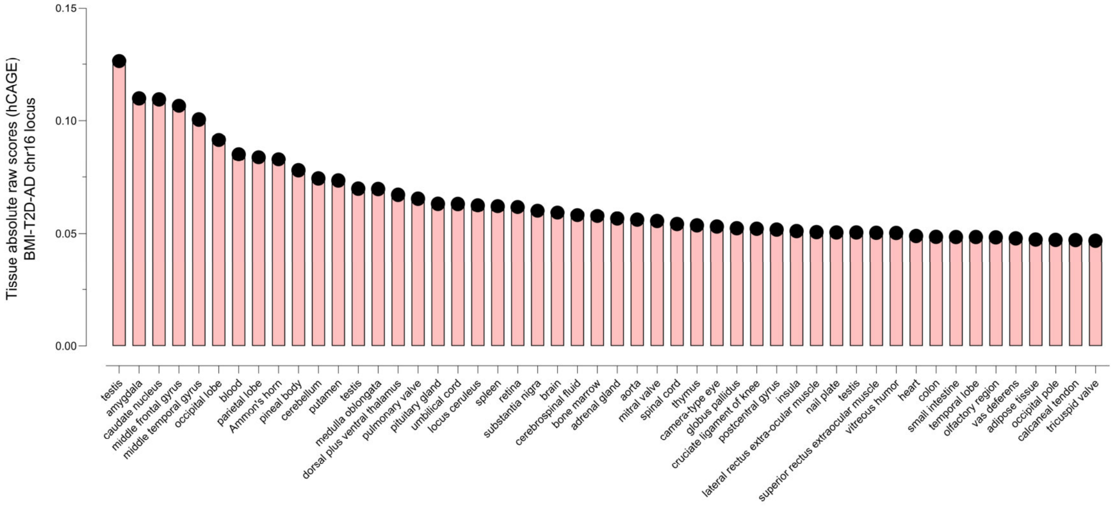


**Fig. S3**C2 | **AlphaGenome**. Top tissue predictions for the effects of rs11150577 in gene expression assessed by hCAGE.

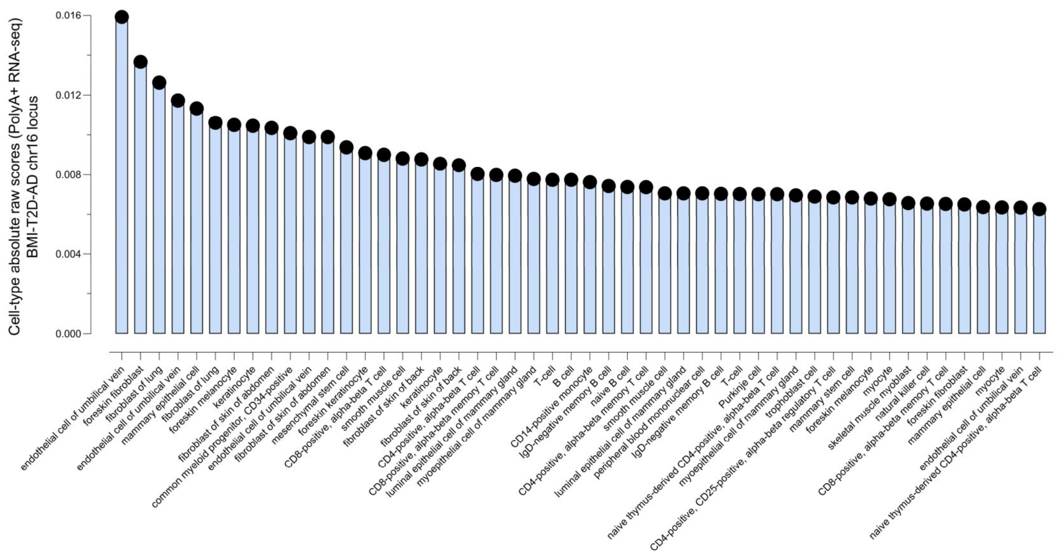


**Fig. S3**D2 | **AlphaGenome**. Top cell-type predictions for the effects of rs11150577 in gene expression assessed by polyA+ RNA-seq.

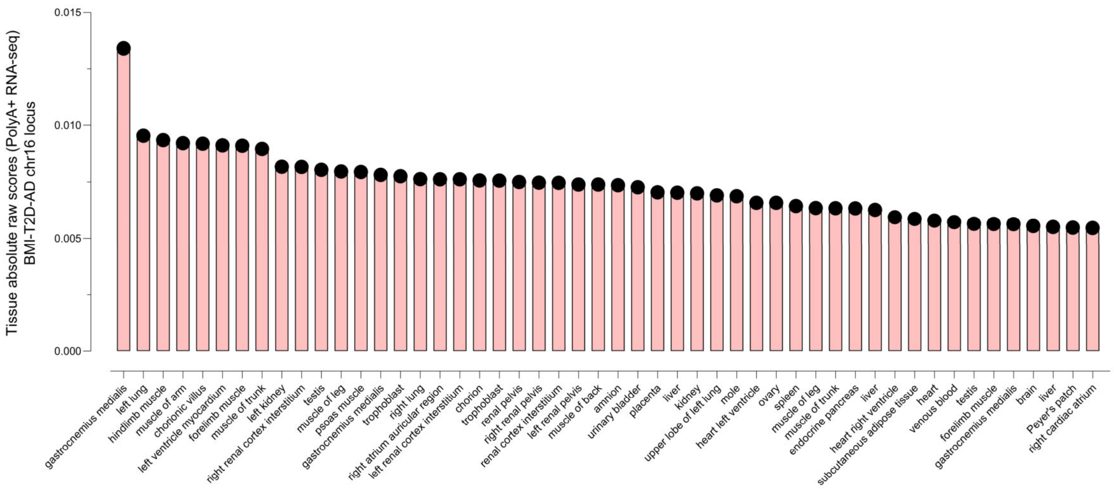


**Fig. S3**E2 | **AlphaGenome**. Top tissue predictions for the effects of rs11150577 in gene expression assessed by polyA+ RNA-seq.

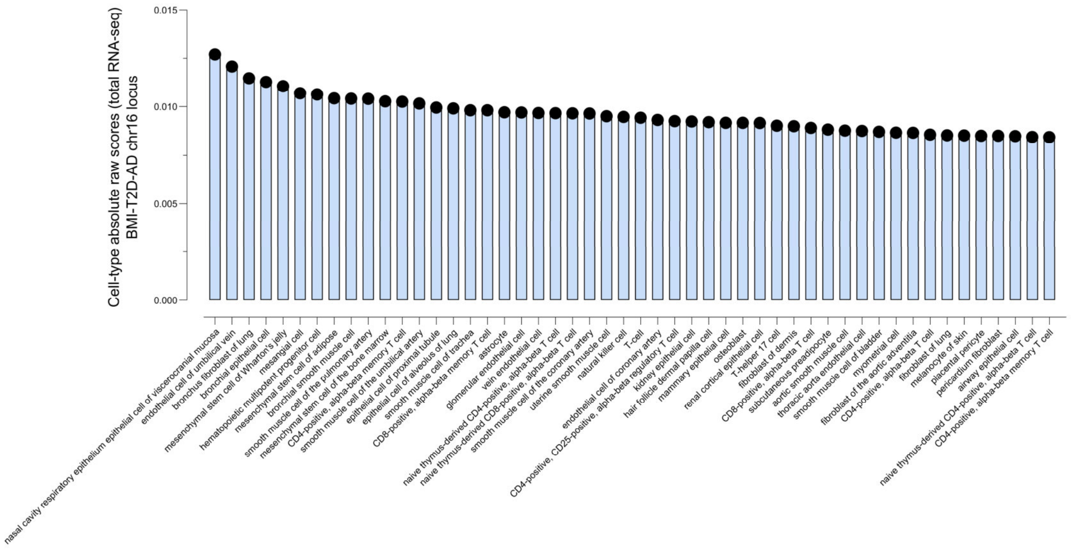


**Fig. S3**F2 | **AlphaGenome**. Top cell-type predictions for the effects of rs11150577 in gene expression assessed by total RNA-seq.

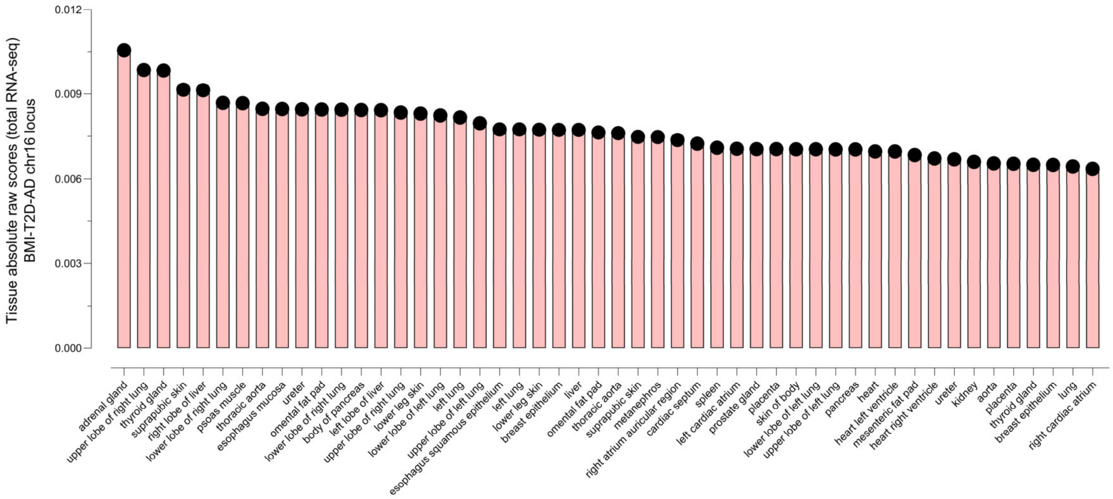


**Fig. S3**G2 | **AlphaGenome**. Top tissue predictions for the effects of rs11150577 in gene expression assessed by total RNA-seq.

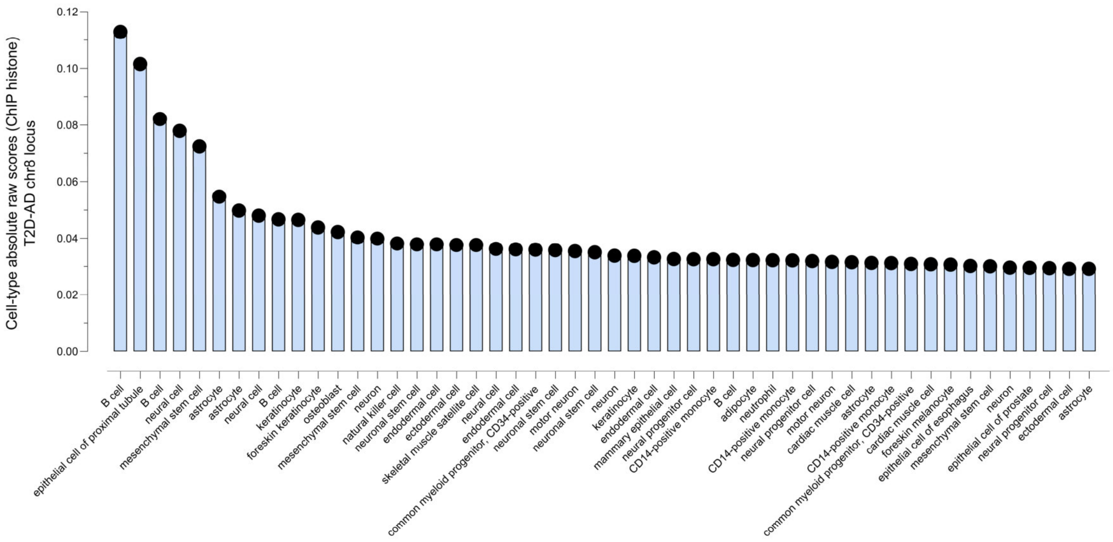


**Fig. S3**H2 | **AlphaGenome**. Top cell-type predictions for the effects of rs13257021 in histone modifications.

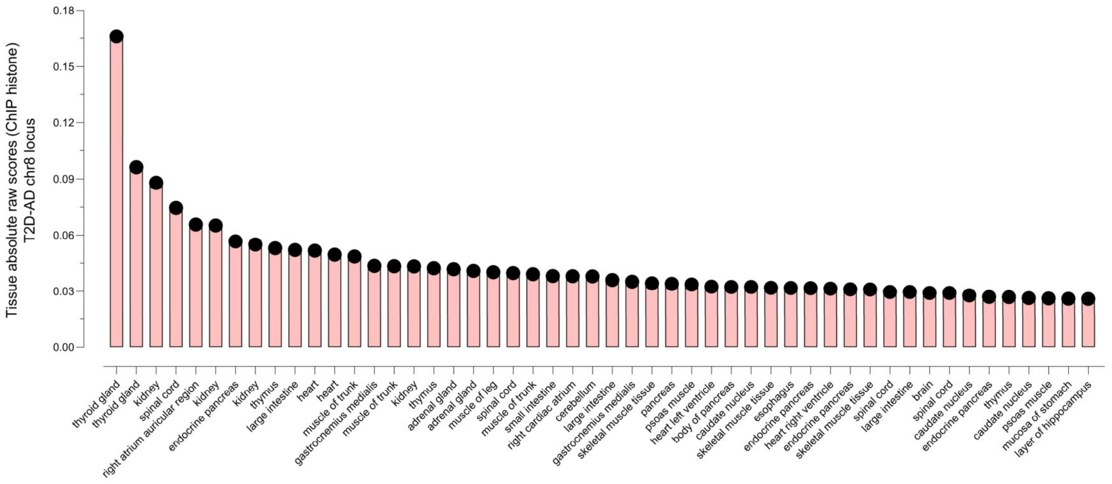


**Fig. S3**I2 | **AlphaGenome**. Top tissue predictions for the effects of rs13257021 in histone modifications.

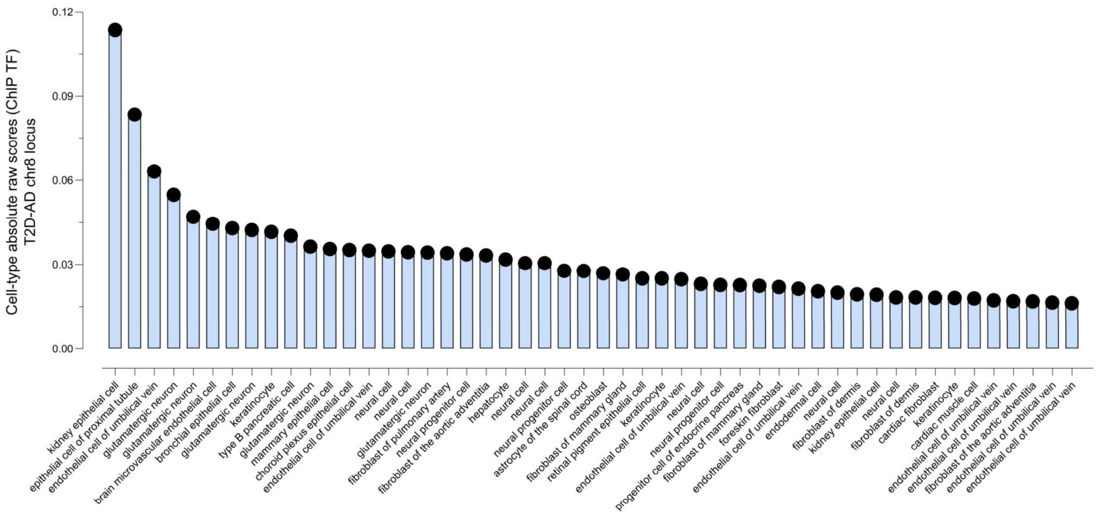


**Fig. S3**J2 | **AlphaGenome**. Top cell-type predictions for the effects of rs13257021 in transcription factor binding.

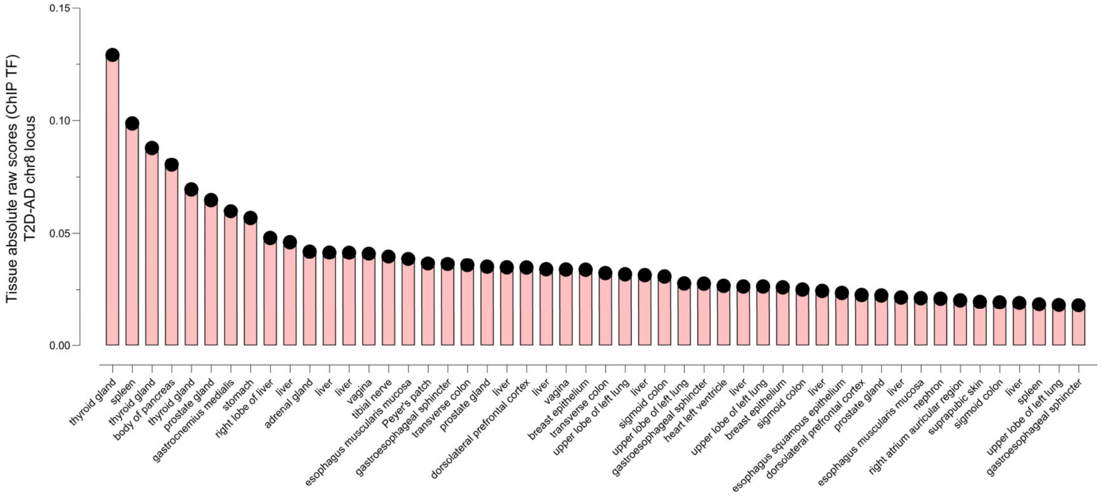


**Fig. S3**K2 | **AlphaGenome**. Top tissue predictions for the effects of rs13257021 in transcription factor binding.

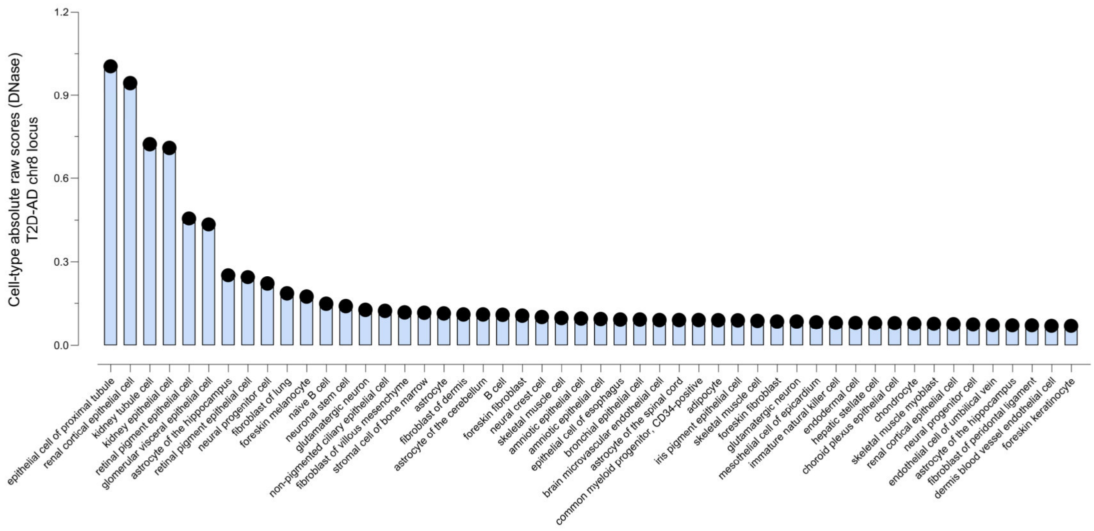


**Fig. S3**L2 | **AlphaGenome**. Top cell-type predictions for the effects of rs13257021 in chromatin accessibility assessed by DNase.

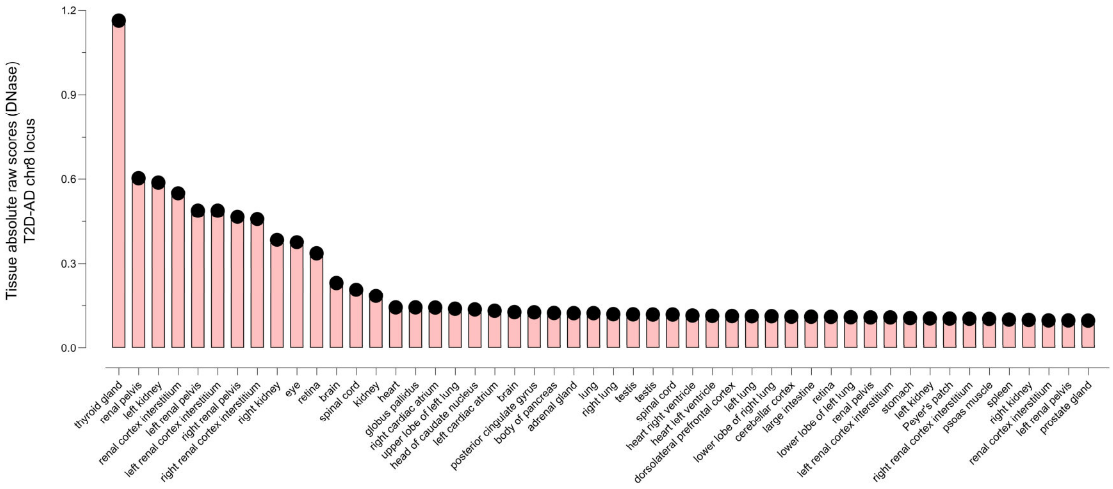


**Fig. S3**M2 | **AlphaGenome**. Top tissue predictions for the effects of rs13257021 in chromatin accessibility assessed by DNase.

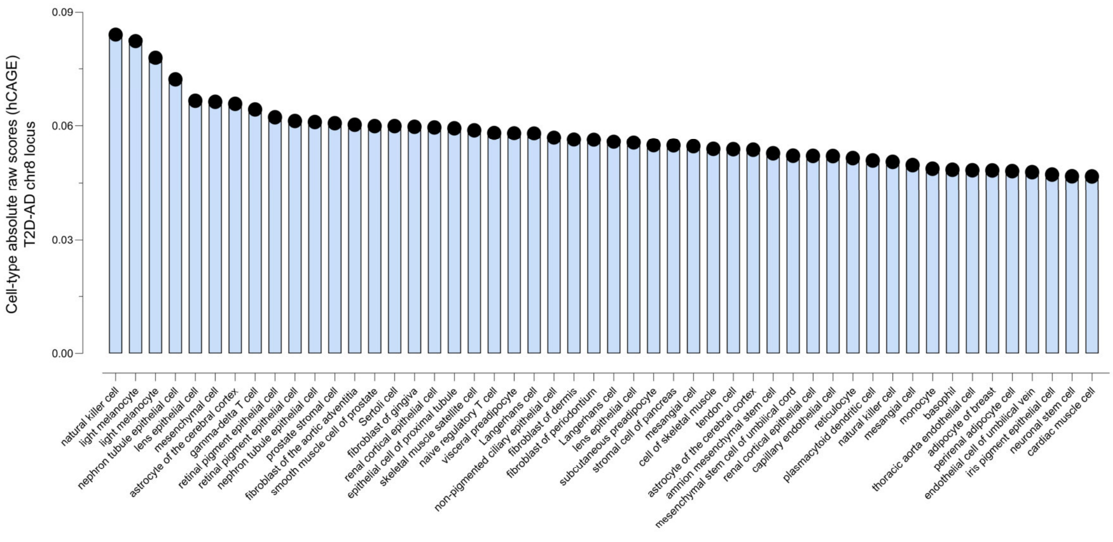


**Fig. S3**N2 | **AlphaGenome**. Top cell-type predictions for the effects of rs13257021 in gene expression assessed by hCAGE.

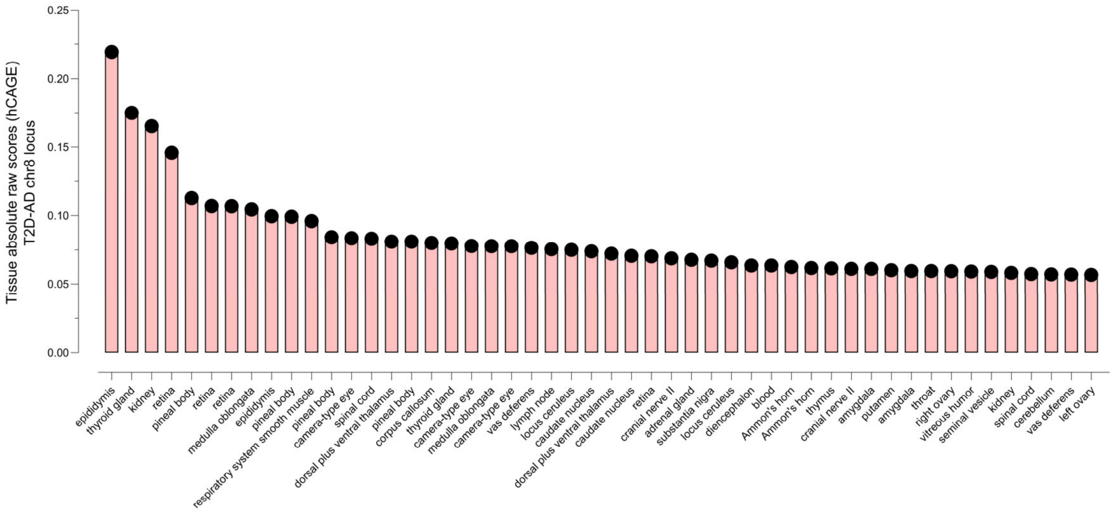


**Fig. S3**O2 | **AlphaGenome**. Top tissue predictions for the effects of rs13257021 in gene expression assessed by hCAGE.

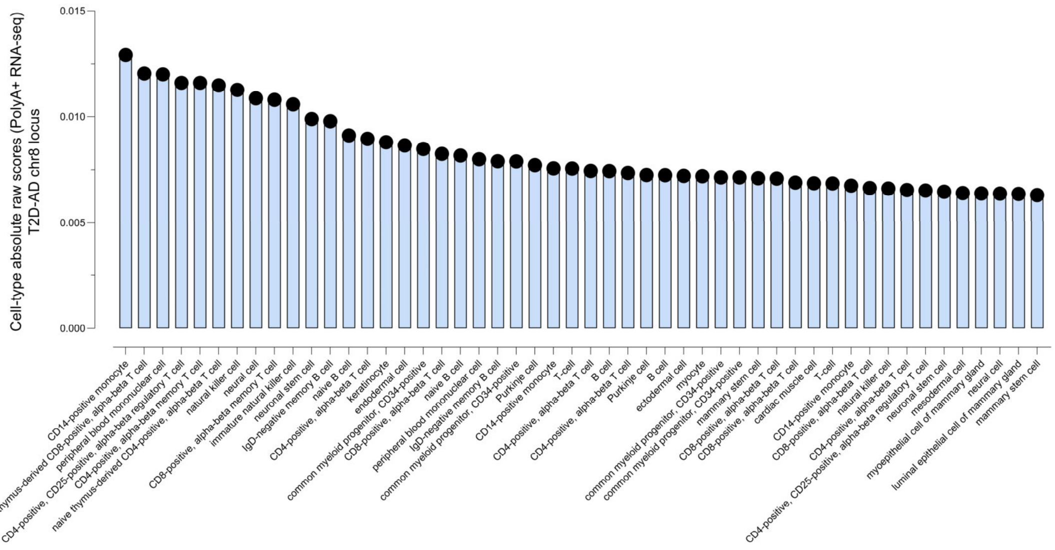


**Fig. S3**P2 | **AlphaGenome**. Top cell-type predictions for the effects of rs13257021 in gene expression assessed by polyA+ RNA-seq.

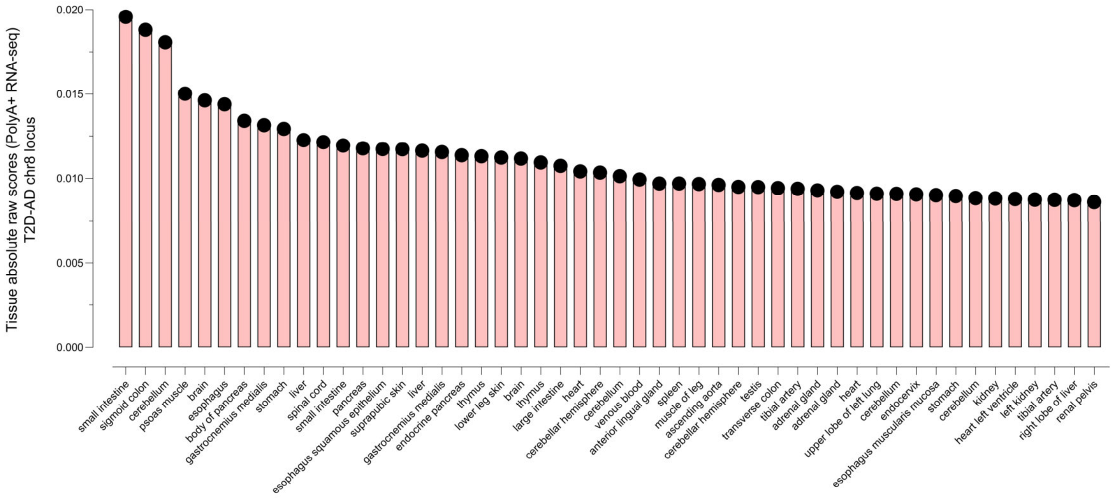


**Fig. S3**Q2 | **AlphaGenome**. Top tissue predictions for the effects of rs13257021 in gene expression assessed by polyA+ RNA-seq.

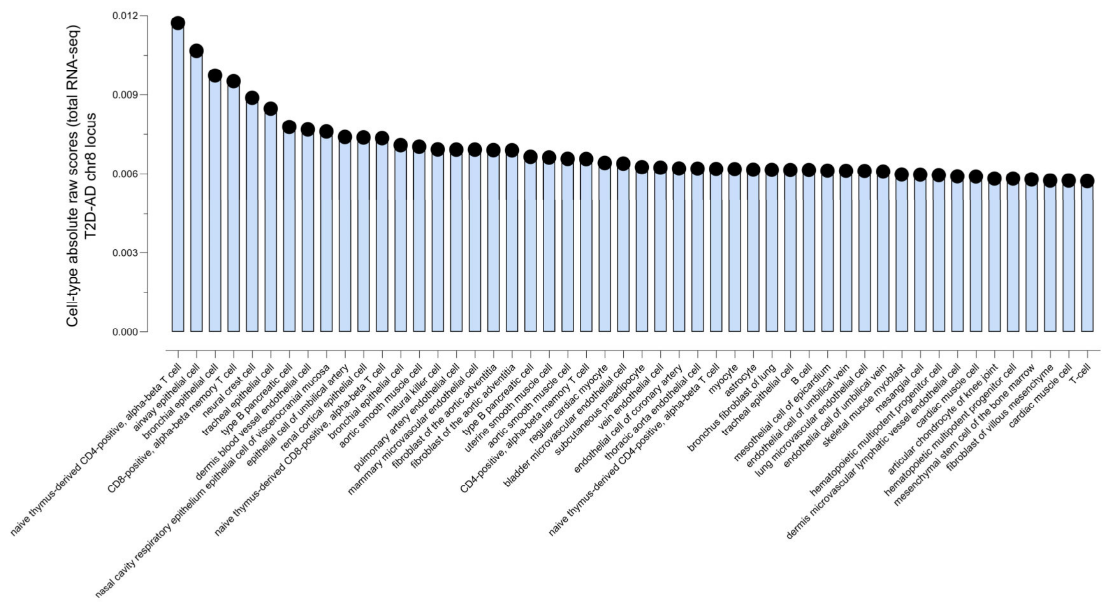


**Fig. S3**R2 | **AlphaGenome**. Top cell-type predictions for the effects of rs13257021 in gene expression assessed by total RNA-seq.

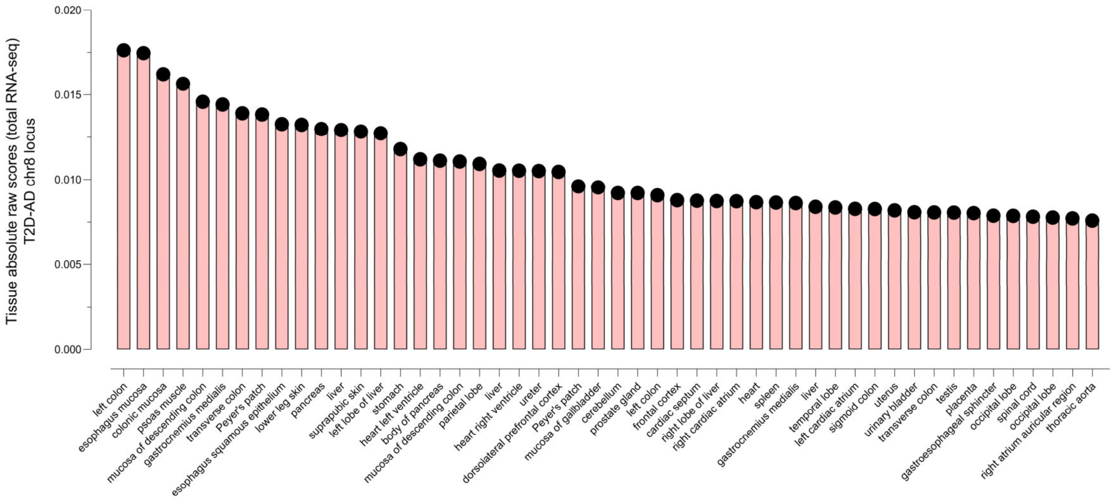


**Fig. S3**S2 | **AlphaGenome**. Top tissue predictions for the effects of rs13257021 in gene expression assessed by total RNA-seq.

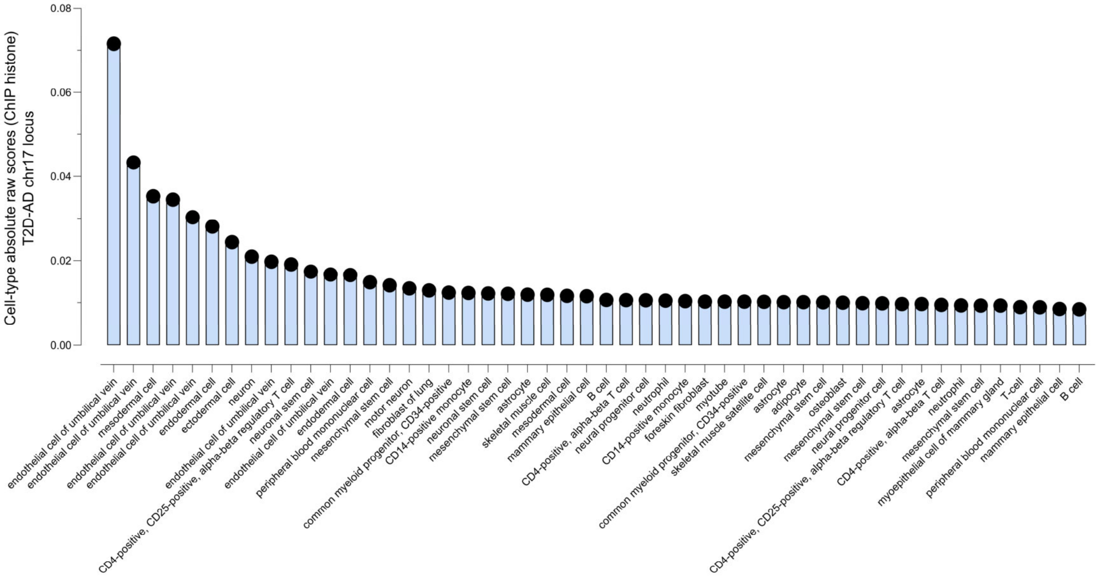


**Fig. S3**T2 | **AlphaGenome**. Top cell-type predictions for the effects of rs4311 in histone modifications.

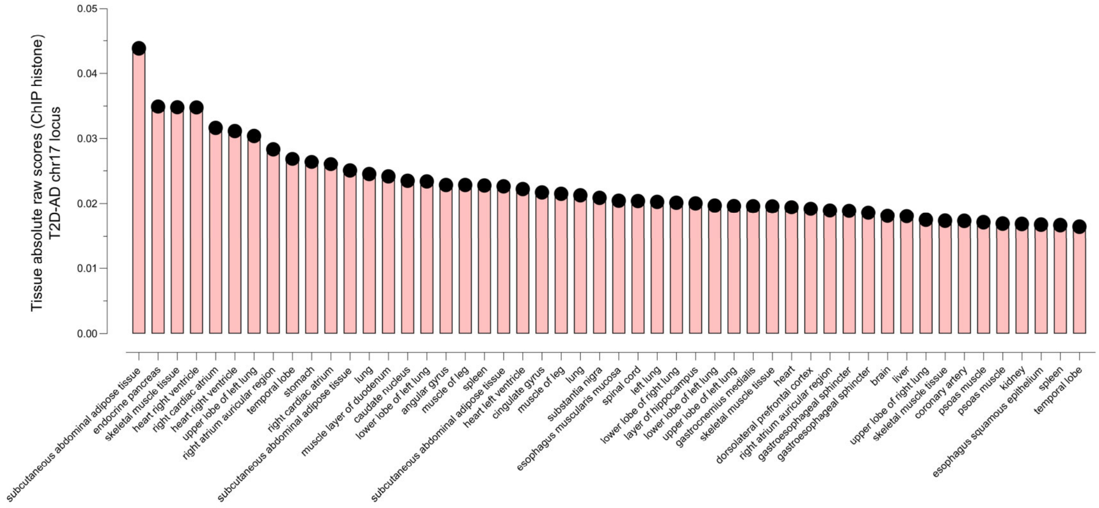


**Fig. S3**U2 | **AlphaGenome**. Top tissue predictions for the effects of rs4311 in histone modifications.

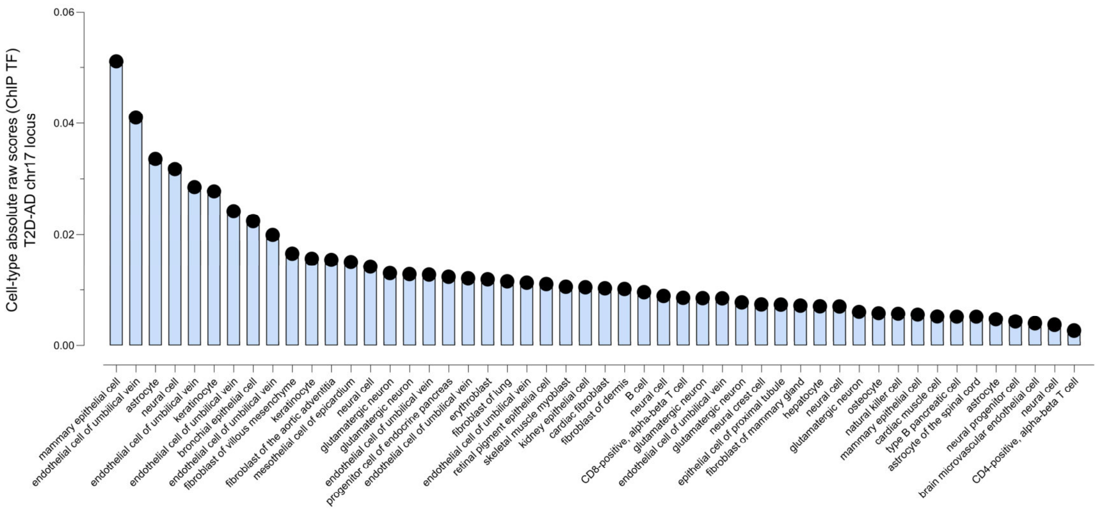


**Fig. S3**V2 | **AlphaGenome**. Top cell-type predictions for the effects of rs4311 in transcription factor binding.

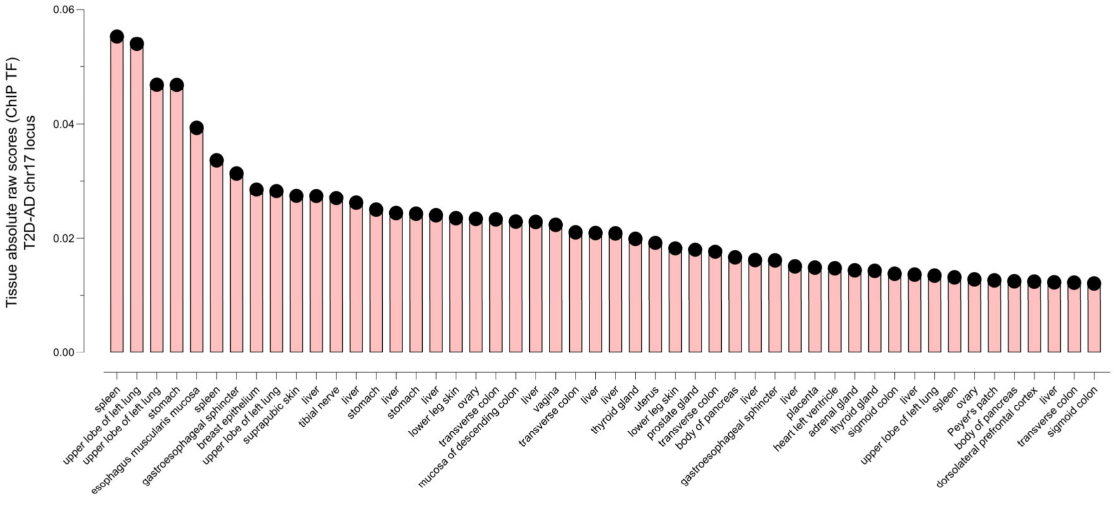


**Fig. S3**W2 | **AlphaGenome**. Top tissue predictions for the effects of rs4311 in transcription factor binding.

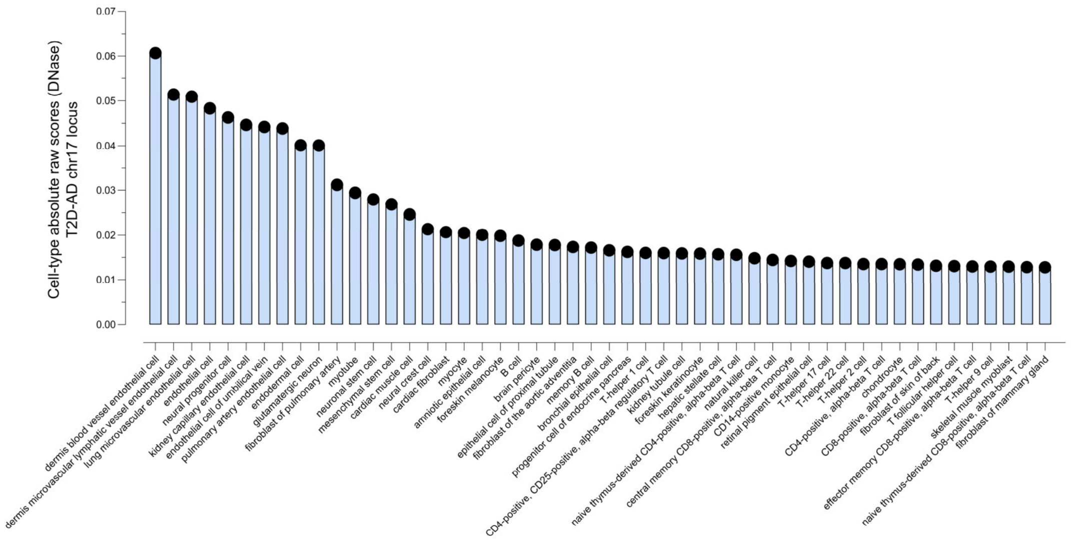


**Fig. S3**X2 | **AlphaGenome**. Top cell-type predictions for the effects of rs4311 in chromatin accessibility assessed by DNase.

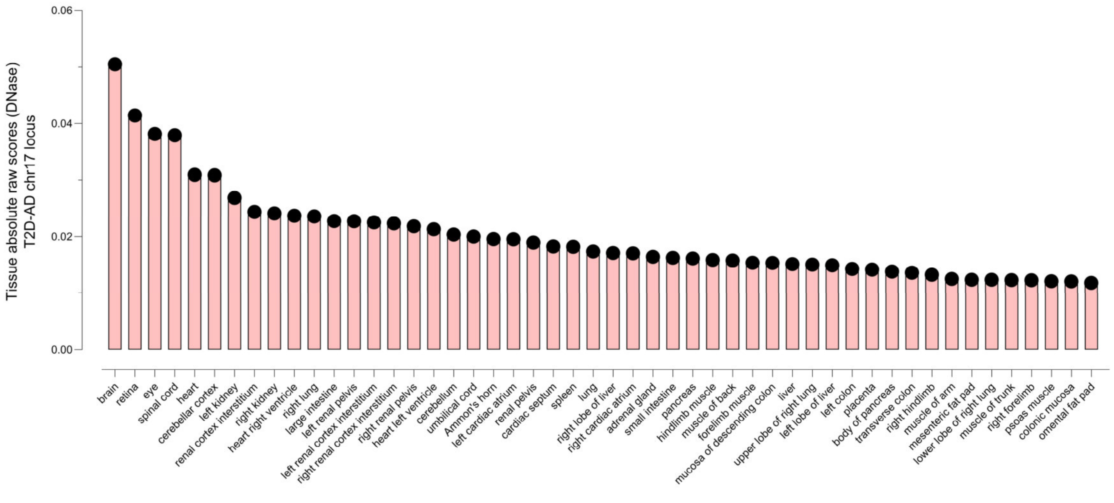


**Fig. S3**Y2 | **AlphaGenome**. Top tissue predictions for the effects of rs4311 in chromatin accessibility assessed by DNase.

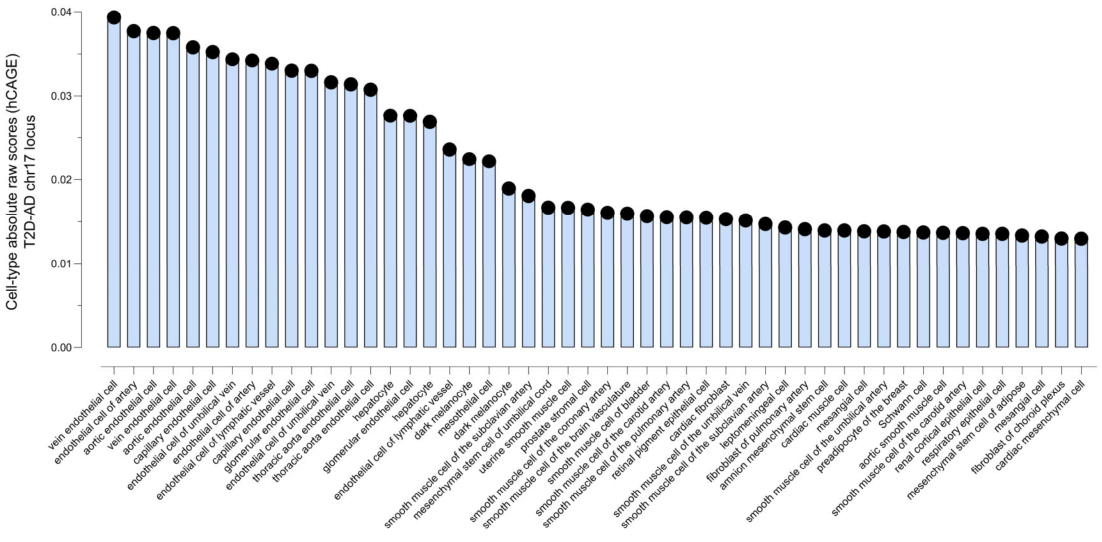


**Fig. S3**Z2 | **AlphaGenome**. Top cell-type predictions for the effects of rs4311 in gene expression assessed by hCAGE.

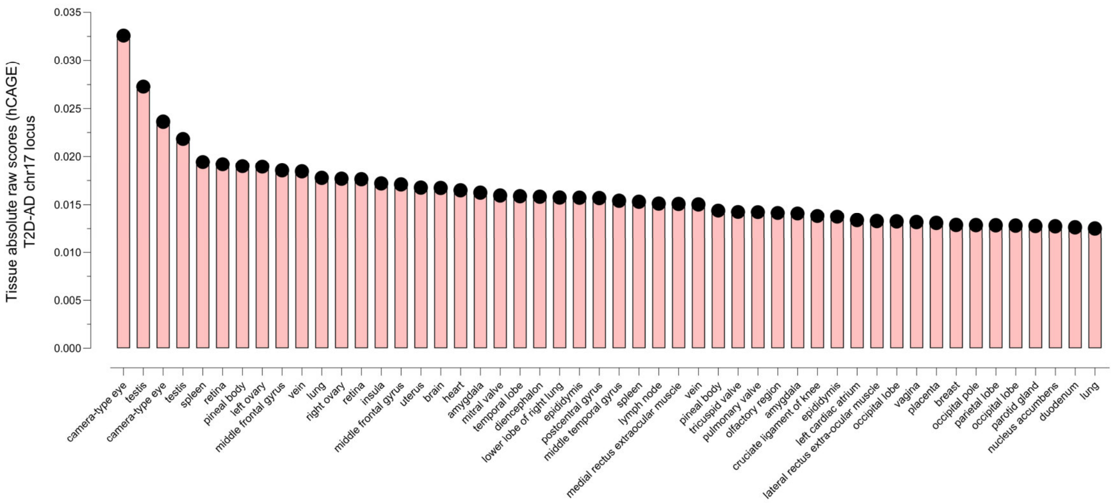


**Fig. S3**A3 | **AlphaGenome**. Top tissue predictions for the effects of rs4311 in gene expression assessed by hCAGE.

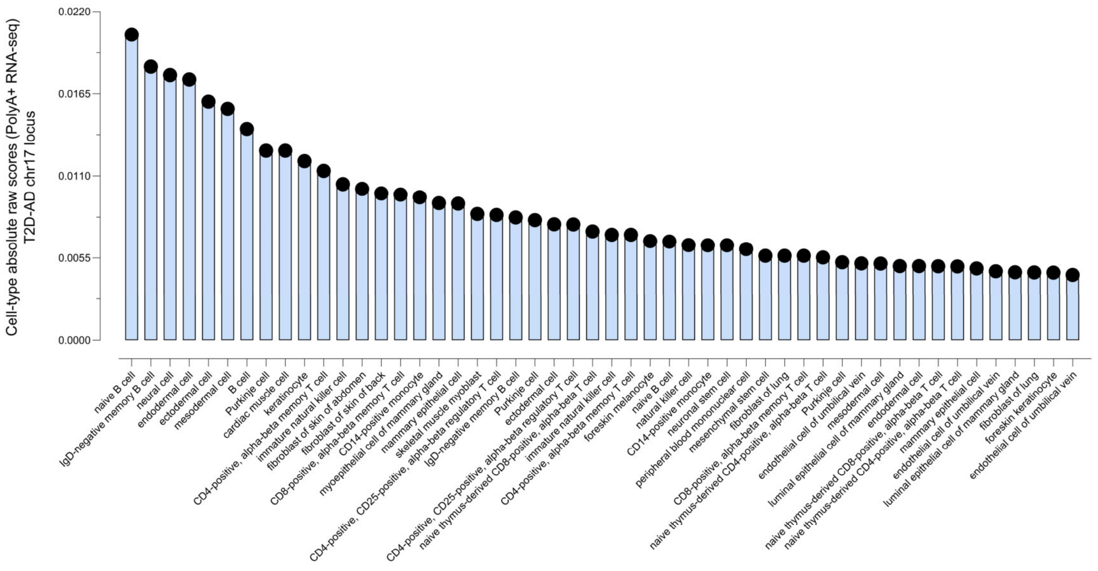


**Fig. S3**B3 | **AlphaGenome**. Top cell-type predictions for the effects of rs4311 in gene expression assessed by polyA+ RNA-seq.

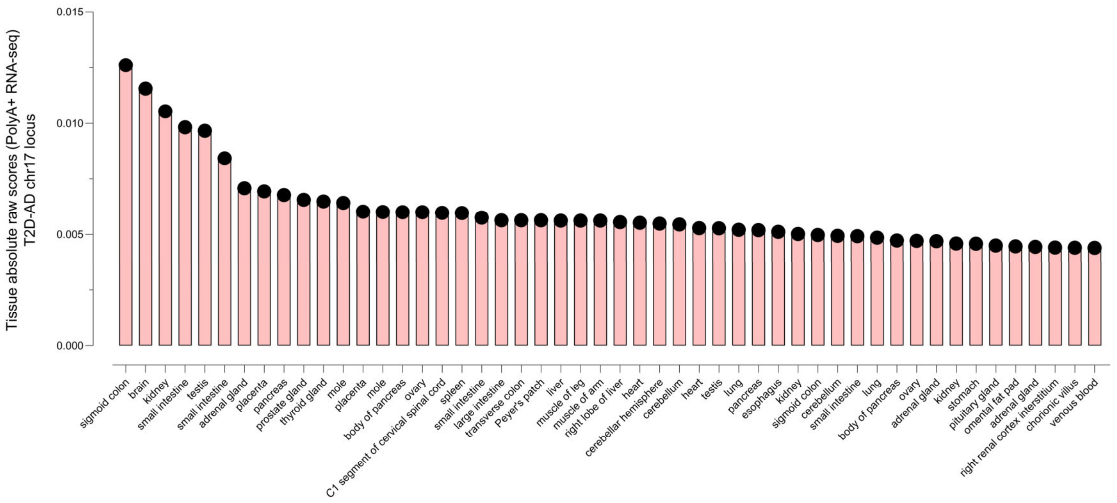


**Fig. S3**C3 | **AlphaGenome**. Top tissue predictions for the effects of rs4311 in gene expression assessed by polyA+ RNA-seq.

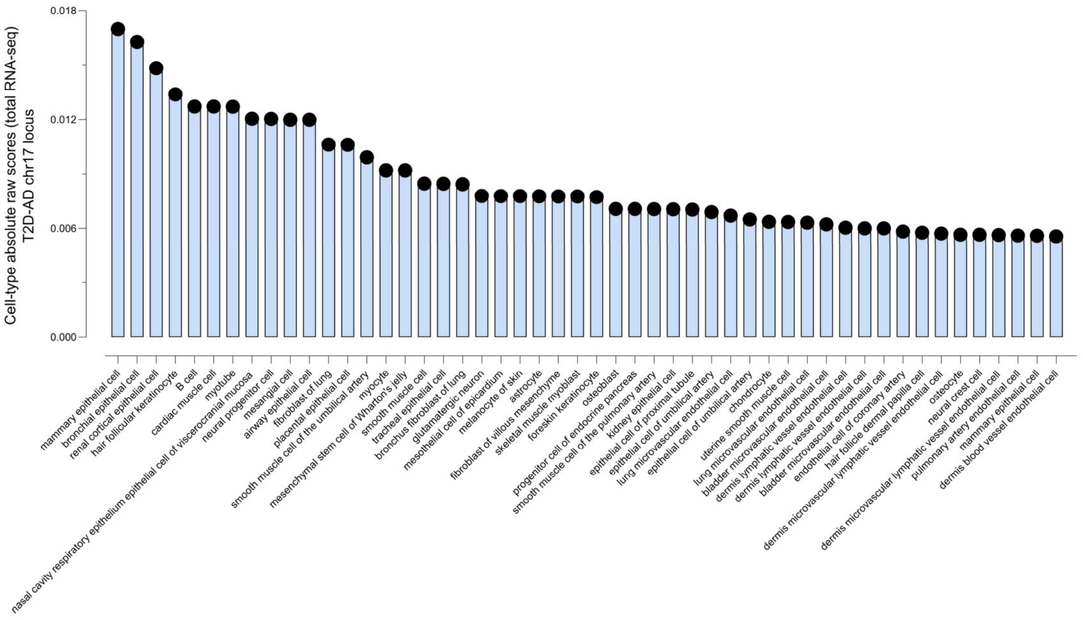


**Fig. S3**D3 | **AlphaGenome**. Top cell-type predictions for the effects of rs4311 in gene expression assessed by total RNA-seq.

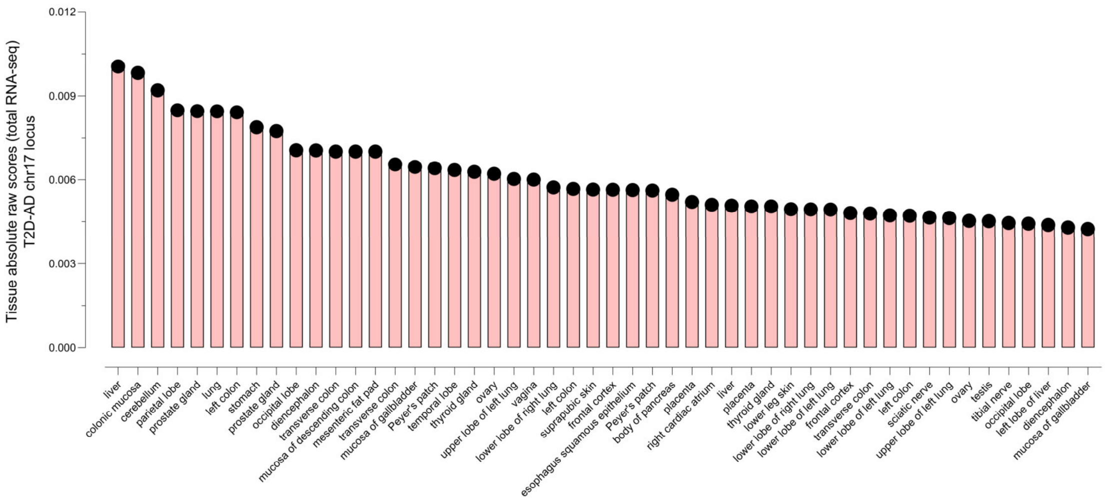


**Fig. S3**E3 | **AlphaGenome**. Top tissue predictions for the effects of rs4311 in gene expression assessed by total RNA-seq.

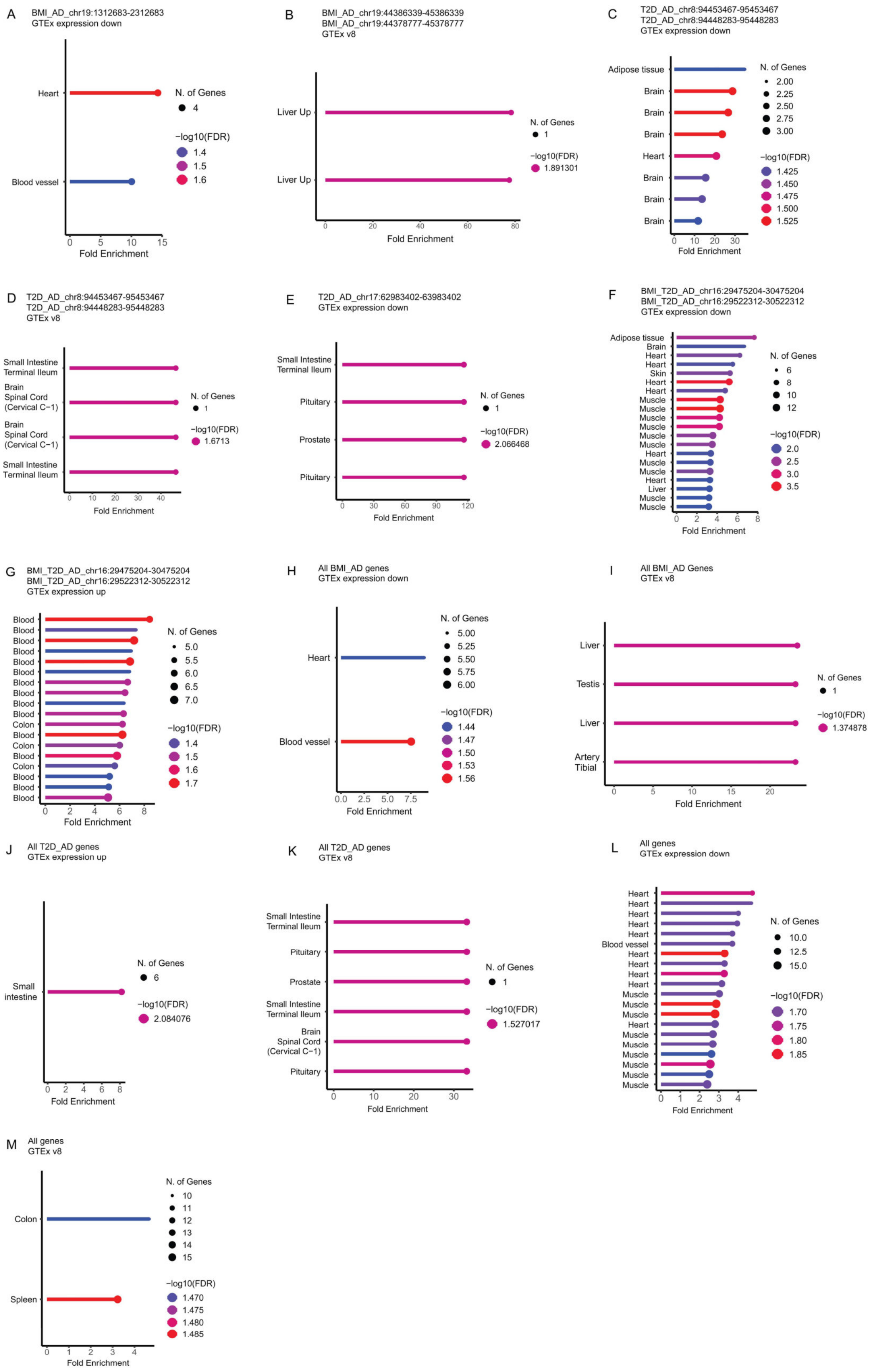


**Fig. S4** | GTEx enrichment analyses.

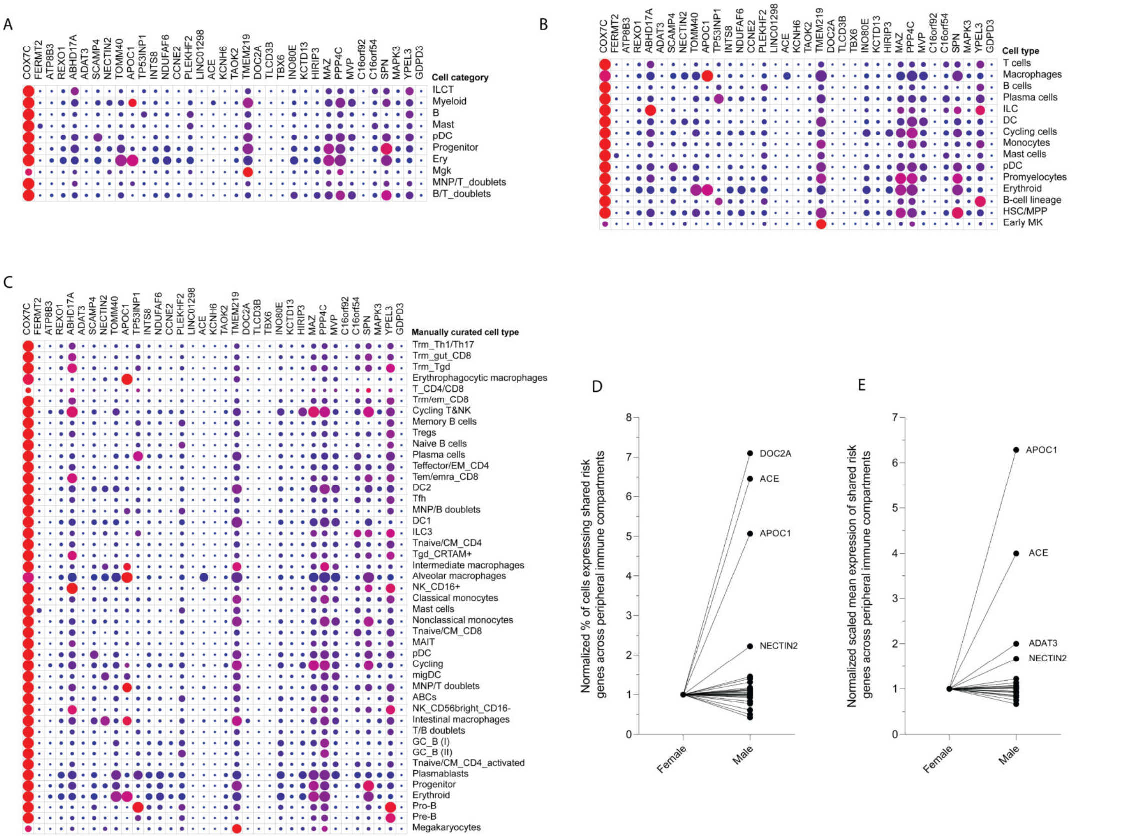


**Fig. S5** | (**A-C**) Individual gene expression and % of expressing cell-types in the immune niches of 17 peripheral tissues. (**D-E**) Sex-stratified individual gene expression and % of expressing cells. Results are normalized to females.

(A) **rs11668832** - chr19:1712682-1912682

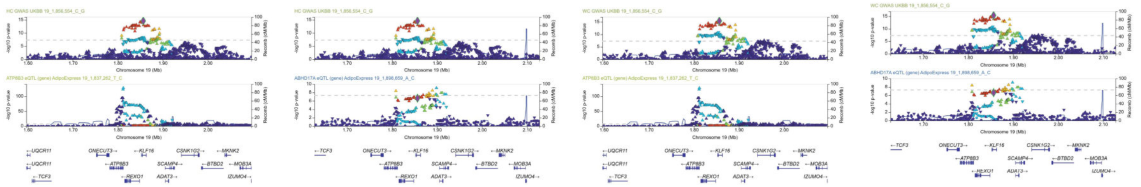

(B) **rs7254892** - chr19:45289596-45489596 / **rs6859** - chr19:45282034-45482034

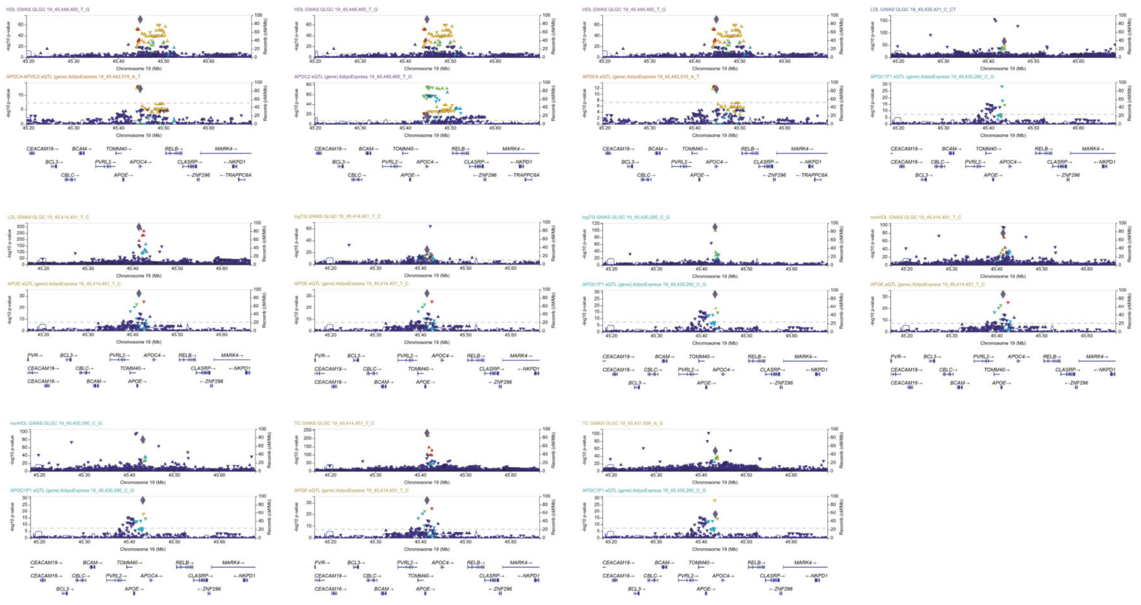

(C) **rs13257021** - chr8:95865695-96065695 / **rs896854** - chr8:95860511-96060511

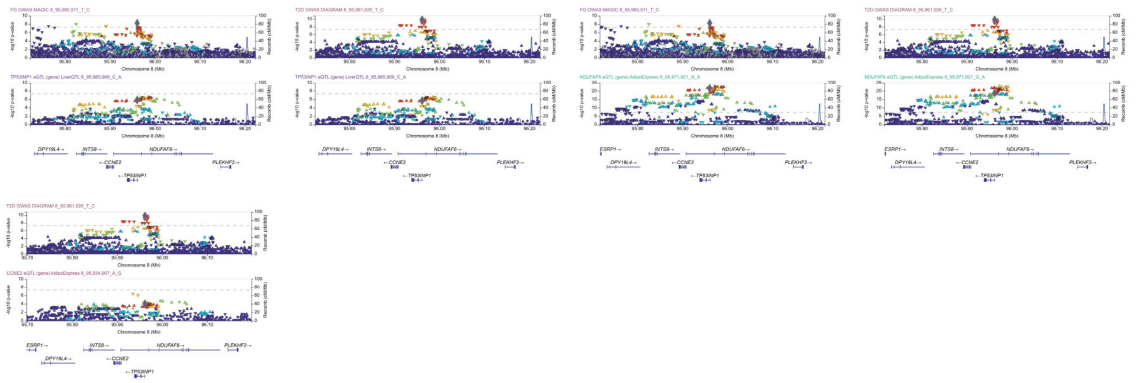

(D) **rs4311** - chr17:61460763-61660763

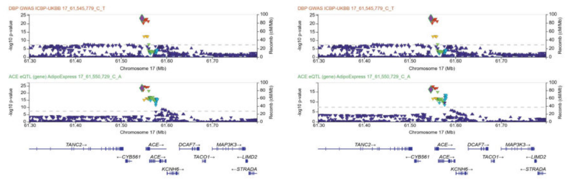

(E) **rs11150577** - chr16:29933633-30133633 / **rs12325539** - chr16:29886525-30086525

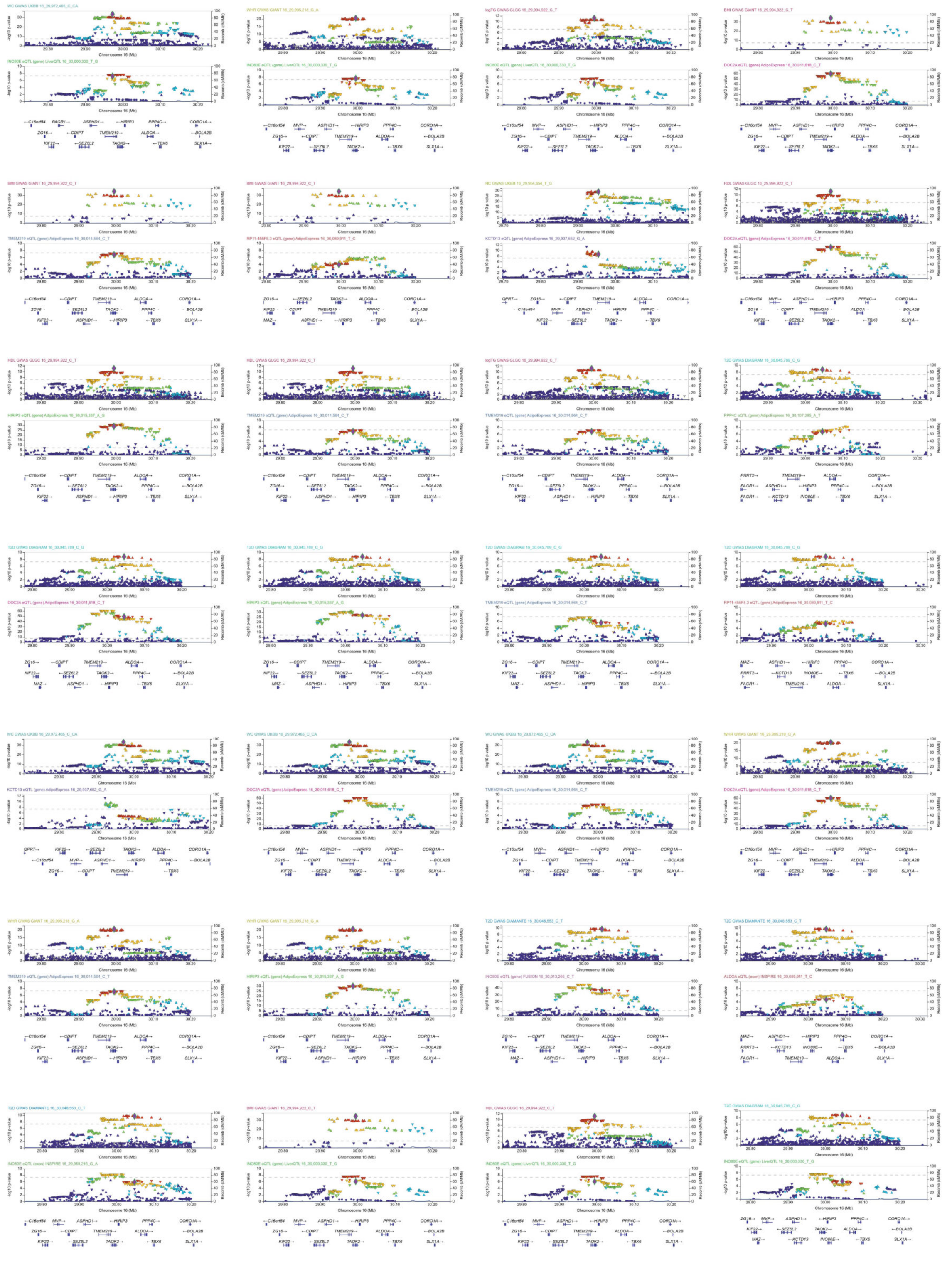

**Fig. S6 | GWAS-eQTL colocalization locuszoom region plots**.

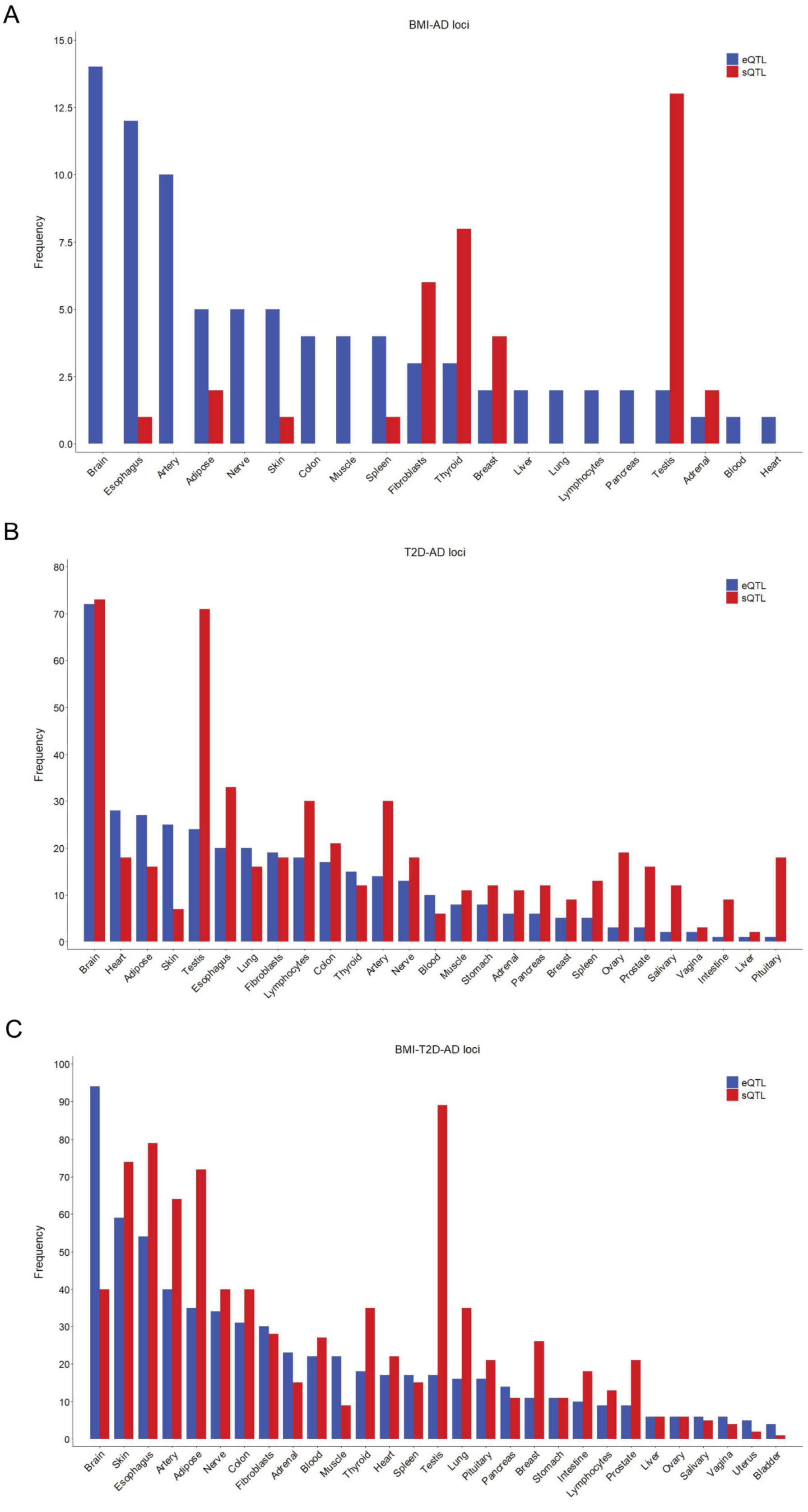


**Fig. S7 | eQTL and sQTL frequency**.

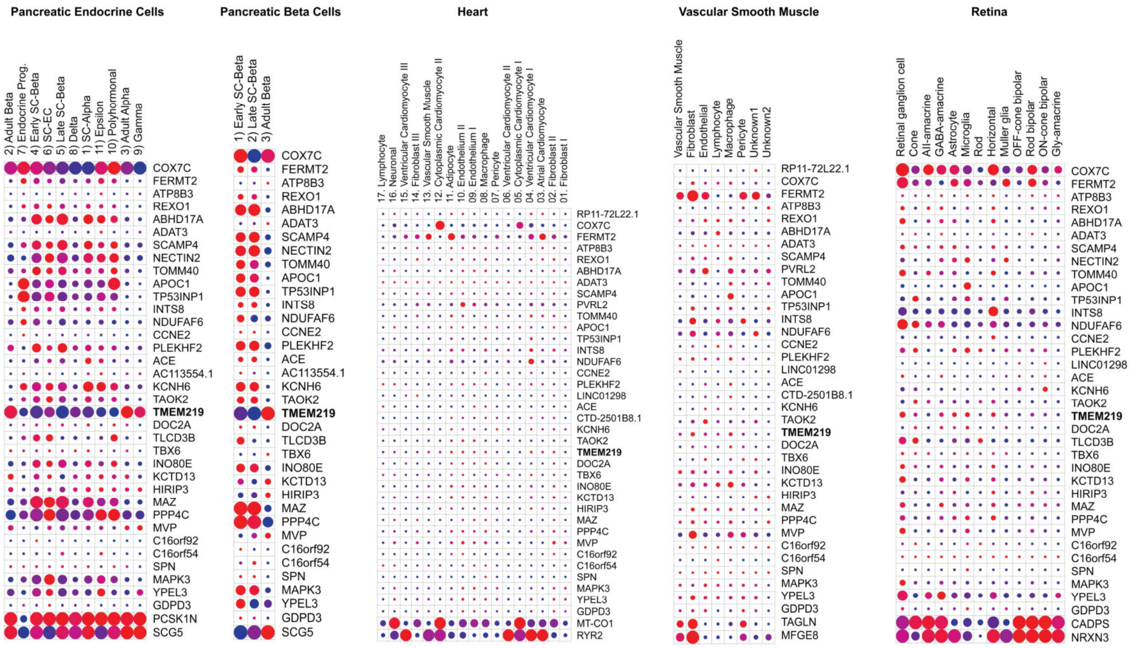


**Fig. S8 | Single-cell transcriptomics analysis of pancreatic endocrine cells, pancreatic beta cells, heart, vascular smooth muscle and retina.**

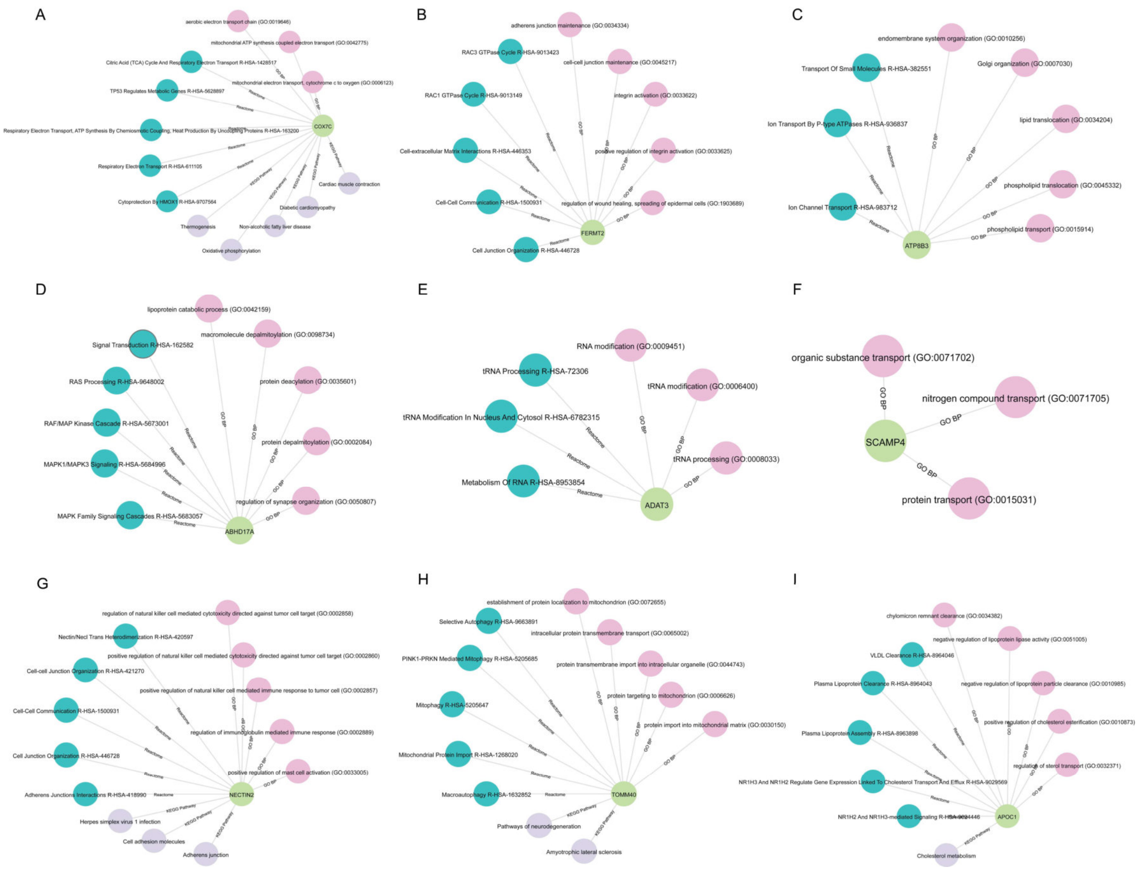


**Fig. S9**A | **Enrichment results using GO terms, KEGG and Reactome pathways for BMI-AD effector genes.**

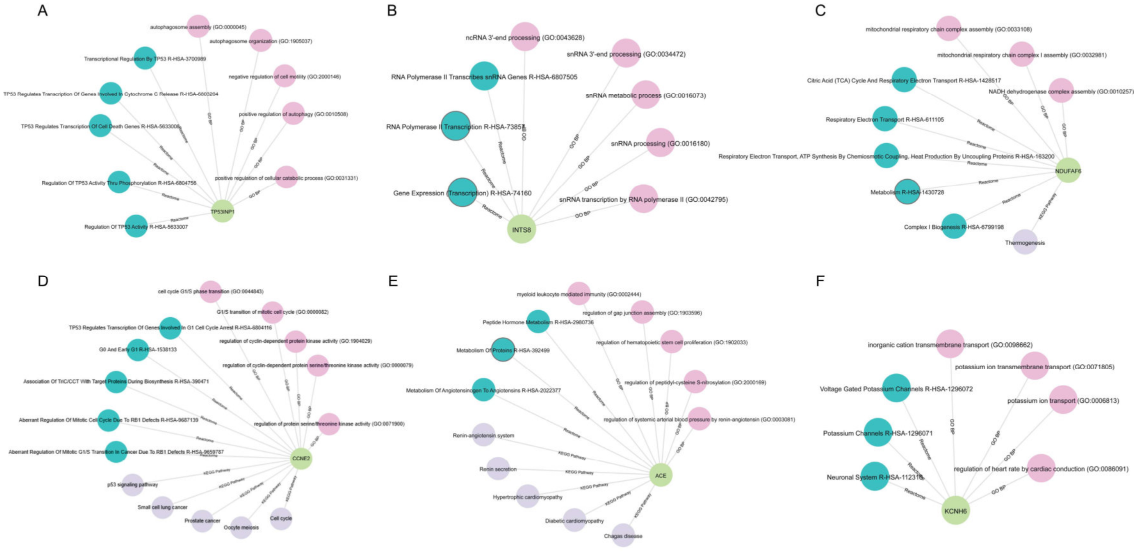


**Fig. S9**B | **Enrichment results using GO terms, KEGG and Reactome pathways for T2D-AD effector genes.**

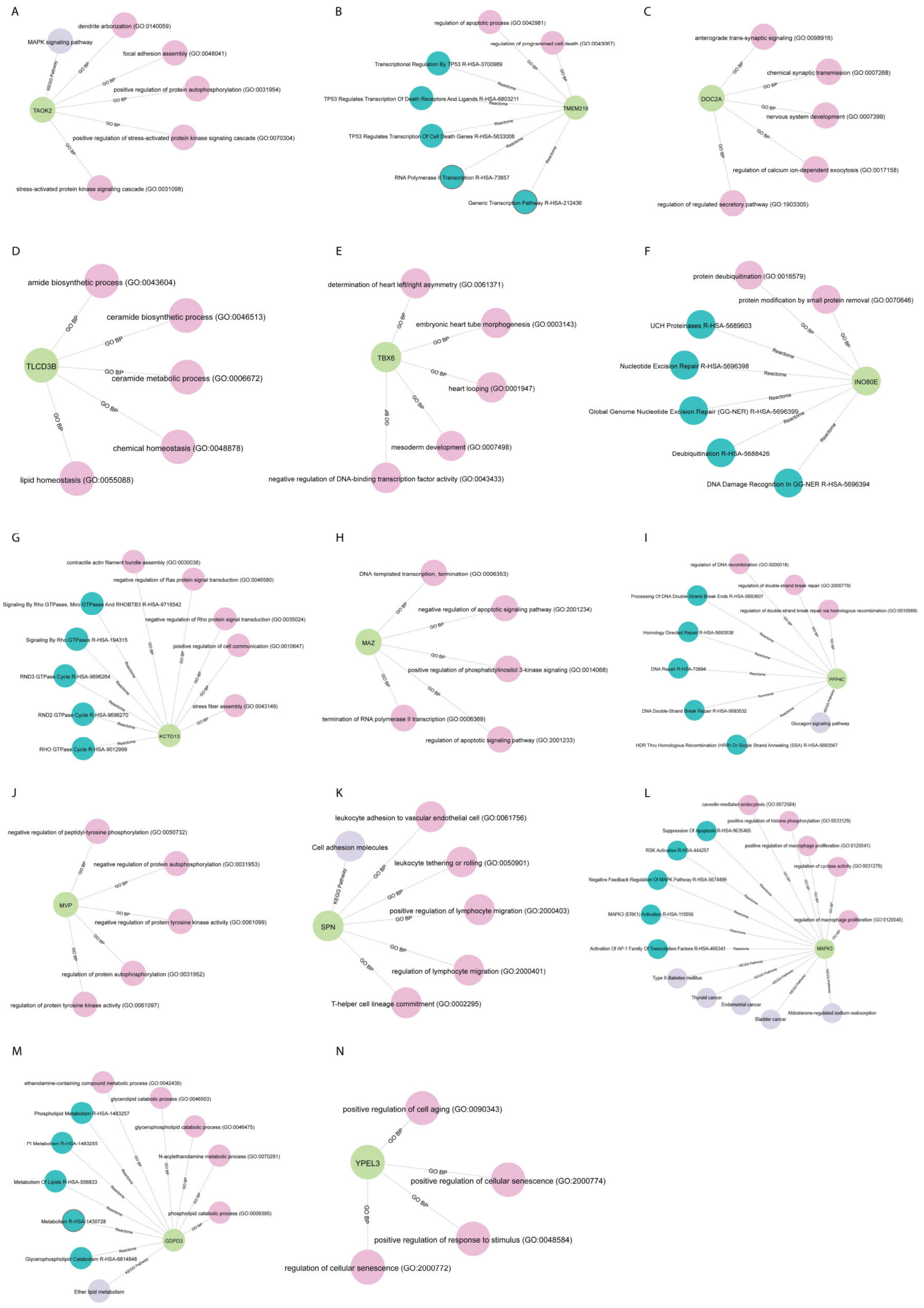


**Fig. S9C | Individual enrichment results using GO terms, KEGG and Reactome pathways for BMI-T2D-AD effector genes.**
