## Supplementary material for "Genomics link obesity and type 2 diabetes to Alzheimer’s disease to unveil novel biological insights": EADB Supplementary Information

### *GWAS cohort descriptions and acknowledgements*.

### 1. GWAS Cohort descriptions and acknowledgements

#### 1.1 European Alzheimer & Dementia Biobank (EADB-TOPMed)

**EADB-France.** In the France node, samples were collected from nine countries (39 centers/studies), and after quality controls (QCs), we obtained 13,867 AD cases and 15,310 controls. All these samples were genotyped at the Centre National de Recherche en Génomique Humaine (CNRGH, Evry, France).
Belgium: The participants were part of a large prospective cohort (PMID: 29589097) of Belgian AD patients and healthy elderly control individuals. The patients were ascertained at the memory clinic of Middelheim and Hoge Beuken (Hospital Network Antwerp, Belgium) and at the memory clinic of the University Hospitals of Leuven, Belgium. The control individuals were the partners of the patients or volunteers from the Belgian community. The study protocols were approved by the ethics committees of the Antwerp University Hospital and the participating neurological centers at the different hospitals of the BELNEU consortium and by the University of Antwerp.
Czech Republic: The Czech Brain Aging Study (CBAS) (PMID: 31857299) is a longitudinal memory-clinic–based study recruiting subjects at risk of dementia (subjects referred for cognitive complaints-SCD, MCI). The CBAS+ study is a cross-sectional study of patients in the early stages of dementia. All subjects signed informed consent and both studies were approved by the local ethics committee.
Denmark: The Copenhagen General Population Study (CGPS) is a prospective study of the Danish general population initiated in 2003 and still recruiting. Individuals were selected randomly based on the national Danish Civil Registration System to reflect the adult Danish population aged 20-100. Data were obtained from a self-administered questionnaire reviewed together with an investigator at the day of attendance, a physical examination, and from blood samples including DNA extraction (PMID: 33022702). The study is approved by the regional ethics committee and by the Steering Committee of the CGPS.
Finland: *The ADGEN cohort (PMID: 25807283):* clinic-based collection of AD patients from Eastern and Northern Finland examined in the Department of Neurology in Kuopio University Hospital and the Department of Neurology in Oulu University Hospital. All the patients were diagnosed with probable AD according to the criteria of the National Institute of Neurological and Communicative Disorders and Stroke and the Alzheimer's disease and Related Disorders Association (NINCDS–ADRDA). The study was approved by the ethics committee of Kuopio University Hospital, Finland (420/2016). *The FINGER study* (PMID: 25771249): a Finnish multi-domain lifestyle RCT enrolling 1,260 older adults with an increased risk of dementia from the general population. The intensive lifestyle intervention lasted for two years, and follow-up extends currently up to seven years. The FINGER study was approved by the coordinating ethics committee of the Hospital District of Helsinki and Uusimaa (94/13/03/00/2009 and HUS/1204/2017), and all the participants gave written informed consent.
France: *The BALTAZAR multicenter (23 memory centers) prospective study* (PMID: 29458036): 1,040 participants from September 2010 to April 2015. They were classified as AD cases (n = 501) according to DSM IV-TR and NINCDS–ADRDA criteria as well as amnestic mild cognitive impairment (MCI) cases (a MCI, n = 417) and non-amnestic MCI cases (na MCI, n = 122) according to Petersen's criteria. A comprehensive battery of cognitive tests was performed, including MMSE, verbal fluency, and FCSRT. All the participants or their legal guardians gave written informed consent. The study was approved by the Paris ethics committee (CPP Ile de France IV Saint Louis Hospital). MEMENTO: a clinic-based study (PMID: 28851447) aimed at better understanding the natural history of AD, dementia, and related diseases. Between 2011 and 2014, 2,323 individuals presenting either recently diagnosed MCI or isolated cognitive complaints were enrolled in 26 memory centers in France. This study was performed in accordance with the guidelines of the Declaration of Helsinki. The MEMENTO study protocol has been approved by the local ethics committee (Comité de Protection des Personnes Sud-Ouest et Outre Mer III; approval number 2010-A01394-35). All the participants provided written informed consent. The CNRMAJ-Rouen study (PMID: 26242991): early onset AD patients (n = 870). The patients or their legal guardians provided written informed consent. This study was approved by the ethics committee of CPP Ile de France II.
Italy: The AD cases and controls were collated through Italy in different centers: Brescia, Cagliari, Florence, Milan, Rome, Pertugia, San Giovani Rotondo and Torino. AD cases were diagnosed according to DSM III-R,IV and NINCDS–ADRDA criteria. Controls were defined a minima as subjects without DMS-III-R dementia criteria and with integrity of their cognitive functions (MMS>25).
Spain: *The Dementia Genetic Spanish Consortium (DEGESCO)* is a national consortium comprising 23 research centers and hospitals across the country, that holds the institutional coverage of The Network Center for Biomedical Research in Neurodegenerative Diseases (CIBERNED). Created in 2013, DEGESCO's

objective is the promotion and conduction of genetic studies aimed at understanding the genetic architecture of neurodegenerative dementias in the Spanish population and participates in coordinated actions in national and international frameworks. All DNA samples are in compliance with the Law of Biomedical Research (Law 14/2007) and the Royal Decree on Biobanks (RD 1716/2011). Patients included in the present study met clinical criteria for probable or possible disease established by the National Institute of Neurological and Communication Disorders and Stroke and the Alzheimer Disease and Related Disorders Association (NINCDS-ADRDA). Cognitively healthy controls were unrelated individuals who had a documented MMSE in the normal range. Contributing centers in the France node genotyping were Centro de Biología Molecular Severo Ochoa (CSIC-UAM (Madrid), the Institute Biodonostia, University of Basque Contry (EHU-UPV, San Sebastián), Institut de Biomedicina de Valencia CSIC (València), and Sant Pau Biomedical Research Institute (Barcelona).

Sweden: *Uppsala.* The Swedish AD patients were ascertained at the Memory Disorder Unit at Uppsala University Hospital. For all patients, the diagnosis was established according to the National Institute on Neurological Disorders and Stroke, and the Alzheimer's Disease and Related Disorders Association (NINDS-ADRDA) guidelines (PMID: 6610841). Healthy control subjects were recruited from the same geographic region following advertisements in local newspapers and displayed no signs of dementia upon Mini Mental State Examination (MMSE). *Karolinska Institutet, Stockholm*. AD patients were ascertained at the Memory Clinic at Karolinska University Hospital - Huddinge, Stockholm Sweden. For all patients, the diagnosis was established according to the National Institute on Neurological Disorders and Stroke, and the Alzheimer's Disease and Related Disorders Association (NINDS-ADRDA) guidelines (PMID: 6610841) and if available supported by CSF beta-amyloid and phospho-tau biomarker analyses. As controls, DNA from participants in the *Swedish National Study on Aging and Care in Kungsholmen* (SNAC-K) data was collected. The original SNAC-K population consisted of 4590 living and eligible persons who lived on the island of Kungsholmen in Central Stockholm, belonged to pre-specified age strata, and were randomly selected to take part in the study. Between 2001 and 2004, 3363 persons participated in the baseline assessment. They belonged to the age cohorts 60, 66, 72, 78, 81, 84, 87, 90, 93, and 96 years and 99 years and older. The examination consists of three parts: a nurse interview, a medical examination, and a neuropsychological testing session. Altogether, the examination takes about six hours. The participants are reexamined each time they reach the next age cohort. All parts of the SNAC-K project have been approved by the ethical committee at Karolinska Institutet or the regional ethical review board. Informed consent was collected from all the participants or, if the person was severely cognitively impaired, from their next of kin.

The UK: MRC. The sample set comprises individuals with AD and healthy controls recruited across the MRC Centre for Neuropsychiatric Genetics and Genomics, Cardiff University, Cardiff, UK; Institute of Psychiatry, London, UK; University of Cambridge, Cambridge, UK. The collection of the samples was through multiple channels, including specialist NHS services and clinics, research registers and Join Dementia Research (JDR) platform. The participants were assessed at home or in research clinics along with an informant, usually a spouse, family member or close friend, who provided information about and on behalf of the individual with dementia. Established measures were used to ascertain the disease severity: Bristol activities of daily living (BADL), Clinical Dementia Rating scale (CDR), Neuropsychiatric Inventory (NPI) and Global Deterioration Scale (GIDS). Individuals with dementia completed the Addenbrooke's Cognitive Examination (ACE-r), Geriatric Depression Scale (GeDS) and National Adult Reading Test (NART) too. Control participants were recruited from GP surgeries and by means of self-referral (including existing studies and Joint Dementia Research platform). For all other recruitment, all AD cases met criteria for either probable (NINCDS-ADRDA, DSM-IV) or definite (CERAD) AD. All elderly controls were screened for dementia using the Mini Mental State Examination (MMSE) or ADAS-cog, were determined to be free from dementia at neuropathological examination or had a Braak score of 2.5 or lower. Control samples were chosen to match case samples for age, gender, ethnicity and country of origin. Informed consent was obtained for all study participants, and the relevant independent ethical committees approved study protocols. SOTON, University of Southampton, Southampton, UK. All AD cases met criteria for either probable (NINCDS-ADRDA, DSM-IV) or definite (CERAD) AD. All elderly controls were screened for dementia using the MMSE or ADAS-cog, were determined to be free from dementia at neuropathological examination or had a Braak score of 2.5 or lower. Nottingham and Manchester, University of Nottingham, Nottingham, UK and Manchester Brain Bank. All AD cases met criteria for either probable (NINCDS-ADRDA, DSM-IV) or definite (CERAD) AD. All elderly controls were screened for dementia using the MMSE or ADAS-cog, were determined to be free from dementia at neuropathological examination or had a Braak score of 2.5 or lower. KCL, London Neurodegenerative Diseases Brain Bank. All AD cases met criteria for either probable (NINCDS-ADRDA, DSM-IV) or definite (CERAD) AD. All elderly controls were screened for dementia using the MMSE or ADAS-cog, were determined to be free from dementia at neuropathological examination or had a Braak score of 2.5 or lower. PRION, All

AD cases met criteria for either probable (NINCDS-ADRDA, DSM-IV) or definite (CERAD) AD. All elderly controls were screened for dementia using the MMSE or ADAS-cog, were determined to be free from dementia at neuropathological examination or had a Braak score of 2.5 or lower. CFAS Wales, The Cognitive Function and Ageing Study Wales (CFAS-Wales) is a longitudinal population-based study of people aged 65 years and over in rural and urban areas of Wales that aims to investigate physical and cognitive health in older age and examine the interactions between health, social networks, activity, and participation. Individuals aged 65 years and over were randomly sampled from general medical practice lists between 2011 and 2013, stratified by age to ensure equal numbers in two age groups, 65-74 years and 75 and over. The baseline sample included 3593 older people and included those living in care homes as well as those living at home. Those who provided written consent to join the study were interviewed in their own homes by trained interviewers and could choose to have the interview conducted through the medium of either English or Welsh. Participants were followed up 2 years later. All AD cases met criteria for either probable (NINCDS-ADRDA, DSM-IV) or definite (CERAD) AD. All elderly controls were screened for dementia using the MMSE or CAMCOG, and were determined to be free from dementia. UCL-DRC. the UCL Alzheimer’s disease cohort of the Dementia Research Centre (UCL - EOAD DRC) included patients seen at the Cognitive Disorders Clinics at The National Hospital for Neurology and Neurosurgery (Queen Square), or affiliated hospitals. Individuals were assessed clinically and diagnosed as having probable Alzheimer’s disease based on contemporary clinical criteria in use at the time, including imaging and neuropsychological testing where appropriate.

**EADB-Germany.** In the German node, samples were collected from seven countries (11 centers/studies) and after QCs, we obtained 4,159 AD cases and 4,545 controls. All these samples were genotyped at Life&brain (Bonn, Germany).

<u>Germany</u>: *DELCODE (the multicenter DZNE-Longitudinal Cognitive Impairment and Dementia Study)*. This is an observational longitudinal memory clinic-based multicenter study in Germany comprising 400 subjects with Subjective cognitive decline (SCD), 200 mild cognitive impairment (MCI) patients, 100 AD dementia patients, 200 control subjects without subjective or objective cognitive decline, and 100 first-degree relatives of patients with a documented diagnosis of AD dementia. All patient groups (SCD, MCI, AD) are referrals, including self-referrals, to the participating memory centers. The control group and the relatives of AD dementia patients are recruited by standardized public advertisement. Ten university-based memory centers are participating, all being collaborators of local DZNE sites. All patient groups (SCD, MCI, AD) were assessed clinically at the respective memory centers before entering DELCODE. The assessments include medical history, psychiatric and neurological examination, neuropsychological testing, blood laboratory work-up, cerebrospinal fluid (CSF) biomarkers, and routine MRI, all according to the local standards. The Consortium to Establish a Registry for Alzheimer’s Disease (CERAD) neuropsychological test battery was applied at all memory centers to measure cognitive function. German age, sex, and education-adjusted norms of the CERAD neuropsychological battery are available online (www.memoryclinic.ch). Detail description is of recruitment protocol is reported elsewhere. *The VOGEL study*: The VOGEL study is a prospective, observational, long-term follow-up study with three time points of investigation within 6–8 years. This cohort includes dementia and healthy subjects. Residents of the city of Würzburg born between 1936 and 1941 were recruited. Every participant underwent physical, psychiatric, and laboratory examinations and performed intense neuropsychological testing as well as VSEP and NIRS according to the published procedures. A total of 604 subjects were included. *The Heidelberg/Mannheim memory clinic sample*: This cohort includes 61 subjects from whom 40 MCI patients were recruited and assessed between 2012 and 2016. Some of those patients converted to dementia by AD or other dementias. *The PAGES study*: This study includes 301 subjects. AD patients were recruited at the memory clinic of the Department of Psychiatry, University of Munich, Germany. Participants in whom dementia associated with AD was diagnosed fulfilled the criteria for probable AD according to the NINCDS–ADRDA. The control group included participants who were randomly selected from the general population of Munich. Controls who had central nervous system diseases or psychotic disorders or who had first-degree relatives with psychotic disorders were excluded. *The Technische Universität München study*: This cohort includes 359 healthy, AD, and other dementias patients recruited from the Centre for Cognitive Disorders. All the participants provided written informed consent. A biobank was submitted to the ethics committee of the Technical University of Munich, School of Medicine (Munich, Germany), which raised no objections and approved the biobank (reference number 347-14*). The Göttingen Universität study*: This study includes 111 in- and outpatients with a healthy or AD dementia status from the Department of Psychiatry of the University of Göttingen. The study’s ethical statement was provided locally at the Göttingen University Medical Centre. *The German Dementia Competence Network (DCN) cohort*: Individuals from the DCN cohort were recruited from 14 university hospital memory clinics across Germany between 2003 and 2005 (PMID: 19339779). The study was approved by the respective ethics committees, and written informed consent was obtained from all the participants prior to inclusion. *The German Study on Aging, Cognition, and Dementia (AgeCoDe)*: The

AgeCoDe study is a general practice (GP) registry-based longitudinal study in elderly individuals that recruited patients aged 75 years and above in six German cities from 2003 to 2004 (PMID: 17848793). The study was approved by the respective ethics committees, and written informed consent was obtained from all the participants prior to inclusion.

Greece: the HELIAD study, comprising 49 AD cases and 1,150 controls. HELIAD is a population-based, multidisciplinary, collaborative study designed to estimate, in the Greek population over the age of 64 years, the prevalence and incidence of MCI, AD, other forms of dementia, and other neuropsychiatric conditions of aging and to investigate associations between nutrition and cognitive dysfunction or age-related neuropsychiatric diseases. The participants were selected through random sampling from the records of two Greek municipalities, Larissa and Marousi. All the participants signed informed consent in Greek.

This study was supported by the following grants: IIRG-09-133014 from the Alzheimer's Association; 189 10276/8/9/2011 from the ESPA-EU program Excellence Grant (ARISTEIA), which is co-funded by the European Social Fund and Greek National Resources; and DY2b/oik.51657/14.4.2009 from the Ministry for Health and Social Solidarity(Greece).

Portugal: the Lisbon study from Portugal, totalling 78 AD cases and 74 controls. This cohort was recruited in 2008–2009 to investigate the connections between oxidative stress and lipid dyshomeostasis in AD. The project includes 190 subjects and was approved by the local ethics committee, and all the participants provided written informed consent. This study includes healthy and dementia-by-AD subjects.

Spain: Those samples are part of DEGESCO. DEGESCO Centers from whom DNA samples were genotyped in the German node (1,778 cases and 470 controls) were the Alzheimer Research Center and Memory Clinic, Fundació ACE, Institut Català de Neurociències Aplicades (Barcelona), the Neurology Service at University Hospital Marqués de Valdecilla (Santander), the Alzheimer's disease and other cognitive disorders, Neurology Department, at Hospital Clínic, IDIBAPS (Barcelona), the Molecular Genetics Laboratory, at the Hospital Universitario Central de Asturias (Oviedo), and Fundació Docència i Recerca Mútua de Terrassa and Movement Disorders Unit, Department of Neurology, University Hospital Mútua de Terrassa (Barcelona).

Switzerland: Two datasets from Switzerland and Austria were combined, totaling 182 AD cases and 388 controls. The Lausanne study: This study includes 137 community-dwelling participants aged 55+ years with cognitive impairment (memory clinic patients with MCI, dementia) or normal cognition (recruited by advertisement, word of mouth). The study's ethical statement was provided locally at the Department of Psychiatry, Geneva University Centre, Switzerland. The VITA study: This is a longitudinal study of 606 individuals (Vienna, Austria) who were 75 years old in 2000, followed up every 30–90 months. This cohort includes dementia and healthy subjects. All the participants gave written informed consent. The study conformed to the latest version of the Declaration of Helsinki and was approved by the ethics committee of the City of Vienna, Austria.

**EADB-Netherlands.** In the Dutch node, samples were collected from six organizations in the Netherlands and after QCs, we obtained 2,438 AD cases and 2,389 controls. All these samples were genotyped at the Erasmus Medical University (Rotterdam, The Netherlands). The Medical Ethics Committee (METC) of the local institutes approved the studies. All the participants and/or their legal guardians gave written informed consent for participation in the clinical and genetic studies. Samples from the following institutes were included. 1) *Erasmus Medical Center*: most individuals were selected from population studies from the epidemiology department and accounted for most of the controls, while a smaller subset of samples originated from the neurology department, where AD was diagnosed according to the National Institute of Neurological and Communicative Disorders and Stroke-Alzheimer's Disease and Related Disorders Association (NINCDS-ADRDA) criteria for AD (PMID: 21514250). 2) *The Amsterdam Dementia Cohort (ADC)* (PMID: 29562540): This cohort comprises patients who visit the memory clinic of the VU University Medical Centre, the Netherlands. The diagnosis of probable AD is based on the clinical criteria formulated by the NINCDS–ADRDA and based on the NIA–AA. Diagnosis of MCI was made according to Petersen and NIA-AA. Controls presented with subjective cognitive decline at the memory clinic, but performed within normal limits on all clinical investigations. 3) *The 100-Plus study*: This study includes Dutch-speaking individuals who (i) can provide official evidence for being aged 100 years or older, (ii) self-report to be cognitively healthy, which is confirmed by a proxy, (iii) consent to the donation of a blood sample, (iv) consent to (at least) two home visits from a researcher, and (v) consent to undergo an interview and neuropsychological test battery (PMID: 30362018). 4) *Parelsnoer Institute*: a collaboration between 8 Dutch University Medical Centers in which clinical data and biomaterials from patients suffering from chronic diseases (so called "Pearls") are collected according to harmonized protocols. The Pearl Neurodegenerative Diseases (PMID: 25551191) includes individuals diagnosed with dementia, mild cognitive impairment, and controls with subjective memory complaints. 5) *The Netherlands Brain Bank*: a non-profit organization that collects human brain tissue of donors with a variety of neurological and psychiatric disorders, but also of non-diseased donors. A clinical diagnosis of AD is based on the clinical criteria of probable AD

(PMID: 6610841, PMID: 17616482). The selected AD patients for this study all received a definitive diagnosis which was based on autopsy. 6) *Maastricht University Medical Center*: a subset of individuals that were referred to the memory clinic for cognitive complaints were included if they participated in the BioBank-Alzheimer Centrum Limburg (BB-ACL). Diagnosis of MCI was made according to the criteria of Petersen, and diagnosis of AD-type dementia was made according to the criteria of the DSM-4, and the NINCDS-ADRDA (PMID: 6610841).

**Genotyping**. EADB genomic DNA samples were transferred to 3 genotyping centers and samples that passed the DNA QC were genotyped with the Illumina Infinium Global Screening Array. Raw probe intensities were shared with the CNRGH, which performed the genotype calling on all samples using the same custom cluster file. During the genotyping QC process, three genotyping batches were considered: (1) 49 genotyping chips were identified as possibly problematic and thus were considered as a separate batch, (2) a batch of samples was genotyped and processed after all other samples and (3) the main batch including all other samples. Further details are available in *Bellenguez et al 2022*.

**Additional support for EADB cohorts was provided by:** The work for this manuscript was further supported by the CoSTREAM project (www.costream.eu) and funding from the European Union's Horizon 2020 research and innovation programme under grant agreement No 667375. Italian Ministry of Health (Ricerca Corrente); Ministero dell'Istruzione, dell'Università e della Ricerca–MIUR project “Dipartimenti di Eccellenza 2018–2022” to Department of Neuroscience “Rita Levi Montalcini”, University of Torino (IR), and AIRAlzh Onlus-ANCC-COOP (SB); Partly supported by “Ministero della Salute”, I.R.C.C.S. Research Program, Ricerca Corrente 2018-2020, Linea n. 2 “Meccanismi genetici, predizione e terapie innovative delle malattie complesse” and by the “5 x 1000” voluntary contribution to the Fondazione I.R.C.C.S. Ospedale “Casa Sollievo della Sofferenza”; and RF-2018-12366665, Fondi per la ricerca 2019 (Sandro Sorbi). Karolinska Institutet AD cohort: Dr. Graff and co-authors of the Karolinska Institutet AD cohort report grants from Swedish Research Council (VR) 2015-02926, 2018-02754, 2015-06799, Swedish Alzheimer Foundation, Stockholm County Council ALF and resarch school, Karolinska Institutet StratNeuro, Swedish Demensfonden, and Swedish brain foundation, during the conduct of the study. ADGEN: This work was supported by Academy of Finland (grant numbers 307866); Sigrid Jusélius Foundation; the Strategic Neuroscience Funding of the University of Eastern Finland; EADB project in the JPNDCO-FUND program (grant number 301220). CBAS: Supported by the project no. LQ1605 from the National Program of Sustainability II (MEYS CR), Supported by Ministry of Health of the Czech Republic, grant nr. NV19-04-00270 (All rights reserved), Grant Agency of Charles University Grants No. 693018 and 654217; the Ministry of Health, Czech Republic—conceptual development of research organization, University Hospital Motol, Prague, Czech Republic Grant No. 00064203; the Czech Ministry of Health Project AZV Grant No. 16—27611A; and Institutional Support of Excellence 2. LF UK Grant No. 699012. CNRMAJ-Rouen: This study received fundings from the Centre National de Référence Malades Alzheimer Jeunes (CNRMAJ). The Finnish Geriatric Intervention Study for the Prevention of Cognitive Impairment and Disability (FINGER) data collection was supported by grants from the Academy of Finland, La Carita Foundation, Juho Vainio Foundation, Novo Nordisk Foundation, Finnish Social Insurance Institution, Ministry of Education and Culture Research Grants, Yrjö Jahnsson Foundation, Finnish Cultural Foundation South Osthrobothnia Regional Fund, and EVO/State Research Funding grants of University Hospitals of Kuopio, Oulu and Turku, Seinäjoki Central Hospital and Oulu City Hospital, Alzheimer's Research & Prevention Foundation USA, AXA Research Fund, Knut and Alice Wallenberg Foundation Sweden, Center for Innovative Medicine (CIMED) at Karolinska Institutet Sweden, and Stiftelsen Stockholms sjukhem Sweden. FINGER cohort genotyping was funded by EADB project in the JPND CO-FUND (grant number 301220). Research at the Belgian EADB site is funded in part by the Alzheimer Research Foundation (SAO-FRA), The Research Foundation Flanders (FWO), and the University of Antwerp Research Fund. FK is supported by a BOF DOCPRO fellowship of the University of Antwerp Research Fund. SNAC-K is financially supported by the Swedish Ministry of Health and Social Affairs, the participating County Councils and Municipalities, and the Swedish Research Council. BDR Bristol: We would like to thank the South West Dementia Brain Bank (SWDBB) for providing brain tissue for this study. The SWDBB is part of the Brains for Dementia Research programme, jointly funded by Alzheimer’s Research UK and Alzheimer’s Society and is supported by BRACE (Bristol Research into Alzheimer’s and Care of the Elderly) and the Medical Research Council. BDR Manchester: We would like to thank the Manchester Brain Bankfor providing brain tissue for this study. The Manchester Brain Bank is part of the Brains for Dementia Research programme, jointly funded by Alzheimer’s

Research UK and Alzheimer's Society. BDR KCL: Human post-mortem tissue was provided by the London Neurodegenerative Diseases Brain Bank which receives funding from the UK Medical Research Council and as part of the Brains for Dementia Research programme, jointly funded by Alzheimer's Research UK and the Alzheimer's Society. The CFAS Wales study was funded by the ESRC (RES-060-25-0060) and HEFCW as 'Maintaining function and well-being in later life: a longitudinal cohort study', (Principal Investigators: R.T Woods, L.Clare, G.Windle, V. Burholt, J. Philips, C. Brayne, C. McCracken, K. Bennett, F. Matthews). We are grateful to the NISCHR Clinical Research Centre for their assistance in tracing participants and in interviewing and in collecting blood samples, and to general practices in the study areas for their cooperation. MRC: We thank all individuals who participated in this study. Cardiff University was supported by the Alzheimer's Society (AS; grant RF014/164) and the Medical Research Council (MRC; grants G0801418/1, MR/K013041/1, MR/L023784/1) (R. Sims is an AS Research Fellow). Cardiff University was also supported by the European Joint Programme for Neurodegenerative Disease (JPND; grant MR/L501517/1), Alzheimer's Research UK (ARUK; grant ARUK-PG2014-1), the Welsh Assembly Government (grant SGR544:CADR), Brain's for dementia Research and a donation from the Moondance Charitable Foundation. Cardiff University acknowledges the support of the UK Dementia Research Institute, of which J. Williams is an associate director. Cambridge University acknowledges support from the MRC. Patient recruitment for the MRC Prion Unit/UCL Department of Neurodegenerative Disease collection was supported by the UCLH/UCL Biomedical Centre and NIHR Queen Square Dementia Biomedical Research Unit. The University of Southampton acknowledges support from the AS. King's College London was supported by the NIHR Biomedical Research Centre for Mental Health and the Biomedical Research Unit for Dementia at the South London and Maudsley NHS Foundation Trust and by King's College London and the MRC. ARUK and the Big Lottery Fund provided support to Nottingham University. Alfredo Ramirez: Part of the work was funded by the JPND EADB grant (German Federal Ministry of Education and Research (BMBF) grant: 01ED1619A). German Study on Ageing, Cognition and Dementia in Primary Care Patients (AgeCoDe): This study/publication is part of the German Research Network on Dementia (KND), the German Research Network on Degenerative Dementia (KNDD; German Study on Ageing, Cognition and Dementia in Primary Care Patients; AgeCoDe), and the Health Service Research Initiative (Study on Needs, health service use, costs and health-related quality of life in a large sample of oldestold primary care patients (85+; AgeQualiDe)) and was funded by the German Federal Ministry of Education and Research (grants KND: 01GI0102, 01GI0420, 01GI0422, 01GI0423, 01GI0429, 01GI0431, 01GI0433, 01GI0434; grants KNDD: 01GI0710, 01GI0711, 01GI0712, 01GI0713, 01GI0714, 01GI0715, 01GI0716; grants Health Service Research Initiative: 01GY1322A, 01GY1322B, 01GY1322C, 01GY1322D, 01GY1322E, 01GY1322F, 01GY1322G). In addition, this publication was supported by the German Federal Ministry of Education and Research (Grant numbers: PreADAPT project 01ED2007A and DESCARTES project 01EK2102B to Alfredo Ramirez, 01EK2102A to Anja Schneider, and 01EK2102C to Michael Wagner. VITA study: The support of the Ludwig Boltzmann Society and the AFI Germany have supported the VITA study. The former VITA study group should be acknowledged: W. Danielczyk, G. Gatterer, K Jellinger, S Jugwirth, KH Tragl, S Zehetmayer. Vogel Study: This work was financed by a research grant of the ''Vogelstiftung Dr. Eckernkamp''. HELIAD study: This study was supported by the grants: IIRG-09-133014 from the Alzheimer's Association, 189 10276/8/9/2011 from the ESPA-EU program Excellence Grant (ARISTEIA) and the ΔΥ2β/οικ.51657/14.4.2009 of the Ministry for Health and Social Solidarity (Greece). Biobank Department of Psychiatry, UMG: Prof. Jens Wiltfang is supported by an Ilídio Pinho professorship and iBiMED (UID/BIM/04501/2013), and FCT project PTDC/DTP_PIC/5587/2014 at the University of Aveiro, Portugal. Lausanne study: This work was supported by grants from the Swiss National Research Foundation (SNF 320030_141179). PAGES study: Harald Hampel is an employee of Eisai Inc. During part of this work he was supported by the AXA Research Fund, the "Fondation partenariale Sorbonne Université" and the "Fondation pour la Recherche sur Alzheimer", Paris, France. Mannheim, Germany Biobank: Department of geriatric Psychiatry, Central Institute for Mental Health, Mannheim, University of Heidelberg, Germany. Genotyping for the Swedish Twin Studies of Aging was supported by NIH/NIA grant R01 AG037985. Genotyping in TwinGene was supported by NIH/NIDDK U01 DK066134. WvdF is recipient of Joint Programming for Neurodegenerative Diseases (JPND) grants PERADES (ANR-13-JPRF-0001) and EADB (733051061). Gothenburg Birth Cohort (GBC) Studies: We would like to thank UCL Genomics for performing the genotyping analyses. The studies were supported by The Stena Foundation, The Swedish Research Council (2015-02830, 2013-8717), The Swedish Research Council for Health, Working Life and Wellfare (2013-1202, 2005-0762, 2008-1210, 2013-2300, 2013- 2496, 2013-0475), The Brain Foundation, Sahlgrenska

University Hospital (ALF), The Alzheimer's Association (IIRG-03-6168), The Alzheimer's Association Zenith Award (ZEN-01-3151), Eivind och Elsa K:son Sylvans Stiftelse, The Swedish Alzheimer Foundation. Clinical AD, Sweden: We would like to thank UCL Genomics for performing the genotyping analyses. Barcelona Brain Biobank: Brain Donors of the Neurological Tissue Bank of the Biobanc-Hospital Clinic-IDIBAPS and their families for their generosity. Hospital Clínic de Barcelona Spanish Ministry of Economy and Competitiveness-Instituto de Salud Carlos III and Fondo Europeo de Desarrollo Regional (FEDER), Unión Europea, "Una manera de hacer Europa" grants (PI16/0235 to Dr. R. Sánchez-Valle and PI17/00670 to Dr. A.Antonelli). AA is funded by Departament de Salut de la Generalitat de Catalunya, PERIS 2016-2020 (SLT002/16/00329). Work at JP-T laboratory was possible thanks to funding from Ciberned and generous gifts from Consuelo Cervera Yuste and Juan Manuel Moreno Cervera. Sydney Memory and Ageing Study (Sydney MAS): We gratefully acknowledge and thank the following for their contributions to Sydney MAS: participants, their supporters and the Sydney MAS Research Team (current and former staff and students). Funding was awarded from the Australian National Health and Medical Research Council (NHMRC) Program Grants (350833, 568969, 109308). AddNeuroMed consortium was led by Simon Lovestone, Bruno Vellas, Patrizia Mecocci, Magda Tsolaki, Iwona Kłoszewska, Hilkka Soininen. This work was supported by InnoMed (Innovative Medicines in Europe), an integrated project funded by the European Union of the Sixth Framework program priority (FP6-2004- LIFESCIHEALTH-5). Oviedo: This work was partly supported by Grant from Fondo de Investigaciones Sanitarias-Fondos FEDER EuropeanUnion to Victoria Alvarez PI15/00878. Pascual Sánchez-Juan is supported by CIBERNED and Carlos III Institute of Health, Spain (PI08/0139, PI12/02288, and PI16/01652), jointly funded by Fondo Europeo de Desarrollo Regional (FEDER), Unión Europea, "Una manera de hacer Europa". Project MinE: The ProjectMinE study was supported by the ALS Foundation Netherlands and the MND association (UK) (Project MinE, www.projectmine.com). The SPIN cohort: We are indebted to patients and their families for their participation in the "Sant Pau Initiative on Neurodegeneration cohort", at the Sant Pau Hospital (Barcelona). This is a multimodal research cohort for biomarker discovery and validation that is partially funded by Generalitat de Catalunya (2017 SGR 547 to JC), as well as from the Institute of Health Carlos III-Subdirección General de Evaluación and the Fondo Europeo de Desarrollo Regional (FEDER- "Una manera de Hacer Europa") (grants PI11/02526, PI14/01126, and PI17/01019 to JF; PI17/01895 to AL), and the Centro de Investigación Biomédica en Red Enfermedades Neurodegenerativas programme (Program 1, Alzheimer Disease to AL). We would also like to thank the Fundació Bancària Obra Social La Caixa (DABNI project) to JF and AL; and Fundación BBVA (to AL), for their support in funding this follow-up study. Adolfo López de Munain is supported by Fundación Salud 2000 (PI2013156), CIBERNED and Diputación Foral de Gipuzkoa (Exp.114/17).

#### 1.2 FinnGen

**Current FinnGen Acknowledgment Statement**

We want to acknowledge the participants and investigators of the FinnGen study. The FinnGen project is funded by two grants from Business Finland (HUS 4685/31/2016 and UH 4386/31/2016) and the following industry partners: AbbVie Inc., AstraZeneca UK Ltd, Biogen MA Inc., Bristol Myers Squibb Inc. (and Celgene Corporation & Celgene International II Sàrl), Genentech Inc., Merck Sharp & Dohme LCC, Pfizer Inc., GlaxoSmithKline Intellectual Property Development Ltd., Sanofi US Services Inc., Maze Therapeutics Inc., Johnson&Johnson Innovative Medicine Inc., Novartis AG, Boehringer Ingelheim International GmbH and Bayer AG. Following biobanks are acknowledged for delivering biobank samples to FinnGen: Auria Biobank (www.auria.fi/biopankki), THL Biobank (www.thl.fi/biobank), Helsinki Biobank (www.helsinginbiopankki.fi), Biobank Borealis of Northern Finland (https://www.ppshp.fi/Tutkimus-ja-opetus/Biopankki/Pages/Biobank-Borealis-briefly-in-English.aspx), Finnish Clinical Biobank Tampere (www.tays.fi/en-US/Research_and_development/Finnish_Clinical_Biobank_Tampere), Biobank of Eastern Finland (www.ita-suomenbiopankki.fi/en), Central Finland Biobank (www.ksshp.fi/fi-FI/Potilaalle/Biopankki), Finnish Red Cross Blood Service Biobank (www.veripalvelu.fi/verenluovutus/biopankkitoiminta), Terveystalo Biobank (www.terveystalo.com/fi/Yritystietoa/Terveystalo-Biopankki/Biopankki/) and Arctic Biobank (https://www.oulu.fi/en/university/faculties-and-units/faculty-medicine/northern-finland-birth-cohorts-and-arctic-biobank). All Finnish Biobanks are members of BBMRI.fi infrastructure (https://www.bbmri-eric.eu/national-nodes/finland/). Finnish Biobank Cooperative -FINBB (https://finbb.fi/) is the coordinator of BBMRI-ERIC operations in Finland. The Finnish biobank data can be accessed through the Fingenious® services (https://site.fingenious.fi/en/) managed by FINBB.

**FinnGen R11 ethics statement and materials & methods**

Study subjects in FinnGen provided informed consent for biobank research, based on the Finnish Biobank Act. Alternatively, separate research cohorts, collected prior the Finnish Biobank Act came into effect (in September 2013) and start of FinnGen (August 2017), were collected based on study-specific consents and later transferred to the Finnish biobanks after approval by Fimea (Finnish Medicines Agency), the National Supervisory Authority for Welfare and Health. Recruitment protocols followed the biobank protocols approved by Fimea. The Coordinating Ethics Committee of the Hospital District of Helsinki and Uusimaa (HUS) statement number for the FinnGen study is Nr HUS/990/2017.

The FinnGen study is approved by Finnish Institute for Health and Welfare (permit numbers: THL/2031/6.02.00/2017, THL/1101/5.05.00/2017, THL/341/6.02.00/2018, THL/2222/6.02.00/2018, THL/283/6.02.00/2019, THL/1721/5.05.00/2019 and THL/1524/5.05.00/2020), Digital and population data service agency (permit numbers: VRK43431/2017-3, VRK/6909/2018-3, VRK/4415/2019-3), the Social Insurance Institution (permit numbers: KELA 58/522/2017, KELA 131/522/2018, KELA 70/522/2019, KELA 98/522/2019, KELA 134/522/2019, KELA 138/522/2019, KELA 2/522/2020, KELA 16/522/2020), Findata permit numbers THL/2364/14.02/2020, THL/4055/14.06.00/2020, THL/3433/14.06.00/2020, THL/4432/14.06/2020, THL/5189/14.06/2020, THL/5894/14.06.00/2020, THL/6619/14.06.00/2020, THL/209/14.06.00/2021, THL/688/14.06.00/2021, THL/1284/14.06.00/2021, THL/1965/14.06.00/2021, THL/5546/14.02.00/2020, THL/2658/14.06.00/2021, THL/4235/14.06.00/2021, Statistics Finland (permit numbers: TK-53-1041-17 and TK/143/07.03.00/2020 (earlier TK-53-90-20) TK/1735/07.03.00/2021, TK/3112/07.03.00/2021) and Finnish Registry for Kidney Diseases permission/extract from the meeting minutes on 4th July 2019.

The Biobank Access Decisions for FinnGen samples and data utilized in FinnGen Data Freeze 11 include: THL Biobank BB2017_55, BB2017_111, BB2018_19, BB_2018_34, BB_2018_67, BB2018_71, BB2019_7, BB2019_8, BB2019_26, BB2020_1, BB2021_65, Finnish Red Cross Blood Service Biobank 7.12.2017, Helsinki Biobank HUS/359/2017, HUS/248/2020, HUS/430/2021 §28, §29, HUS/150/2022 §12, §13, §14, §15, §16, §17, §18, §23, §58 and §59, Auria Biobank AB17-5154 and amendment #1 (August 17 2020) and amendments BB_2021-0140, BB_2021-0156 (August 26 2021, Feb 2 2022), BB_2021-0169, BB_2021-0179, BB_2021-0161, AB20-5926 and amendment #1 (April 23 2020) and it´s modification (Sep 22 2021), BB_2022-0262, BB_2022-0256, Biobank Borealis of Northern Finland_2017_1013, 2021_5010, 2021_5018, 2021_5015, 2021_5015 Amendment, 2021_5023, 2021_5023 Amendment, 2021_5017, 2022_6001, 2022_6006 Amendment, BB22-0067, 2022_0262, Biobank of Eastern Finland 1186/2018 and amendment 22§/2020, 53§/2021, 13§/2022, 14§/2022, 15§/2022, 27§/2022, 28§/2022, 29§/2022, 33§/2022, 35§/2022, 36§/2022, 37§/2022, 39§/2022, 7§/2023, Finnish Clinical Biobank Tampere MH0004 and amendments (21.02.2020 & 06.10.2020), 8§/2021, 9§/2021, §9/2022, §10/2022, §12/2022, 13§/2022, §20/2022, §21/2022, §22/2022, §23/2022, 28§/2022, 29§/2022, 30§/2022, 31§/2022, 32§/2022, 38§/2022, 40§/2022, 42§/2022, 1§/2023, Central Finland Biobank 1-2017, BB_2021-0161, BB_2021-0169, BB_2021-0179, BB_2021-0170, BB_2022-0256, and Terveystalo Biobank STB 2018001 and amendment 25th Aug 2020, Finnish Hematological Registry and Clinical Biobank decision 18th June 2021, Arctic biobank P0844: ARC_2021_1001.

#### 1.3 UK Biobank (UKB)

The UK Biobank is comprised of environmental, lifestyle, and genetic data for 500,000 participants. Cases and controls for this analysis were included based on similar criteria to what was used for the FinnGen cohort. AD cases included participants with an ICD10 code of F00 or G30 (UKB codes p130836 and p131036). Controls included participants that did not have an algorithmically defined AD diagnosis as assigned by UKB (UKB code p42020) or any ICD10 code of A81, F01, F02, G31, F03, or I673 (UKB codes p130136, p130838, p130840, p131038, p130842 or p41270). Controls were also filtered for participants currently over 60 years of age to approximate the age distribution of the cases. Cases and controls were filtered for Caucasian ancestry and related individuals removed.

UKB genetic data was genotyped and underwent quality control as described elsewhere (PMID: 30305743). For this manuscript, data imputed using the TOPMed reference panel was used for determining *APOE* genotype and for all subsequent analyses. Data was accessed and analysed on the UKB Research Analysis Platform (RAP) under UK Biobank application ID 33601.

## 1.4 GR@CE

The GR@ACE study (PMID: 31473137) recruited Alzheimer's disease (AD) patients from Fundació ACE, Institut Català de Neurociències Aplicades (Catalonia, Spain), and control individuals from three centers: Fundació ACE (Barcelona, Spain), Valme University Hospital (Seville, Spain), and the Spanish National DNA Bank–Carlos III (University of Salamanca, Spain) (http://www.bancoadn.org). At all sites, AD diagnosis was established by a multidisciplinary working group—including neurologists, neuropsychologists, and social workers—according to the DSM-IV criteria for dementia and the National Institute on Aging and Alzheimer's Association's (NIA–AA) 2011 guidelines for diagnosing AD. In our study, we considered as AD cases any individuals with dementia diagnosed with probable or possible AD at any point in their clinical course. Genotyping was conducted using the Axiom 815K Spanish biobank array (Thermo Fisher) at the Spanish National Centre for Genotyping (CeGEN, Santiago de Compostela, Spain). The genotyping array not only is an adaptation of the Axiom biobank genotyping array but also contains rare population-specific variations observed in the Spanish population.

The Genome Research @ Fundació ACE project (GR@ACE) is supported by Grifols SA, Fundación bancaria 'La Caixa', Fundació ACE, and CIBERNED (Centro de Investigación Biomédica en Red Enfermedades Neurodegenerativas (Program 1, Alzheimer Disease to MB and AR)). A.R. and M.B. receive support from the European Union/EFPIA Innovative Medicines Initiative Joint undertaking ADAPTED and MOPEAD projects (grant numbers 115975 and 115985, respectively). M.B. and A.R. are also supported by national grants PI13/02434, PI16/01861, PI17/01474 and PI19/01240. Acción Estratégica en Salud is integrated into the Spanish National R + D + I Plan and funded by ISCIII (Instituto de Salud Carlos III)–Subdirección General de Evaluación and the Fondo Europeo de Desarrollo Regional (FEDER–'Una manera de hacer Europa'). Some control samples and data from patients included in this study were provided in part by the National DNA Bank Carlos III (www.bancoadn.org, University of Salamanca, Spain) and Hospital Universitario Virgen de Valme (Sevilla, Spain); they were processed following standard operating procedures with the appropriate approval of the Ethical and Scientific Committee.

#### 1.5 European Alzheimer's Disease Initiative (EADI) Consortium

EADI is composed of several case-control studies and one population-based cohort, the 3C study (PMID: 14598854, PMID: 19734903). Case-control studies are comprised of AD cases and cognitively normal controls across France. The population-based cohort, the 3C study, is a prospective study of the relationship between vascular factors and dementia carried out in the three French cities Bordeaux, Montpellier and Dijon. The AD status was defined based on 12 years follow-up for Dijon participants, 14-15 years follow-up for Montpellier participants and 17-18 years follow-up for Bordeaux participants. All other non demented subjects of 3C were included as controls. All AD cases, both in the case-control studies and the 3C study, were ascertained by neurologists and the clinical diagnosis of probable AD was established according to the DSM-III-R and NINCDS-ADRDA criteria. Samples that passed DNA quality control were genotyped with Illumina Human 610-Quad BeadChips.

This work has been developed and supported by the LABEX (laboratory of excellence program investment for the future) DISTALZ grant (Development of Innovative Strategies for a Transdisciplinary approach to ALZheimer's disease) including funding from MEL (Metropole européenne de Lille), ERDF (European Regional Development Fund) and Conseil Régional Nord Pas de Calais. This work was supported by INSERM, the National Foundation for Alzheimer's disease and related disorders, the Institut Pasteur de Lille and the Centre National de Recherche en Génomique Humaine, CEA, the JPND PERADES, the Laboratory of Excellence GENMED (Medical Genomics) grant no. ANR-10-LABX-0013 managed by the National Research Agency (ANR) part of the Investment for the Future program, and the FP7 AgedBrainSysBio. The Three-City Study was performed as part of collaboration between the Institut National de la Santé et de la Recherche Médicale (Inserm), the Victor Segalen Bordeaux II University and Sanofi-Synthélabo. The Fondation pour la Recherche Médicale funded the preparation and initiation of the study. The 3C Study was also funded by the Caisse Nationale Maladie des Travailleurs Salariés, Direction Générale de la Santé, MGEN, Institut de la Longévité,

Agence Française de Sécurité Sanitaire des Produits de Santé, the Aquitaine and Bourgogne Regional Councils, Agence Nationale de la Recherche, ANR supported the COGINUT and COVADIS projects. Fondation de France and the joint French Ministry of Research/INSERM "Cohortes et collections de données biologiques" programme. Lille Génopôle received an unconditional grant from Eisai. The Three-city biological bank was developed and maintained by the laboratory for genomic analysis LAG-BRC - Institut Pasteur de Lille.

#### 1.6 Genetic and Environmental Risk in AD (GERAD) Consortium

Genetic and Environmental Risk in AD (GERAD) Consortium/Defining Genetic, Polygenic, and Environmental Risk for Alzheimer's Disease (PERADES) Consortium
The GERAD/PERADES sample comprises 3,177 Alzheimer's disease cases and 7,277 controls with available age and gender data (PMID: 19734902). Cases and elderly screened controls were recruited by the Medical Research Council (MRC) Genetic Resource for Alzheimer's disease (Cardiff University; Institute of Psychiatry, London; Cambridge University; Trinity College Dublin), the Alzheimer's Research Trust (ART) Collaboration (University of Nottingham; University of Manchester; University of Southampton; University of Bristol; Queen's University Belfast; the Oxford Project to Investigate Memory and Ageing (OPTIMA), Oxford University); Washington University, St Louis, United States; MRC PRION Unit, University College London; London and the South East Region Alzheimer's disease project (LASER-AD), University College London; Competence Network of Dementia (CND) and Department of Psychiatry, University of Bonn, Germany; the National Institute of Mental Health (NIMH) Alzheimer's disease Genetics Initiative. 6129 population controls were drawn from large existing cohorts with available GWAS data, including the 1958 British Birth Cohort (1958BC) (http://www.b58cgene.sgul.ac.uk), the KORA F4 Study and the Heinz Nixdorf Recall Study. All Alzheimer's disease cases met criteria for either probable (NINCDS-ADRDA, DSM-IV) or definite (CERAD) Alzheimer's disease. All elderly controls were screened for dementia using the MMSE or ADAS-cog, were determined to be free from dementia at neuropathological examination or had a Braak score of 2.5 or lower. Genotypes from all cases and 4617 controls were previously included in the AD GWAS by Harold and colleagues (PMID: 19734902). Genotypes for the remaining 2660 population controls were obtained from WTCCC2.

We thank all individuals who participated in this study. Cardiff University was supported by the Wellcome Trust, Alzheimer's Society (AS; grant RF014/164), the Medical Research Council (MRC; grants G0801418/1, MR/K013041/1, MR/L023784/1), the European Joint Programme for Neurodegenerative Disease (JPND, grant MR/L501517/1), Alzheimer's Research UK (ARUK, grant ARUK-PG2014-1), Welsh Assembly Government (grant SGR544:CADR), a donation from the Moondance Charitable Foundation, UK Dementia's Platform (DPUK, reference MR/L023784/1), and the UK Dementia Research Institute at Cardiff. Cambridge University acknowledges support from the MRC. ARUK supported sample collections at the Kings College London, the South West Dementia Bank, Universities of Cambridge, Nottingham, Manchester and Belfast. King's College London was supported by the NIHR Biomedical Research Centre for Mental Health and Biomedical Research Unit for Dementia at the South London and Maudsley NHS Foundation Trust and Kings College London and the MRC. Alzheimer's Research UK (ARUK) and the Big Lottery Fund provided support to Nottingham University. Ulster Garden Villages, AS, ARUK, American Federation for Aging Research, NI R&D Office and the Royal College of Physicians/Dunhill Medical Trust provided support for Queen's University, Belfast. The University of Southampton acknowledges support from the AS. The MRC and Mercer's Institute for Research on Ageing supported the Trinity College group. DCR is a Wellcome Trust Principal Research fellow. The South West Dementia Brain Bank acknowledges support from Bristol Research into Alzheimer's and Care of the Elderly. The Charles Wolfson Charitable Trust supported the OPTIMA group. Washington University was funded by NIH grants, Barnes Jewish Foundation and the Charles and Joanne Knight Alzheimer's Research Initiative. Patient recruitment for the MRC Prion Unit/UCL Department of Neurodegenerative Disease collection was supported by the UCLH/UCL Biomedical Research Centre and their work was supported by the NIHR Queen Square Dementia BRU, the Alzheimer's Research UK and the Alzheimer's Society. LASER-AD was funded by Lundbeck SA. The AgeCoDe study group was supported by the German Federal Ministry for Education and Research grants 01 GI 0710, 01 GI 0712, 01 GI 0713, 01 GI 0714, 01 GI 0715, 01 GI 0716, 01 GI 0717. Genotyping of the Bonn case-control sample was funded by the German centre for Neurodegenerative Diseases (DZNE), Germany. The GERAD Consortium also used samples ascertained by the NIMH AD Genetics Initiative. HH was supported by a grant of the Katharina-Hardt-Foundation, Bad

Homburg vor der Höhe, Germany. The KORA F4 studies were financed by Helmholtz Zentrum München; German Research Center for Environmental Health; BMBF; German National Genome Research Network and the Munich Center of Health Sciences. The Heinz Nixdorf Recall cohort was funded by the Heinz Nixdorf Foundation (Dr. Jur. G.Schmidt, Chairman) and BMBF. We acknowledge use of genotype data from the 1958 Birth Cohort collection and National Blood Service, funded by the MRC and the Wellcome Trust which was genotyped by the Wellcome Trust Case Control Consortium and the Type-1 Diabetes Genetics Consortium, sponsored by the National Institute of Diabetes and Digestive and Kidney Diseases, National Institute of Allergy and Infectious Diseases, National Human Genome Research Institute, National Institute of Child Health and Human Development and Juvenile Diabetes Research Foundation International. The project is also supported through the following funding organisations under the aegis of JPND - www.jpnd.eu (United Kingdom, Medical Research Council (MR/L501529/1; MR/R024804/1) and Economic and Social Research Council (ES/L008238/1)) and through the Motor Neurone Disease Association. This study represents independent research part funded by the National Institute for Health Research (NIHR) Biomedical Research Centre at South London and Maudsley NHS Foundation Trust and King's College London. Prof Jens Wiltfang is supported by an Ilídio Pinho professorship and iBiMED (UID/BIM/04501/2013), at the University of Aveiro, Portugal.

#### 1.7 Bonn studies

The Bonn group would like to thank Dr. Heike Koelsch for her scientific support.
The Bonn samples are part of the German Dementia Competence Network (DCN) and the German Research Network on Degenerative Dementia (KNDD, AgeCoDe study group), which are funded by the German Federal Ministry of Education and Research (BMBF): grants DCN: 01G10102, 01GI0420, 01GI0422, 01GI0423, 01GI0429, 01GI0431, 01GI0433, 04GI0434, 01GI0711; grants KNDD: 01GI1007A, 01GI0710, 01GI0711, 01GI0712, 01GI0713, 01GI0714, 01GI0715, 01GI0716, 01GI0717, 01ET1006B. Controls for the Bonn samples were also obtained from the Heinz Nixdorf Recall cohort that was funded by the Heinz Nixdorf Foundation (Dr. Jur. G.Schmidt, Chairman) and BMBF. Genotyping of the Bonn case-control sample was funded by the German Centre for Neurodegenerative Diseases (DZNE), Germany. Markus M Nöthen and Alfredo Ramirez are members of the German Research Foundation (DFG) cluster of excellence ImmunoSensation.

Bonn OMNI cohort: the Bonn OMNI cohort consists of AD patients and controls derived from a larger German GWAS cohort which was recruited from the following three sources: (i) the German Dementia Competence Network; (ii) the German study on Aging, Cognition, and Dementia in primary care patients (AgeCoDe); and (iii) the interdisciplinary Memory Clinic at the University Hospital of Bonn. The control sample comprised of individuals from the population-based study Heinz Nixdorf Recall (HNR) study cohort. This sample was previously used for replication in Lambert et al.[33]. *The German study on aging, cognition and dementia (AgeCoDe)*: see description above. *The German competence network cohort (DCN)*: The DCN cohort includes 1,095 patients with mild cognitive impairment (MCI) and 648 cases with mild Alzheimer's disease (AD) clinical dementia syndrome that were recruited from 14 university hospital memory clinics across Germany between 2003 and 2005[9]. Exclusion criteria were substance abuse or dependence, insufficient German language skills, multi-morbidity, comorbid condition with excess mortality, circumstances that would have made regular attendance at follow-up visits questionable and lack of an informant. The diagnosis of mild dementia according to ICD-10 criteria required a decline of cognitive ability (at least 1 SD) from a previous level in at least 2 domains as evidenced by age-corrected standardized tests, impairment in activities of daily living (i.e. B-ADL > 6), changes in personality, drive, social behavior or control of emotion but no clouding of consciousness. These changes must have persisted for at least 3 months. The etiological diagnosis of AD was assigned according to NINCDS-ADRDA criteria[8]. *Memory clinic Bonn*: The interdisciplinary Memory Clinic of the Department of Psychiatry and Department of Neurology at the University Hospital in Bonn provided further patients. Diagnoses were assigned according the NINCDS/ADRDA criteria[8] and on the basis of clinical history, physical examination, neuropsychological testing (using the CERAD neuropsychological battery, including the MMSE), laboratory assessments, and brain imaging. *Control sample:* In the Heinz Nixdorf Recall (Risk Factors, Evaluation of Coronary Calcification, and Lifestyle) study, participants were randomly sampled in three cities in Germany. The study design has previously been described[34,35]. Briefly, 4814 participants aged 45 to 75 years were enrolled between 2000 and 2003 (t0, baseline). Cognitive performance of participants was evaluated at follow up scheduled 5 years after baseline (t1, n = 4157, 2005–2008) and then again at follow up 5 years after t1 (t2, n = 3087, 2010–2015). Controls sample was selected if participant did not present cognitive impairment as reported at the last available evaluation. Cognitive evaluation has been described extensively previously[36,37]. Herein, cognitive impairment at t1 was defined as a performance of one standard deviation (SD) below the age- and education-adjusted mean except for the clock-drawing test, where a performance ≥3 was rated as impaired (for a detailed description, see the study by Winkler et al.[36]). The study was approved by the University of Duisburg-Essen Institutional Review Board and followed established guidelines of good epidemiological practice.
